# The Burden of Digestive Diseases in the United States Population

**DOI:** 10.1101/2023.08.16.23294166

**Authors:** Aynur Unalp-Arida, Constance E. Ruhl

## Abstract

**Background and rationale.** Digestive diseases are common and lead to significant morbidity, mortality, and health care utilization. We used national survey and claims databases to expand on earlier findings and investigate current trends in the digestive disease burden in the United States. **Methods.** The National Ambulatory Medical Care Survey, Nationwide Emergency Department Sample, National Inpatient Sample, Vital Statistics of the U.S., Surveillance, Epidemiology, and End Results Program, Optum Clinformatics® Data Mart, and Centers for Medicare and Medicaid Services Medicare 5% Sample databases were used to estimate medical care, mortality, cancer incidence, and claims-based prevalence with a digestive disease diagnosis. Rates were age-adjusted (for national databases) and shown per 100,000 population. **Results.** For all digestive diseases, prevalence (claims-based, all-listed diagnoses) was 30.5% among commercial insurance enrollees (2020) and 53.1% among Medicare beneficiaries (2019). In the U.S. population, digestive diseases contributed to approximately 126 million ambulatory care visits (2015), 41 million emergency department visits (2018), 16 million hospital discharges (2018), and 472,000 deaths (2019) annually. Prevalence, medical care, and mortality rates with a digestive disease diagnosis were higher among children and younger adults (except for emergency department visits) and then increased with age. Women had higher prevalence and medical care rates with a digestive disease diagnosis, but mortality rates were higher among men. Prevalence and medical care rates with a digestive disease diagnosis were higher among Blacks, followed by Whites, then Hispanics, and lowest among Asians. Mortality rates were higher among Blacks compared with Whites and lower among Hispanics compared with non-Hispanics. Between 2004 and the most recent year, ambulatory care visit rates with a digestive disease diagnosis increased by 4%, hospital discharge rates decreased by 3%, and mortality rates decreased by 7%. Among commercial insurance enrollees, rates were higher compared with national data for ambulatory care visits and hospital discharges, but lower for emergency department visits. The medical care use and mortality burdens varied among individual digestive diseases. **Conclusion.** The digestive disease burden in the United States is substantial, particularly among Blacks and older adults.

## INTRODUCTION

Digestive diseases affect multiple organs and systems including the alimentary tract, liver and biliary system, and pancreas. These disorders have diverse causes such as congenital and genetic anomalies, acute and chronic infections, environmental risk factors, and adverse effects of drugs and toxins. The burden of digestive diseases ranges from the inconvenience of a diarrheal disease to chronic debilitating illnesses requiring continuous medical care to fatal conditions such as pancreatic cancer. There have been significant changes in the prevalence and overall burden of digestive diseases in the 21^st^ century building on earlier public health gains from improved sanitation, food safety, and development of new drugs, vaccines, diagnostic tests, and minimally invasive procedures. The well-being of the population of the United States requires a continued investment in digestive disease research and public health initiatives supported by a health care system capable of providing these advances to all equitably.

As outlined in the National Institute of Diabetes and Digestive and Kidney Diseases (NIDDK) Strategic Plan for Research,(1) a better understanding of social determinants of health, such as access to healthy food and safe places to exercise, may help to reduce health disparities and achieve health equity which may in turn decrease the burden of digestive diseases among racial and ethnic minority and other underserved populations. In this descriptive epidemiology paper, we present essential health data to reduce the burden of digestive diseases and to inform health care providers, researchers, administrators, public officials, professional and patient-based organizations, and the public to act. Our work follows the tradition of National Institutes of Health (NIH) sponsored publications *Digestive diseases in the United States: epidemiology and impact* in 1994,(2) and *Opportunities and challenges in digestive diseases research: recommendations of the National Commission on Digestive Diseases* in 2009,(3) and its companion report on the burden of digestive diseases.(4) Close examination of these reports reveals interesting trends in various digestive diseases despite numerous limitations on the types of data that can be obtained in the diverse and decentralized U.S. health care system. Despite many limitations of available data sources, there are several certainties that may be gleaned, such as a century of progress in health care delivery, scientific advances, and a still staggering burden of digestive diseases in the United States.

## METHODS

Digestive disease morbidity was identified by an International Classification of Diseases (ICD), Tenth Revision, Clinical Modification (ICD-10-CM) codes and digestive disease mortality by ICD, Tenth Revision (ICD-10) codes (**Appendix 1**). The first-listed diagnosis was considered the primary diagnosis and all remaining diagnoses were considered secondary and included under ‘all-listed’. For national event-level data sources, diagnoses were counted only once under the all-listed category, irrespective of the number of actual diagnoses listed on a medical record or death certificate.

The National Ambulatory Medical Care Survey (NAMCS), Healthcare Cost and Utilization Project (HCUP) Nationwide Emergency Department Sample (NEDS), HCUP National Inpatient Sample (NIS), Vital Statistics of the U.S., Surveillance, Epidemiology, and End Results (SEER) Program, Optum Clinformatics® Data Mart (CDM), and Centers for Medicare and Medicaid Services (CMS) Medicare 5% Sample databases were used to estimate medical care, mortality, years of potential life lost, cancer incidence, and claims-based prevalence with primary or other digestive disease diagnoses. These data sources are described in more detail in **Appendix 2**. Estimates for the total population and demographic groups were calculated for the most recent year of data available. Rates were age-adjusted (for national databases) and shown per 100,000 population. For national data sources, ambulatory care, emergency department, and hospital numbers and rates represent events, not persons with an event. Statistical methodology is described in more detail in **Appendix 3**.

There were an estimated 66 million ambulatory care visits to office-based physicians with a first-listed diagnosis of a digestive disease and 126 million visits with a digestive disease diagnosis listed in any position (2015) (Table 1). The ambulatory care visit rate (per 100,000 U.S. population) was 19,962 with a first-listed diagnosis and 37,173 with an all-listed diagnosis. In other words, for every 100 U.S. residents, there were 37 ambulatory care visits at which a digestive disease diagnosis was noted and 20 visits for which a digestive disease diagnosis was primary. All-listed diagnosis rates were higher among children compared with adolescents and younger adults and then increased with age. Age-adjusted rates of all-listed diagnoses were higher among women compared with men, Blacks compared with Whites, and Hispanics compared with non-Hispanics. Between 2004 and 2015, the number of ambulatory care visits with a primary diagnosis of a digestive disease decreased from an estimated 72 million to 66 million and the number with an all-listed diagnosis increased from 105 million to 126 million. Age-adjusted ambulatory care visit rates (per 100,000) with a primary digestive disease diagnosis decreased by 19% overall from 24,543 to 19,962 and decreased among sex and race groups.(4,5) All-listed diagnosis rates increased by 4% overall from 35,684 to 37,173 and increased among women, Whites, and Blacks, but decreased among men.

**Table 1:**
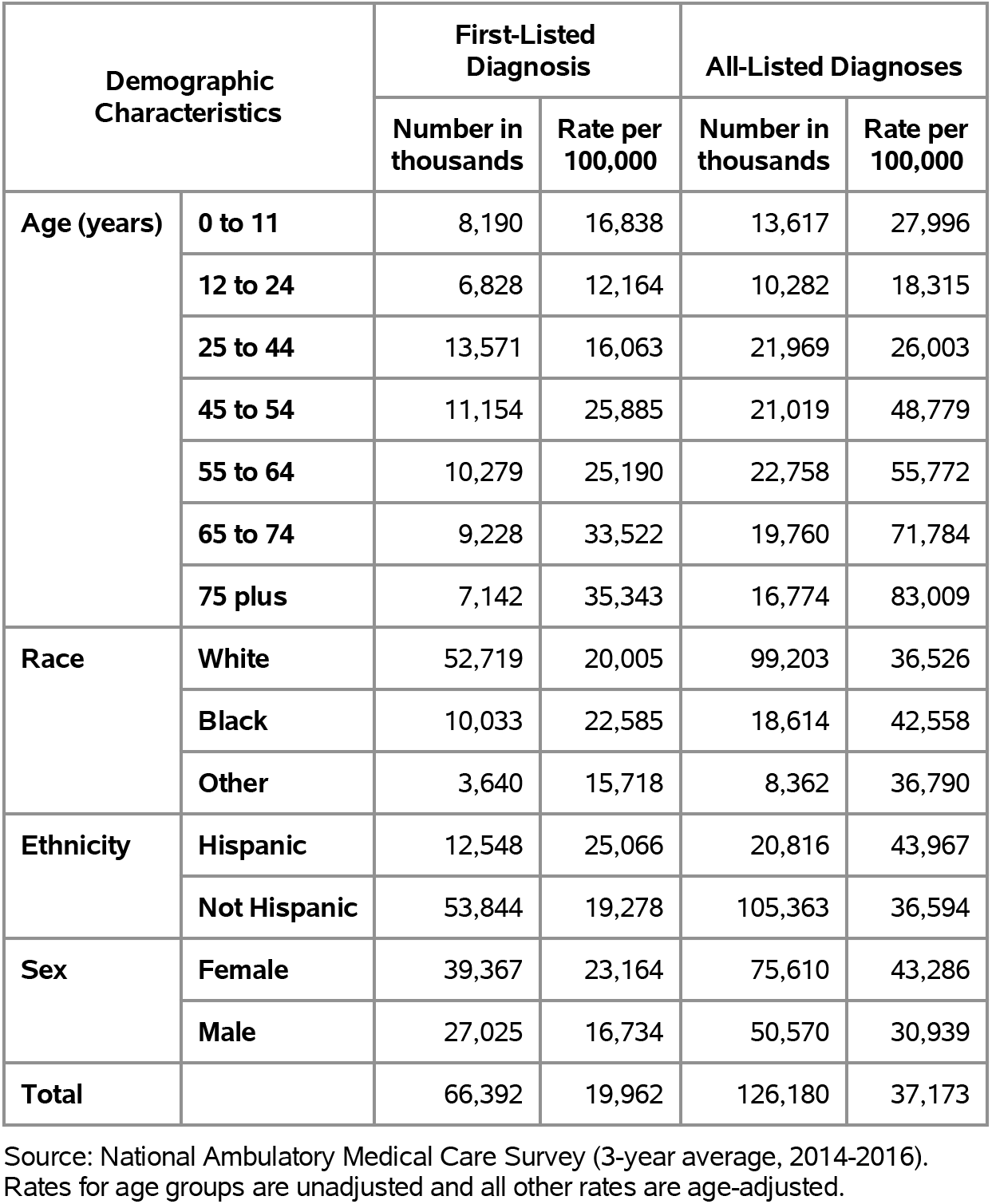
All Digestive Diseases: Ambulatory care visits with first-listed and all-listed diagnoses by age, race, ethnicity, and sex in the United States, 2015.

| Demographic Characteristics |  | First-Listed Diagnosis |  | All-Listed Diagnoses |  |
| --- | --- | --- | --- | --- | --- |
|  |  | Number in thousands | Rate per 100,000 | Number in thousands | Rate per 100,000 |
| Age (years) | 0 to 11 | 8,190 | 16,838 | 13,617 | 27,996 |
|  | 12 to 24 | 6,828 | 12,164 | 10,282 | 18,315 |
|  | 25 to 44 | 13,571 | 16,063 | 21,969 | 26,003 |
|  | 45 to 54 | 11,154 | 25,885 | 21,019 | 48,779 |
|  | 55 to 64 | 10,279 | 25,190 | 22,758 | 55,772 |
|  | 65 to 74 | 9,228 | 33,522 | 19,760 | 71,784 |
|  | 75 plus | 7,142 | 35,343 | 16,774 | 83,009 |
| Race | White | 52,719 | 20,005 | 99,203 | 36,526 |
|  | Black | 10,033 | 22,585 | 18,614 | 42,558 |
|  | Other | 3,640 | 15,718 | 8,362 | 36,790 |
| Ethnicity | Hispanic | 12,548 | 25,066 | 20,816 | 43,967 |
|  | Not Hispanic | 53,844 | 19,278 | 105,363 | 36,594 |
| Sex | Female | 39,367 | 23,164 | 75,610 | 43,286 |
|  | Male | 27,025 | 16,734 | 50,570 | 30,939 |
| Total |  | 66,392 | 19,962 | 126,180 | 37,173 |

Source: National Ambulatory Medical Care Survey (3-year average, 2014-2016). Rates for age groups are unadjusted and all other rates are age-adjusted.

There were an estimated 19 million emergency department visits with a first-listed diagnosis of a digestive disease and 41 million visits with a digestive disease diagnosis listed in any position in 2018 (Table 2). The emergency department visit rate (per 100,000) was 5,697 with a first-listed diagnosis and 12,064 with an all-listed diagnosis. This equates to 6 emergency department visits with a first-listed digestive disease diagnosis per 100 U.S. residents and 12 visits where a digestive disease was listed in any position. All-listed diagnosis rates increased with age, especially among persons 75 years and over. Age-adjusted rates were higher among women compared with men. National emergency department visit rates were not included in The Burden of Digestive Diseases in the United States compendium so rate trends cannot be examined.(4)

**Table 2:**
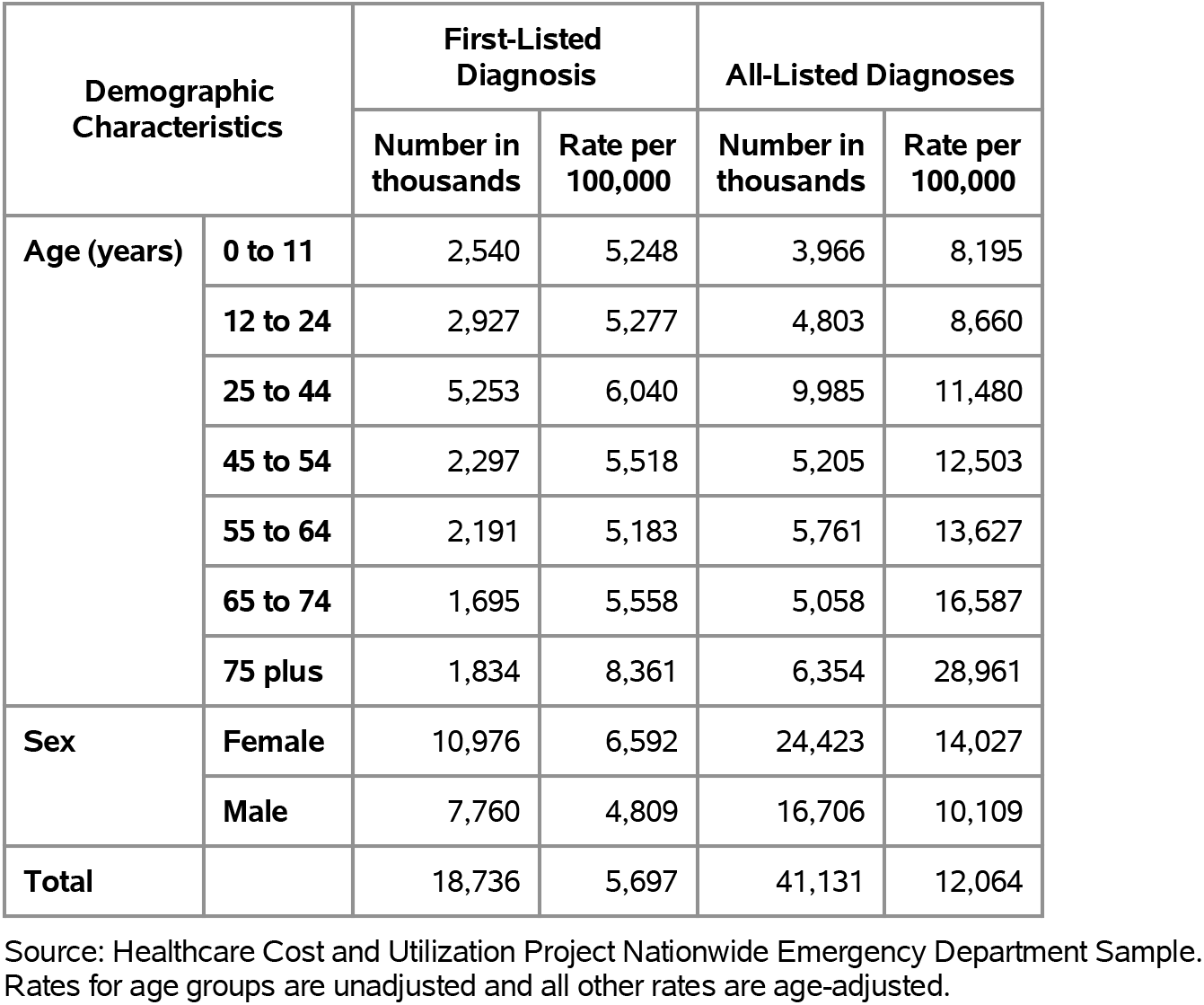
All Digestive Diseases: Emergency department visits with first-listed and all-listed diagnoses by age and sex in the United States, 2018.

There were an estimated 4.0 million hospital discharges with a first-listed diagnosis of a digestive disease and 16 million discharges with a digestive disease diagnosis listed in any position in 2018 (Table 3). The hospital discharge rate (per 100,000) was 1,098 with a first-listed diagnosis and 4,478 with an all-listed diagnosis. This equates to 1 overnight hospital stay per 100 U.S. residents with a first-listed digestive disease diagnosis and 4 visits that included a digestive disease as a discharge diagnosis in any position. All-listed diagnosis rates were much higher among children compared with adolescents and the youngest adults and then increased with age. Age-adjusted rates were higher among women compared with men, Blacks compared with Whites, and non-Hispanics compared with Hispanics. Between 2004 and 2018, the number of hospital discharges with a primary diagnosis of a digestive disease decreased from an estimated 4.6 million to 4.0 million and the number with an all-listed diagnosis increased from 14 million to 16 million. Age-adjusted hospital discharge rates (per 100,000) with a primary digestive disease diagnosis decreased by 30% overall from 1,563 to 1,098 and decreased among sex and race groups.(4,5) All-listed diagnosis rates decreased by only 3% overall from 4,608 to 4,478 and decreased among women, but increased among men, Whites, and Blacks.

**Table 3:** All Digestive Diseases: Hospital discharges with first-listed and all-listed diagnoses by age, race, ethnicity, and sex in the United States, 2018.

| Demographic Characteristics |  | First-Listed Diagnosis |  | All-Listed Diagnoses |  |
| --- | --- | --- | --- | --- | --- |
|  |  | Number in thousands | Rate per 100,000 | Number in thousands | Rate per 100,000 |
| Age (years) | 0 to 11 | 154 | 318 | 1,185 | 2,449 |
|  | 12 to 24 | 187 | 338 | 549 | 990 |
|  | 25 to 44 | 674 | 775 | 2,193 | 2,521 |
|  | 45 to 54 | 559 | 1,343 | 1,874 | 4,500 |
|  | 55 to 64 | 774 | 1,830 | 2,995 | 7,085 |
|  | 65 to 74 | 760 | 2,493 | 3,328 | 10,914 |
|  | 75 plus | 885 | 4,033 | 4,327 | 19,724 |
| Race | White | 3,233 | 1,102 | 13,188 | 4,407 |
|  | Black | 531 | 1,205 | 2,331 | 5,280 |
|  | Other | 267 | 1,044 | 1,029 | 4,121 |
| Ethnicity | Hispanic | 541 | 1,099 | 1,831 | 3,810 |
|  | Not Hispanic | 3,490 | 1,111 | 14,716 | 4,628 |
| Sex | Female | 2,135 | 1,116 | 8,920 | 4,603 |
|  | Male | 1,857 | 1,082 | 7,529 | 4,369 |
| Total |  | 3,993 | 1,098 | 16,450 | 4,478 |

Source: Healthcare Cost and Utilization Project National Inpatient Sample. Rates for age groups are unadjusted and all other rates are age-adjusted.

A digestive disease was the underlying cause of 294,000 deaths and an underlying or contributing cause of 472,000 deaths in 2019 (Table 4). Mortality rates (per 100,000) were 72.4 as underlying cause and 116.3 as underlying or other cause. Mortality rates (underlying or other cause) were higher among children compared with adolescents and the youngest adults and then increased with age. In contrast to health care rates, age-adjusted mortality rates were higher among men compared with women. They were higher among Blacks compared with Whites and among non-Hispanics compared with Hispanics. Between 2004 and 2019, the number of deaths with a digestive disease as underlying cause increased from 236,000 to 294,000 and the number with a digestive disease as underlying or other cause increased from 366,000 to 472,000. Age-adjusted mortality rates (per 100,000) with a digestive disease as underlying cause decreased by 10% overall from 80.4 to 72.4 and decreased among sex and race groups.(4,5) Rates with a digestive disease as underlying or other cause decreased by 7% overall from 124.8 to 116.3 and decreased among sex and race groups.

**Table 4:**
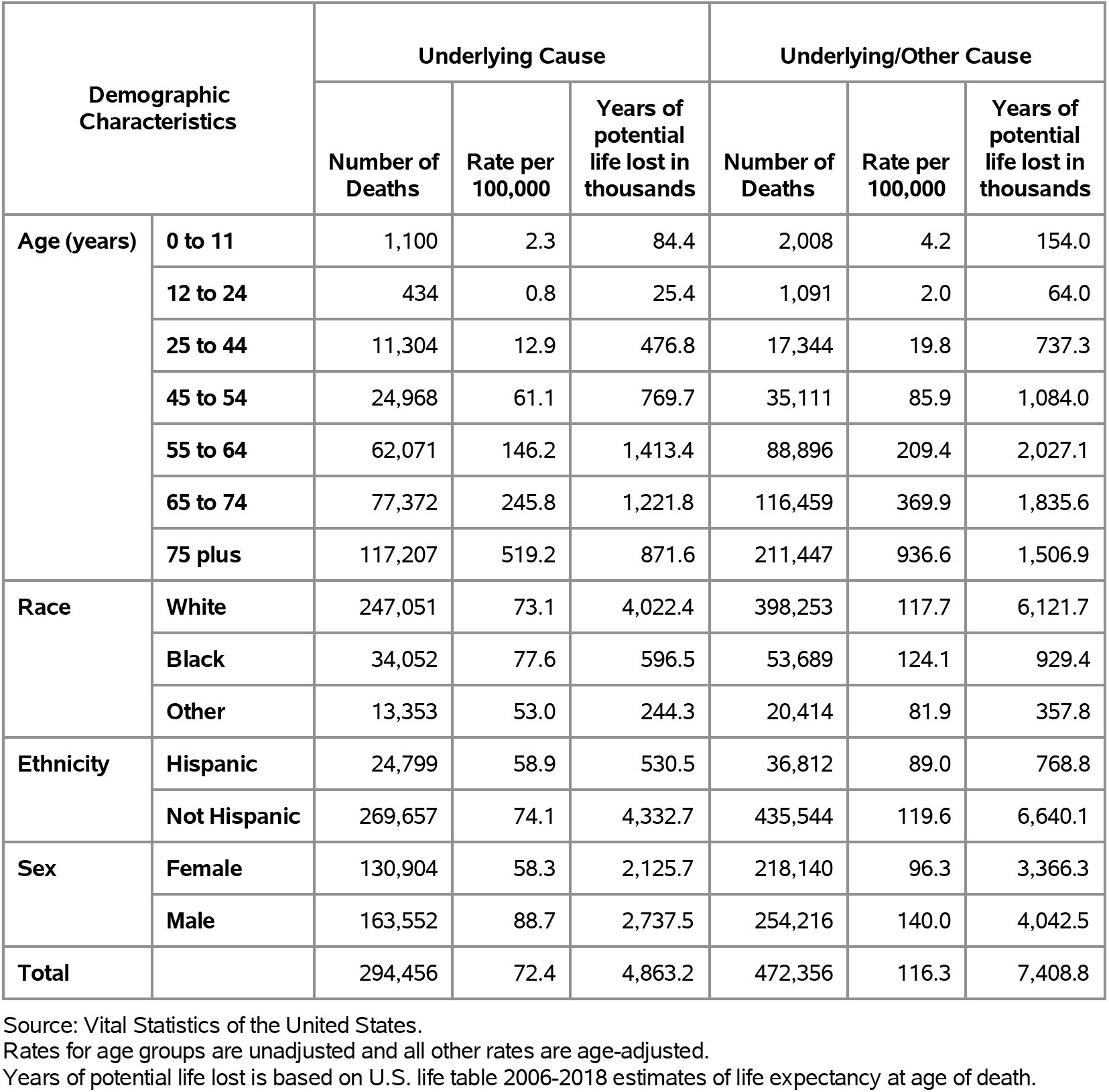
All Digestive Diseases: Deaths with underlying or underlying/other cause and lifetime years of life lost by age, race, ethnicity, and sex in the United States, 2019.

In contrast to national data that represent events, Optum data are at the person level which enables calculation of claims-based prevalence. Among 13.7 million private insurance enrollees (2020), 2.2 million had a first-listed digestive disease diagnosis (claims-based prevalence of 16.1%) and 4.2 million had an all-listed diagnosis (claims-based prevalence of 30.5%) (Table 5). Prevalence (all-listed diagnoses) was higher among children compared with adolescents and the youngest adults and then increased with age until the oldest age group. Prevalence was higher among women compared with men. It was highest among Blacks, followed by Whites, then Hispanics, and lowest among Asians. Prevalence was higher in the Northeast and South compared with the Midwest and West.

**Table 5:** All Digestive Diseases: Claims-based prevalence with first-listed and all-listed diagnoses by age, race-ethnicity, sex and region among privately insured enrollees, 2020.

| Demographic Characteristics |  | Number of enrollees | First-Listed Diagnosis |  | All-Listed Diagnoses |  |
| --- | --- | --- | --- | --- | --- | --- |
|  |  |  | Number of enrollees with disease | Percent of enrollees with disease | Number of enrollees with disease | Percent of enrollees with disease |
| Age (years) | 0 to 11 | 1,101,978 | 93,661 | 8.5% | 172,707 | 15.7% |
|  | 12 to 24 | 1,451,892 | 111,327 | 7.7% | 176,361 | 12.1% |
|  | 25 to 44 | 2,656,672 | 284,972 | 10.7% | 492,626 | 18.5% |
|  | 45 to 54 | 1,452,320 | 248,092 | 17.1% | 464,662 | 32.0% |
|  | 55 to 64 | 1,640,216 | 341,348 | 20.8% | 654,682 | 39.9% |
|  | 65 to 74 | 2,840,615 | 631,047 | 22.2% | 1,188,635 | 41.8% |
|  | 75 plus | 2,576,241 | 504,913 | 19.6% | 1,030,999 | 40.0% |
| Race-ethnicity | White | 8,657,702 | 1,449,645 | 16.7% | 2,737,997 | 31.6% |
|  | Black | 1,242,477 | 228,501 | 18.4% | 433,613 | 34.9% |
|  | Hispanic | 1,493,457 | 239,333 | 16.0% | 446,788 | 29.9% |
|  | Asian | 607,284 | 74,540 | 12.3% | 140,459 | 23.1% |
|  | Unknown | 1,719,014 | 223,341 | 13.0% | 421,815 | 24.5% |
| Sex | Female | 7,199,363 | 1,262,266 | 17.5% | 2,396,094 | 33.3% |
|  | Male | 6,520,571 | 953,094 | 14.6% | 1,784,578 | 27.4% |
| Region | Northeast | 1,526,275 | 277,389 | 18.2% | 500,914 | 32.8% |
|  | Midwest | 3,322,846 | 494,252 | 14.9% | 942,800 | 28.4% |
|  | South | 5,619,435 | 985,884 | 17.5% | 1,868,071 | 33.2% |
|  | West | 3,251,378 | 457,835 | 14.1% | 868,887 | 26.7% |
| Total |  | 13,719,934 | 2,215,360 | 16.1% | 4,180,672 | 30.5% |

Source: De-identified Optum Clinformatic® Data Mart.
Enrollees with full enrollment in a commercial health plan or Medicare during the year.

Among commercial insurance enrollees, ambulatory care visit rates with both first-listed and all-listed digestive disease diagnoses were much higher compared with national data; however, availability of more recent data for commercial insurance enrollees (2020 vs. 2015) may limit comparability (Table 6). Like national data, ambulatory care visit rates (all-listed diagnoses) were higher among children compared with adolescents and the youngest adults and then increased with age and were higher among women compared with men. Optum data include information on ambulatory care visits among Asians that are not available in national data used in this paper. Among persons with known race-ethnicity, rates were highest among Blacks, followed by Whites and Hispanics, and lowest among Asians. Ambulatory care visit rates were higher in the South and Northeast compared with the Midwest and West.

**Table 6:** All Digestive Diseases: Ambulatory care visits with first-listed and all-listed diagnoses by age, race-ethnicity, sex and region among privately insured enrollees, 2020.

| Demographic Characteristics |  | First-Listed Diagnosis |  | All-Listed Diagnoses |  |
| --- | --- | --- | --- | --- | --- |
|  |  | Number of events | Event rate per 100,000 | Number of events | Event rate per 100,000 |
| Age (years) | 0 to 11 | 138,742 | 12,590 | 283,215 | 25,701 |
|  | 12 to 24 | 194,991 | 13,430 | 322,026 | 22,180 |
|  | 25 to 44 | 558,515 | 21,023 | 1,032,553 | 38,866 |
|  | 45 to 54 | 513,234 | 35,339 | 1,073,396 | 73,909 |
|  | 55 to 64 | 737,616 | 44,971 | 1,644,058 | 100,234 |
|  | 65 to 74 | 1,380,964 | 48,615 | 3,081,539 | 108,481 |
|  | 75 plus | 1,124,690 | 43,656 | 2,911,397 | 113,009 |
| Race-ethnicity | White | 3,045,440 | 35,176 | 6,749,160 | 77,956 |
|  | Black | 491,995 | 39,598 | 1,162,933 | 93,598 |
|  | Hispanic | 521,380 | 34,911 | 1,139,865 | 76,324 |
|  | Asian | 150,771 | 24,827 | 322,664 | 53,132 |
|  | Unknown | 439,166 | 25,548 | 973,562 | 56,635 |
| Sex | Female | 2,657,694 | 36,916 | 6,082,612 | 84,488 |
|  | Male | 1,991,058 | 30,535 | 4,265,572 | 65,417 |
| Region | Northeast | 598,611 | 39,220 | 1,289,784 | 84,505 |
|  | Midwest | 969,127 | 29,166 | 2,153,490 | 64,809 |
|  | South | 2,128,406 | 37,876 | 4,802,169 | 85,456 |
|  | West | 952,608 | 29,299 | 2,102,741 | 64,672 |
| Total |  | 4,648,752 | 33,883 | 10,348,184 | 75,424 |

Source: De-identified Optum Clinformatic® Data Mart.
Enrollees with full enrollment in a commercial health plan or Medicare during the year.

Among commercial insurance enrollees, emergency department visit rates with both first-listed and all-listed digestive disease diagnoses were much lower compared with national data; however, availability of more recent data for commercial insurance enrollees (2020 vs. 2018) may limit comparability (Table 7). Like national data, emergency department visit rates (all-listed diagnoses) increased with age and were higher among women compared with men. Optum data include information on emergency department visits by race-ethnicity that are not available in national data used in this paper. Among persons with known race-ethnicity, rates were highest among Blacks, followed by Whites and Hispanics, and lowest among Asians. Emergency department visit rates were higher in the South and Northeast compared with the Midwest and West.

**Table 7:** All Digestive Diseases: Emergency department visits with first-listed and all-listed diagnoses by age, race-ethnicity, sex and region among privately insured enrollees, 2020.

| Demographic Characteristics |  | First-Listed Diagnosis |  | All-Listed Diagnoses |  |
| --- | --- | --- | --- | --- | --- |
|  |  | Number of events | Event rate per 100,000 | Number of events | Event rate per 100,000 |
| Age (years) | 0 to 11 | 965 | 88 | 1,415 | 128 |
|  | 12 to 24 | 3,398 | 234 | 5,561 | 383 |
|  | 25 to 44 | 18,980 | 714 | 32,916 | 1,239 |
|  | 45 to 54 | 21,424 | 1,475 | 32,401 | 2,231 |
|  | 55 to 64 | 42,370 | 2,583 | 67,205 | 4,097 |
|  | 65 to 74 | 105,255 | 3,705 | 172,755 | 6,082 |
|  | 75 plus | 127,163 | 4,936 | 214,422 | 8,323 |
| Race-ethnicity | White | 202,814 | 2,343 | 335,012 | 3,870 |
|  | Black | 47,209 | 3,800 | 78,914 | 6,351 |
|  | Hispanic | 36,174 | 2,422 | 57,680 | 3,862 |
|  | Asian | 6,270 | 1,032 | 10,296 | 1,695 |
|  | Unknown | 27,088 | 1,576 | 44,773 | 2,605 |
| Sex | Female | 191,534 | 2,660 | 320,081 | 4,446 |
|  | Male | 128,021 | 1,963 | 206,594 | 3,168 |
| Region | Northeast | 39,112 | 2,563 | 64,396 | 4,219 |
|  | Midwest | 66,201 | 1,992 | 109,225 | 3,287 |
|  | South | 149,266 | 2,656 | 246,329 | 4,384 |
|  | West | 64,976 | 1,998 | 106,725 | 3,282 |
| Total |  | 319,555 | 2,329 | 526,675 | 3,839 |

Source: De-identified Optum Clinformatic® Data Mart.
Enrollees with full enrollment in a commercial health plan or Medicare during the year.

Among commercial insurance enrollees, hospital discharge rates with both first-listed and all-listed digestive disease diagnoses were higher compared with national data; however, availability of more recent data for commercial insurance enrollees (2020 vs. 2018) may limit comparability (Table 8). Like national data, hospital discharge rates (all-listed diagnoses) were higher among children compared with adolescents and the youngest adults and then increased with age and were higher among women compared with men. Optum data include information on hospital discharges among Asians that are not available in national data used in this paper. Among persons with known race-ethnicity, rates were highest among Blacks, followed by Whites, then Hispanics, and lowest among Asians. Hospital discharge rates were highest in the Northeast and South, intermediate in the Midwest, and lowest in the West.

**Table 8:** All Digestive Diseases: Hospital discharges with first-listed and all-listed diagnoses by age, race-ethnicity, sex and region among privately insured enrollees, 2020.

| Demographic Characteristics |  | First-Listed Diagnosis |  | All-Listed Diagnoses |  |
| --- | --- | --- | --- | --- | --- |
|  |  | Number of events | Event rate per 100,000 | Number of events | Event rate per 100,000 |
| Age (years) | 0 to 11 | 1,770 | 161 | 22,408 | 2,033 |
|  | 12 to 24 | 2,258 | 156 | 9,403 | 648 |
|  | 25 to 44 | 10,289 | 387 | 39,131 | 1,473 |
|  | 45 to 54 | 11,494 | 791 | 44,452 | 3,061 |
|  | 55 to 64 | 22,199 | 1,353 | 103,973 | 6,339 |
|  | 65 to 74 | 47,950 | 1,688 | 233,696 | 8,227 |
|  | 75 plus | 62,345 | 2,420 | 335,855 | 13,037 |
| Race-ethnicity | White | 105,032 | 1,213 | 514,339 | 5,941 |
|  | Black | 19,757 | 1,590 | 106,043 | 8,535 |
|  | Hispanic | 16,777 | 1,123 | 74,156 | 4,965 |
|  | Asian | 3,466 | 571 | 15,349 | 2,527 |
|  | Unknown | 13,273 | 772 | 79,031 | 4,597 |
| Sex | Female | 87,500 | 1,215 | 434,432 | 6,034 |
|  | Male | 70,805 | 1,086 | 354,486 | 5,436 |
| Region | Northeast | 20,735 | 1,359 | 101,404 | 6,644 |
|  | Midwest | 35,011 | 1,054 | 176,590 | 5,314 |
|  | South | 71,551 | 1,273 | 366,456 | 6,521 |
|  | West | 31,008 | 954 | 144,468 | 4,443 |
| Total |  | 158,305 | 1,154 | 788,918 | 5,750 |

Source: De-identified Optum Clinformatic® Data Mart.
Enrollees with full enrollment in a commercial health plan or Medicare during the year.

Like Optum data, Medicare data are at the person level and enables calculation of claims-based prevalence. Among 27 million age-eligible Medicare beneficiaries (2019), 7.8 million had a first-listed digestive disease diagnosis (claims-based prevalence of 29.1%) and 14 million had an all-listed diagnosis (claims-based prevalence of 53.1%) (Table 9). Prevalence (all-listed diagnoses) increased with age and was higher among women compared with men and Whites compared with Blacks. Prevalence was highest in the South, followed by the Northeast, then the Midwest, and lowest in the West. Medicare claims data were not utilized in The Burden of Digestive Diseases in the United States compendium so rate trends cannot be examined.(4)

**Table 9:** All Digestive Diseases: Claims-based prevalence with first-listed and all-listed diagnoses by age, race, sex and region among fee-for-service, age-eligible Medicare beneficiaries, 2019.

| Demographic Characteristics |  | Number of enrollees | First-Listed Diagnosis |  | All-Listed Diagnoses |  |
| --- | --- | --- | --- | --- | --- | --- |
|  |  |  | Number of enrollees with disease | Percent of enrollees with disease | Number of enrollees with disease | Percent of enrollees with disease |
| Age (years) | 65 to 69 | 8,066,320 | 2,262,980 | 28.1% | 4,016,480 | 49.8% |
|  | 70 to 74 | 6,936,940 | 2,068,880 | 29.8% | 3,674,040 | 53.0% |
|  | 75 to 79 | 4,894,060 | 1,494,340 | 30.5% | 2,686,880 | 54.9% |
|  | 80 to 84 | 3,335,340 | 985,700 | 29.6% | 1,847,460 | 55.4% |
|  | 85+ | 3,578,120 | 992,580 | 27.7% | 2,003,400 | 56.0% |
| Race | White | 22,023,800 | 6,446,940 | 29.3% | 11,797,960 | 53.6% |
|  | Black | 1,861,660 | 527,600 | 28.3% | 964,080 | 51.8% |
|  | Other | 2,925,320 | 829,940 | 28.4% | 1,466,220 | 50.1% |
| Sex | Female | 14,975,560 | 4,537,040 | 30.3% | 8,328,400 | 55.6% |
|  | Male | 11,835,220 | 3,267,440 | 27.6% | 5,899,860 | 49.9% |
| Region | Northeast | 4,748,700 | 1,443,740 | 30.4% | 2,549,620 | 53.7% |
|  | Midwest | 6,069,800 | 1,658,760 | 27.3% | 3,158,060 | 52.0% |
|  | South | 10,595,900 | 3,210,640 | 30.3% | 5,865,440 | 55.4% |
|  | West | 5,396,380 | 1,491,340 | 27.6% | 2,655,140 | 49.2% |
| Total |  | 26,810,780 | 7,804,480 | 29.1% | 14,228,260 | 53.1% |

Source: Centers for Medicare & Medicaid Services, 5% Denominator and Claims Files. Beneficiaries aged 65+ years with continuous and full Parts A and B enrollment and no Health Maintenance Organization enrollment during the year. Unweighted counts were multiplied by 20 to produce counts for this table.

Among age-eligible Medicare beneficiaries, there were an estimated 19 million ambulatory care visits with a first-listed diagnosis of a digestive disease and 43 million visits with a digestive disease diagnosis listed in any position (2019) (Table 10). The ambulatory care visit rate (per 100,000 population) was 69,119 with a first-listed diagnosis and 161,206 with an all-listed diagnosis. All-listed diagnosis rates increased with age until 85 years and were higher among women compared with men and Blacks compared with Whites. Rates were higher in the South and Northeast compared with the West and Midwest.

**Table 10:** All Digestive Diseases: Ambulatory care visits with first-listed and all-listed diagnoses by age, race, sex and region among fee-for-service, age-eligible Medicare beneficiaries, 2019.

| Demographic Characteristics |  | First-Listed Diagnosis |  | All-Listed Diagnoses |  |
| --- | --- | --- | --- | --- | --- |
|  |  | Number of events | Event rate per 100,000 | Number of events | Event rate per 100,000 |
| Age (years) | 65 to 69 | 5,348,520 | 66,307 | 11,581,900 | 143,583 |
|  | 70 to 74 | 4,978,480 | 71,768 | 11,005,700 | 158,654 |
|  | 75 to 79 | 3,683,640 | 75,268 | 8,447,660 | 172,610 |
|  | 80 to 84 | 2,386,820 | 71,562 | 5,910,200 | 177,199 |
|  | 85+ | 2,133,980 | 59,640 | 6,275,240 | 175,378 |
| Race | White | 15,277,320 | 69,367 | 35,437,700 | 160,906 |
|  | Black | 1,208,800 | 64,931 | 3,083,400 | 165,626 |
|  | Other | 2,045,320 | 69,918 | 4,699,600 | 160,653 |
| Sex | Female | 10,749,960 | 71,783 | 25,908,960 | 173,008 |
|  | Male | 7,781,480 | 65,749 | 17,311,740 | 146,273 |
| Region | Northeast | 3,573,040 | 75,242 | 8,092,200 | 170,409 |
|  | Midwest | 3,681,060 | 60,645 | 8,847,180 | 145,757 |
|  | South | 7,763,360 | 73,268 | 18,370,940 | 173,378 |
|  | West | 3,513,980 | 65,117 | 7,910,380 | 146,587 |
| Total |  | 18,531,440 | 69,119 | 43,220,700 | 161,206 |

Source: Centers for Medicare & Medicaid Services, 5% Denominator and Claims Files. Beneficiaries aged 65+ years with continuous and full Parts A and B enrollment and no Health Maintenance Organization enrollment during the year. Unweighted counts were multiplied by 20 to produce counts for this table.

Among age-eligible Medicare beneficiaries, there were an estimated 2.4 million emergency department visits with a first-listed diagnosis of a digestive disease and 7.1 million visits with a digestive disease diagnosis listed in any position (2019) (Table 11). The emergency department visit rate (per 100,000 population) was 9,135 with a first-listed diagnosis and 26,607 with an all-listed diagnosis. All-listed diagnosis rates increased with age and were higher among women compared with men and Blacks compared with Whites. Rates were highest in the South, followed by the Northeast and Midwest, and lowest in the West.

**Table 11:**
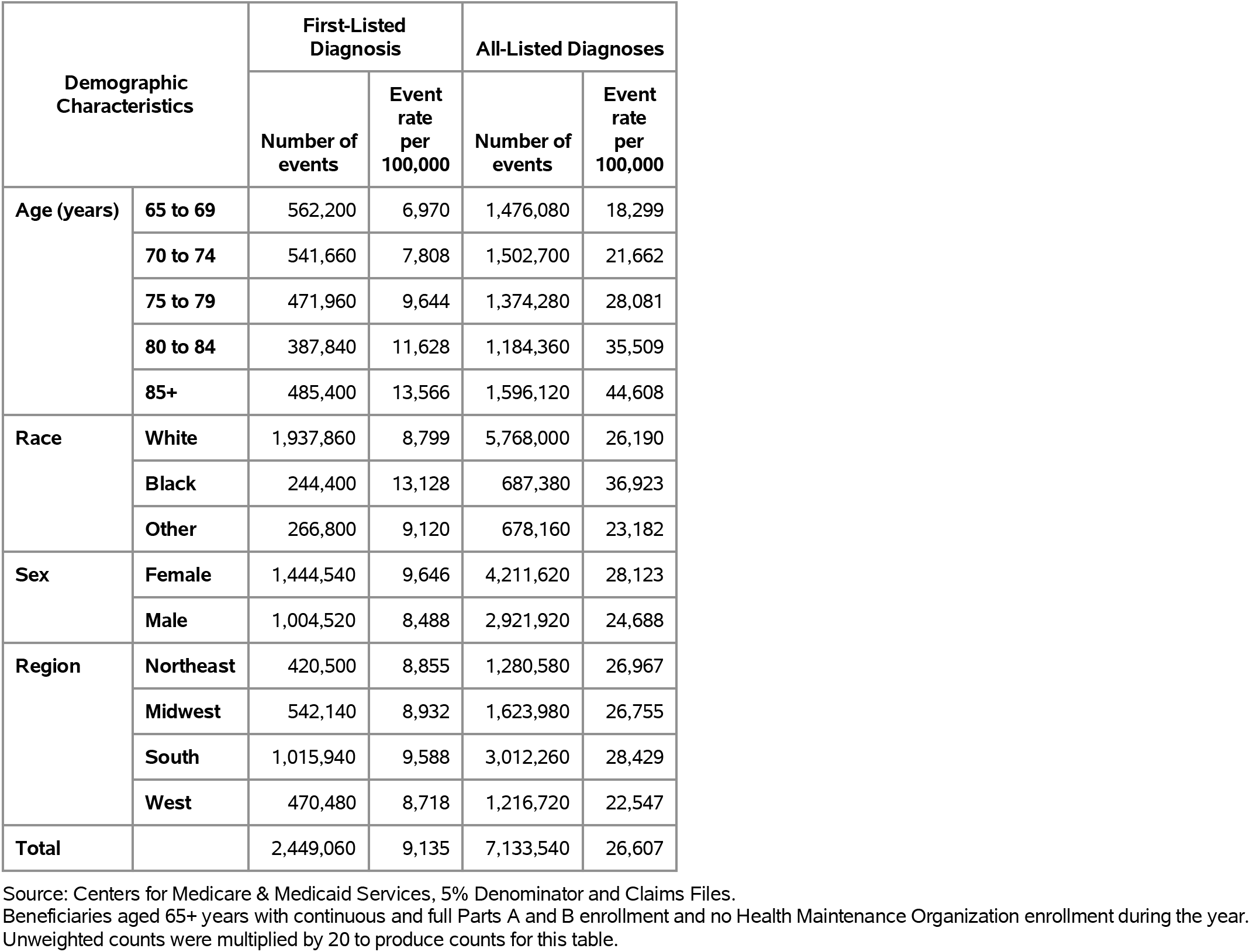
All Digestive Diseases: Emergency department visits with first-listed and all-listed diagnoses by age, race, sex and region among fee-for-service, age-eligible Medicare beneficiaries, 2019.

| Demographic Characteristics |  | First-Listed Diagnosis |  | All-Listed Diagnoses |  |
| --- | --- | --- | --- | --- | --- |
|  |  | Number of events | Event rate per 100,000 | Number of events | Event rate per 100,000 |
| Age (years) | 65 to 69 | 562,200 | 6,970 | 1,476,080 | 18,299 |
|  | 70 to 74 | 541,660 | 7,808 | 1,502,700 | 21,662 |
|  | 75 to 79 | 471,960 | 9,644 | 1,374,280 | 28,081 |
|  | 80 to 84 | 387,840 | 11,628 | 1,184,360 | 35,509 |
|  | 85+ | 485,400 | 13,566 | 1,596,120 | 44,608 |
| Race | White | 1,937,860 | 8,799 | 5,768,000 | 26,190 |
|  | Black | 244,400 | 13,128 | 687,380 | 36,923 |
|  | Other | 266,800 | 9,120 | 678,160 | 23,182 |
| Sex | Female | 1,444,540 | 9,646 | 4,211,620 | 28,123 |
|  | Male | 1,004,520 | 8,488 | 2,921,920 | 24,688 |
| Region | Northeast | 420,500 | 8,855 | 1,280,580 | 26,967 |
|  | Midwest | 542,140 | 8,932 | 1,623,980 | 26,755 |
|  | South | 1,015,940 | 9,588 | 3,012,260 | 28,429 |
|  | West | 470,480 | 8,718 | 1,216,720 | 22,547 |
| Total |  | 2,449,060 | 9,135 | 7,133,540 | 26,607 |

Source: Centers for Medicare & Medicaid Services, 5% Denominator and Claims Files. Beneficiaries aged 65+ years with continuous and full Parts A and B enrollment and no Health Maintenance Organization enrollment during the year. Unweighted counts were multiplied by 20 to produce counts for this table.

Among age-eligible Medicare beneficiaries, there were an estimated 915,000 hospital discharges with a first-listed diagnosis of a digestive disease and 17,000 discharges with a digestive disease diagnosis listed in any position (2019) (Table 12). The hospital discharge rate (per 100,000 population) was 3,413 with a first-listed diagnosis and 17,407 with an all-listed diagnosis. All-listed diagnosis rates increased with age and were higher among Blacks compared with Whites but differed little by sex. Rates were lower in the West compared with the South, Midwest, and Northeast.

**Table 12:** All Digestive Diseases: Hospital discharges with first-listed and all-listed diagnoses by age, race, sex and region among fee-for-service, age-eligible Medicare beneficiaries, 2019.

| Demographic Characteristics |  | First-Listed Diagnosis |  | All-Listed Diagnoses |  |
| --- | --- | --- | --- | --- | --- |
|  |  | Number of events | Event rate per 100,000 | Number of events | Event rate per 100,000 |
| Age (years) | 65 to 69 | 202,620 | 2,512 | 934,600 | 11,586 |
|  | 70 to 74 | 203,160 | 2,929 | 993,820 | 14,326 |
|  | 75 to 79 | 179,100 | 3,660 | 919,520 | 18,788 |
|  | 80 to 84 | 147,660 | 4,427 | 781,660 | 23,436 |
|  | 85+ | 182,500 | 5,100 | 1,037,280 | 28,990 |
| Race | White | 740,160 | 3,361 | 3,839,000 | 17,431 |
|  | Black | 82,120 | 4,411 | 407,000 | 21,862 |
|  | Other | 92,760 | 3,171 | 420,880 | 14,387 |
| Sex | Female | 518,640 | 3,463 | 2,634,420 | 17,591 |
|  | Male | 396,400 | 3,349 | 2,032,460 | 17,173 |
| Region | Northeast | 169,840 | 3,577 | 857,580 | 18,059 |
|  | Midwest | 212,360 | 3,499 | 1,097,960 | 18,089 |
|  | South | 373,640 | 3,526 | 1,952,780 | 18,430 |
|  | West | 159,200 | 2,950 | 758,560 | 14,057 |
| Total |  | 915,040 | 3,413 | 4,666,880 | 17,407 |

Source: Centers for Medicare & Medicaid Services, 5% Denominator and Claims Files. Beneficiaries aged 65+ years with continuous and full Parts A and B enrollment and no Health Maintenance Organization enrollment during the year. Unweighted counts were multiplied by 20 to produce counts for this table.

Gastroesophageal reflux disease contributed to 26.7 million ambulatory visits (2015) (Table 13). Ambulatory care visit rates (all-listed diagnoses) were higher among children compared with adolescents and the youngest adults and then increased with age. Age-adjusted rates were higher among women compared with men, Blacks compared with Whites, and Hispanics compared with non-Hispanics.

**Table 13:** Gastroesophageal Reflux Disease: Ambulatory care visits with first-listed and all-listed diagnoses by age, race, ethnicity, and sex in the United States, 2015.

| Demographic Characteristics |  | First-Listed Diagnosis |  | All-Listed Diagnoses |  |
| --- | --- | --- | --- | --- | --- |
|  |  | Number in thousands | Rate per 100,000 | Number in thousands | Rate per 100,000 |
| Age (years) | 0 to 11 | 362 | 745 | 1,195 | 2,457 |
|  | 12 to 24 | 541 | 964 | 1,146 | 2,042 |
|  | 25 to 44 | 1,065 | 1,260 | 3,463 | 4,098 |
|  | 45 to 54 | 952 | 2,210 | 4,592 | 10,657 |
|  | 55 to 64 | 985 | 2,414 | 5,635 | 13,810 |
|  | 65 to 74 | 974 | 3,539 | 5,945 | 21,597 |
|  | 75 plus | 562 | 2,780 | 4,735 | 23,432 |
| Race | White | 4,387 | 1,638 | 21,392 | 7,488 |
|  | Black | 797 | 1,835 | 3,561 | 8,328 |
|  | Other | 257 | 1,087 | 1,759 | 7,906 |
| Ethnicity | Hispanic | 912 | 2,232 | 4,242 | 10,206 |
|  | Not Hispanic | 4,529 | 1,568 | 22,469 | 7,340 |
| Sex | Female | 3,572 | 2,009 | 16,082 | 8,673 |
|  | Male | 1,869 | 1,175 | 10,629 | 6,421 |
| Total |  | 5,441 | 1,609 | 26,711 | 7,578 |

Source: National Ambulatory Medical Care Survey (3-year average, 2014-2016). Rates for age groups are unadjusted and all other rates are age-adjusted.

Gastroesophageal reflux disease contributed to 9.5 million emergency department visits in 2018 (Table 14). Emergency department visit rates (all-listed diagnoses) increased with age. Age-adjusted rates were higher among women compared with men.

**Table 14:** Gastroesophageal Reflux Disease: Emergency department visits with first-listed and all-listed diagnoses by age and sex in the United States, 2018.

| Demographic Characteristics |  | First-Listed Diagnosis |  | All-Listed Diagnoses |  |
| --- | --- | --- | --- | --- | --- |
|  |  | Number in thousands | Rate per 100,000 | Number in thousands | Rate per 100,000 |
| Age (years) | 0 to 11 | 44 | 91 | 226 | 466 |
|  | 12 to 24 | 60 | 109 | 326 | 589 |
|  | 25 to 44 | 123 | 141 | 1,447 | 1,663 |
|  | 45 to 54 | 61 | 147 | 1,335 | 3,208 |
|  | 55 to 64 | 58 | 138 | 1,791 | 4,236 |
|  | 65 to 74 | 45 | 147 | 1,841 | 6,037 |
|  | 75 plus | 46 | 211 | 2,518 | 11,478 |
| Sex | Female | 245 | 144 | 5,657 | 2,913 |
|  | Male | 192 | 118 | 3,826 | 2,228 |
| Total |  | 438 | 131 | 9,484 | 2,582 |

Source: Healthcare Cost and Utilization Project Nationwide Emergency Department Sample. Rates for age groups are unadjusted and all other rates are age-adjusted.

Gastroesophageal reflux disease contributed to 6.2 million hospital discharges in 2018 (Table 15). Hospital discharge rates (all-listed diagnoses) increased with age. Age-adjusted rates were higher among women compared with men, Blacks compared with Whites, and non-Hispanics compared with Hispanics. Between 2004 and 2018, age-adjusted hospital discharge rates (per 100,000) with an all-listed diagnosis increased by 50% from 1,086 to 1628.(4,5)

**Table 15:** Gastroesophageal Reflux Disease: Hospital discharges with first-listed and all-listed diagnoses by age, race, ethnicity, and sex in the United States, 2018.

| Demographic Characteristics |  | First-Listed Diagnosis |  | All-Listed Diagnoses |  |
| --- | --- | --- | --- | --- | --- |
|  |  | Number in thousands | Rate per 100,000 | Number in thousands | Rate per 100,000 |
| Age (years) | 0 to 11 | 6 | 12 | 78 | 162 |
|  | 12 to 24 | 2 | 4 | 104 | 188 |
|  | 25 to 44 | 10 | 12 | 628 | 722 |
|  | 45 to 54 | 11 | 27 | 704 | 1,691 |
|  | 55 to 64 | 17 | 40 | 1,233 | 2,917 |
|  | 65 to 74 | 16 | 52 | 1,510 | 4,953 |
|  | 75 plus | 20 | 90 | 1,967 | 8,967 |
| Race | White | 65 | 22 | 5,207 | 1,653 |
|  | Black | 13 | 30 | 800 | 1,841 |
|  | Other | 5 | 20 | 274 | 1,112 |
| Ethnicity | Hispanic | 10 | 21 | 476 | 1,103 |
|  | Not Hispanic | 73 | 23 | 5,806 | 1,726 |
| Sex | Female | 41 | 21 | 3,599 | 1,759 |
|  | Male | 41 | 24 | 2,626 | 1,489 |
| Total |  | 82 | 22 | 6,225 | 1,628 |

Source: Healthcare Cost and Utilization Project National Inpatient Sample. Rates for age groups are unadjusted and all other rates are age-adjusted.

Gastroesophageal reflux disease contributed to 12,000 deaths in 2019 (Table 16). Mortality rates (underlying or other cause) were higher among children compared with adolescents and the youngest adults and then increased with age. Age-adjusted mortality rates were higher among men, Whites, and non-Hispanics. Between 2004 and 2019, age-adjusted mortality rates (per 100,000) with gastroesophageal reflux disease as underlying or other cause increased by 7% from 2.7 to 2.9.(4)

**Table 16:** Gastroesophageal Reflux Disease: Deaths with underlying or underlying/other cause and lifetime years of life lost by age, race, ethnicity, and sex in the United States, 2019.

| Demographic Characteristics |  | Underlying Cause |  |  | Underlying/Other Cause |  |  |
| --- | --- | --- | --- | --- | --- | --- | --- |
|  |  | Number of Deaths | Rate per 100,000 | Years of potential life lost in thousands | Number of Deaths | Rate per 100,000 | Years of potential life lost in thousands |
| Age (years) | 0 to 11 | 10 | 0.0 | 0.8 | 43 | 0.1 | 3.2 |
|  | 12 to 24 | 2 | 0.0 | 0.1 | 27 | 0.0 | 1.6 |
|  | 25 to 44 | 33 | 0.0 | 1.4 | 194 | 0.2 | 8.2 |
|  | 45 to 54 | 45 | 0.1 | 1.4 | 449 | 1.1 | 13.8 |
|  | 55 to 64 | 117 | 0.3 | 2.7 | 1,269 | 3.0 | 28.8 |
|  | 65 to 74 | 194 | 0.6 | 3.0 | 2,216 | 7.0 | 34.4 |
|  | 75 plus | 824 | 3.7 | 4.8 | 7,515 | 33.3 | 47.8 |
| Race | White | 1,107 | 0.3 | 12.3 | 10,412 | 3.0 | 118.8 |
|  | Black | 81 | 0.2 | 1.3 | 1,008 | 2.5 | 14.9 |
|  | Other | 37 | 0.2 | 0.6 | 293 | 1.2 | 4.1 |
| Ethnicity | Hispanic | 64 | 0.2 | 1.2 | 524 | 1.4 | 8.5 |
|  | Not Hispanic | 1,161 | 0.3 | 13.0 | 11,189 | 3.0 | 129.3 |
| Sex | Female | 603 | 0.3 | 6.7 | 6,312 | 2.7 | 70.0 |
|  | Male | 622 | 0.4 | 7.5 | 5,401 | 3.1 | 67.9 |
| Total |  | 1,225 | 0.3 | 14.2 | 11,713 | 2.9 | 137.9 |

Source: Vital Statistics of the United States.
Rates for age groups are unadjusted and all other rates are age-adjusted.
Years of potential life lost is based on U.S. life table 2006-2018 estimates of life expectancy at age of death.

Among privately insured enrollees, the claims-based prevalence of gastroesophageal reflux disease (based on all-listed diagnoses) was 11.1% (Table 17). Prevalence was similar among children compared with adolescents and the youngest adults and then increased with age. It was higher among women. It was highest among Blacks, followed by Whites and Hispanics, and lowest among Asians. It was highest in the South, followed by the Northeast, then the Midwest, and lowest in the West.

**Table 17:** Gastroesophageal Reflux Disease: Claims-based prevalence with first-listed and all-listed diagnoses by age, race-ethnicity, sex and region among privately insured enrollees, 2020.

| Demographic Characteristics |  | Number of enrollees | First-Listed Diagnosis |  | All-Listed Diagnoses |  |
| --- | --- | --- | --- | --- | --- | --- |
|  |  |  | Number of enrollees with disease | Percent of enrollees with disease | Number of enrollees with disease | Percent of enrollees with disease |
| Age (years) | 0 to 11 | 1,101,978 | 9,567 | 0.9% | 20,842 | 1.9% |
|  | 12 to 24 | 1,451,892 | 10,511 | 0.7% | 25,914 | 1.8% |
|  | 25 to 44 | 2,656,672 | 43,031 | 1.6% | 132,690 | 5.0% |
|  | 45 to 54 | 1,452,320 | 36,937 | 2.5% | 145,474 | 10.0% |
|  | 55 to 64 | 1,640,216 | 52,413 | 3.2% | 241,952 | 14.8% |
|  | 65 to 74 | 2,840,615 | 103,549 | 3.6% | 492,699 | 17.3% |
|  | 75 plus | 2,576,241 | 80,437 | 3.1% | 470,137 | 18.2% |
| Race-ethnicity | White | 8,657,702 | 219,262 | 2.5% | 1,039,266 | 12.0% |
|  | Black | 1,242,477 | 37,054 | 3.0% | 177,232 | 14.3% |
|  | Hispanic | 1,493,457 | 37,219 | 2.5% | 150,528 | 10.1% |
|  | Asian | 607,284 | 10,905 | 1.8% | 40,131 | 6.6% |
|  | Unknown | 1,719,014 | 32,005 | 1.9% | 122,551 | 7.1% |
| Sex | Female | 7,199,363 | 204,551 | 2.8% | 912,394 | 12.7% |
|  | Male | 6,520,571 | 131,894 | 2.0% | 617,314 | 9.5% |
| Region | Northeast | 1,526,275 | 44,716 | 2.9% | 181,214 | 11.9% |
|  | Midwest | 3,322,846 | 65,408 | 2.0% | 339,807 | 10.2% |
|  | South | 5,619,435 | 165,939 | 3.0% | 738,811 | 13.1% |
|  | West | 3,251,378 | 60,382 | 1.9% | 269,876 | 8.3% |
| Total |  | 13,719,934 | 336,445 | 2.5% | 1,529,708 | 11.1% |

Source: De-identified Optum Clinformatic® Data Mart.
Enrollees with full enrollment in a commercial health plan or Medicare during the year.

Among commercial insurance enrollees, ambulatory care visit rates with gastroesophageal reflux disease (all-listed diagnoses) were higher among children compared with adolescents and the youngest adults and then increased with age and were higher among women compared with men (Table 18). Among persons with known race-ethnicity, rates were highest among Blacks, followed by Whites, then Hispanics, and lowest among Asians. Rates were highest in the South, followed by the Northeast, then the Midwest, and lowest in the West.

**Table 18:**
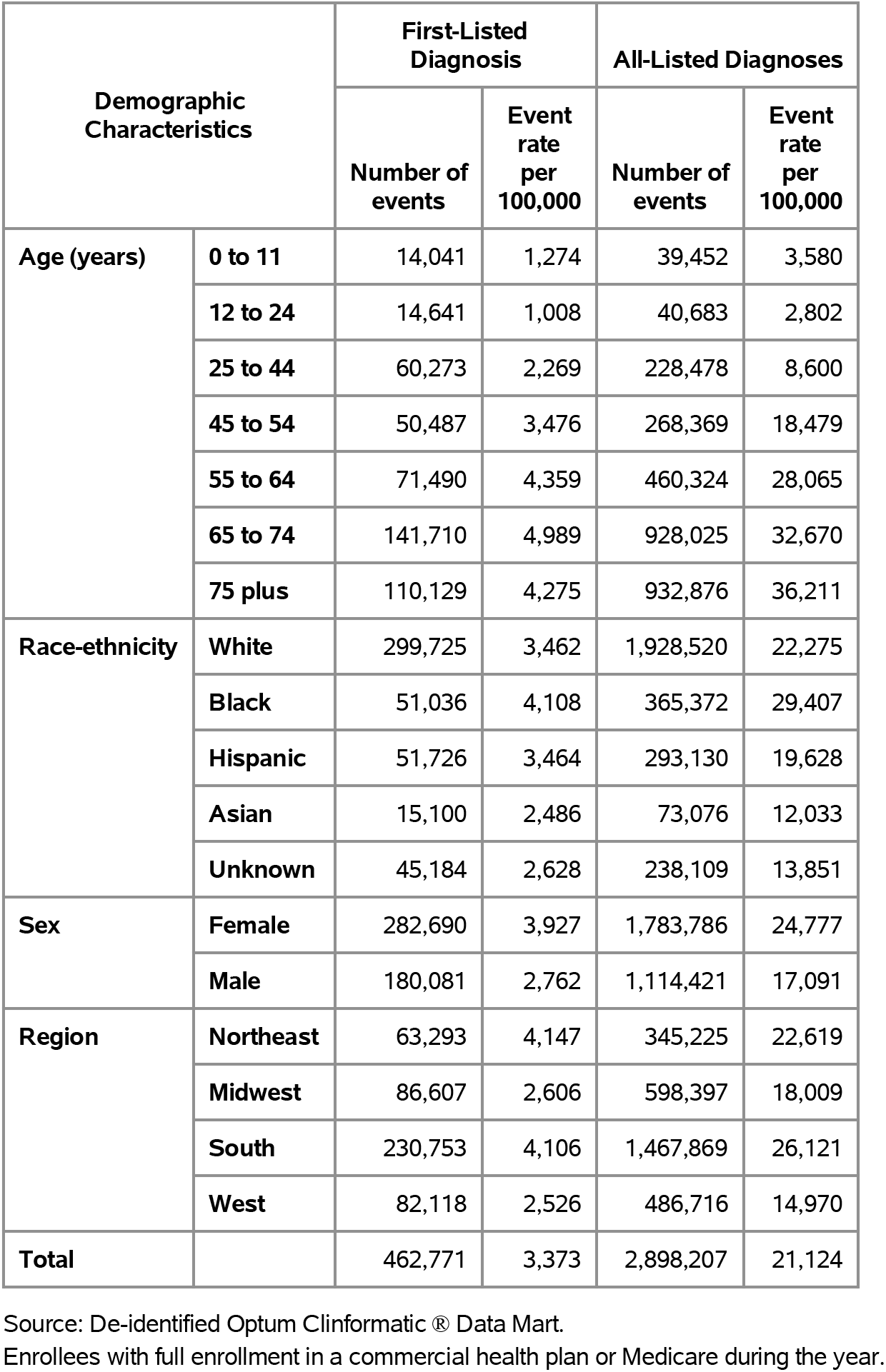
Gastroesophageal Reflux Disease: Ambulatory care visits with first-listed and all-listed diagnoses by age, race-ethnicity, sex and region among privately insured enrollees, 2020.

| Demographic Characteristics |  | First-Listed Diagnosis |  | All-Listed Diagnoses |  |
| --- | --- | --- | --- | --- | --- |
|  |  | Number of events | Event rate per 100,000 | Number of events | Event rate per 100,000 |
| Age (years) | 0 to 11 | 14,041 | 1,274 | 39,452 | 3,580 |
|  | 12 to 24 | 14,641 | 1,008 | 40,683 | 2,802 |
|  | 25 to 44 | 60,273 | 2,269 | 228,478 | 8,600 |
|  | 45 to 54 | 50,487 | 3,476 | 268,369 | 18,479 |
|  | 55 to 64 | 71,490 | 4,359 | 460,324 | 28,065 |
|  | 65 to 74 | 141,710 | 4,989 | 928,025 | 32,670 |
|  | 75 plus | 110,129 | 4,275 | 932,876 | 36,211 |
| Race-ethnicity | White | 299,725 | 3,462 | 1,928,520 | 22,275 |
|  | Black | 51,036 | 4,108 | 365,372 | 29,407 |
|  | Hispanic | 51,726 | 3,464 | 293,130 | 19,628 |
|  | Asian | 15,100 | 2,486 | 73,076 | 12,033 |
|  | Unknown | 45,184 | 2,628 | 238,109 | 13,851 |
| Sex | Female | 282,690 | 3,927 | 1,783,786 | 24,777 |
|  | Male | 180,081 | 2,762 | 1,114,421 | 17,091 |
| Region | Northeast | 63,293 | 4,147 | 345,225 | 22,619 |
|  | Midwest | 86,607 | 2,606 | 598,397 | 18,009 |
|  | South | 230,753 | 4,106 | 1,467,869 | 26,121 |
|  | West | 82,118 | 2,526 | 486,716 | 14,970 |
| Total |  | 462,771 | 3,373 | 2,898,207 | 21,124 |

Source: De-identified Optum Clinformatic® Data Mart.
Enrollees with full enrollment in a commercial health plan or Medicare during the year.

Among commercial insurance enrollees, emergency department visit rates with gastroesophageal reflux disease (all-listed diagnoses) increased with age and were higher among women compared with men (Table 19). Among persons with known race-ethnicity, rates were highest among Blacks, followed by Whites, then Hispanics, and lowest among Asians. Rates were highest in the Northeast, followed by the Midwest and South, and lowest in the West.

**Table 19:** Gastroesophageal Reflux Disease: Emergency department visits with first-listed and all-listed diagnoses by age, race-ethnicity, sex and region among privately insured enrollees, 2020.

| Demographic Characteristics |  | First-Listed Diagnosis |  | All-Listed Diagnoses |  |
| --- | --- | --- | --- | --- | --- |
|  |  | Number of events | Event rate per 100,000 | Number of events | Event rate per 100,000 |
| Age (years) | 0 to 11 | 32 | 3 | 73 | 7 |
|  | 12 to 24 | 139 | 10 | 371 | 26 |
|  | 25 to 44 | 688 | 26 | 2,394 | 90 |
|  | 45 to 54 | 536 | 37 | 2,071 | 143 |
|  | 55 to 64 | 907 | 55 | 3,921 | 239 |
|  | 65 to 74 | 1,776 | 63 | 7,832 | 276 |
|  | 75 plus | 1,882 | 73 | 8,846 | 343 |
| Race-ethnicity | White | 3,595 | 42 | 16,654 | 192 |
|  | Black | 1,051 | 85 | 4,064 | 327 |
|  | Hispanic | 662 | 44 | 2,141 | 143 |
|  | Asian | 105 | 17 | 467 | 77 |
|  | Unknown | 547 | 32 | 2,182 | 127 |
| Sex | Female | 3,576 | 50 | 15,940 | 221 |
|  | Male | 2,384 | 37 | 9,568 | 147 |
| Region | Northeast | 743 | 49 | 3,910 | 256 |
|  | Midwest | 1,249 | 38 | 6,771 | 204 |
|  | South | 3,055 | 54 | 11,272 | 201 |
|  | West | 913 | 28 | 3,555 | 109 |
| Total |  | 5,960 | 43 | 25,508 | 186 |

Source: De-identified Optum Clinformatic® Data Mart.
Enrollees with full enrollment in a commercial health plan or Medicare during the year.

Among commercial insurance enrollees, hospital discharge rates with gastroesophageal reflux disease (all-listed diagnoses) increased with age and were higher among women compared with men (Table 20). Among persons with known race-ethnicity, rates were highest among Blacks, followed by Whites, then Hispanics, and lowest among Asians. Rates were highest in the South, followed by the Northeast, then the Midwest, and lowest in the West.

**Table 20:** Gastroesophageal Reflux Disease: Hospital discharges with first-listed and all-listed diagnoses by age, race-ethnicity, sex and region among privately insured enrollees, 2020.

| Demographic Characteristics |  | First-Listed Diagnosis |  | All-Listed Diagnoses |  |
| --- | --- | --- | --- | --- | --- |
|  |  | Number of events | Event rate per 100,000 | Number of events | Event rate per 100,000 |
| Age (years) | 0 to 11 | 55 | 5 | 933 | 85 |
|  | 12 to 24 | 25 | 2 | 1,570 | 108 |
|  | 25 to 44 | 108 | 4 | 11,259 | 424 |
|  | 45 to 54 | 143 | 10 | 17,165 | 1,182 |
|  | 55 to 64 | 405 | 25 | 44,429 | 2,709 |
|  | 65 to 74 | 750 | 26 | 99,583 | 3,506 |
|  | 75 plus | 1,130 | 44 | 143,001 | 5,551 |
| Race-ethnicity | White | 1,709 | 20 | 221,785 | 2,562 |
|  | Black | 385 | 31 | 43,846 | 3,529 |
|  | Hispanic | 247 | 17 | 24,559 | 1,644 |
|  | Asian | 42 | 7 | 4,729 | 779 |
|  | Unknown | 233 | 14 | 23,021 | 1,339 |
| Sex | Female | 1,390 | 19 | 185,026 | 2,570 |
|  | Male | 1,226 | 19 | 132,914 | 2,038 |
| Region | Northeast | 312 | 20 | 39,742 | 2,604 |
|  | Midwest | 554 | 17 | 75,875 | 2,283 |
|  | South | 1,185 | 21 | 152,116 | 2,707 |
|  | West | 565 | 17 | 50,207 | 1,544 |
| Total |  | 2,616 | 19 | 317,940 | 2,317 |

Source: De-identified Optum Clinformatic® Data Mart.
Enrollees with full enrollment in a commercial health plan or Medicare during the year.

Among Medicare beneficiaries, the claims-based prevalence of gastroesophageal reflux disease (based on all-listed diagnoses) was 23.9% (Table 21). Prevalence increased with age until the oldest age group and was higher among women and Whites. It was highest in the South, followed by the Midwest and Northeast, and lowest in the West.

**Table 21:** Gastroesophageal Reflux Disease: Claims-based prevalence with first-listed and all-listed diagnoses by age, race, sex and region among fee-for-service, age-eligible Medicare beneficiaries, 2019.

| Demographic Characteristics |  | Number of enrollees | First-Listed Diagnosis |  | All-Listed Diagnoses |  |
| --- | --- | --- | --- | --- | --- | --- |
|  |  |  | Number of enrollees with disease | Percent of enrollees with disease | Number of enrollees with disease | Percent of enrollees with disease |
| Age (years) | 65 to 69 | 8,066,320 | 392,040 | 4.9% | 1,659,120 | 20.6% |
|  | 70 to 74 | 6,936,940 | 368,140 | 5.3% | 1,629,540 | 23.5% |
|  | 75 to 79 | 4,894,060 | 268,220 | 5.5% | 1,273,780 | 26.0% |
|  | 80 to 84 | 3,335,340 | 174,080 | 5.2% | 893,240 | 26.8% |
|  | 85+ | 3,578,120 | 155,280 | 4.3% | 940,280 | 26.3% |
| Race | White | 22,023,800 | 1,127,640 | 5.1% | 5,374,440 | 24.4% |
|  | Black | 1,861,660 | 86,000 | 4.6% | 424,540 | 22.8% |
|  | Other | 2,925,320 | 144,120 | 4.9% | 596,980 | 20.4% |
| Sex | Female | 14,975,560 | 862,780 | 5.8% | 3,926,880 | 26.2% |
|  | Male | 11,835,220 | 494,980 | 4.2% | 2,469,080 | 20.9% |
| Region | Northeast | 4,748,700 | 253,520 | 5.3% | 1,099,980 | 23.2% |
|  | Midwest | 6,069,800 | 256,180 | 4.2% | 1,429,680 | 23.6% |
|  | South | 10,595,900 | 614,400 | 5.8% | 2,851,060 | 26.9% |
|  | West | 5,396,380 | 233,660 | 4.3% | 1,015,240 | 18.8% |
| Total |  | 26,810,780 | 1,357,760 | 5.1% | 6,395,960 | 23.9% |

Source: Centers for Medicare & Medicaid Services, 5% Denominator and Claims Files. Beneficiaries aged 65+ years with continuous and full Parts A and B enrollment and no Health Maintenance Organization enrollment during the year. Unweighted counts were multiplied by 20 to produce counts for this table.

Among Medicare beneficiaries, ambulatory care visit rates with gastroesophageal reflux disease (all-listed diagnoses) increased with age and were higher among women compared with men and Blacks compared with Whites (Table 22). Rates were highest in the South, followed by the Northeast, then the Midwest, and lowest in the West.

**Table 22:** Gastroesophageal Reflux Disease: Ambulatory care visits with first-listed and all-listed diagnoses by age, race, sex and region among fee-for-service, age-eligible Medicare beneficiaries, 2019.

| Demographic Characteristics |  | First-Listed Diagnosis |  | All-Listed Diagnoses |  |
| --- | --- | --- | --- | --- | --- |
|  |  | Number of events | Event rate per 100,000 | Number of events | Event rate per 100,000 |
| Age (years) | 65 to 69 | 557,060 | 6,906 | 3,213,040 | 39,833 |
|  | 70 to 74 | 532,500 | 7,676 | 3,213,460 | 46,324 |
|  | 75 to 79 | 378,820 | 7,740 | 2,518,320 | 51,457 |
|  | 80 to 84 | 247,340 | 7,416 | 1,801,280 | 54,006 |
|  | 85+ | 234,320 | 6,549 | 1,979,600 | 55,325 |
| Race | White | 1,606,080 | 7,292 | 10,517,200 | 47,754 |
|  | Black | 130,000 | 6,983 | 930,960 | 50,007 |
|  | Other | 213,960 | 7,314 | 1,277,540 | 43,672 |
| Sex | Female | 1,259,740 | 8,412 | 8,152,640 | 54,440 |
|  | Male | 690,300 | 5,833 | 4,573,060 | 38,639 |
| Region | Northeast | 371,400 | 7,821 | 2,241,740 | 47,207 |
|  | Midwest | 348,820 | 5,747 | 2,533,080 | 41,733 |
|  | South | 894,920 | 8,446 | 6,014,200 | 56,760 |
|  | West | 334,900 | 6,206 | 1,936,680 | 35,889 |
| Total |  | 1,950,040 | 7,273 | 12,725,700 | 47,465 |

Source: Centers for Medicare & Medicaid Services, 5% Denominator and Claims Files. Beneficiaries aged 65+ years with continuous and full Parts A and B enrollment and no Health Maintenance Organization enrollment during the year. Unweighted counts were multiplied by 20 to produce counts for this table.

Among Medicare beneficiaries, emergency department visit rates with gastroesophageal reflux disease (all-listed diagnoses) increased with age and were higher among women compared with men and Blacks compared with Whites (Table 23). Rates were lower in the West compared with the South, Midwest, and Northeast.

**Table 23:** Gastroesophageal Reflux Disease: Emergency department visits with first-listed and all-listed diagnoses by age, race, sex and region among fee-for-service, age-eligible Medicare beneficiaries, 2019.

| Demographic Characteristics |  | First-Listed Diagnosis |  | All-Listed Diagnoses |  |
| --- | --- | --- | --- | --- | --- |
|  |  | Number of events | Event rate per 100,000 | Number of events | Event rate per 100,000 |
| Age (years) | 65 to 69 | 13,720 | 170 | 515,880 | 6,395 |
|  | 70 to 74 | 12,660 | 183 | 562,800 | 8,113 |
|  | 75 to 79 | 11,500 | 235 | 537,080 | 10,974 |
|  | 80 to 84 | 9,880 | 296 | 462,380 | 13,863 |
|  | 85+ | 11,580 | 324 | 623,620 | 17,429 |
| Race | White | 45,860 | 208 | 2,251,100 | 10,221 |
|  | Black | 7,660 | 411 | 247,140 | 13,275 |
|  | Other | 5,820 | 199 | 203,520 | 6,957 |
| Sex | Female | 35,000 | 234 | 1,657,340 | 11,067 |
|  | Male | 24,340 | 206 | 1,044,420 | 8,825 |
| Region | Northeast | 9,040 | 190 | 475,160 | 10,006 |
|  | Midwest | 14,360 | 237 | 653,820 | 10,772 |
|  | South | 26,040 | 246 | 1,200,480 | 11,330 |
|  | West | 9,900 | 183 | 372,300 | 6,899 |
| Total |  | 59,340 | 221 | 2,701,760 | 10,077 |

Source: Centers for Medicare & Medicaid Services, 5% Denominator and Claims Files. Beneficiaries aged 65+ years with continuous and full Parts A and B enrollment and no Health Maintenance Organization enrollment during the year. Unweighted counts were multiplied by 20 to produce counts for this table.

Among Medicare beneficiaries, hospital discharge rates with gastroesophageal reflux disease (all-listed diagnoses) increased with age and were higher among women compared with men and Blacks compared with Whites (Table 24). Rates were lower in the West compared with the South, Midwest, and Northeast.

**Table 24:** Gastroesophageal Reflux Disease: Hospital discharges with first-listed and all-listed diagnoses by age, race, sex and region among fee-for-service, age-eligible Medicare beneficiaries, 2019.

| Demographic Characteristics |  | First-Listed Diagnosis |  | All-Listed Diagnoses |  |
| --- | --- | --- | --- | --- | --- |
|  |  | Number of events | Event rate per 100,000 | Number of events | Event rate per 100,000 |
| Age (years) | 65 to 69 | 3,560 | 44 | 431,460 | 5,349 |
|  | 70 to 74 | 3,940 | 57 | 484,000 | 6,977 |
|  | 75 to 79 | 3,580 | 73 | 444,260 | 9,078 |
|  | 80 to 84 | 3,120 | 94 | 362,980 | 10,883 |
|  | 85+ | 4,240 | 118 | 468,520 | 13,094 |
| Race | White | 14,700 | 67 | 1,859,960 | 8,445 |
|  | Black | 2,020 | 109 | 167,660 | 9,006 |
|  | Other | 1,720 | 59 | 163,600 | 5,593 |
| Sex | Female | 10,220 | 68 | 1,293,460 | 8,637 |
|  | Male | 8,220 | 69 | 897,760 | 7,585 |
| Region | Northeast | 3,440 | 72 | 391,460 | 8,244 |
|  | Midwest | 4,400 | 72 | 539,980 | 8,896 |
|  | South | 7,200 | 68 | 951,340 | 8,978 |
|  | West | 3,400 | 63 | 308,440 | 5,716 |
| Total |  | 18,440 | 69 | 2,191,220 | 8,173 |

Source: Centers for Medicare & Medicaid Services, 5% Denominator and Claims Files. Beneficiaries aged 65+ years with continuous and full Parts A and B enrollment and no Health Maintenance Organization enrollment during the year. Unweighted counts were multiplied by 20 to produce counts for this table.

Peptic ulcer disease contributed to 1.4 million ambulatory visits (2015) (Table 25). Ambulatory care visits were uncommon during childhood and then rates (all-listed diagnoses) increased with age until the oldest age group. Age-adjusted ambulatory care visit rates were higher among women compared with men, Blacks compared with Whites, and Hispanics compared with non-Hispanics.

**Table 25:** Peptic Ulcer Disease: Ambulatory care visits with first-listed and all-listed diagnoses by age, race, ethnicity, and sex in the United States, 2015.

| Demographic Characteristics |  | First-Listed Diagnosis |  | All-Listed Diagnoses |  |
| --- | --- | --- | --- | --- | --- |
|  |  | Number in thousands | Rate per 100,000 | Number in thousands | Rate per 100,000 |
| Age (years) | 0 to 11 | . | . | . | . |
|  | 12 to 24 | 5 | 8 | 32 | 56 |
|  | 25 to 44 | 85 | 101 | 128 | 152 |
|  | 45 to 54 | 43 | 100 | 230 | 533 |
|  | 55 to 64 | 105 | 257 | 236 | 577 |
|  | 65 to 74 | 155 | 563 | 499 | 1,812 |
|  | 75 plus | 38 | 189 | 241 | 1,191 |
| Race | White | 235 | 92 | 965 | 329 |
|  | Black | 154 | 319 | 263 | 588 |
|  | Other | 42 | 188 | 137 | 548 |
| Ethnicity | Hispanic | 60 | 157 | 333 | 871 |
|  | Not Hispanic | 371 | 113 | 1,032 | 331 |
| Sex | Female | 288 | 148 | 779 | 404 |
|  | Male | 144 | 83 | 586 | 339 |
| Total |  | 431 | 117 | 1,365 | 372 |

Source: National Ambulatory Medical Care Survey (3-year average, 2014-2016). Rates for age groups are unadjusted and all other rates are age-adjusted.

Peptic ulcer disease contributed to 471,000 emergency department visits in 2018 (Table 26). Emergency department visit rates increased with age. Age-adjusted emergency department visit rates were higher among men compared with women.

**Table 26:** Peptic Ulcer Disease: Emergency department visits with first-listed and all-listed diagnoses by age and sex in the United States, 2018.

| Demographic Characteristics |  | First-Listed Diagnosis |  | All-Listed Diagnoses |  |
| --- | --- | --- | --- | --- | --- |
|  |  | Number in thousands | Rate per 100,000 | Number in thousands | Rate per 100,000 |
| Age (years) | 0 to 11 | 0 | 1 | 1 | 2 |
|  | 12 to 24 | 6 | 10 | 12 | 22 |
|  | 25 to 44 | 26 | 30 | 65 | 75 |
|  | 45 to 54 | 22 | 53 | 63 | 152 |
|  | 55 to 64 | 33 | 77 | 94 | 222 |
|  | 65 to 74 | 34 | 113 | 100 | 328 |
|  | 75 plus | 47 | 213 | 135 | 615 |
| Sex | Female | 83 | 42 | 240 | 121 |
|  | Male | 84 | 48 | 231 | 132 |
| Total |  | 168 | 45 | 471 | 126 |

Source: Healthcare Cost and Utilization Project Nationwide Emergency Department Sample. Rates for age groups are unadjusted and all other rates are age-adjusted.

Peptic ulcer disease contributed to 439,000 hospital discharges in 2018 (Table 27). Hospital discharge rates increased with age. Hospital discharge rates were higher among men compared with women, Blacks compared with Whites, and non-Hispanics compared with Hispanics. Between 2004 and 2018, age-adjusted hospital discharge rates (per 100,000) with an all-listed diagnosis decreased by 31% from 166 to 114.(4)

**Table 27:** Peptic Ulcer Disease: Hospital discharges with first-listed and all-listed diagnoses by age, race, ethnicity, and sex in the United States, 2018.

| Demographic Characteristics |  | First-Listed Diagnosis |  | All-Listed Diagnoses |  |
| --- | --- | --- | --- | --- | --- |
|  |  | Number in thousands | Rate per 100,000 | Number in thousands | Rate per 100,000 |
| Age (years) | 0 to 11 | 0 | 0 | 1 | 2 |
|  | 12 to 24 | 2 | 4 | 6 | 10 |
|  | 25 to 44 | 15 | 17 | 43 | 49 |
|  | 45 to 54 | 18 | 44 | 52 | 124 |
|  | 55 to 64 | 31 | 74 | 90 | 214 |
|  | 65 to 74 | 36 | 117 | 105 | 346 |
|  | 75 plus | 49 | 225 | 142 | 645 |
| Race | White | 122 | 39 | 349 | 110 |
|  | Black | 20 | 46 | 63 | 145 |
|  | Other | 11 | 46 | 31 | 127 |
| Ethnicity | Hispanic | 13 | 31 | 44 | 105 |
|  | Not Hispanic | 140 | 41 | 399 | 118 |
| Sex | Female | 74 | 36 | 217 | 105 |
|  | Male | 78 | 44 | 222 | 126 |
| Total |  | 152 | 40 | 439 | 114 |

Source: Healthcare Cost and Utilization Project National Inpatient Sample. Rates for age groups are unadjusted and all other rates are age-adjusted.

Peptic ulcer disease contributed to 7,000 deaths in 2019 (Table 28). Mortality was uncommon among the youngest age groups and then rates (underlying or other cause) increased with age. Age-adjusted mortality rates were higher among men, Whites, and non-Hispanics. Between 2004 and 2019, age-adjusted mortality rates (per 100,000) with peptic ulcer disease as underlying or other cause decreased by 39% from 2.8 to 1.7.(4)

**Table 28:** Peptic Ulcer Disease: Deaths with underlying or underlying/other cause and lifetime years of life lost by age, race, ethnicity, and sex in the United States, 2019.

| Demographic Characteristics |  | Underlying Cause |  |  | Underlying/Other Cause |  |  |
| --- | --- | --- | --- | --- | --- | --- | --- |
|  |  | Number of Deaths | Rate per 100,000 | Years of potential life lost in thousands | Number of Deaths | Rate per 100,000 | Years of potential life lost in thousands |
| Age (years) | 0 to 11 | 1 | 0.0 | 0.1 | 2 | 0.0 | 0.1 |
|  | 12 to 24 | 5 | 0.0 | 0.3 | 12 | 0.0 | 0.7 |
|  | 25 to 44 | 122 | 0.1 | 5.1 | 202 | 0.2 | 8.5 |
|  | 45 to 54 | 237 | 0.6 | 7.2 | 437 | 1.1 | 13.4 |
|  | 55 to 64 | 584 | 1.4 | 13.4 | 1,136 | 2.7 | 26.0 |
|  | 65 to 74 | 758 | 2.4 | 12.0 | 1,607 | 5.1 | 25.3 |
|  | 75 plus | 1,848 | 8.2 | 12.9 | 3,662 | 16.2 | 25.7 |
| Race | White | 3,022 | 0.9 | 42.4 | 6,009 | 1.8 | 83.1 |
|  | Black | 353 | 0.8 | 5.9 | 677 | 1.6 | 11.1 |
|  | Other | 180 | 0.8 | 2.7 | 372 | 1.5 | 5.6 |
| Ethnicity | Hispanic | 227 | 0.6 | 3.8 | 482 | 1.3 | 8.2 |
|  | Not Hispanic | 3,328 | 0.9 | 47.2 | 6,576 | 1.8 | 91.6 |
| Sex | Female | 1,751 | 0.8 | 23.8 | 3,336 | 1.5 | 45.5 |
|  | Male | 1,804 | 1.0 | 27.2 | 3,722 | 2.1 | 54.3 |
| Total |  | 3,555 | 0.9 | 51.0 | 7,058 | 1.7 | 99.8 |

Source: Vital Statistics of the United States.
Rates for age groups are unadjusted and all other rates are age-adjusted.
Years of potential life lost is based on U.S. life table 2006-2018 estimates of life expectancy at age of death.

Among privately insured enrollees, the claims-based prevalence of peptic ulcer disease (based on all-listed diagnoses) was 0.5% (Table 29). Prevalence increased with age and was higher among women. It was highest among Blacks, similar among Whites and Hispanics, and lowest among Asians. It was highest in the Northeast and South, and lowest in the West.

**Table 29:**
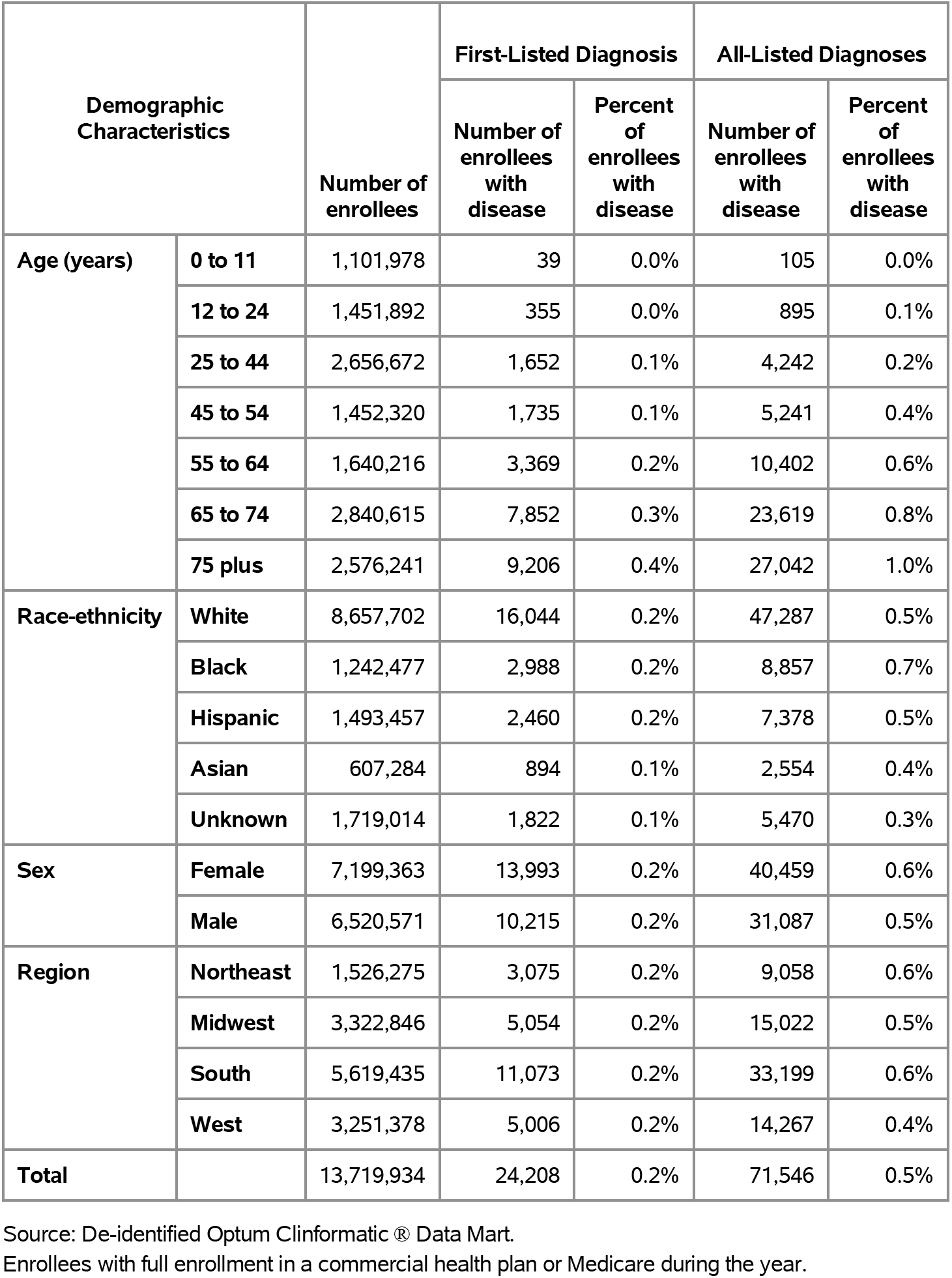
Peptic Ulcer Disease: Claims-based prevalence with first-listed and all-listed diagnoses by age, race-ethnicity, sex and region among privately insured enrollees, 2020.

Among commercial insurance enrollees, ambulatory care visit rates with peptic ulcer disease (all-listed diagnoses) increased with age and were higher among women compared with men (Table 30). Among persons with known race-ethnicity, rates were highest among Blacks, followed by Whites, then Hispanics, and lowest among Asians. Rates were highest in the South, followed by the Northeast, then the West, and lowest in the Midwest.

**Table 30:** Peptic Ulcer Disease: Ambulatory care visits with first-listed and all-listed diagnoses by age, race-ethnicity, sex and region among privately insured enrollees, 2020.

| Demographic Characteristics |  | First-Listed Diagnosis |  | All-Listed Diagnoses |  |
| --- | --- | --- | --- | --- | --- |
|  |  | Number of events | Event rate per 100,000 | Number of events | Event rate per 100,000 |
| Age (years) | 0 to 11 | 46 | 4 | 130 | 12 |
|  | 12 to 24 | 444 | 31 | 1,210 | 83 |
|  | 25 to 44 | 1,962 | 74 | 5,714 | 215 |
|  | 45 to 54 | 2,090 | 144 | 7,129 | 491 |
|  | 55 to 64 | 3,817 | 233 | 13,880 | 846 |
|  | 65 to 74 | 8,754 | 308 | 31,724 | 1,117 |
|  | 75 plus | 9,500 | 369 | 37,610 | 1,460 |
| Race-ethnicity | White | 17,436 | 201 | 64,734 | 748 |
|  | Black | 3,433 | 276 | 11,898 | 958 |
|  | Hispanic | 2,804 | 188 | 10,039 | 672 |
|  | Asian | 1,038 | 171 | 3,648 | 601 |
|  | Unknown | 1,902 | 111 | 7,078 | 412 |
| Sex | Female | 16,007 | 222 | 57,474 | 798 |
|  | Male | 10,606 | 163 | 39,923 | 612 |
| Region | Northeast | 3,337 | 219 | 12,003 | 786 |
|  | Midwest | 5,311 | 160 | 19,397 | 584 |
|  | South | 12,513 | 223 | 45,431 | 808 |
|  | West | 5,452 | 168 | 20,566 | 633 |
| Total |  | 26,613 | 194 | 97,397 | 710 |

Source: De-identified Optum Clinformatics® Data Mart.
Enrollees with full enrollment in a commercial health plan or Medicare during the year.

Among commercial insurance enrollees, emergency department visit rates with peptic ulcer disease (all-listed diagnoses) increased with age and differed little by sex (Table 31). Among persons with known race-ethnicity, rates were highest among Blacks, followed by Whites, then Hispanics, and lowest among Asians. Rates were highest in the Northeast, followed by the South, and lowest in the Midwest and West.

**Table 31:** Peptic Ulcer Disease: Emergency department visits with first-listed and all-listed diagnoses by age, race-ethnicity, sex and region among privately insured enrollees, 2020.

| Demographic Characteristics |  | First-Listed Diagnosis |  | All-Listed Diagnoses |  |
| --- | --- | --- | --- | --- | --- |
|  |  | Number of events | Event rate per 100,000 | Number of events | Event rate per 100,000 |
| Age (years) | 0 to 11 | 1 | 0 | 3 | 0 |
|  | 12 to 24 | 4 | 0 | 22 | 2 |
|  | 25 to 44 | 50 | 2 | 130 | 5 |
|  | 45 to 54 | 51 | 4 | 157 | 11 |
|  | 55 to 64 | 153 | 9 | 366 | 22 |
|  | 65 to 74 | 381 | 13 | 787 | 28 |
|  | 75 plus | 439 | 17 | 939 | 36 |
| Race-ethnicity | White | 768 | 9 | 1,629 | 19 |
|  | Black | 132 | 11 | 314 | 25 |
|  | Hispanic | 79 | 5 | 208 | 14 |
|  | Asian | 21 | 3 | 57 | 9 |
|  | Unknown | 79 | 5 | 196 | 11 |
| Sex | Female | 601 | 8 | 1,326 | 18 |
|  | Male | 478 | 7 | 1,078 | 17 |
| Region | Northeast | 158 | 10 | 334 | 22 |
|  | Midwest | 283 | 9 | 539 | 16 |
|  | South | 433 | 8 | 1,039 | 18 |
|  | West | 205 | 6 | 492 | 15 |
| Total |  | 1,079 | 8 | 2,404 | 18 |

Source: De-identified Optum Clinformatics® Data Mart.
Enrollees with full enrollment in a commercial health plan or Medicare during the year.

Among commercial insurance enrollees, hospital discharge rates with peptic ulcer disease (all-listed diagnoses) increased with age and were higher among men compared with women (Table 32). Among persons with known race-ethnicity, rates were highest among Blacks, followed by Whites, then Hispanics, and lowest among Asians. Rates were higher in the Northeast and South compared with the Midwest and West.

**Table 32:** Peptic Ulcer Disease: Hospital discharges with first-listed and all-listed diagnoses by age, race-ethnicity, sex and region among privately insured enrollees, 2020.

| Demographic Characteristics |  | First-Listed Diagnosis |  | All-Listed Diagnoses |  |
| --- | --- | --- | --- | --- | --- |
|  |  | Number of events | Event rate per 100,000 | Number of events | Event rate per 100,000 |
| Age (years) | 0 to 11 | 3 | 0 | 18 | 2 |
|  | 12 to 24 | 24 | 2 | 100 | 7 |
|  | 25 to 44 | 192 | 7 | 694 | 26 |
|  | 45 to 54 | 337 | 23 | 1,230 | 85 |
|  | 55 to 64 | 919 | 56 | 3,360 | 205 |
|  | 65 to 74 | 2,523 | 89 | 8,812 | 310 |
|  | 75 plus | 4,211 | 163 | 13,342 | 518 |
| Race-ethnicity | White | 5,725 | 66 | 18,415 | 213 |
|  | Black | 941 | 76 | 3,651 | 294 |
|  | Hispanic | 710 | 48 | 2,621 | 175 |
|  | Asian | 240 | 40 | 722 | 119 |
|  | Unknown | 593 | 34 | 2,147 | 125 |
| Sex | Female | 4,213 | 59 | 14,152 | 197 |
|  | Male | 3,996 | 61 | 13,404 | 206 |
| Region | Northeast | 978 | 64 | 3,431 | 225 |
|  | Midwest | 1,866 | 56 | 5,958 | 179 |
|  | South | 3,516 | 63 | 12,581 | 224 |
|  | West | 1,849 | 57 | 5,586 | 172 |
| Total |  | 8,209 | 60 | 27,556 | 201 |

Source: De-identified Optum Clinformatic® Data Mart.
Enrollees with full enrollment in a commercial health plan or Medicare during the year.

Among Medicare beneficiaries, the claims-based prevalence of peptic ulcer disease (based on all-listed diagnoses) was 1.2% (Table 33). Prevalence increased with age until the oldest age group and was higher among women and Blacks. It differed little by region.

**Table 33:** Peptic Ulcer Disease: Claims-based prevalence with first-listed and all-listed diagnoses by age, race, sex and region among fee-for-service, age-eligible Medicare beneficiaries, 2019.

| Demographic Characteristics |  | Number of enrollees | First-Listed Diagnosis |  | All-Listed Diagnoses |  |
| --- | --- | --- | --- | --- | --- | --- |
|  |  |  | Number of enrollees with disease | Percent of enrollees with disease | Number of enrollees with disease | Percent of enrollees with disease |
| Age (years) | 65 to 69 | 8,066,320 | 26,060 | 0.3% | 77,660 | 1.0% |
|  | 70 to 74 | 6,936,940 | 27,640 | 0.4% | 79,920 | 1.2% |
|  | 75 to 79 | 4,894,060 | 23,420 | 0.5% | 68,360 | 1.4% |
|  | 80 to 84 | 3,335,340 | 18,240 | 0.5% | 50,660 | 1.5% |
|  | 85+ | 3,578,120 | 18,260 | 0.5% | 53,480 | 1.5% |
| Race | White | 22,023,800 | 90,720 | 0.4% | 264,140 | 1.2% |
|  | Black | 1,861,660 | 8,340 | 0.4% | 25,460 | 1.4% |
|  | Other | 2,925,320 | 14,560 | 0.5% | 40,480 | 1.4% |
| Sex | Female | 14,975,560 | 65,180 | 0.4% | 188,600 | 1.3% |
|  | Male | 11,835,220 | 48,440 | 0.4% | 141,480 | 1.2% |
| Region | Northeast | 4,748,700 | 19,620 | 0.4% | 56,500 | 1.2% |
|  | Midwest | 6,069,800 | 24,540 | 0.4% | 72,540 | 1.2% |
|  | South | 10,595,900 | 45,840 | 0.4% | 135,980 | 1.3% |
|  | West | 5,396,380 | 23,620 | 0.4% | 65,060 | 1.2% |
| Total |  | 26,810,780 | 113,620 | 0.4% | 330,080 | 1.2% |

Source: Centers for Medicare & Medicaid Services, 5% Denominator and Claims Files. Beneficiaries aged 65+ years with continuous and full Parts A and B enrollment and no Health Maintenance Organization enrollment during the year. Unweighted counts were multiplied by 20 to produce counts for this table.

Among Medicare beneficiaries, ambulatory care visit rates with peptic ulcer disease (all-listed diagnoses) increased with age until 85 years and were higher among women compared with men but differed little by race (Table 34). Rates were highest in the West and South, followed by the Northeast, and lowest in the Midwest.

**Table 34:** Peptic Ulcer Disease: Ambulatory care visits with first-listed and all-listed diagnoses by age, race, sex and region among fee-for-service, age-eligible Medicare beneficiaries, 2019.

| Demographic Characteristics |  | First-Listed Diagnosis |  | All-Listed Diagnoses |  |
| --- | --- | --- | --- | --- | --- |
|  |  | Number of events | Event rate per 100,000 | Number of events | Event rate per 100,000 |
| Age (years) | 65 to 69 | 28,320 | 351 | 108,400 | 1,344 |
|  | 70 to 74 | 30,300 | 437 | 116,640 | 1,681 |
|  | 75 to 79 | 24,660 | 504 | 96,200 | 1,966 |
|  | 80 to 84 | 18,900 | 567 | 75,600 | 2,267 |
|  | 85+ | 17,480 | 489 | 74,560 | 2,084 |
| Race | White | 94,780 | 430 | 374,340 | 1,700 |
|  | Black | 8,280 | 445 | 33,320 | 1,790 |
|  | Other | 16,600 | 567 | 63,740 | 2,179 |
| Sex | Female | 72,060 | 481 | 281,780 | 1,882 |
|  | Male | 47,600 | 402 | 189,620 | 1,602 |
| Region | Northeast | 21,620 | 455 | 81,420 | 1,715 |
|  | Midwest | 24,320 | 401 | 96,520 | 1,590 |
|  | South | 48,400 | 457 | 194,020 | 1,831 |
|  | West | 25,320 | 469 | 99,440 | 1,843 |
| Total |  | 119,660 | 446 | 471,400 | 1,758 |

Source: Centers for Medicare & Medicaid Services, 5% Denominator and Claims Files. Beneficiaries aged 65+ years with continuous and full Parts A and B enrollment and no Health Maintenance Organization enrollment during the year. Unweighted counts were multiplied by 20 to produce counts for this table.

Among Medicare beneficiaries, emergency department visit rates with peptic ulcer disease (all-listed diagnoses) increased with age and were higher among men compared with women and Blacks compared with Whites (Table 35). Rates were higher in the South compared with the Northeast and Midwest and lowest in the West.

**Table 35:**
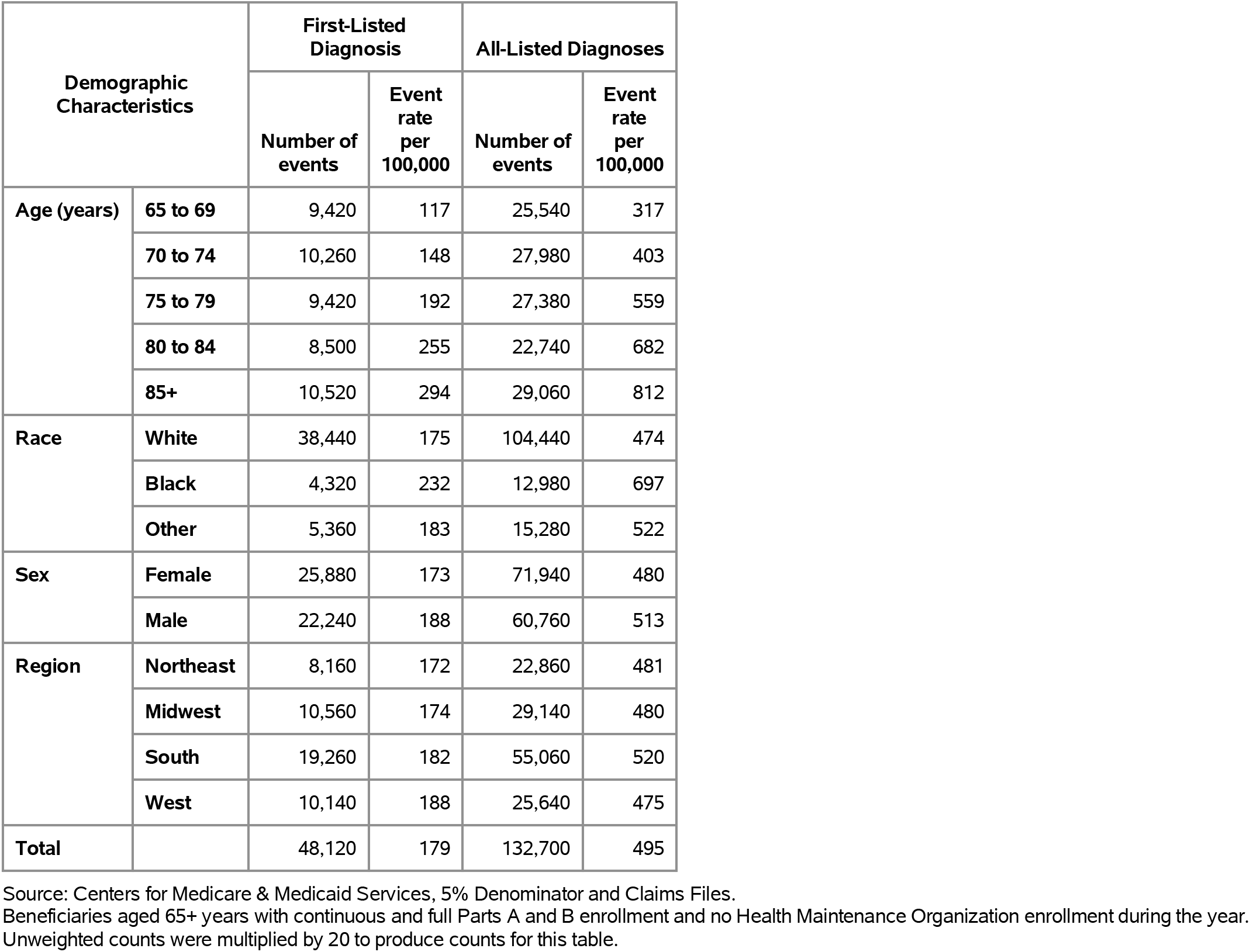
Peptic Ulcer Disease: Emergency department visits with first-listed and all-listed diagnoses by age, race, sex and region among fee-for-service, age-eligible Medicare beneficiaries, 2019.

Among Medicare beneficiaries, hospital discharge rates with peptic ulcer disease (all-listed diagnoses) increased with age and were higher among men compared with women and Blacks compared with Whites (Table 36). Rates were highest in the South, followed by the Midwest, then the West, and lowest in the Northeast.

**Table 36:** Peptic Ulcer Disease: Hospital discharges with first-listed and all-listed diagnoses by age, race, sex and region among fee-for-service, age-eligible Medicare beneficiaries, 2019.

| Demographic Characteristics |  | First-Listed Diagnosis |  | All-Listed Diagnoses |  |
| --- | --- | --- | --- | --- | --- |
|  |  | Number of events | Event rate per 100,000 | Number of events | Event rate per 100,000 |
| Age (years) | 65 to 69 | 9,360 | 116 | 26,920 | 334 |
|  | 70 to 74 | 10,200 | 147 | 29,580 | 426 |
|  | 75 to 79 | 9,440 | 193 | 29,580 | 604 |
|  | 80 to 84 | 8,700 | 261 | 24,140 | 724 |
|  | 85+ | 10,760 | 301 | 30,000 | 838 |
| Race | White | 39,420 | 179 | 111,460 | 506 |
|  | Black | 3,900 | 209 | 12,700 | 682 |
|  | Other | 5,140 | 176 | 16,060 | 549 |
| Sex | Female | 25,600 | 171 | 74,320 | 496 |
|  | Male | 22,860 | 193 | 65,900 | 557 |
| Region | Northeast | 8,020 | 169 | 23,420 | 493 |
|  | Midwest | 10,980 | 181 | 31,920 | 526 |
|  | South | 19,380 | 183 | 57,940 | 547 |
|  | West | 10,080 | 187 | 26,940 | 499 |
| Total |  | 48,460 | 181 | 140,220 | 523 |

Source: Centers for Medicare & Medicaid Services, 5% Denominator and Claims Files. Beneficiaries aged 65+ years with continuous and full Parts A and B enrollment and no Health Maintenance Organization enrollment during the year. Unweighted counts were multiplied by 20 to produce counts for this table.

Functional disorders contributed to 12.2 million ambulatory visits (2015) (Table 37). Ambulatory care visit rates (all-listed diagnoses) were higher among children compared with adolescents and younger adults and then generally increased with age. Age-adjusted rates were higher among women compared with men, Blacks compared with Whites, and Hispanics compared with non-Hispanics.

**Table 37:** Functional Disorders: Ambulatory care visits with first-listed and all-listed diagnoses by age, race, ethnicity, and sex in the United States, 2015.

| Demographic Characteristics |  | First-Listed Diagnosis |  | All-Listed Diagnoses |  |
| --- | --- | --- | --- | --- | --- |
|  |  | Number in thousands | Rate per 100,000 | Number in thousands | Rate per 100,000 |
| Age (years) | 0 to 11 | 871 | 1,790 | 1,815 | 3,732 |
|  | 12 to 24 | 444 | 791 | 991 | 1,765 |
|  | 25 to 44 | 577 | 683 | 2,304 | 2,727 |
|  | 45 to 54 | 444 | 1,030 | 1,921 | 4,459 |
|  | 55 to 64 | 299 | 732 | 1,770 | 4,336 |
|  | 65 to 74 | 612 | 2,224 | 1,571 | 5,707 |
|  | 75 plus | 664 | 3,287 | 1,862 | 9,215 |
| Race | White | 2,837 | 1,128 | 9,633 | 3,626 |
|  | Black | 772 | 1,882 | 1,833 | 4,388 |
|  | Other | 301 | 1,488 | 768 | 3,504 |
| Ethnicity | Hispanic | 874 | 1,923 | 2,434 | 5,097 |
|  | Not Hispanic | 3,036 | 1,125 | 9,800 | 3,498 |
| Sex | Female | 2,298 | 1,407 | 8,394 | 4,921 |
|  | Male | 1,613 | 1,069 | 3,841 | 2,479 |
| Total |  | 3,910 | 1,230 | 12,234 | 3,707 |

Source: National Ambulatory Medical Care Survey (3-year average, 2014-2016). Rates for age groups are unadjusted and all other rates are age-adjusted.

Functional disorders contributed to 4.5 million emergency department visits in 2018 (Table 38). Emergency department visit rates (all-listed diagnoses) were higher among children compared with adolescents and younger adults and then generally increased with age. Age-adjusted rates were higher among women compared with men.

**Table 38:**
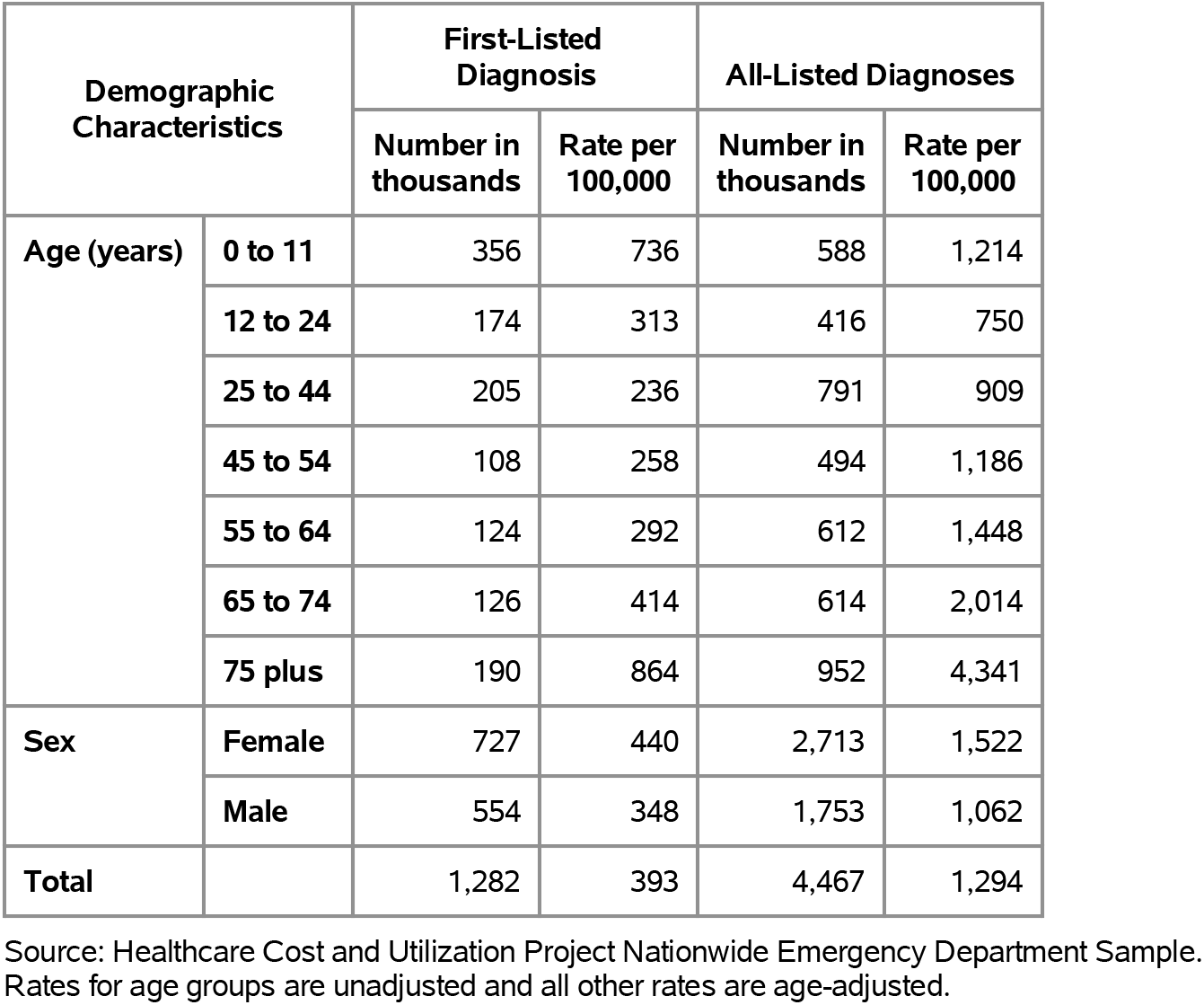
Functional Disorders: Emergency department visits with first-listed and all-listed diagnoses by age and sex in the United States, 2018.

Functional disorders contributed to 2.6 million hospital discharges in 2018 (Table 39). Hospital discharge rates (all-listed diagnoses) increased with age. Age-adjusted rates were higher among women compared with men, Blacks compared with Whites, and non-Hispanics compared with Hispanics. Between 2004 and 2018, age-adjusted hospital discharge rates (per 100,000) with an all-listed diagnosis increased by 64% from 423 to 695.(4)

**Table 39:** Functional Disorders: Hospital discharges with first-listed and all-listed diagnoses by age, race, ethnicity, and sex in the United States, 2018.

| Demographic Characteristics |  | First-Listed Diagnosis |  | All-Listed Diagnoses |  |
| --- | --- | --- | --- | --- | --- |
|  |  | Number in thousands | Rate per 100,000 | Number in thousands | Rate per 100,000 |
| Age (years) | 0 to 11 | 10 | 22 | 82 | 170 |
|  | 12 to 24 | 9 | 16 | 114 | 205 |
|  | 25 to 44 | 21 | 24 | 356 | 409 |
|  | 45 to 54 | 17 | 40 | 298 | 716 |
|  | 55 to 64 | 23 | 54 | 466 | 1,103 |
|  | 65 to 74 | 24 | 77 | 520 | 1,704 |
|  | 75 plus | 27 | 125 | 736 | 3,354 |
| Race | White | 103 | 36 | 2,023 | 668 |
|  | Black | 20 | 46 | 419 | 957 |
|  | Other | 8 | 31 | 150 | 604 |
| Ethnicity | Hispanic | 15 | 31 | 263 | 571 |
|  | Not Hispanic | 116 | 38 | 2,329 | 725 |
| Sex | Female | 76 | 41 | 1,509 | 771 |
|  | Male | 54 | 32 | 1,062 | 614 |
| Total |  | 130 | 37 | 2,571 | 695 |

Source: Healthcare Cost and Utilization Project National Inpatient Sample. Rates for age groups are unadjusted and all other rates are age-adjusted.

Functional disorders contributed to 4,000 deaths in 2019 (Table 40). Mortality rates (underlying or other cause) increased with age. Age-adjusted mortality rates were higher among women, Whites, and non-Hispanics. Between 2004 and 2019, age-adjusted mortality rates (per 100,000) with functional disorders as underlying or other cause increased by 57% from 0.7 to 1.1.(4)

**Table 40:** Functional Disorders: Deaths with underlying or underlying/other cause and lifetime years of life lost by age, race, ethnicity, and sex in the United States, 2019.

| Demographic Characteristics |  | Underlying Cause |  |  | Underlying/Other Cause |  |  |
| --- | --- | --- | --- | --- | --- | --- | --- |
|  |  | Number of Deaths | Rate per 100,000 | Years of potential life lost in thousands | Number of Deaths | Rate per 100,000 | Years of potential life lost in thousands |
| Age (years) | 0 to 11 | 13 | 0.0 | 1.0 | 39 | 0.1 | 2.9 |
|  | 12 to 24 | 11 | 0.0 | 0.6 | 41 | 0.1 | 2.5 |
|  | 25 to 44 | 43 | 0.0 | 1.9 | 179 | 0.2 | 8.0 |
|  | 45 to 54 | 59 | 0.1 | 1.8 | 252 | 0.6 | 7.9 |
|  | 55 to 64 | 127 | 0.3 | 2.9 | 598 | 1.4 | 13.8 |
|  | 65 to 74 | 207 | 0.7 | 3.3 | 924 | 2.9 | 14.6 |
|  | 75 plus | 493 | 2.2 | 3.3 | 2,259 | 10.0 | 15.3 |
| Race | White | 835 | 0.2 | 12.5 | 3,717 | 1.1 | 54.3 |
|  | Black | 99 | 0.2 | 2.1 | 436 | 1.0 | 8.3 |
|  | Other | 19 | 0.1 | 0.3 | 139 | 0.6 | 2.3 |
| Ethnicity | Hispanic | 48 | 0.1 | 1.2 | 254 | 0.6 | 6.0 |
|  | Not Hispanic | 905 | 0.3 | 13.7 | 4,038 | 1.1 | 59.0 |
| Sex | Female | 589 | 0.3 | 8.6 | 2,536 | 1.1 | 36.8 |
|  | Male | 364 | 0.2 | 6.3 | 1,756 | 1.0 | 28.2 |
| Total |  | 953 | 0.2 | 14.9 | 4,292 | 1.1 | 64.9 |

Source: Vital Statistics of the United States.
Rates for age groups are unadjusted and all other rates are age-adjusted.
Years of potential life lost is based on U.S. life table 2006-2018 estimates of life expectancy at age of death.

Among privately insured enrollees, the claims-based prevalence of functional disorders (based on all-listed diagnoses) was 5.1% (Table 41). Prevalence was higher among children compared with adolescents and younger adults and then increased with age. It was higher among women. It was highest among Blacks, followed by Whites and Hispanics, and lowest among Asians. It was higher in the Northeast and South compared with the Midwest and West.

**Table 41:** Functional Disorders: Claims-based prevalence with first-listed and all-listed diagnoses by age, race-ethnicity, sex and region among privately insured enrollees, 2020.

| Demographic Characteristics |  | Number of enrollees | First-Listed Diagnosis |  | All-Listed Diagnoses |  |
| --- | --- | --- | --- | --- | --- | --- |
|  |  |  | Number of enrollees with disease | Percent of enrollees with disease | Number of enrollees with disease | Percent of enrollees with disease |
| Age (years) | 0 to 11 | 1,101,978 | 17,221 | 1.6% | 41,010 | 3.7% |
|  | 12 to 24 | 1,451,892 | 12,495 | 0.9% | 31,145 | 2.1% |
|  | 25 to 44 | 2,656,672 | 24,790 | 0.9% | 71,710 | 2.7% |
|  | 45 to 54 | 1,452,320 | 18,234 | 1.3% | 59,554 | 4.1% |
|  | 55 to 64 | 1,640,216 | 26,001 | 1.6% | 89,930 | 5.5% |
|  | 65 to 74 | 2,840,615 | 53,853 | 1.9% | 179,930 | 6.3% |
|  | 75 plus | 2,576,241 | 65,723 | 2.6% | 227,352 | 8.8% |
| Race-ethnicity | White | 8,657,702 | 135,735 | 1.6% | 444,685 | 5.1% |
|  | Black | 1,242,477 | 25,978 | 2.1% | 84,337 | 6.8% |
|  | Hispanic | 1,493,457 | 23,554 | 1.6% | 74,925 | 5.0% |
|  | Asian | 607,284 | 6,968 | 1.1% | 21,398 | 3.5% |
|  | Unknown | 1,719,014 | 26,082 | 1.5% | 75,286 | 4.4% |
| Sex | Female | 7,199,363 | 142,418 | 2.0% | 458,945 | 6.4% |
|  | Male | 6,520,571 | 75,899 | 1.2% | 241,686 | 3.7% |
| Region | Northeast | 1,526,275 | 30,308 | 2.0% | 90,664 | 5.9% |
|  | Midwest | 3,322,846 | 43,513 | 1.3% | 146,587 | 4.4% |
|  | South | 5,619,435 | 102,125 | 1.8% | 325,473 | 5.8% |
|  | West | 3,251,378 | 42,371 | 1.3% | 137,907 | 4.2% |
| Total |  | 13,719,934 | 218,317 | 1.6% | 700,631 | 5.1% |

Source: De-identified Optum Clinformatic® Data Mart.
Enrollees with full enrollment in a commercial health plan or Medicare during the year.

Among commercial insurance enrollees, ambulatory care visit rates with functional disorders (all-listed diagnoses) were higher among children compared with adolescents and younger adults and then increased with age and were higher among women compared with men (Table 42). Among persons with known race-ethnicity, rates were highest among Blacks, followed by Whites and Hispanics, and lowest among Asians. Rates were highest in the Northeast, followed by the South, and lowest in the West and Midwest.

**Table 42:** Functional Disorders: Ambulatory care visits with first-listed and all-listed diagnoses by age, race-ethnicity, sex and region among privately insured enrollees, 2020.

| Demographic Characteristics |  | First-Listed Diagnosis |  | All-Listed Diagnoses |  |
| --- | --- | --- | --- | --- | --- |
|  |  | Number of events | Event rate per 100,000 | Number of events | Event rate per 100,000 |
| Age (years) | 0 to 11 | 22,308 | 2,024 | 57,585 | 5,226 |
|  | 12 to 24 | 16,865 | 1,162 | 45,951 | 3,165 |
|  | 25 to 44 | 33,288 | 1,253 | 112,940 | 4,251 |
|  | 45 to 54 | 24,548 | 1,690 | 98,347 | 6,772 |
|  | 55 to 64 | 34,877 | 2,126 | 148,806 | 9,072 |
|  | 65 to 74 | 71,228 | 2,507 | 288,154 | 10,144 |
|  | 75 plus | 89,635 | 3,479 | 393,339 | 15,268 |
| Race-ethnicity | White | 182,846 | 2,112 | 726,991 | 8,397 |
|  | Black | 35,038 | 2,820 | 141,648 | 11,400 |
|  | Hispanic | 31,103 | 2,083 | 122,070 | 8,174 |
|  | Asian | 8,907 | 1,467 | 33,771 | 5,561 |
|  | Unknown | 34,855 | 2,028 | 120,642 | 7,018 |
| Sex | Female | 193,110 | 2,682 | 775,604 | 10,773 |
|  | Male | 99,639 | 1,528 | 369,518 | 5,667 |
| Region | Northeast | 42,798 | 2,804 | 155,976 | 10,219 |
|  | Midwest | 56,744 | 1,708 | 225,756 | 6,794 |
|  | South | 136,425 | 2,428 | 537,446 | 9,564 |
|  | West | 56,782 | 1,746 | 225,944 | 6,949 |
| Total |  | 292,749 | 2,134 | 1,145,122 | 8,346 |

Source: De-identified Optum Clinformatics® Data Mart.
Enrollees with full enrollment in a commercial health plan or Medicare during the year.

Among commercial insurance enrollees, emergency department visit rates with functional disorders (all-listed diagnoses) increased with age and were higher among women compared with men (Table 43). Among persons with known race-ethnicity, rates were highest among Blacks, followed by Whites, then Hispanics, and lowest among Asians. Rates were highest in the South, followed by the Northeast, then the West, and lowest in the Midwest.

**Table 43:**
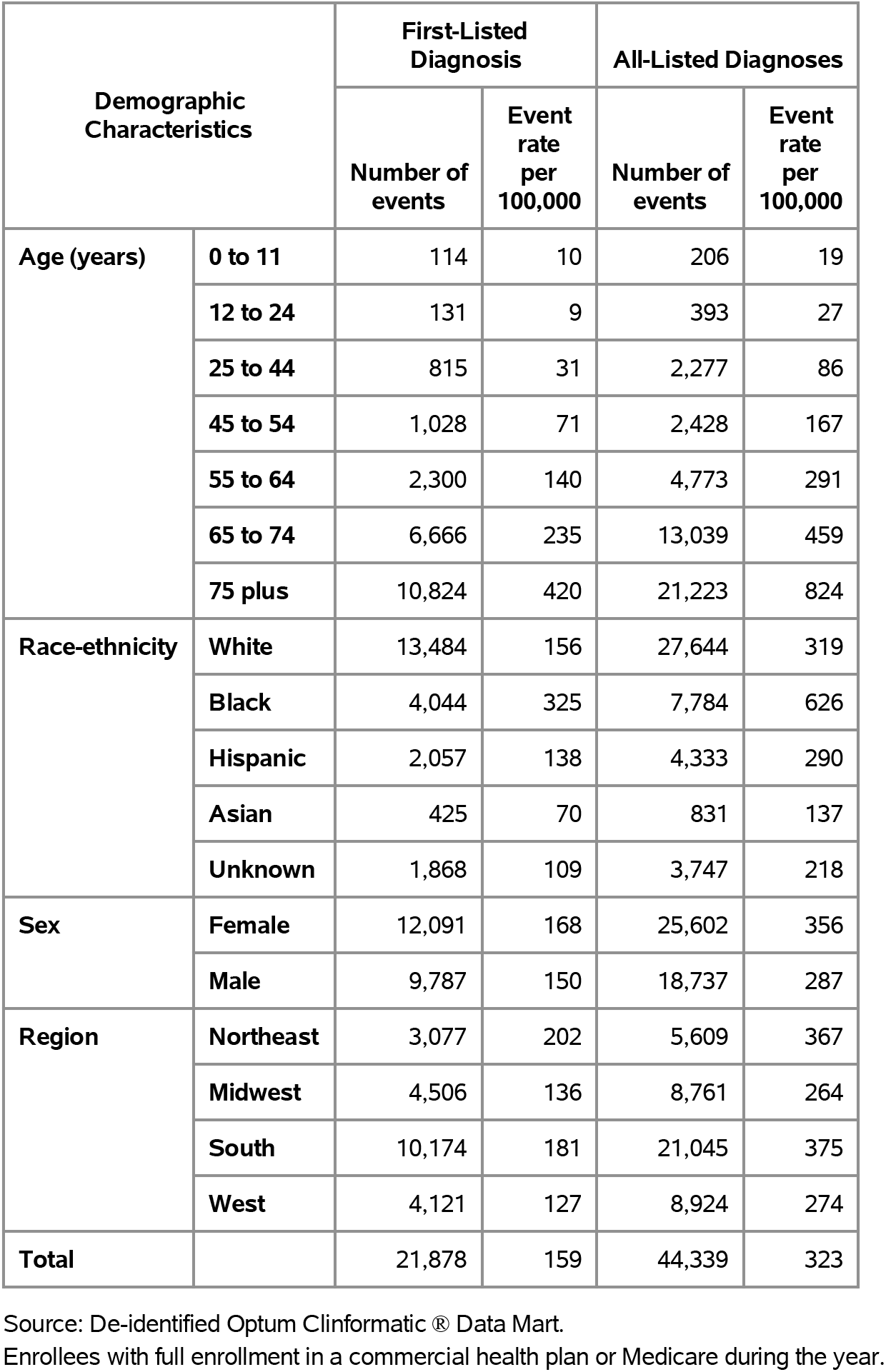
Functional Disorders: Emergency department visits with first-listed and all-listed diagnoses by age, race-ethnicity, sex and region among privately insured enrollees, 2020.

| Demographic Characteristics |  | First-Listed Diagnosis |  | All-Listed Diagnoses |  |
| --- | --- | --- | --- | --- | --- |
|  |  | Number of events | Event rate per 100,000 | Number of events | Event rate per 100,000 |
| Age (years) | 0 to 11 | 114 | 10 | 206 | 19 |
|  | 12 to 24 | 131 | 9 | 393 | 27 |
|  | 25 to 44 | 815 | 31 | 2,277 | 86 |
|  | 45 to 54 | 1,028 | 71 | 2,428 | 167 |
|  | 55 to 64 | 2,300 | 140 | 4,773 | 291 |
|  | 65 to 74 | 6,666 | 235 | 13,039 | 459 |
|  | 75 plus | 10,824 | 420 | 21,223 | 824 |
| Race-ethnicity | White | 13,484 | 156 | 27,644 | 319 |
|  | Black | 4,044 | 325 | 7,784 | 626 |
|  | Hispanic | 2,057 | 138 | 4,333 | 290 |
|  | Asian | 425 | 70 | 831 | 137 |
|  | Unknown | 1,868 | 109 | 3,747 | 218 |
| Sex | Female | 12,091 | 168 | 25,602 | 356 |
|  | Male | 9,787 | 150 | 18,737 | 287 |
| Region | Northeast | 3,077 | 202 | 5,609 | 367 |
|  | Midwest | 4,506 | 136 | 8,761 | 264 |
|  | South | 10,174 | 181 | 21,045 | 375 |
|  | West | 4,121 | 127 | 8,924 | 274 |
| Total |  | 21,878 | 159 | 44,339 | 323 |

Source: De-identified Optum Clinformatics® Data Mart.
Enrollees with full enrollment in a commercial health plan or Medicare during the year.

Among commercial insurance enrollees, hospital discharge rates with functional disorders (all-listed diagnoses) increased with age and were higher among women compared with men (Table 44). Among persons with known race-ethnicity, rates were highest among Blacks, followed by Whites, then Hispanics, and lowest among Asians. Rates were highest in the Northeast, followed by the South, then the Midwest, and lowest in the West.

**Table 44:** Functional Disorders: Hospital discharges with first-listed and all-listed diagnoses by age, race-ethnicity, sex and region among privately insured enrollees, 2020.

| Demographic Characteristics |  | First-Listed Diagnosis |  | All-Listed Diagnoses |  |
| --- | --- | --- | --- | --- | --- |
|  |  | Number of events | Event rate per 100,000 | Number of events | Event rate per 100,000 |
| Age (years) | 0 to 11 | 81 | 7 | 1,222 | 111 |
|  | 12 to 24 | 120 | 8 | 2,601 | 179 |
|  | 25 to 44 | 318 | 12 | 6,990 | 263 |
|  | 45 to 54 | 387 | 27 | 8,714 | 600 |
|  | 55 to 64 | 684 | 42 | 19,259 | 1,174 |
|  | 65 to 74 | 1,219 | 43 | 40,825 | 1,437 |
|  | 75 plus | 1,517 | 59 | 60,455 | 2,347 |
| Race-ethnicity | White | 2,894 | 33 | 92,269 | 1,066 |
|  | Black | 553 | 45 | 20,808 | 1,675 |
|  | Hispanic | 405 | 27 | 12,692 | 850 |
|  | Asian | 93 | 15 | 2,846 | 469 |
|  | Unknown | 381 | 22 | 11,451 | 666 |
| Sex | Female | 2,521 | 35 | 81,522 | 1,132 |
|  | Male | 1,805 | 28 | 58,544 | 898 |
| Region | Northeast | 611 | 40 | 18,619 | 1,220 |
|  | Midwest | 919 | 28 | 30,462 | 917 |
|  | South | 1,985 | 35 | 66,446 | 1,182 |
|  | West | 811 | 25 | 24,539 | 755 |
| Total |  | 4,326 | 32 | 140,066 | 1,021 |

Source: De-identified Optum Clinformatic® Data Mart.
Enrollees with full enrollment in a commercial health plan or Medicare during the year.

Among Medicare beneficiaries, the claims-based prevalence of functional disorders (based on all-listed diagnoses) was 11.3% (Table 45). Prevalence increased with age and was higher among women and Blacks. It was highest in the South, followed by the Northeast, then the Midwest, and lowest in the West.

**Table 45:** Functional Disorders: Claims-based prevalence with first-listed and all-listed diagnoses by age, race, sex and region among fee-for-service, age-eligible Medicare beneficiaries, 2019.

| Demographic Characteristics |  | Number of enrollees | First-Listed Diagnosis |  | All-Listed Diagnoses |  |
| --- | --- | --- | --- | --- | --- | --- |
|  |  |  | Number of enrollees with disease | Percent of enrollees with disease | Number of enrollees with disease | Percent of enrollees with disease |
| Age (years) | 65 to 69 | 8,066,320 | 200,600 | 2.5% | 651,060 | 8.1% |
|  | 70 to 74 | 6,936,940 | 211,120 | 3.0% | 672,680 | 9.7% |
|  | 75 to 79 | 4,894,060 | 185,700 | 3.8% | 584,700 | 11.9% |
|  | 80 to 84 | 3,335,340 | 149,220 | 4.5% | 487,200 | 14.6% |
|  | 85+ | 3,578,120 | 192,180 | 5.4% | 644,840 | 18.0% |
| Race | White | 22,023,800 | 760,060 | 3.5% | 2,477,760 | 11.3% |
|  | Black | 1,861,660 | 78,480 | 4.2% | 242,680 | 13.0% |
|  | Other | 2,925,320 | 100,280 | 3.4% | 320,040 | 10.9% |
| Sex | Female | 14,975,560 | 609,300 | 4.1% | 1,981,680 | 13.2% |
|  | Male | 11,835,220 | 329,520 | 2.8% | 1,058,800 | 8.9% |
| Region | Northeast | 4,748,700 | 183,080 | 3.9% | 565,260 | 11.9% |
|  | Midwest | 6,069,800 | 185,320 | 3.1% | 648,220 | 10.7% |
|  | South | 10,595,900 | 399,780 | 3.8% | 1,286,480 | 12.1% |
|  | West | 5,396,380 | 170,640 | 3.2% | 540,520 | 10.0% |
| Total |  | 26,810,780 | 938,820 | 3.5% | 3,040,480 | 11.3% |

Source: Centers for Medicare & Medicaid Services, 5% Denominator and Claims Files. Beneficiaries aged 65+ years with continuous and full Parts A and B enrollment and no Health Maintenance Organization enrollment during the year. Unweighted counts were multiplied by 20 to produce counts for this table.

Among Medicare beneficiaries, ambulatory care visit rates with functional disorders (all-listed diagnoses) increased with age and were higher among women compared with men and Blacks compared with Whites (Table 46). Rates were highest in the Northeast, followed by the South, then the Midwest, and lowest in the West.

**Table 46:** Functional Disorders: Ambulatory care visits with first-listed and all-listed diagnoses by age, race, sex and region among fee-for-service, age-eligible Medicare beneficiaries, 2019.

| Demographic Characteristics |  | First-Listed Diagnosis |  | All-Listed Diagnoses |  |
| --- | --- | --- | --- | --- | --- |
|  |  | Number of events | Event rate per 100,000 | Number of events | Event rate per 100,000 |
| Age (years) | 65 to 69 | 246,240 | 3,053 | 1,048,200 | 12,995 |
|  | 70 to 74 | 263,460 | 3,798 | 1,102,520 | 15,893 |
|  | 75 to 79 | 233,340 | 4,768 | 979,480 | 20,014 |
|  | 80 to 84 | 190,220 | 5,703 | 834,440 | 25,018 |
|  | 85+ | 250,060 | 6,989 | 1,238,160 | 34,604 |
| Race | White | 962,360 | 4,370 | 4,196,640 | 19,055 |
|  | Black | 94,800 | 5,092 | 431,000 | 23,151 |
|  | Other | 126,160 | 4,313 | 575,160 | 19,661 |
| Sex | Female | 801,320 | 5,351 | 3,560,620 | 23,776 |
|  | Male | 382,000 | 3,228 | 1,642,180 | 13,875 |
| Region | Northeast | 245,840 | 5,177 | 1,012,540 | 21,322 |
|  | Midwest | 221,640 | 3,652 | 1,048,280 | 17,270 |
|  | South | 506,440 | 4,780 | 2,235,620 | 21,099 |
|  | West | 209,400 | 3,880 | 906,360 | 16,796 |
| Total |  | 1,183,320 | 4,414 | 5,202,800 | 19,406 |

Source: Centers for Medicare & Medicaid Services, 5% Denominator and Claims Files. Beneficiaries aged 65+ years with continuous and full Parts A and B enrollment and no Health Maintenance Organization enrollment during the year. Unweighted counts were multiplied by 20 to produce counts for this table.

Among Medicare beneficiaries, emergency department visit rates with functional disorders (all-listed diagnoses) increased with age and were higher among women compared with men and Blacks compared with Whites (Table 47). Rates were highest in the Northeast and South, followed by the Midwest, and lowest in the West.

**Table 47:** Functional Disorders: Emergency department visits with first-listed and all-listed diagnoses by age, race, sex and region among fee-for-service, age-eligible Medicare beneficiaries, 2019.

| Demographic Characteristics |  | First-Listed Diagnosis |  | All-Listed Diagnoses |  |
| --- | --- | --- | --- | --- | --- |
|  |  | Number of events | Event rate per 100,000 | Number of events | Event rate per 100,000 |
| Age (years) | 65 to 69 | 43,220 | 536 | 181,220 | 2,247 |
|  | 70 to 74 | 44,820 | 646 | 191,960 | 2,767 |
|  | 75 to 79 | 44,900 | 917 | 184,600 | 3,772 |
|  | 80 to 84 | 38,840 | 1,164 | 169,820 | 5,092 |
|  | 85+ | 54,680 | 1,528 | 261,520 | 7,309 |
| Race | White | 173,280 | 787 | 789,220 | 3,583 |
|  | Black | 28,860 | 1,550 | 106,400 | 5,715 |
|  | Other | 24,320 | 831 | 93,500 | 3,196 |
| Sex | Female | 123,240 | 823 | 599,640 | 4,004 |
|  | Male | 103,220 | 872 | 389,480 | 3,291 |
| Region | Northeast | 38,840 | 818 | 185,980 | 3,916 |
|  | Midwest | 49,440 | 815 | 219,240 | 3,612 |
|  | South | 94,520 | 892 | 414,480 | 3,912 |
|  | West | 43,660 | 809 | 169,420 | 3,140 |
| Total |  | 226,460 | 845 | 989,120 | 3,689 |

Source: Centers for Medicare & Medicaid Services, 5% Denominator and Claims Files. Beneficiaries aged 65+ years with continuous and full Parts A and B enrollment and no Health Maintenance Organization enrollment during the year. Unweighted counts were multiplied by 20 to produce counts for this table.

Among Medicare beneficiaries, hospital discharge rates with functional disorders (all-listed diagnoses) increased with age and were higher among women compared with men and Blacks compared with Whites (Table 48). Rates were lower in the West compared with the South, Northeast, and Midwest.

**Table 48:** Functional Disorders: Hospital discharges with first-listed and all-listed diagnoses by age, race, sex and region among fee-for-service, age-eligible Medicare beneficiaries, 2019.

| Demographic Characteristics |  | First-Listed Diagnosis |  | All-Listed Diagnoses |  |
| --- | --- | --- | --- | --- | --- |
|  |  | Number of events | Event rate per 100,000 | Number of events | Event rate per 100,000 |
| Age (years) | 65 to 69 | 7,560 | 94 | 159,320 | 1,975 |
|  | 70 to 74 | 6,840 | 99 | 168,820 | 2,434 |
|  | 75 to 79 | 6,080 | 124 | 153,340 | 3,133 |
|  | 80 to 84 | 4,220 | 127 | 137,960 | 4,136 |
|  | 85+ | 5,900 | 165 | 201,980 | 5,645 |
| Race | White | 23,540 | 107 | 669,340 | 3,039 |
|  | Black | 3,660 | 197 | 77,220 | 4,148 |
|  | Other | 3,400 | 116 | 74,860 | 2,559 |
| Sex | Female | 16,700 | 112 | 489,400 | 3,268 |
|  | Male | 13,900 | 117 | 332,020 | 2,805 |
| Region | Northeast | 5,960 | 126 | 154,940 | 3,263 |
|  | Midwest | 6,540 | 108 | 187,240 | 3,085 |
|  | South | 12,400 | 117 | 348,160 | 3,286 |
|  | West | 5,700 | 106 | 131,080 | 2,429 |
| Total |  | 30,600 | 114 | 821,420 | 3,064 |

Source: Centers for Medicare & Medicaid Services, 5% Denominator and Claims Files. Beneficiaries aged 65+ years with continuous and full Parts A and B enrollment and no Health Maintenance Organization enrollment during the year. Unweighted counts were multiplied by 20 to produce counts for this table.

Appendicitis contributed to 739,000 ambulatory visits (2015) (Table 49). Ambulatory care visit rates (all-listed diagnoses) were highest among persons 55-64 years and lowest among those 75 years and over. Age-adjusted ambulatory care visit rates were higher among men compared with women, Whites compared with Blacks, and Hispanics compared with non-Hispanics.

**Table 49:** Appendicitis: Ambulatory care visits with first-listed and all-listed diagnoses by age, race, ethnicity, and sex in the United States, 2015.

| Demographic Characteristics |  | First-Listed Diagnosis |  | All-Listed Diagnoses |  |
| --- | --- | --- | --- | --- | --- |
|  |  | Number in thousands | Rate per 100,000 | Number in thousands | Rate per 100,000 |
| Age (years) | 0 to 11 | 35 | 72 | 40 | 82 |
|  | 12 to 24 | 135 | 240 | 155 | 277 |
|  | 25 to 44 | 125 | 148 | 166 | 197 |
|  | 45 to 54 | 65 | 151 | 74 | 172 |
|  | 55 to 64 | 204 | 501 | 223 | 546 |
|  | 65 to 74 | 75 | 274 | 77 | 281 |
|  | 75 plus | 3 | 17 | 3 | 17 |
| Race | White | 539 | 194 | 601 | 220 |
|  | Black | 39 | 83 | 70 | 152 |
|  | Other | 65 | 258 | 67 | 269 |
| Ethnicity | Hispanic | 230 | 464 | 245 | 490 |
|  | Not Hispanic | 412 | 151 | 494 | 181 |
| Sex | Female | 296 | 170 | 325 | 189 |
|  | Male | 346 | 195 | 414 | 236 |
| Total |  | 643 | 182 | 739 | 212 |

Source: National Ambulatory Medical Care Survey (3-year average, 2014-2016). Rates for age groups are unadjusted and all other rates are age-adjusted.

Appendicitis contributed to 421,000 emergency department visits in 2018 (Table 50). Emergency department visit rates (all-listed diagnoses) peaked among persons 12-24 years and then decreased with age. Age-adjusted rates were higher among men compared with women.

**Table 50:** Appendicitis: Emergency department visits with first-listed and all-listed diagnoses by age and sex in the United States, 2018.

| Demographic Characteristics |  | First-Listed Diagnosis |  | All-Listed Diagnoses |  |
| --- | --- | --- | --- | --- | --- |
|  |  | Number in thousands | Rate per 100,000 | Number in thousands | Rate per 100,000 |
| Age (years) | 0 to 11 | 52 | 108 | 55 | 113 |
|  | 12 to 24 | 108 | 196 | 115 | 208 |
|  | 25 to 44 | 112 | 129 | 123 | 141 |
|  | 45 to 54 | 43 | 104 | 48 | 115 |
|  | 55 to 64 | 35 | 83 | 41 | 96 |
|  | 65 to 74 | 21 | 68 | 25 | 83 |
|  | 75 plus | 10 | 47 | 14 | 66 |
| Sex | Female | 177 | 111 | 198 | 123 |
|  | Male | 205 | 131 | 223 | 142 |
| Total |  | 382 | 121 | 421 | 133 |

Source: Healthcare Cost and Utilization Project Nationwide Emergency Department Sample. Rates for age groups are unadjusted and all other rates are age-adjusted.

Appendicitis contributed to 193,000 hospital discharges in 2018 (Table 51). Hospital discharge rates (all-listed diagnoses) peaked at 12-24 years and were similar for other age groups. Age-adjusted rates were higher among men compared with women, Whites compared with Blacks, and Hispanics compared with non-Hispanics. Between 2004 and 2018, age-adjusted hospital discharge rates (per 100,000) with an all-listed diagnosis decreased by 47% from 111 to 59. (4,6)

**Table 51:** Appendicitis: Hospital discharges with first-listed and all-listed diagnoses by age, race, ethnicity, and sex in the United States, 2018.

| Demographic Characteristics |  | First-Listed Diagnosis |  | All-Listed Diagnoses |  |
| --- | --- | --- | --- | --- | --- |
|  |  | Number in thousands | Rate per 100,000 | Number in thousands | Rate per 100,000 |
| Age (years) | 0 to 11 | 22 | 46 | 24 | 50 |
|  | 12 to 24 | 36 | 64 | 40 | 72 |
|  | 25 to 44 | 41 | 47 | 49 | 56 |
|  | 45 to 54 | 20 | 48 | 25 | 60 |
|  | 55 to 64 | 19 | 45 | 25 | 58 |
|  | 65 to 74 | 14 | 45 | 18 | 60 |
|  | 75 plus | 8 | 38 | 12 | 56 |
| Race | White | 131 | 53 | 159 | 63 |
|  | Black | 12 | 27 | 16 | 36 |
|  | Other | 16 | 63 | 19 | 74 |
| Ethnicity | Hispanic | 41 | 66 | 48 | 79 |
|  | Not Hispanic | 119 | 45 | 147 | 54 |
| Sex | Female | 73 | 44 | 91 | 54 |
|  | Male | 86 | 54 | 103 | 64 |
| Total |  | 159 | 49 | 193 | 59 |

Source: Healthcare Cost and Utilization Project National Inpatient Sample. Rates for age groups are unadjusted and all other rates are age-adjusted.

Appendicitis contributed to 1,000 deaths in 2019 (Table 52). Mortality was uncommon through young adulthood after which rates (underlying or other cause) increased with age. Age-adjusted mortality rates did not differ by sex, race, or ethnicity. Between 2004 and 2019, age-adjusted mortality rates (per 100,000) with appendicitis as underlying or other cause decreased by a third from 0.3 to 0.2.(4)

**Table 52:** Appendicitis: Deaths with underlying or underlying/other cause and lifetime years of life lost by age, race, ethnicity, and sex in the United States, 2019.

| Demographic Characteristics |  | Underlying Cause |  |  | Underlying/Other Cause |  |  |
| --- | --- | --- | --- | --- | --- | --- | --- |
|  |  | Number of Deaths | Rate per 100,000 | Years of potential life lost in thousands | Number of Deaths | Rate per 100,000 | Years of potential life lost in thousands |
| Age (years) | 0 to 11 | 6 | 0.0 | 0.4 | 10 | 0.0 | 0.7 |
|  | 12 to 24 | 7 | 0.0 | 0.4 | 14 | 0.0 | 0.9 |
|  | 25 to 44 | 24 | 0.0 | 1.0 | 38 | 0.0 | 1.6 |
|  | 45 to 54 | 22 | 0.1 | 0.7 | 54 | 0.1 | 1.7 |
|  | 55 to 64 | 54 | 0.1 | 1.3 | 126 | 0.3 | 2.9 |
|  | 65 to 74 | 77 | 0.2 | 1.2 | 153 | 0.5 | 2.4 |
|  | 75 plus | 201 | 0.9 | 1.4 | 336 | 1.5 | 2.4 |
| Race | White | 328 | 0.1 | 5.1 | 619 | 0.2 | 10.2 |
|  | Black | 42 | 0.1 | 1.0 | 73 | 0.2 | 1.7 |
|  | Other | 21 | 0.1 | 0.3 | 39 | 0.2 | 0.7 |
| Ethnicity | Hispanic | 33 | 0.1 | 0.9 | 70 | 0.2 | 1.9 |
|  | Not Hispanic | 358 | 0.1 | 5.6 | 661 | 0.2 | 10.7 |
| Sex | Female | 177 | 0.1 | 2.7 | 336 | 0.2 | 5.8 |
|  | Male | 214 | 0.1 | 3.7 | 395 | 0.2 | 6.8 |
| Total |  | 391 | 0.1 | 6.4 | 731 | 0.2 | 12.6 |

Source: Vital Statistics of the United States.
Rates for age groups are unadjusted and all other rates are age-adjusted.
Years of potential life lost is based on U.S. life table 2006-2018 estimates of life expectancy at age of death.

Among privately insured enrollees, the claims-based prevalence of appendicitis (based on all-listed diagnoses) was 0.1% (Table 53). Prevalence was highest among adolescents and younger adults and did not differ by sex. It was highest among Hispanics and similar among Whites, Blacks, and Asians. It did not differ by region.

**Table 53:**
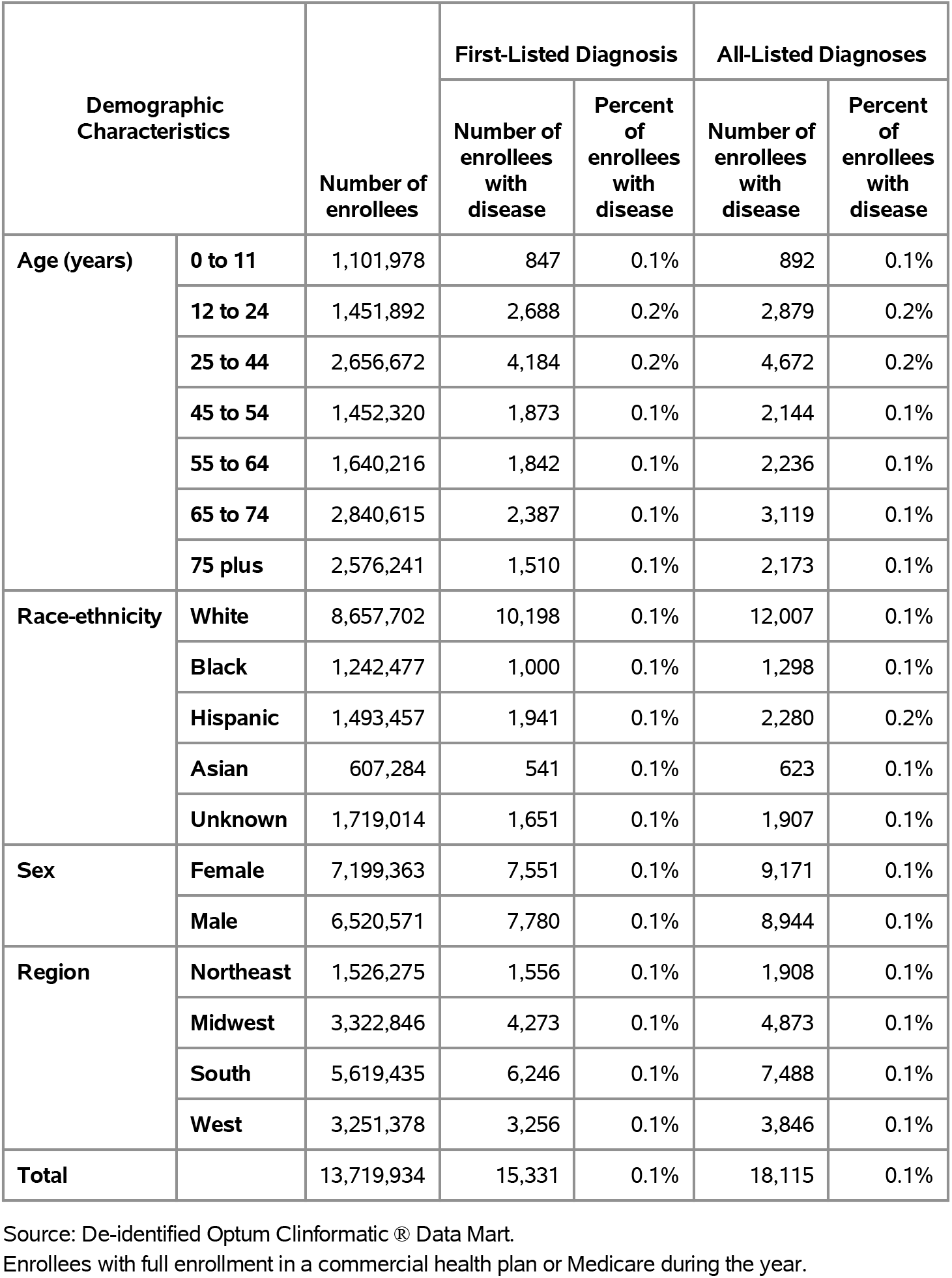
Appendicitis: Claims-based prevalence with first-listed and all-listed diagnoses by age, race-ethnicity, sex and region among privately insured enrollees, 2020.

Among commercial insurance enrollees, ambulatory care visit rates with appendicitis (all-listed diagnoses) peaked among adolescents and the youngest adults and then decreased with age and were higher among men compared with women (Table 54). Among persons with known race-ethnicity, rates were highest among Hispanics, followed by Whites, then Asians, and lowest among Blacks. Rates were highest in the Midwest, followed by the South and Northeast, and lowest in the West.

**Table 54:** Appendicitis: Ambulatory care visits with first-listed and all-listed diagnoses by age, race-ethnicity, sex and region among privately insured enrollees, 2020.

| Demographic Characteristics |  | First-Listed Diagnosis |  | All-Listed Diagnoses |  |
| --- | --- | --- | --- | --- | --- |
|  |  | Number of events | Event rate per 100,000 | Number of events | Event rate per 100,000 |
| Age (years) | 0 to 11 | 1,105 | 100 | 1,191 | 108 |
|  | 12 to 24 | 3,575 | 246 | 3,879 | 267 |
|  | 25 to 44 | 5,492 | 207 | 6,210 | 234 |
|  | 45 to 54 | 2,503 | 172 | 2,938 | 202 |
|  | 55 to 64 | 2,313 | 141 | 2,821 | 172 |
|  | 65 to 74 | 2,725 | 96 | 3,445 | 121 |
|  | 75 plus | 1,750 | 68 | 2,517 | 98 |
| Race-ethnicity | White | 12,963 | 150 | 15,250 | 176 |
|  | Black | 1,274 | 103 | 1,576 | 127 |
|  | Hispanic | 2,447 | 164 | 2,924 | 196 |
|  | Asian | 689 | 113 | 809 | 133 |
|  | Unknown | 2,090 | 122 | 2,442 | 142 |
| Sex | Female | 9,717 | 135 | 11,752 | 163 |
|  | Male | 9,746 | 149 | 11,249 | 173 |
| Region | Northeast | 2,113 | 138 | 2,566 | 168 |
|  | Midwest | 5,385 | 162 | 6,135 | 185 |
|  | South | 7,960 | 142 | 9,527 | 170 |
|  | West | 4,005 | 123 | 4,773 | 147 |
| Total |  | 19,463 | 142 | 23,001 | 168 |

Source: De-identified Optum Clinformatic® Data Mart.
Enrollees with full enrollment in a commercial health plan or Medicare during the year.

Among commercial insurance enrollees, emergency department visit rates with appendicitis (all-listed diagnoses) increased with age until 75 years and were higher among women compared with men (Table 55). Among persons with known race-ethnicity, rates were highest among Whites and Hispanics, followed by Blacks, and lowest among Asians. Rates were higher in the Midwest and West compared with the Northeast and South.

**Table 55:**
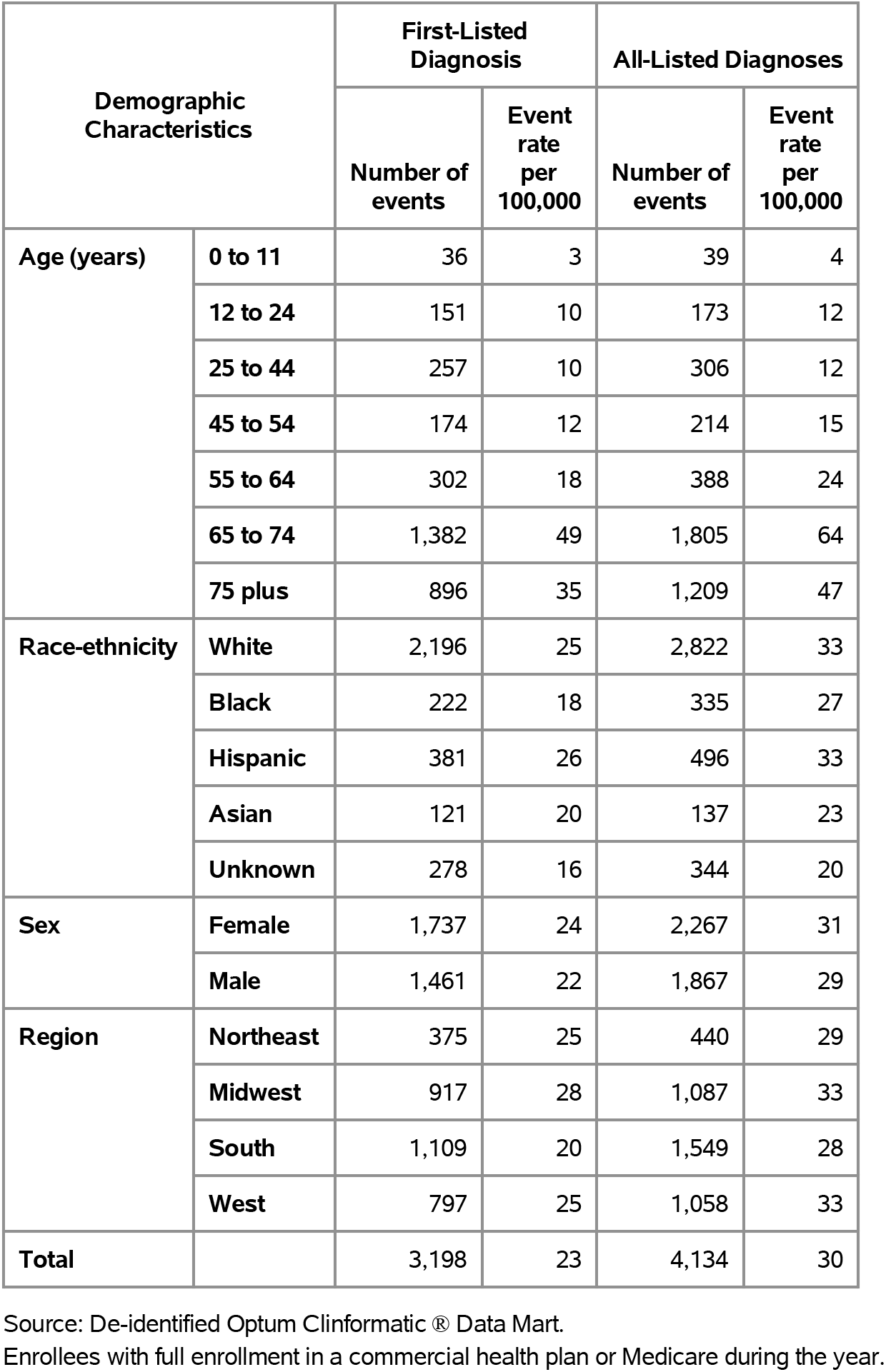
Appendicitis: Emergency department visits with first-listed and all-listed diagnoses by age, race-ethnicity, sex and region among privately insured enrollees, 2020.

Among commercial insurance enrollees, hospital discharge rates with appendicitis (all-listed diagnoses) generally increased with age and were higher among men compared with women (Table 56). Among persons with known race-ethnicity, rates were highest among Hispanics, followed by Whites and Blacks, and lowest among Asians. Rates were highest in the Northeast, followed by the South and Midwest, and lowest in the West.

**Table 56:**
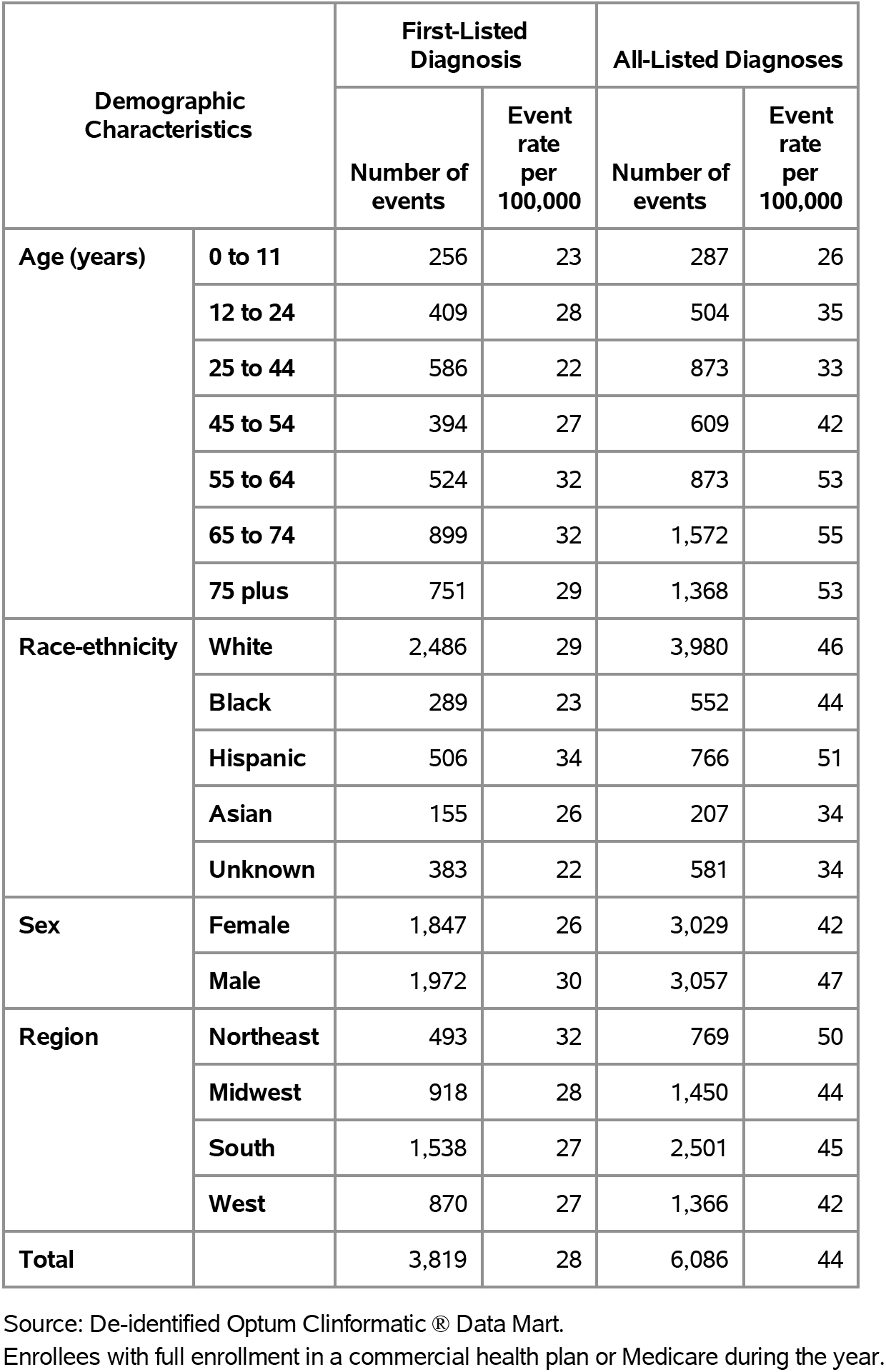
Appendicitis: Hospital discharges with first-listed and all-listed diagnoses by age, race-ethnicity, sex and region among privately insured enrollees, 2020.

| Demographic Characteristics |  | First-Listed Diagnosis |  | All-Listed Diagnoses |  |
| --- | --- | --- | --- | --- | --- |
|  |  | Number of events | Event rate per 100,000 | Number of events | Event rate per 100,000 |
| Age (years) | 0 to 11 | 256 | 23 | 287 | 26 |
|  | 12 to 24 | 409 | 28 | 504 | 35 |
|  | 25 to 44 | 586 | 22 | 873 | 33 |
|  | 45 to 54 | 394 | 27 | 609 | 42 |
|  | 55 to 64 | 524 | 32 | 873 | 53 |
|  | 65 to 74 | 899 | 32 | 1,572 | 55 |
|  | 75 plus | 751 | 29 | 1,368 | 53 |
| Race-ethnicity | White | 2,486 | 29 | 3,980 | 46 |
|  | Black | 289 | 23 | 552 | 44 |
|  | Hispanic | 506 | 34 | 766 | 51 |
|  | Asian | 155 | 26 | 207 | 34 |
|  | Unknown | 383 | 22 | 581 | 34 |
| Sex | Female | 1,847 | 26 | 3,029 | 42 |
|  | Male | 1,972 | 30 | 3,057 | 47 |
| Region | Northeast | 493 | 32 | 769 | 50 |
|  | Midwest | 918 | 28 | 1,450 | 44 |
|  | South | 1,538 | 27 | 2,501 | 45 |
|  | West | 870 | 27 | 1,366 | 42 |
| Total |  | 3,819 | 28 | 6,086 | 44 |

Source: De-identified Optum Clinformatics® Data Mart.
Enrollees with full enrollment in a commercial health plan or Medicare during the year.

Among Medicare beneficiaries, the claims-based prevalence of appendicitis (based on all-listed diagnoses) was 0.1% (Table 57). Prevalence did not differ by age, sex, or race. It did not differ by region.

**Table 57:** Appendicitis: Claims-based prevalence with first-listed and all-listed diagnoses by age, race, sex and region among fee-for-service, age-eligible Medicare beneficiaries, 2019.

| Demographic Characteristics |  | Number of enrollees | First-Listed Diagnosis |  | All-Listed Diagnoses |  |
| --- | --- | --- | --- | --- | --- | --- |
|  |  |  | Number of enrollees with disease | Percent of enrollees with disease | Number of enrollees with disease | Percent of enrollees with disease |
| Age (years) | 65 to 69 | 8,066,320 | 8,880 | 0.1% | 10,440 | 0.1% |
|  | 70 to 74 | 6,936,940 | 6,960 | 0.1% | 8,720 | 0.1% |
|  | 75 to 79 | 4,894,060 | 4,160 | 0.1% | 5,560 | 0.1% |
|  | 80 to 84 | 3,335,340 | 2,220 | 0.1% | 2,900 | 0.1% |
|  | 85+ | 3,578,120 | 2,100 | 0.1% | 2,940 | 0.1% |
| Race | White | 22,023,800 | 20,360 | 0.1% | 25,260 | 0.1% |
|  | Black | 1,861,660 | 1,060 | 0.1% | 1,500 | 0.1% |
|  | Other | 2,925,320 | 2,900 | 0.1% | 3,800 | 0.1% |
| Sex | Female | 14,975,560 | 13,420 | 0.1% | 16,720 | 0.1% |
|  | Male | 11,835,220 | 10,900 | 0.1% | 13,840 | 0.1% |
| Region | Northeast | 4,748,700 | 4,060 | 0.1% | 5,040 | 0.1% |
|  | Midwest | 6,069,800 | 5,280 | 0.1% | 6,560 | 0.1% |
|  | South | 10,595,900 | 9,680 | 0.1% | 12,300 | 0.1% |
|  | West | 5,396,380 | 5,300 | 0.1% | 6,660 | 0.1% |
| Total |  | 26,810,780 | 24,320 | 0.1% | 30,560 | 0.1% |

Source: Centers for Medicare & Medicaid Services, 5% Denominator and Claims Files. Beneficiaries aged 65+ years with continuous and full Parts A and B enrollment and no Health Maintenance Organization enrollment during the year. Unweighted counts were multiplied by 20 to produce counts for this table.

Among Medicare beneficiaries, ambulatory care visit rates with appendicitis (all-listed diagnoses) decreased with age, differed little by sex, and were higher among Whites compared with Blacks (Table 58). Rates were highest in the West, followed by the South, then the Midwest, and lowest in the Northeast.

**Table 58:**
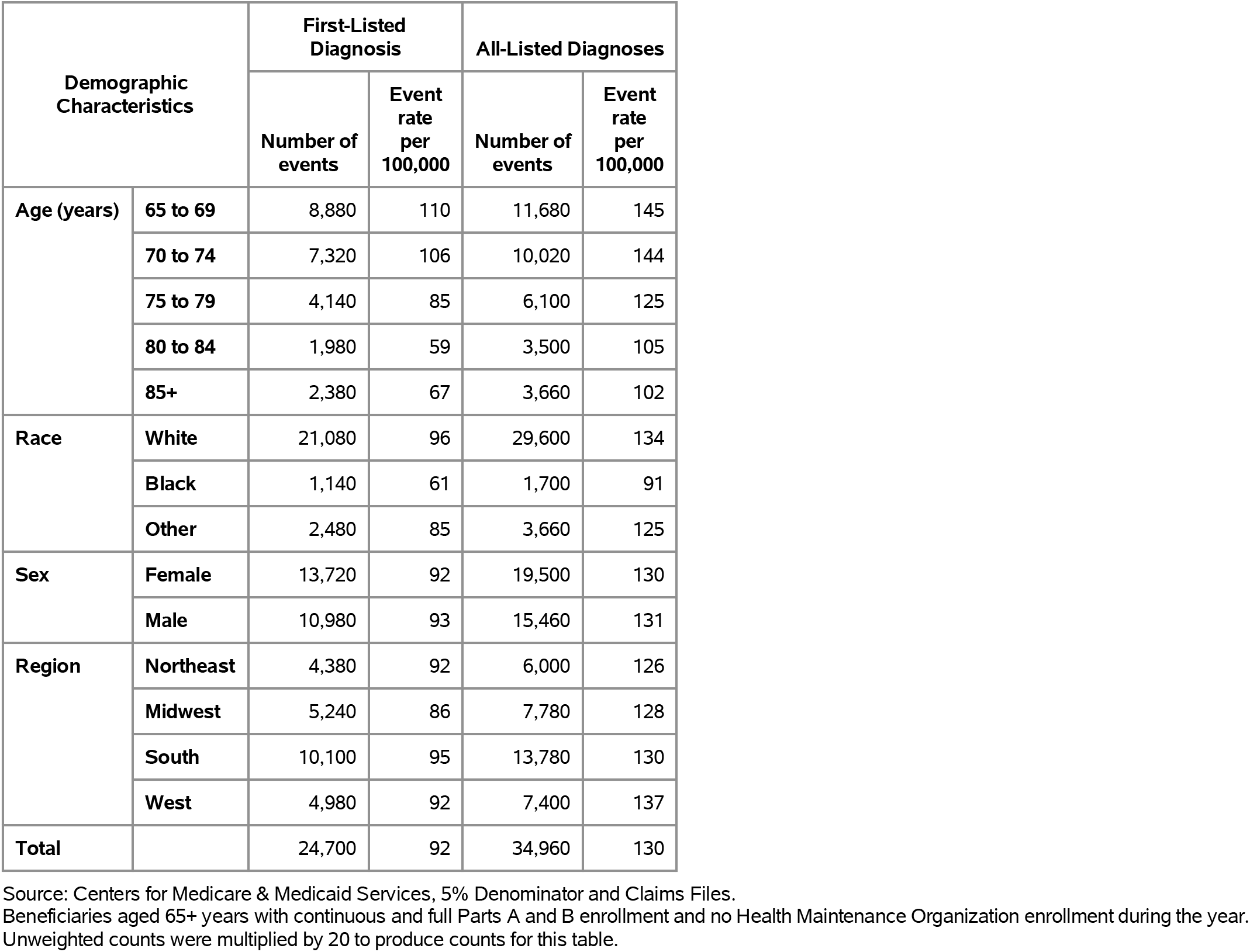
Appendicitis: Ambulatory care visits with first-listed and all-listed diagnoses by age, race, sex and region among fee-for-service, age-eligible Medicare beneficiaries, 2019.

| Demographic Characteristics |  | First-Listed Diagnosis |  | All-Listed Diagnoses |  |
| --- | --- | --- | --- | --- | --- |
|  |  | Number of events | Event rate per 100,000 | Number of events | Event rate per 100,000 |
| Age (years) | 65 to 69 | 8,880 | 110 | 11,680 | 145 |
|  | 70 to 74 | 7,320 | 106 | 10,020 | 144 |
|  | 75 to 79 | 4,140 | 85 | 6,100 | 125 |
|  | 80 to 84 | 1,980 | 59 | 3,500 | 105 |
|  | 85+ | 2,380 | 67 | 3,660 | 102 |
| Race | White | 21,080 | 96 | 29,600 | 134 |
|  | Black | 1,140 | 61 | 1,700 | 91 |
|  | Other | 2,480 | 85 | 3,660 | 125 |
| Sex | Female | 13,720 | 92 | 19,500 | 130 |
|  | Male | 10,980 | 93 | 15,460 | 131 |
| Region | Northeast | 4,380 | 92 | 6,000 | 126 |
|  | Midwest | 5,240 | 86 | 7,780 | 128 |
|  | South | 10,100 | 95 | 13,780 | 130 |
|  | West | 4,980 | 92 | 7,400 | 137 |
| Total |  | 24,700 | 92 | 34,960 | 130 |

Source: Centers for Medicare & Medicaid Services, 5% Denominator and Claims Files. Beneficiaries aged 65+ years with continuous and full Parts A and B enrollment and no Health Maintenance Organization enrollment during the year. Unweighted counts were multiplied by 20 to produce counts for this table.

Among Medicare beneficiaries, emergency department visit rates with appendicitis (all-listed diagnoses) decreased with age until 85 years and were higher among men compared with women and Whites compared with Blacks (Table 59). Rates were highest in the West, followed by the South and Northeast, and lowest in the Midwest.

**Table 59:** Appendicitis: Emergency department visits with first-listed and all-listed diagnoses by age, race, sex and region among fee-for-service, age-eligible Medicare beneficiaries, 2019.

| Demographic Characteristics |  | First-Listed Diagnosis |  | All-Listed Diagnoses |  |
| --- | --- | --- | --- | --- | --- |
|  |  | Number of events | Event rate per 100,000 | Number of events | Event rate per 100,000 |
| Age (years) | 65 to 69 | 8,040 | 100 | 9,080 | 113 |
|  | 70 to 74 | 6,020 | 87 | 7,500 | 108 |
|  | 75 to 79 | 3,640 | 74 | 4,600 | 94 |
|  | 80 to 84 | 1,760 | 53 | 2,220 | 67 |
|  | 85+ | 1,940 | 54 | 2,600 | 73 |
| Race | White | 17,780 | 81 | 21,300 | 97 |
|  | Black | 1,000 | 54 | 1,360 | 73 |
|  | Other | 2,620 | 90 | 3,340 | 114 |
| Sex | Female | 11,680 | 78 | 13,980 | 93 |
|  | Male | 9,720 | 82 | 12,020 | 102 |
| Region | Northeast | 3,760 | 79 | 4,640 | 98 |
|  | Midwest | 4,280 | 71 | 5,220 | 86 |
|  | South | 8,640 | 82 | 10,500 | 99 |
|  | West | 4,720 | 87 | 5,640 | 105 |
| Total |  | 21,400 | 80 | 26,000 | 97 |

Source: Centers for Medicare & Medicaid Services, 5% Denominator and Claims Files. Beneficiaries aged 65+ years with continuous and full Parts A and B enrollment and no Health Maintenance Organization enrollment during the year. Unweighted counts were multiplied by 20 to produce counts for this table.

Among Medicare beneficiaries, hospital discharge rates with appendicitis (all-listed diagnoses) peaked among persons 70 to 74 years and were higher among men compared with women and Whites compared with Blacks (Table 60). Rates were highest in the West, followed by the South, then the Northeast, and lowest in the Midwest.

**Table 60:** Appendicitis: Hospital discharges with first-listed and all-listed diagnoses by age, race, sex and region among fee-for-service, age-eligible Medicare beneficiaries, 2019.

| Demographic Characteristics |  | First-Listed Diagnosis |  | All-Listed Diagnoses |  |
| --- | --- | --- | --- | --- | --- |
|  |  | Number of events | Event rate per 100,000 | Number of events | Event rate per 100,000 |
| Age (years) | 65 to 69 | 4,240 | 53 | 5,360 | 66 |
|  | 70 to 74 | 3,460 | 50 | 4,940 | 71 |
|  | 75 to 79 | 2,020 | 41 | 3,280 | 67 |
|  | 80 to 84 | 1,360 | 41 | 1,820 | 55 |
|  | 85+ | 1,040 | 29 | 1,920 | 54 |
| Race | White | 9,820 | 45 | 13,920 | 63 |
|  | Black | 760 | 41 | 1,100 | 59 |
|  | Other | 1,540 | 53 | 2,300 | 79 |
| Sex | Female | 6,600 | 44 | 9,180 | 61 |
|  | Male | 5,520 | 47 | 8,140 | 69 |
| Region | Northeast | 2,160 | 45 | 3,000 | 63 |
|  | Midwest | 2,540 | 42 | 3,560 | 59 |
|  | South | 4,840 | 46 | 6,960 | 66 |
|  | West | 2,580 | 48 | 3,800 | 70 |
| Total |  | 12,120 | 45 | 17,320 | 65 |

Source: Centers for Medicare & Medicaid Services, 5% Denominator and Claims Files. Beneficiaries aged 65+ years with continuous and full Parts A and B enrollment and no Health Maintenance Organization enrollment during the year. Unweighted counts were multiplied by 20 to produce counts for this table.

Abdominal wall hernia contributed to 5.1 million ambulatory visits (2015) (Table 61). Ambulatory care visit rates (all-listed diagnoses) were higher among children compared with adolescents and the youngest adults and then increased with age. Age-adjusted rates were higher among men compared with women, Blacks compared with Whites, and Hispanics compared with non-Hispanics.

**Table 61:**
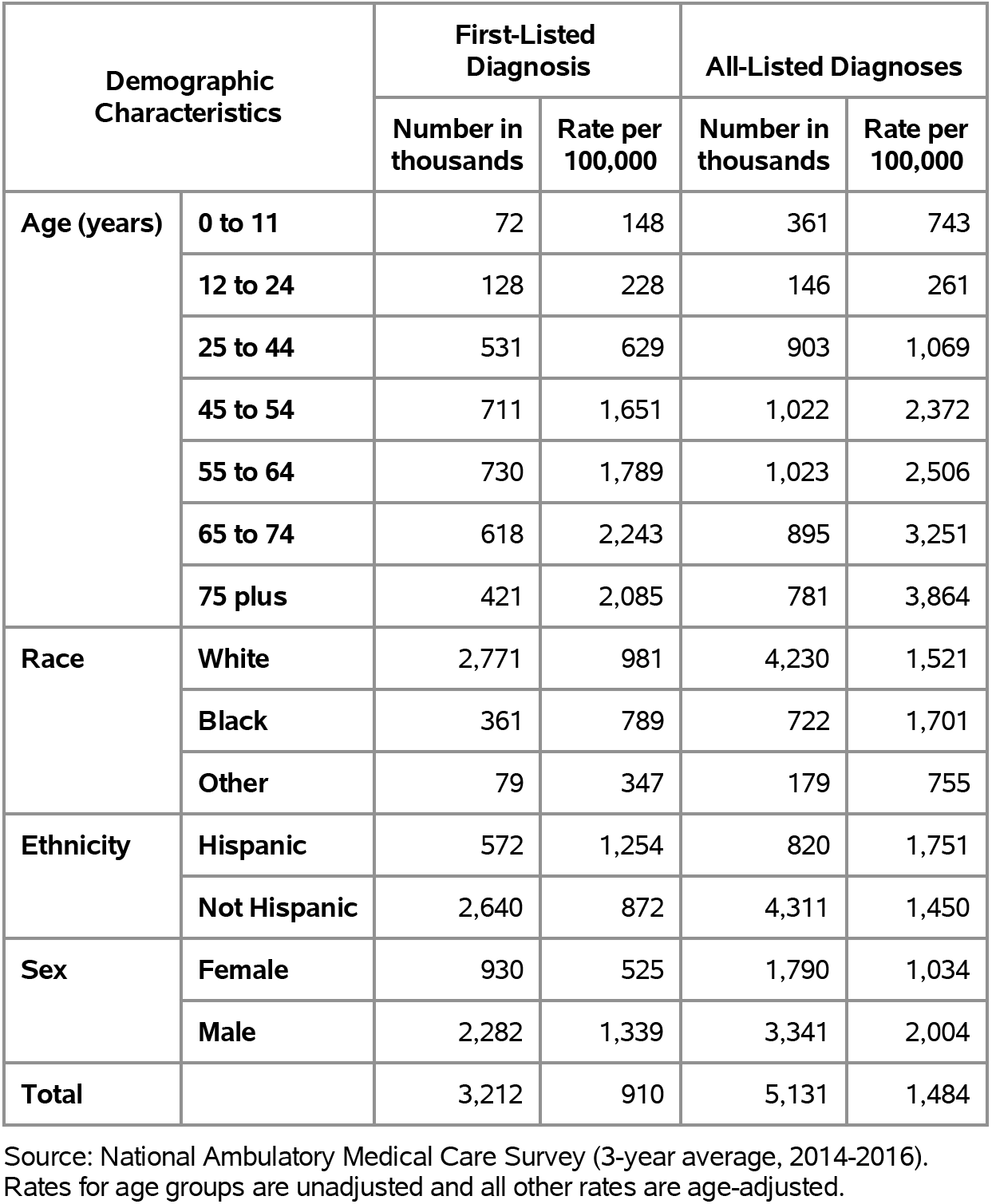
Abdominal Wall Hernia: Ambulatory care visits with first-listed and all-listed diagnoses by age, race, ethnicity, and sex in the United States, 2015.

Abdominal wall hernia contributed to 761,000 emergency department visits in 2018 (Table 62). Emergency department visit rates (all-listed diagnoses) were higher among children compared with adolescents and the youngest adults and then increased with age. Age-adjusted rates were higher among men compared with women.

**Table 62:** Abdominal Wall Hernia: Emergency department visits with first-listed and all-listed diagnoses by age and sex in the United States, 2018.

| Demographic Characteristics |  | First-Listed Diagnosis |  | All-Listed Diagnoses |  |
| --- | --- | --- | --- | --- | --- |
|  |  | Number in thousands | Rate per 100,000 | Number in thousands | Rate per 100,000 |
| Age (years) | 0 to 11 | 15 | 31 | 35 | 72 |
|  | 12 to 24 | 12 | 22 | 21 | 38 |
|  | 25 to 44 | 82 | 94 | 159 | 183 |
|  | 45 to 54 | 59 | 141 | 130 | 313 |
|  | 55 to 64 | 60 | 141 | 148 | 351 |
|  | 65 to 74 | 43 | 141 | 121 | 397 |
|  | 75 plus | 48 | 218 | 146 | 665 |
| Sex | Female | 115 | 64 | 304 | 165 |
|  | Male | 203 | 121 | 457 | 269 |
| Total |  | 318 | 92 | 761 | 214 |

Source: Healthcare Cost and Utilization Project Nationwide Emergency Department Sample. Rates for age groups are unadjusted and all other rates are age-adjusted.

Abdominal wall hernia contributed to 452,000 hospital discharges in 2018 (Table 63). Hospital discharge rates (all-listed diagnoses) were higher among children compared with adolescents and the youngest adults and then increased with age. Age-adjusted rates were higher among men compared with women and Blacks compared with Whites but did not differ by ethnicity. Between 2004 and 2018, age-adjusted hospital discharge rates (per 100,000) with an all-listed diagnosis decreased by 5% from 127 to 121.(4,6)

**Table 63:** Abdominal Wall Hernia: Hospital discharges with first-listed and all-listed diagnoses by age, race, ethnicity, and sex in the United States, 2018.

| Demographic Characteristics |  | First-Listed Diagnosis |  | All-Listed Diagnoses |  |
| --- | --- | --- | --- | --- | --- |
|  |  | Number in thousands | Rate per 100,000 | Number in thousands | Rate per 100,000 |
| Age (years) | 0 to 11 | 2 | 5 | 27 | 56 |
|  | 12 to 24 | 1 | 2 | 4 | 8 |
|  | 25 to 44 | 16 | 18 | 51 | 58 |
|  | 45 to 54 | 22 | 53 | 65 | 155 |
|  | 55 to 64 | 33 | 77 | 100 | 237 |
|  | 65 to 74 | 31 | 100 | 97 | 319 |
|  | 75 plus | 31 | 143 | 108 | 491 |
| Race | White | 113 | 37 | 364 | 119 |
|  | Black | 17 | 39 | 68 | 153 |
|  | Other | 6 | 25 | 23 | 92 |
| Ethnicity | Hispanic | 17 | 37 | 57 | 122 |
|  | Not Hispanic | 120 | 36 | 399 | 123 |
| Sex | Female | 71 | 36 | 210 | 108 |
|  | Male | 64 | 36 | 242 | 138 |
| Total |  | 135 | 36 | 452 | 121 |

Source: Healthcare Cost and Utilization Project National Inpatient Sample. Rates for age groups are unadjusted and all other rates are age-adjusted.

Abdominal wall hernia contributed to 2,000 deaths in 2019 (Table 64). Mortality was uncommon among the youngest age groups after which rates (underlying or other cause) increased with age. Age-adjusted mortality rates were higher among men, Blacks, and non-Hispanics. Between 2004 and 2019, age-adjusted mortality rates (per 100,000) with abdominal hernia as underlying or other cause decreased by 14% from 0.7 to 0.6.(4)

**Table 64:** Abdominal Wall Hernia: Deaths with underlying or underlying/other cause and lifetime years of life lost by age, race, ethnicity, and sex in the United States, 2019.

| Demographic Characteristics |  | Underlying Cause |  |  | Underlying/Other Cause |  |  |
| --- | --- | --- | --- | --- | --- | --- | --- |
|  |  | Number of Deaths | Rate per 100,000 | Years of potential life lost in thousands | Number of Deaths | Rate per 100,000 | Years of potential life lost in thousands |
| Age (years) | 0 to 11 | 12 | 0.0 | 0.9 | 22 | 0.0 | 1.6 |
|  | 12 to 24 | 4 | 0.0 | 0.2 | 9 | 0.0 | 0.5 |
|  | 25 to 44 | 34 | 0.0 | 1.4 | 51 | 0.1 | 2.1 |
|  | 45 to 54 | 87 | 0.2 | 2.7 | 154 | 0.4 | 4.8 |
|  | 55 to 64 | 209 | 0.5 | 4.7 | 343 | 0.8 | 7.8 |
|  | 65 to 74 | 328 | 1.0 | 5.1 | 548 | 1.7 | 8.6 |
|  | 75 plus | 884 | 3.9 | 6.0 | 1,345 | 6.0 | 9.2 |
| Race | White | 1,352 | 0.4 | 17.8 | 2,124 | 0.6 | 28.9 |
|  | Black | 166 | 0.4 | 2.7 | 272 | 0.7 | 4.5 |
|  | Other | 40 | 0.2 | 0.7 | 76 | 0.3 | 1.2 |
| Ethnicity | Hispanic | 110 | 0.3 | 1.9 | 183 | 0.5 | 3.3 |
|  | Not Hispanic | 1,448 | 0.4 | 19.2 | 2,289 | 0.6 | 31.2 |
| Sex | Female | 766 | 0.3 | 9.9 | 1,133 | 0.5 | 15.3 |
|  | Male | 792 | 0.5 | 11.2 | 1,339 | 0.8 | 19.3 |
| Total |  | 1,558 | 0.4 | 21.1 | 2,472 | 0.6 | 34.6 |

Source: Vital Statistics of the United States.
Rates for age groups are unadjusted and all other rates are age-adjusted.
Years of potential life lost is based on U.S. life table 2006-2018 estimates of life expectancy at age of death.

Among privately insured enrollees, the claims-based prevalence of abdominal wall hernia (based on all-listed diagnoses) was 1.1% (Table 65). Prevalence was higher among children compared with adolescents and younger adults and then increased with age. It was higher among men. It was similar among Whites, Blacks, and Hispanics, and lowest among Asians. It differed little by region.

**Table 65:** Abdominal Wall Hernia: Claims-based prevalence with first-listed and all-listed diagnoses by age, race-ethnicity, sex and region among privately insured enrollees, 2020.

| Demographic Characteristics |  | Number of enrollees | First-Listed Diagnosis |  | All-Listed Diagnoses |  |
| --- | --- | --- | --- | --- | --- | --- |
|  |  |  | Number of enrollees with disease | Percent of enrollees with disease | Number of enrollees with disease | Percent of enrollees with disease |
| Age (years) | 0 to 11 | 1,101,978 | 2,295 | 0.2% | 7,404 | 0.7% |
|  | 12 to 24 | 1,451,892 | 1,172 | 0.1% | 1,650 | 0.1% |
|  | 25 to 44 | 2,656,672 | 9,438 | 0.4% | 14,243 | 0.5% |
|  | 45 to 54 | 1,452,320 | 9,486 | 0.7% | 15,840 | 1.1% |
|  | 55 to 64 | 1,640,216 | 14,120 | 0.9% | 24,223 | 1.5% |
|  | 65 to 74 | 2,840,615 | 25,399 | 0.9% | 43,577 | 1.5% |
|  | 75 plus | 2,576,241 | 21,367 | 0.8% | 40,099 | 1.6% |
| Race-ethnicity | White | 8,657,702 | 58,240 | 0.7% | 98,395 | 1.1% |
|  | Black | 1,242,477 | 7,471 | 0.6% | 13,666 | 1.1% |
|  | Hispanic | 1,493,457 | 8,382 | 0.6% | 15,699 | 1.1% |
|  | Asian | 607,284 | 1,488 | 0.2% | 2,675 | 0.4% |
|  | Unknown | 1,719,014 | 7,696 | 0.4% | 16,601 | 1.0% |
| Sex | Female | 7,199,363 | 25,247 | 0.4% | 48,911 | 0.7% |
|  | Male | 6,520,571 | 58,030 | 0.9% | 98,125 | 1.5% |
| Region | Northeast | 1,526,275 | 10,040 | 0.7% | 17,758 | 1.2% |
|  | Midwest | 3,322,846 | 19,518 | 0.6% | 33,129 | 1.0% |
|  | South | 5,619,435 | 34,651 | 0.6% | 62,930 | 1.1% |
|  | West | 3,251,378 | 19,068 | 0.6% | 33,219 | 1.0% |
| Total |  | 13,719,934 | 83,277 | 0.6% | 147,036 | 1.1% |

Source: De-identified Optum Clinformatic® Data Mart.
Enrollees with full enrollment in a commercial health plan or Medicare during the year.

Among commercial insurance enrollees, ambulatory care visit rates with abdominal wall hernia (all-listed diagnoses) were higher among children compared with adolescents and younger adults and then increased with age until 75 years (Table 66). They were higher among men compared with women. Among persons with known race-ethnicity, rates were highest among Whites, followed by Blacks, then Hispanics, and lowest among Asians. Rates were highest in the Northeast, followed by the South, then the West, and lowest in the Midwest.

**Table 66:** Abdominal Wall Hernia: Ambulatory care visits with first-listed and all-listed diagnoses by age, race-ethnicity, sex and region among privately insured enrollees, 2020.

| Demographic Characteristics |  | First-Listed Diagnosis |  | All-Listed Diagnoses |  |
| --- | --- | --- | --- | --- | --- |
|  |  | Number of events | Event rate per 100,000 | Number of events | Event rate per 100,000 |
| Age (years) | 0 to 11 | 3,646 | 331 | 11,690 | 1,061 |
|  | 12 to 24 | 2,088 | 144 | 2,869 | 198 |
|  | 25 to 44 | 17,177 | 647 | 26,042 | 980 |
|  | 45 to 54 | 17,590 | 1,211 | 29,299 | 2,017 |
|  | 55 to 64 | 25,941 | 1,582 | 44,819 | 2,733 |
|  | 65 to 74 | 45,825 | 1,613 | 79,931 | 2,814 |
|  | 75 plus | 37,007 | 1,436 | 69,688 | 2,705 |
| Race-ethnicity | White | 105,416 | 1,218 | 178,404 | 2,061 |
|  | Black | 12,994 | 1,046 | 24,102 | 1,940 |
|  | Hispanic | 14,757 | 988 | 28,056 | 1,879 |
|  | Asian | 2,750 | 453 | 4,822 | 794 |
|  | Unknown | 13,357 | 777 | 28,954 | 1,684 |
| Sex | Female | 41,413 | 575 | 82,451 | 1,145 |
|  | Male | 107,861 | 1,654 | 181,887 | 2,789 |
| Region | Northeast | 17,803 | 1,166 | 31,852 | 2,087 |
|  | Midwest | 34,524 | 1,039 | 60,305 | 1,815 |
|  | South | 62,477 | 1,112 | 111,778 | 1,989 |
|  | West | 34,470 | 1,060 | 60,403 | 1,858 |
| Total |  | 149,274 | 1,088 | 264,338 | 1,927 |

Source: De-identified Optum Clinformatic® Data Mart.
Enrollees with full enrollment in a commercial health plan or Medicare during the year.

Among commercial insurance enrollees, emergency department visit rates with abdominal wall hernia (all-listed diagnoses) increased with age and were higher among men compared with women (Table 67). Among persons with known race-ethnicity, rates were highest among Blacks, followed by Hispanics and Whites, and much lower among Asians. Rates were highest in the Northeast, followed by the South, then the West, and lowest in the Midwest.

**Table 67:** Abdominal Wall Hernia: Emergency department visits with first-listed and all-listed diagnoses by age, race-ethnicity, sex and region among privately insured enrollees, 2020.

| Demographic Characteristics |  | First-Listed Diagnosis |  | All-Listed Diagnoses |  |
| --- | --- | --- | --- | --- | --- |
|  |  | Number of events | Event rate per 100,000 | Number of events | Event rate per 100,000 |
| Age (years) | 0 to 11 | 14 | 1 | 22 | 2 |
|  | 12 to 24 | 8 | 1 | 10 | 1 |
|  | 25 to 44 | 161 | 6 | 315 | 12 |
|  | 45 to 54 | 225 | 15 | 433 | 30 |
|  | 55 to 64 | 615 | 37 | 1,156 | 70 |
|  | 65 to 74 | 1,832 | 64 | 3,394 | 119 |
|  | 75 plus | 2,586 | 100 | 4,676 | 182 |
| Race-ethnicity | White | 3,588 | 41 | 6,501 | 75 |
|  | Black | 708 | 57 | 1,335 | 107 |
|  | Hispanic | 591 | 40 | 1,176 | 79 |
|  | Asian | 69 | 11 | 121 | 20 |
|  | Unknown | 485 | 28 | 873 | 51 |
| Sex | Female | 2,042 | 28 | 4,208 | 58 |
|  | Male | 3,399 | 52 | 5,798 | 89 |
| Region | Northeast | 820 | 54 | 1,294 | 85 |
|  | Midwest | 1,155 | 35 | 2,020 | 61 |
|  | South | 2,181 | 39 | 4,319 | 77 |
|  | West | 1,285 | 40 | 2,373 | 73 |
| Total |  | 5,441 | 40 | 10,006 | 73 |

Source: De-identified Optum Clinformatics® Data Mart.
Enrollees with full enrollment in a commercial health plan or Medicare during the year.

Among commercial insurance enrollees, hospital discharge rates with abdominal wall hernia (all-listed diagnoses) were higher among children compared with adolescents and younger adults and then increased with age and were higher among men compared with women (Table 68). Among persons with known race-ethnicity, rates were highest among Blacks, followed by Hispanics and Whites, and much lower among Asians. Rates were highest in the Northeast, followed by the South, and lowest in the Midwest and West.

**Table 68:**
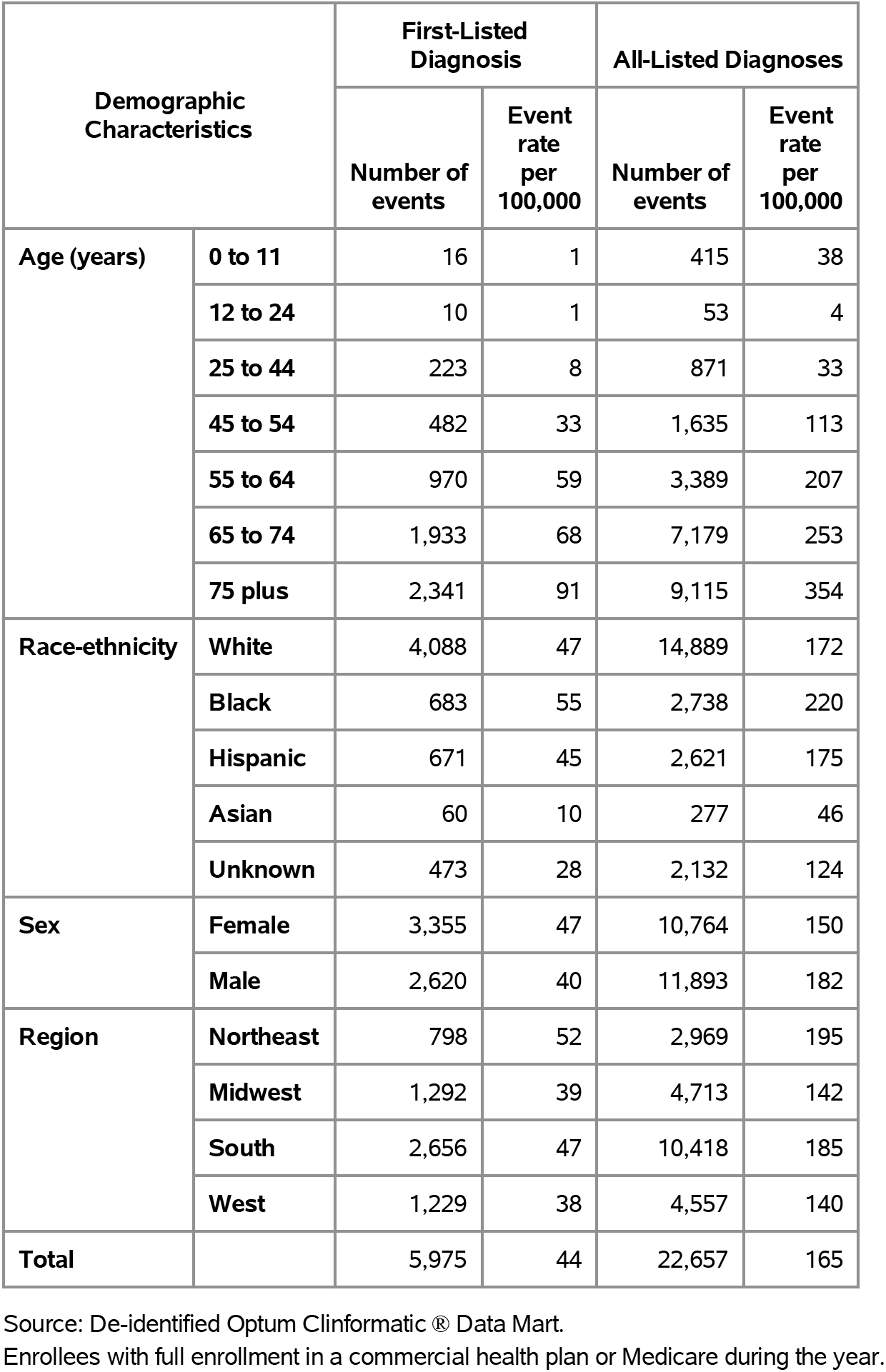
Abdominal Wall Hernia: Hospital discharges with first-listed and all-listed diagnoses by age, race-ethnicity, sex and region among privately insured enrollees, 2020.

| Demographic Characteristics |  | First-Listed Diagnosis |  | All-Listed Diagnoses |  |
| --- | --- | --- | --- | --- | --- |
|  |  | Number of events | Event rate per 100,000 | Number of events | Event rate per 100,000 |
| Age (years) | 0 to 11 | 16 | 1 | 415 | 38 |
|  | 12 to 24 | 10 | 1 | 53 | 4 |
|  | 25 to 44 | 223 | 8 | 871 | 33 |
|  | 45 to 54 | 482 | 33 | 1,635 | 113 |
|  | 55 to 64 | 970 | 59 | 3,389 | 207 |
|  | 65 to 74 | 1,933 | 68 | 7,179 | 253 |
|  | 75 plus | 2,341 | 91 | 9,115 | 354 |
| Race-ethnicity | White | 4,088 | 47 | 14,889 | 172 |
|  | Black | 683 | 55 | 2,738 | 220 |
|  | Hispanic | 671 | 45 | 2,621 | 175 |
|  | Asian | 60 | 10 | 277 | 46 |
|  | Unknown | 473 | 28 | 2,132 | 124 |
| Sex | Female | 3,355 | 47 | 10,764 | 150 |
|  | Male | 2,620 | 40 | 11,893 | 182 |
| Region | Northeast | 798 | 52 | 2,969 | 195 |
|  | Midwest | 1,292 | 39 | 4,713 | 142 |
|  | South | 2,656 | 47 | 10,418 | 185 |
|  | West | 1,229 | 38 | 4,557 | 140 |
| Total |  | 5,975 | 44 | 22,657 | 165 |

Source: De-identified Optum Clinformatics® Data Mart.
Enrollees with full enrollment in a commercial health plan or Medicare during the year.

Among Medicare beneficiaries, the claims-based prevalence of abdominal wall hernia (based on all-listed diagnoses) was 2.3% (Table 69). Prevalence was highest among persons 75-84 years and was higher among men and Whites. It was higher in the Northeast compared to other regions.

**Table 69:** Abdominal Wall Hernia: Claims-based prevalence with first-listed and all-listed diagnoses by age, race, sex and region among fee-for-service, age-eligible Medicare beneficiaries, 2019.

| Demographic Characteristics |  | Number of enrollees | First-Listed Diagnosis |  | All-Listed Diagnoses |  |
| --- | --- | --- | --- | --- | --- | --- |
|  |  |  | Number of enrollees with disease | Percent of enrollees with disease | Number of enrollees with disease | Percent of enrollees with disease |
| Age (years) | 65 to 69 | 8,066,320 | 97,040 | 1.2% | 172,600 | 2.1% |
|  | 70 to 74 | 6,936,940 | 88,460 | 1.3% | 160,700 | 2.3% |
|  | 75 to 79 | 4,894,060 | 64,960 | 1.3% | 124,660 | 2.5% |
|  | 80 to 84 | 3,335,340 | 42,360 | 1.3% | 83,440 | 2.5% |
|  | 85+ | 3,578,120 | 37,000 | 1.0% | 83,860 | 2.3% |
| Race | White | 22,023,800 | 279,360 | 1.3% | 527,980 | 2.4% |
|  | Black | 1,861,660 | 17,580 | 0.9% | 36,220 | 1.9% |
|  | Other | 2,925,320 | 32,880 | 1.1% | 61,060 | 2.1% |
| Sex | Female | 14,975,560 | 104,140 | 0.7% | 218,560 | 1.5% |
|  | Male | 11,835,220 | 225,680 | 1.9% | 406,700 | 3.4% |
| Region | Northeast | 4,748,700 | 61,540 | 1.3% | 121,220 | 2.6% |
|  | Midwest | 6,069,800 | 71,240 | 1.2% | 134,160 | 2.2% |
|  | South | 10,595,900 | 127,460 | 1.2% | 244,080 | 2.3% |
|  | West | 5,396,380 | 69,580 | 1.3% | 125,800 | 2.3% |
| Total |  | 26,810,780 | 329,820 | 1.2% | 625,260 | 2.3% |

Source: Centers for Medicare & Medicaid Services, 5% Denominator and Claims Files. Beneficiaries aged 65+ years with continuous and full Parts A and B enrollment and no Health Maintenance Organization enrollment during the year. Unweighted counts were multiplied by 20 to produce counts for this table.

Among Medicare beneficiaries, ambulatory care visit rates with abdominal wall hernia (all-listed diagnoses) peaked among persons 75 to 79 years and were over twice as high among men compared with women and Whites compared with Blacks (Table 70). Rates were highest in the Northeast, followed by the West, then the South, and lowest in the Midwest.

**Table 70:** Abdominal Wall Hernia: Ambulatory care visits with first-listed and all-listed diagnoses by age, race, sex and region among fee-for-service, age-eligible Medicare beneficiaries, 2019.

| Demographic Characteristics |  | First-Listed Diagnosis |  | All-Listed Diagnoses |  |
| --- | --- | --- | --- | --- | --- |
|  |  | Number of events | Event rate per 100,000 | Number of events | Event rate per 100,000 |
| Age (years) | 65 to 69 | 176,900 | 2,193 | 313,600 | 3,888 |
|  | 70 to 74 | 161,920 | 2,334 | 291,860 | 4,207 |
|  | 75 to 79 | 116,000 | 2,370 | 218,780 | 4,470 |
|  | 80 to 84 | 73,800 | 2,213 | 144,340 | 4,328 |
|  | 85+ | 54,140 | 1,513 | 119,120 | 3,329 |
| Race | White | 496,400 | 2,254 | 919,520 | 4,175 |
|  | Black | 27,700 | 1,488 | 57,620 | 3,095 |
|  | Other | 58,660 | 2,005 | 110,560 | 3,779 |
| Sex | Female | 164,580 | 1,099 | 342,560 | 2,287 |
|  | Male | 418,180 | 3,533 | 745,140 | 6,296 |
| Region | Northeast | 109,020 | 2,296 | 211,560 | 4,455 |
|  | Midwest | 123,680 | 2,038 | 231,100 | 3,807 |
|  | South | 227,320 | 2,145 | 416,720 | 3,933 |
|  | West | 122,740 | 2,274 | 228,320 | 4,231 |
| Total |  | 582,760 | 2,174 | 1,087,700 | 4,057 |

Source: Centers for Medicare & Medicaid Services, 5% Denominator and Claims Files. Beneficiaries aged 65+ years with continuous and full Parts A and B enrollment and no Health Maintenance Organization enrollment during the year. Unweighted counts were multiplied by 20 to produce counts for this table.

Among Medicare beneficiaries, emergency department visit rates with abdominal wall hernia (all-listed diagnoses) increased with age and were higher among men compared with women and Blacks compared with Whites (Table 71). Rates were highest in the Northeast, followed by the South, and lowest in the Midwest and West.

**Table 71:** Abdominal Wall Hernia: Emergency department visits with first-listed and all-listed diagnoses by age, race, sex and region among fee-for-service, age-eligible Medicare beneficiaries, 2019.

| Demographic Characteristics |  | First-Listed Diagnosis |  | All-Listed Diagnoses |  |
| --- | --- | --- | --- | --- | --- |
|  |  | Number of events | Event rate per 100,000 | Number of events | Event rate per 100,000 |
| Age (years) | 65 to 69 | 13,280 | 165 | 37,340 | 463 |
|  | 70 to 74 | 13,900 | 200 | 38,220 | 551 |
|  | 75 to 79 | 11,300 | 231 | 33,340 | 681 |
|  | 80 to 84 | 8,900 | 267 | 27,800 | 833 |
|  | 85+ | 12,180 | 340 | 37,160 | 1,039 |
| Race | White | 48,080 | 218 | 141,060 | 640 |
|  | Black | 5,520 | 297 | 15,820 | 850 |
|  | Other | 5,960 | 204 | 16,980 | 580 |
| Sex | Female | 25,280 | 169 | 76,720 | 512 |
|  | Male | 34,280 | 290 | 97,140 | 821 |
| Region | Northeast | 12,420 | 262 | 34,600 | 729 |
|  | Midwest | 13,420 | 221 | 37,060 | 611 |
|  | South | 22,140 | 209 | 69,420 | 655 |
|  | West | 11,580 | 215 | 32,780 | 607 |
| Total |  | 59,560 | 222 | 173,860 | 648 |

Source: Centers for Medicare & Medicaid Services, 5% Denominator and Claims Files. Beneficiaries aged 65+ years with continuous and full Parts A and B enrollment and no Health Maintenance Organization enrollment during the year. Unweighted counts were multiplied by 20 to produce counts for this table.

Among Medicare beneficiaries, hospital discharge rates with abdominal wall hernia (all-listed diagnoses) increased with age and were higher among men compared with women and Blacks compared with Whites (Table 72). Rates were highest in the Northeast, followed by the South, then the Midwest, and lowest in the West.

**Table 72:** Abdominal Wall Hernia: Hospital discharges with first-listed and all-listed diagnoses by age, race, sex and region among fee-for-service, age-eligible Medicare beneficiaries, 2019.

| Demographic Characteristics |  | First-Listed Diagnosis |  | All-Listed Diagnoses |  |
| --- | --- | --- | --- | --- | --- |
|  |  | Number of events | Event rate per 100,000 | Number of events | Event rate per 100,000 |
| Age (years) | 65 to 69 | 7,840 | 97 | 26,380 | 327 |
|  | 70 to 74 | 7,780 | 112 | 26,140 | 377 |
|  | 75 to 79 | 6,560 | 134 | 23,440 | 479 |
|  | 80 to 84 | 5,140 | 154 | 18,140 | 544 |
|  | 85+ | 5,640 | 158 | 21,720 | 607 |
| Race | White | 27,220 | 124 | 95,320 | 433 |
|  | Black | 2,540 | 136 | 9,500 | 510 |
|  | Other | 3,200 | 109 | 11,000 | 376 |
| Sex | Female | 18,060 | 121 | 55,960 | 374 |
|  | Male | 14,900 | 126 | 59,860 | 506 |
| Region | Northeast | 6,500 | 137 | 21,880 | 461 |
|  | Midwest | 7,380 | 122 | 25,740 | 424 |
|  | South | 13,520 | 128 | 47,640 | 450 |
|  | West | 5,560 | 103 | 20,560 | 381 |
| Total |  | 32,960 | 123 | 115,820 | 432 |

Source: Centers for Medicare & Medicaid Services, 5% Denominator and Claims Files. Beneficiaries aged 65+ years with continuous and full Parts A and B enrollment and no Health Maintenance Organization enrollment during the year. Unweighted counts were multiplied by 20 to produce counts for this table.

Crohn’s disease contributed to 1.0 million ambulatory visits (2015) (Table 73). Ambulatory care visit rates (all-listed diagnoses) were highest among persons 65-74 years. Age-adjusted rates were higher among women compared with men, Whites compared with Blacks, and non-Hispanics compared with Hispanics.

**Table 73:** Crohns Disease: Ambulatory care visits with first-listed and all-listed diagnoses by age, race, ethnicity, and sex in the United States, 2015.

| Demographic Characteristics |  | First-Listed Diagnosis |  | All-Listed Diagnoses |  |
| --- | --- | --- | --- | --- | --- |
|  |  | Number in thousands | Rate per 100,000 | Number in thousands | Rate per 100,000 |
| Age (years) | 0 to 11 | . | . | 1 | 2 |
|  | 12 to 24 | 26 | 46 | 63 | 112 |
|  | 25 to 44 | 148 | 176 | 244 | 288 |
|  | 45 to 54 | 110 | 255 | 257 | 596 |
|  | 55 to 64 | 110 | 269 | 211 | 516 |
|  | 65 to 74 | 144 | 524 | 213 | 774 |
|  | 75 plus | 8 | 41 | 54 | 270 |
| Race | White | 445 | 159 | 827 | 294 |
|  | Black | 31 | 58 | 105 | 240 |
|  | Other | 71 | 289 | 111 | 518 |
| Ethnicity | Hispanic | 41 | 63 | 92 | 227 |
|  | Not Hispanic | 506 | 169 | 951 | 318 |
| Sex | Female | 319 | 169 | 604 | 323 |
|  | Male | 228 | 141 | 438 | 276 |
| Total |  | 547 | 156 | 1,043 | 300 |

Source: National Ambulatory Medical Care Survey (3-year average, 2014-2016). Rates for age groups are unadjusted and all other rates are age-adjusted.

Crohn’s disease contributed to 375,000 emergency department visits in 2018 (Table 74). Emergency department visit rates (all-listed diagnoses) were highest among persons 25-44 years. Age-adjusted rates were higher among women compared with men.

**Table 74:** Crohns Disease: Emergency department visits with first-listed and all-listed diagnoses by age and sex in the United States, 2018.

| Demographic Characteristics |  | First-Listed Diagnosis |  | All-Listed Diagnoses |  |
| --- | --- | --- | --- | --- | --- |
|  |  | Number in thousands | Rate per 100,000 | Number in thousands | Rate per 100,000 |
| Age (years) | 0 to 11 | 1 | 2 | 3 | 6 |
|  | 12 to 24 | 15 | 28 | 38 | 68 |
|  | 25 to 44 | 45 | 52 | 143 | 164 |
|  | 45 to 54 | 14 | 34 | 63 | 150 |
|  | 55 to 64 | 10 | 22 | 53 | 124 |
|  | 65 to 74 | 6 | 19 | 41 | 135 |
|  | 75 plus | 3 | 16 | 35 | 160 |
| Sex | Female | 51 | 31 | 220 | 129 |
|  | Male | 44 | 28 | 155 | 95 |
| Total |  | 95 | 30 | 375 | 113 |

Source: Healthcare Cost and Utilization Project Nationwide Emergency Department Sample. Rates for age groups are unadjusted and all other rates are age-adjusted.

Crohn’s disease contributed to 207,000 hospital discharges in 2018 (Table 75). Hospital discharge rates (all-listed diagnoses) increased with age. Age-adjusted rates were higher among women compared with men, Whites compared with Blacks, and non-Hispanics compared with Hispanics. Between 2004 and 2018, age-adjusted hospital discharge rates (per 100,000) with an all-listed diagnosis increased by 23% from 48 to 59.(4)

**Table 75:** Crohns Disease: Hospital discharges with first-listed and all-listed diagnoses by age, race, ethnicity, and sex in the United States, 2018.

| Demographic Characteristics |  | First-Listed Diagnosis |  | All-Listed Diagnoses |  |
| --- | --- | --- | --- | --- | --- |
|  |  | Number in thousands | Rate per 100,000 | Number in thousands | Rate per 100,000 |
| Age (years) | 0 to 11 | 1 | 2 | 1 | 3 |
|  | 12 to 24 | 10 | 18 | 18 | 32 |
|  | 25 to 44 | 25 | 28 | 60 | 69 |
|  | 45 to 54 | 10 | 23 | 32 | 76 |
|  | 55 to 64 | 7 | 18 | 35 | 83 |
|  | 65 to 74 | 5 | 17 | 34 | 110 |
|  | 75 plus | 3 | 14 | 27 | 124 |
| Race | White | 49 | 19 | 177 | 64 |
|  | Black | 9 | 20 | 25 | 55 |
|  | Other | 3 | 10 | 7 | 27 |
| Ethnicity | Hispanic | 4 | 8 | 12 | 23 |
|  | Not Hispanic | 57 | 21 | 197 | 67 |
| Sex | Female | 33 | 20 | 119 | 66 |
|  | Male | 28 | 18 | 88 | 52 |
| Total |  | 61 | 19 | 207 | 59 |

Source: Healthcare Cost and Utilization Project National Inpatient Sample. Rates for age groups are unadjusted and all other rates are age-adjusted.

Crohn’s disease contributed to 2,000 deaths in 2019 (Table 76). Mortality was uncommon among the youngest age groups after which rates (underlying or other cause) increased with age. Age-adjusted mortality rates were higher among men, Whites, and non-Hispanics. Between 2004 and 2019, age-adjusted mortality rates (per 100,000) with Crohn’s disease as underlying or other cause increased by 20% from 0.5 to 0.6.(4)

**Table 76:** Crohns Disease: Deaths with underlying or underlying/other cause and lifetime years of life lost by age, race, ethnicity, and sex in the United States, 2019.

| Demographic Characteristics |  | Underlying Cause |  |  | Underlying/Other Cause |  |  |
| --- | --- | --- | --- | --- | --- | --- | --- |
|  |  | Number of Deaths | Rate per 100,000 | Years of potential life lost in thousands | Number of Deaths | Rate per 100,000 | Years of potential life lost in thousands |
| Age (years) | 0 to 11 | . | . | . | . | . | . |
|  | 12 to 24 | 4 | 0.0 | 0.2 | 8 | 0.0 | 0.5 |
|  | 25 to 44 | 57 | 0.1 | 2.4 | 141 | 0.2 | 6.0 |
|  | 45 to 54 | 71 | 0.2 | 2.2 | 201 | 0.5 | 6.2 |
|  | 55 to 64 | 100 | 0.2 | 2.3 | 360 | 0.8 | 8.4 |
|  | 65 to 74 | 183 | 0.6 | 2.9 | 576 | 1.8 | 9.1 |
|  | 75 plus | 264 | 1.2 | 2.1 | 899 | 4.0 | 6.9 |
| Race | White | 625 | 0.2 | 10.9 | 2,002 | 0.6 | 33.2 |
|  | Black | 45 | 0.1 | 1.1 | 151 | 0.4 | 3.3 |
|  | Other | 9 | 0.0 | 0.2 | 32 | 0.1 | 0.6 |
| Ethnicity | Hispanic | 15 | 0.0 | 0.3 | 45 | 0.1 | 0.9 |
|  | Not Hispanic | 664 | 0.2 | 11.8 | 2,140 | 0.6 | 36.2 |
| Sex | Female | 394 | 0.2 | 6.8 | 1,202 | 0.5 | 20.6 |
|  | Male | 285 | 0.2 | 5.3 | 983 | 0.6 | 16.5 |
| Total |  | 679 | 0.2 | 12.1 | 2,185 | 0.6 | 37.1 |

Source: Vital Statistics of the United States.
Rates for age groups are unadjusted and all other rates are age-adjusted.
Years of potential life lost is based on U.S. life table 2006-2018 estimates of life expectancy at age of death.

Among privately insured enrollees, the claims-based prevalence of Crohn’s disease (based on all-listed diagnoses) was 0.3% (Table 77). Prevalence was highest in middle age and did not differ by sex. It was highest among Whites and Blacks, followed by Hispanics, and lowest among Asians. It differed little by region.

**Table 77:**
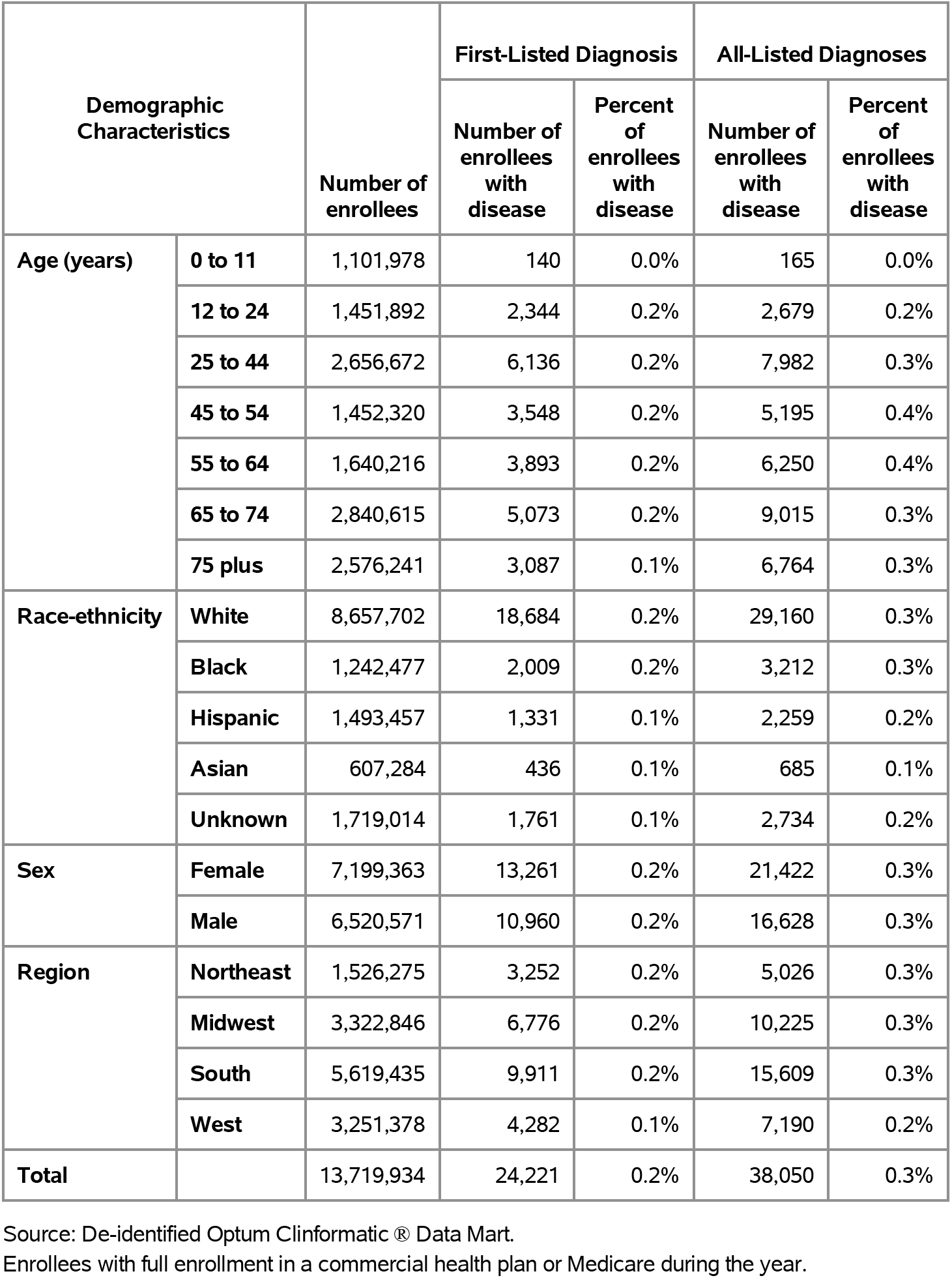
Crohns Disease: Claims-based prevalence with first-listed and all-listed diagnoses by age, race-ethnicity, sex and region among privately insured enrollees, 2020.

Among commercial insurance enrollees, ambulatory care visit rates with Crohn’s disease (all-listed diagnoses) peaked among persons 55 to 64 years and were higher among women compared with men (Table 78). Among persons with known race-ethnicity, rates were highest among Whites, followed by Blacks, then Hispanics, and lowest among Asians. Rates were highest in the Northeast, followed by the South, then the Midwest, and lowest in the West.

**Table 78:**
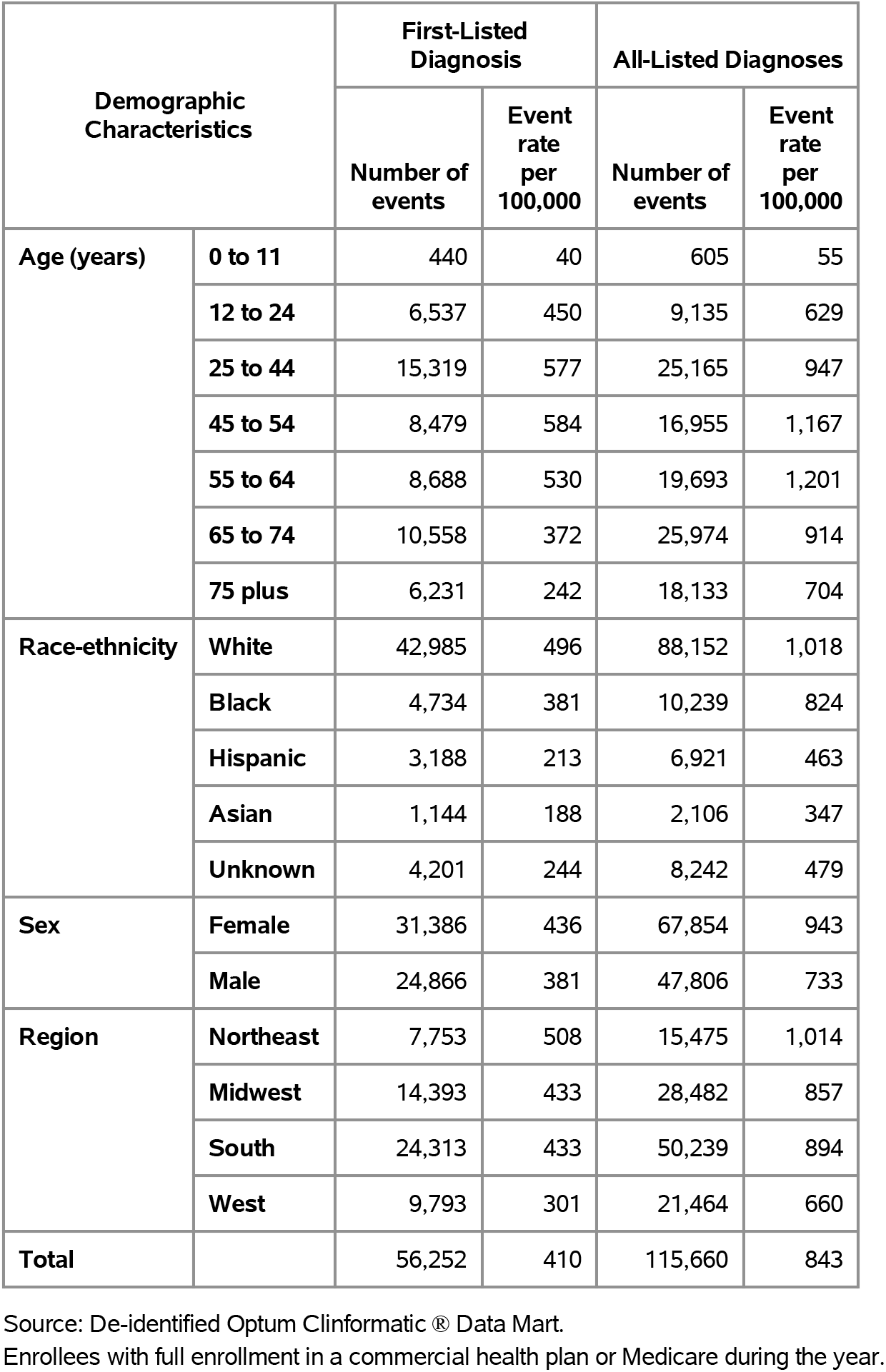
Crohns Disease: Ambulatory care visits with first-listed and all-listed diagnoses by age, race-ethnicity, sex and region among privately insured enrollees, 2020.

| Demographic Characteristics |  | First-Listed Diagnosis |  | All-Listed Diagnoses |  |
| --- | --- | --- | --- | --- | --- |
|  |  | Number of events | Event rate per 100,000 | Number of events | Event rate per 100,000 |
| Age (years) | 0 to 11 | 440 | 40 | 605 | 55 |
|  | 12 to 24 | 6,537 | 450 | 9,135 | 629 |
|  | 25 to 44 | 15,319 | 577 | 25,165 | 947 |
|  | 45 to 54 | 8,479 | 584 | 16,955 | 1,167 |
|  | 55 to 64 | 8,688 | 530 | 19,693 | 1,201 |
|  | 65 to 74 | 10,558 | 372 | 25,974 | 914 |
|  | 75 plus | 6,231 | 242 | 18,133 | 704 |
| Race-ethnicity | White | 42,985 | 496 | 88,152 | 1,018 |
|  | Black | 4,734 | 381 | 10,239 | 824 |
|  | Hispanic | 3,188 | 213 | 6,921 | 463 |
|  | Asian | 1,144 | 188 | 2,106 | 347 |
|  | Unknown | 4,201 | 244 | 8,242 | 479 |
| Sex | Female | 31,386 | 436 | 67,854 | 943 |
|  | Male | 24,866 | 381 | 47,806 | 733 |
| Region | Northeast | 7,753 | 508 | 15,475 | 1,014 |
|  | Midwest | 14,393 | 433 | 28,482 | 857 |
|  | South | 24,313 | 433 | 50,239 | 894 |
|  | West | 9,793 | 301 | 21,464 | 660 |
| Total |  | 56,252 | 410 | 115,660 | 843 |

Source: De-identified Optum Clinformatic® Data Mart.
Enrollees with full enrollment in a commercial health plan or Medicare during the year.

Among commercial insurance enrollees, emergency department visit rates with Crohn’s disease (all-listed diagnoses) peaked among persons 45 to 54 years and were higher among women compared with men (Table 79). Among persons with known race-ethnicity, rates were highest among Whites and Blacks, followed by Hispanics, and lowest among Asians. Rates were highest in the Northeast, followed by the South and Midwest, and lowest in the West.

**Table 79:** Crohns Disease: Emergency department visits with first-listed and all-listed diagnoses by age, race-ethnicity, sex and region among privately insured enrollees, 2020.

| Demographic Characteristics |  | First-Listed Diagnosis |  | All-Listed Diagnoses |  |
| --- | --- | --- | --- | --- | --- |
|  |  | Number of events | Event rate per 100,000 | Number of events | Event rate per 100,000 |
| Age (years) | 0 to 11 | 2 | 0 | 2 | 0 |
|  | 12 to 24 | 40 | 3 | 67 | 5 |
|  | 25 to 44 | 308 | 12 | 663 | 25 |
|  | 45 to 54 | 231 | 16 | 421 | 29 |
|  | 55 to 64 | 147 | 9 | 328 | 20 |
|  | 65 to 74 | 170 | 6 | 473 | 17 |
|  | 75 plus | 106 | 4 | 382 | 15 |
| Race-ethnicity | White | 738 | 9 | 1,761 | 20 |
|  | Black | 115 | 9 | 242 | 19 |
|  | Hispanic | 59 | 4 | 129 | 9 |
|  | Asian | 13 | 2 | 24 | 4 |
|  | Unknown | 79 | 5 | 180 | 10 |
| Sex | Female | 568 | 8 | 1,446 | 20 |
|  | Male | 436 | 7 | 890 | 14 |
| Region | Northeast | 126 | 8 | 368 | 24 |
|  | Midwest | 231 | 7 | 551 | 17 |
|  | South | 496 | 9 | 1,036 | 18 |
|  | West | 151 | 5 | 381 | 12 |
| Total |  | 1,004 | 7 | 2,336 | 17 |

Source: De-identified Optum Clinformatics® Data Mart.
Enrollees with full enrollment in a commercial health plan or Medicare during the year.

Among commercial insurance enrollees, hospital discharge rates with Crohn’s disease (all-listed diagnoses) increased with age until 65 years and were higher among women compared with men (Table 80). Among persons with known race-ethnicity, rates were highest among Blacks and Whites, followed by Hispanics, and lowest among Asians. Rates were highest in the Northeast, followed by the South, then the Midwest, and lowest in the West.

**Table 80:** Crohns Disease: Hospital discharges with first-listed and all-listed diagnoses by age, race-ethnicity, sex and region among privately insured enrollees, 2020.

| Demographic Characteristics |  | First-Listed Diagnosis |  | All-Listed Diagnoses |  |
| --- | --- | --- | --- | --- | --- |
|  |  | Number of events | Event rate per 100,000 | Number of events | Event rate per 100,000 |
| Age (years) | 0 to 11 | 18 | 2 | 23 | 2 |
|  | 12 to 24 | 208 | 14 | 378 | 26 |
|  | 25 to 44 | 512 | 19 | 1,418 | 53 |
|  | 45 to 54 | 268 | 18 | 1,031 | 71 |
|  | 55 to 64 | 286 | 17 | 1,536 | 94 |
|  | 65 to 74 | 319 | 11 | 2,414 | 85 |
|  | 75 plus | 231 | 9 | 2,412 | 94 |
| Race-ethnicity | White | 1,302 | 15 | 6,853 | 79 |
|  | Black | 198 | 16 | 988 | 80 |
|  | Hispanic | 113 | 8 | 531 | 36 |
|  | Asian | 40 | 7 | 118 | 19 |
|  | Unknown | 189 | 11 | 722 | 42 |
| Sex | Female | 1,043 | 14 | 5,429 | 75 |
|  | Male | 799 | 12 | 3,783 | 58 |
| Region | Northeast | 256 | 17 | 1,208 | 79 |
|  | Midwest | 458 | 14 | 2,272 | 68 |
|  | South | 815 | 15 | 4,042 | 72 |
|  | West | 313 | 10 | 1,690 | 52 |
| Total |  | 1,842 | 13 | 9,212 | 67 |

Source: De-identified Optum Clinformatic® Data Mart.
Enrollees with full enrollment in a commercial health plan or Medicare during the year.

Among Medicare beneficiaries, the claims-based prevalence of Crohn’s disease (based on all-listed diagnoses) was 0.4% (Table 81). Prevalence differed little by age, did not differ by sex, and was higher among Whites. It was highest in the Northeast and lowest in the West.

**Table 81:** Crohns Disease: Claims-based prevalence with first-listed and all-listed diagnoses by age, race, sex and region among fee-for-service, age-eligible Medicare beneficiaries, 2019.

| Demographic Characteristics |  | Number of enrollees | First-Listed Diagnosis |  | All-Listed Diagnoses |  |
| --- | --- | --- | --- | --- | --- | --- |
|  |  |  | Number of enrollees with disease | Percent of enrollees with disease | Number of enrollees with disease | Percent of enrollees with disease |
| Age (years) | 65 to 69 | 8,066,320 | 21,000 | 0.3% | 33,780 | 0.4% |
|  | 70 to 74 | 6,936,940 | 17,420 | 0.3% | 28,860 | 0.4% |
|  | 75 to 79 | 4,894,060 | 11,540 | 0.2% | 20,140 | 0.4% |
|  | 80 to 84 | 3,335,340 | 6,400 | 0.2% | 12,200 | 0.4% |
|  | 85+ | 3,578,120 | 3,780 | 0.1% | 8,360 | 0.2% |
| Race | White | 22,023,800 | 53,380 | 0.2% | 92,340 | 0.4% |
|  | Black | 1,861,660 | 2,520 | 0.1% | 4,360 | 0.2% |
|  | Other | 2,925,320 | 4,240 | 0.1% | 6,640 | 0.2% |
| Sex | Female | 14,975,560 | 34,860 | 0.2% | 60,900 | 0.4% |
|  | Male | 11,835,220 | 25,280 | 0.2% | 42,440 | 0.4% |
| Region | Northeast | 4,748,700 | 14,000 | 0.3% | 22,860 | 0.5% |
|  | Midwest | 6,069,800 | 14,120 | 0.2% | 24,700 | 0.4% |
|  | South | 10,595,900 | 22,580 | 0.2% | 39,340 | 0.4% |
|  | West | 5,396,380 | 9,440 | 0.2% | 16,440 | 0.3% |
| Total |  | 26,810,780 | 60,140 | 0.2% | 103,340 | 0.4% |

Source: Centers for Medicare & Medicaid Services, 5% Denominator and Claims Files. Beneficiaries aged 65+ years with continuous and full Parts A and B enrollment and no Health Maintenance Organization enrollment during the year. Unweighted counts were multiplied by 20 to produce counts for this table.

Among Medicare beneficiaries, ambulatory care visit rates with Crohn’s disease (all-listed diagnoses) decreased with age and were higher among women compared with men and Whites compared with Blacks (Table 82). Rates were highest in the Northeast, followed by the South and Midwest, and lowest in the West.

**Table 82:** Crohns Disease: Ambulatory care visits with first-listed and all-listed diagnoses by age, race, sex and region among fee-for-service, age-eligible Medicare beneficiaries, 2019.

| Demographic Characteristics |  | First-Listed Diagnosis |  | All-Listed Diagnoses |  |
| --- | --- | --- | --- | --- | --- |
|  |  | Number of events | Event rate per 100,000 | Number of events | Event rate per 100,000 |
| Age (years) | 65 to 69 | 63,380 | 786 | 127,640 | 1,582 |
|  | 70 to 74 | 53,740 | 775 | 109,200 | 1,574 |
|  | 75 to 79 | 32,420 | 662 | 72,000 | 1,471 |
|  | 80 to 84 | 18,900 | 567 | 41,500 | 1,244 |
|  | 85+ | 9,180 | 257 | 26,100 | 729 |
| Race | White | 157,740 | 716 | 335,440 | 1,523 |
|  | Black | 7,100 | 381 | 15,620 | 839 |
|  | Other | 12,780 | 437 | 25,380 | 868 |
| Sex | Female | 100,300 | 670 | 223,340 | 1,491 |
|  | Male | 77,320 | 653 | 153,100 | 1,294 |
| Region | Northeast | 39,740 | 837 | 85,980 | 1,811 |
|  | Midwest | 39,720 | 654 | 84,440 | 1,391 |
|  | South | 70,520 | 666 | 147,480 | 1,392 |
|  | West | 27,640 | 512 | 58,540 | 1,085 |
| Total |  | 177,620 | 662 | 376,440 | 1,404 |

Source: Centers for Medicare & Medicaid Services, 5% Denominator and Claims Files. Beneficiaries aged 65+ years with continuous and full Parts A and B enrollment and no Health Maintenance Organization enrollment during the year. Unweighted counts were multiplied by 20 to produce counts for this table.

Among Medicare beneficiaries, emergency department visit rates with Crohn’s disease (all-listed diagnoses) increased with age until 85 years and were higher among women compared with men and Whites compared with Blacks (Table 83). Rates were highest in the Northeast, followed by the Midwest, then the South, and lowest in the West.

**Table 83:** Crohns Disease: Emergency department visits with first-listed and all-listed diagnoses by age, race, sex and region among fee-for-service, age-eligible Medicare beneficiaries, 2019.

| Demographic Characteristics |  | First-Listed Diagnosis |  | All-Listed Diagnoses |  |
| --- | --- | --- | --- | --- | --- |
|  |  | Number of events | Event rate per 100,000 | Number of events | Event rate per 100,000 |
| Age (years) | 65 to 69 | 2,000 | 25 | 13,420 | 166 |
|  | 70 to 74 | 1,580 | 23 | 12,240 | 176 |
|  | 75 to 79 | 1,300 | 27 | 10,540 | 215 |
|  | 80 to 84 | 960 | 29 | 7,500 | 225 |
|  | 85+ | 340 | 10 | 5,740 | 160 |
| Race | White | 5,280 | 24 | 43,680 | 198 |
|  | Black | 420 | 23 | 2,800 | 150 |
|  | Other | 480 | 16 | 2,960 | 101 |
| Sex | Female | 4,080 | 27 | 30,940 | 207 |
|  | Male | 2,100 | 18 | 18,500 | 156 |
| Region | Northeast | 1,560 | 33 | 11,340 | 239 |
|  | Midwest | 1,260 | 21 | 12,000 | 198 |
|  | South | 2,520 | 24 | 18,820 | 178 |
|  | West | 840 | 16 | 7,280 | 135 |
| Total |  | 6,180 | 23 | 49,440 | 184 |

Source: Centers for Medicare & Medicaid Services, 5% Denominator and Claims Files. Beneficiaries aged 65+ years with continuous and full Parts A and B enrollment and no Health Maintenance Organization enrollment during the year. Unweighted counts were multiplied by 20 to produce counts for this table.

Among Medicare beneficiaries, hospital discharge rates with Crohn’s disease (all-listed diagnoses) peaked among persons 75 to 79 years and were higher among women compared with men and Whites compared with Blacks (Table 84). Rates were highest in the Northeast, followed by the Midwest, then the South, and lowest in the West.

**Table 84:** Crohns Disease: Hospital discharges with first-listed and all-listed diagnoses by age, race, sex and region among fee-for-service, age-eligible Medicare beneficiaries, 2019.

| Demographic Characteristics |  | First-Listed Diagnosis |  | All-Listed Diagnoses |  |
| --- | --- | --- | --- | --- | --- |
|  |  | Number of events | Event rate per 100,000 | Number of events | Event rate per 100,000 |
| Age (years) | 65 to 69 | 1,500 | 19 | 9,540 | 118 |
|  | 70 to 74 | 1,280 | 18 | 9,600 | 138 |
|  | 75 to 79 | 860 | 18 | 8,720 | 178 |
|  | 80 to 84 | 600 | 18 | 5,360 | 161 |
|  | 85+ | 380 | 11 | 4,500 | 126 |
| Race | White | 4,000 | 18 | 33,520 | 152 |
|  | Black | 280 | 15 | 2,060 | 111 |
|  | Other | 340 | 12 | 2,140 | 73 |
| Sex | Female | 2,700 | 18 | 23,340 | 156 |
|  | Male | 1,920 | 16 | 14,380 | 122 |
| Region | Northeast | 1,200 | 25 | 8,380 | 176 |
|  | Midwest | 1,000 | 16 | 9,760 | 161 |
|  | South | 1,860 | 18 | 14,200 | 134 |
|  | West | 560 | 10 | 5,380 | 100 |
| Total |  | 4,620 | 17 | 37,720 | 141 |

Source: Centers for Medicare & Medicaid Services, 5% Denominator and Claims Files. Beneficiaries aged 65+ years with continuous and full Parts A and B enrollment and no Health Maintenance Organization enrollment during the year. Unweighted counts were multiplied by 20 to produce counts for this table.

Ulcerative colitis contributed to 1.3 million ambulatory visits (2015) (Table 85). Ambulatory care visit rates (all-listed diagnoses) were highest among persons 65-74 years. Age-adjusted rates were higher among women compared with men, Blacks compared with Whites, and Hispanics compared with non-Hispanics.

**Table 85:** Ulcerative Colitis: Ambulatory care visits with first-listed and all-listed diagnoses by age, race, ethnicity, and sex in the United States, 2015.

| Demographic Characteristics |  | First-Listed Diagnosis |  | All-Listed Diagnoses |  |
| --- | --- | --- | --- | --- | --- |
|  |  | Number in thousands | Rate per 100,000 | Number in thousands | Rate per 100,000 |
| Age (years) | 0 to 11 | . | . | . | . |
|  | 12 to 24 | 38 | 67 | 110 | 196 |
|  | 25 to 44 | 64 | 76 | 128 | 152 |
|  | 45 to 54 | 343 | 795 | 372 | 864 |
|  | 55 to 64 | 240 | 587 | 358 | 878 |
|  | 65 to 74 | 65 | 235 | 269 | 976 |
|  | 75 plus | 39 | 195 | 68 | 338 |
| Race | White | 565 | 203 | 1,011 | 351 |
|  | Black | 188 | 385 | 248 | 510 |
|  | Other | 36 | 218 | 47 | 288 |
| Ethnicity | Hispanic | 259 | 598 | 339 | 807 |
|  | Not Hispanic | 530 | 167 | 968 | 304 |
| Sex | Female | 418 | 235 | 796 | 431 |
|  | Male | 370 | 211 | 510 | 285 |
| Total |  | 789 | 221 | 1,306 | 357 |

Source: National Ambulatory Medical Care Survey (3-year average, 2014-2016). Rates for age groups are unadjusted and all other rates are age-adjusted.

Ulcerative colitis contributed to 164,000 emergency department visits in 2018 (Table 86). Emergency department visit rates (all-listed diagnoses) increased with age. Age-adjusted rates were higher among women compared with men.

**Table 86:**
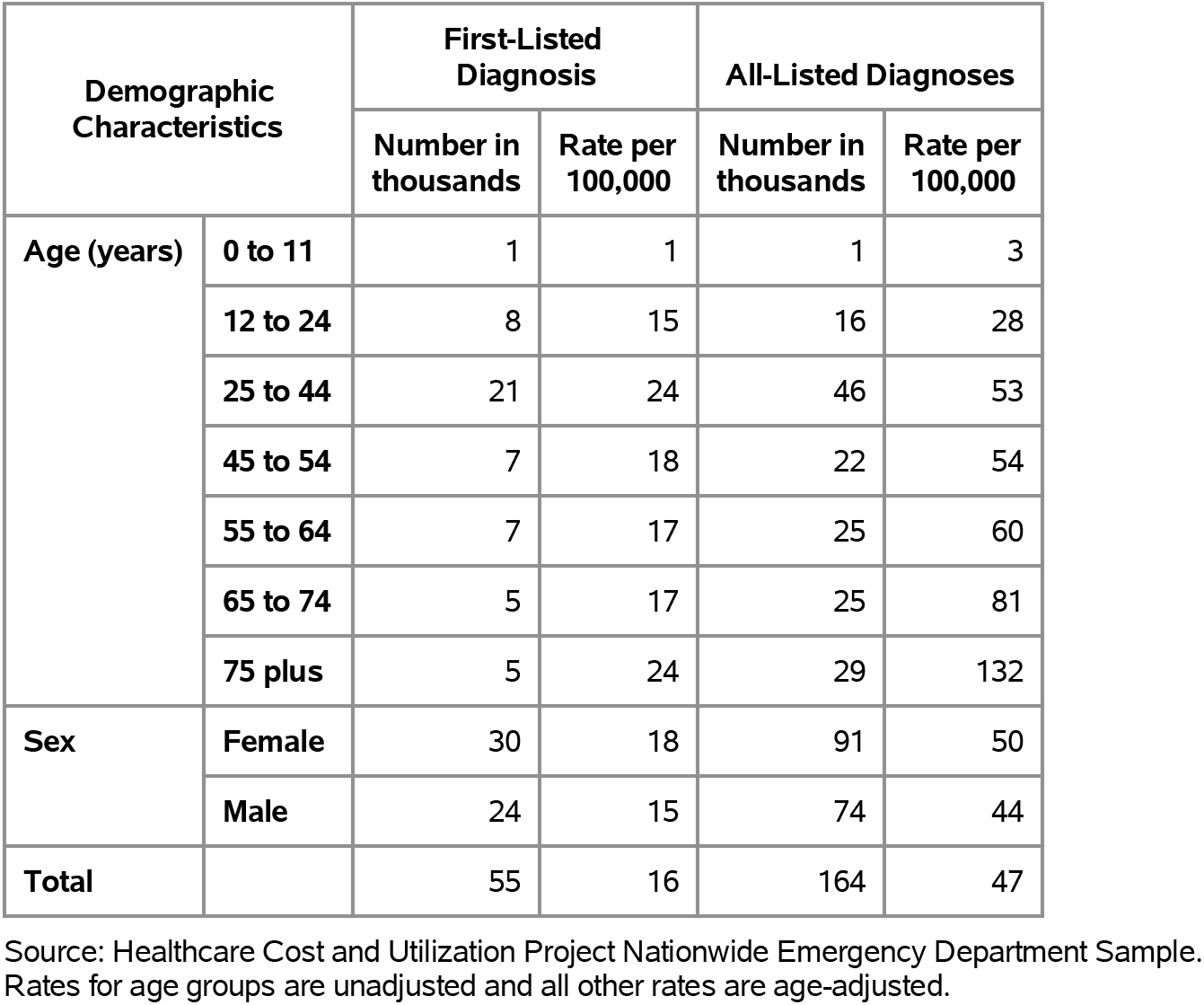
Ulcerative Colitis: Emergency department visits with first-listed and all-listed diagnoses by age and sex in the United States, 2018.

Ulcerative colitis contributed to 128,000 hospital discharges in 2018 (Table 87). Hospital discharge rates (all-listed diagnoses) increased with age. Age-adjusted rates were higher among women compared with men, Whites compared with Blacks, and non-Hispanics compared with Hispanics. Between 2004 and 2018, age-adjusted hospital discharge rates (per 100,000) with an all-listed diagnosis increased by 29% from 28 to 36.(4)

**Table 87:**
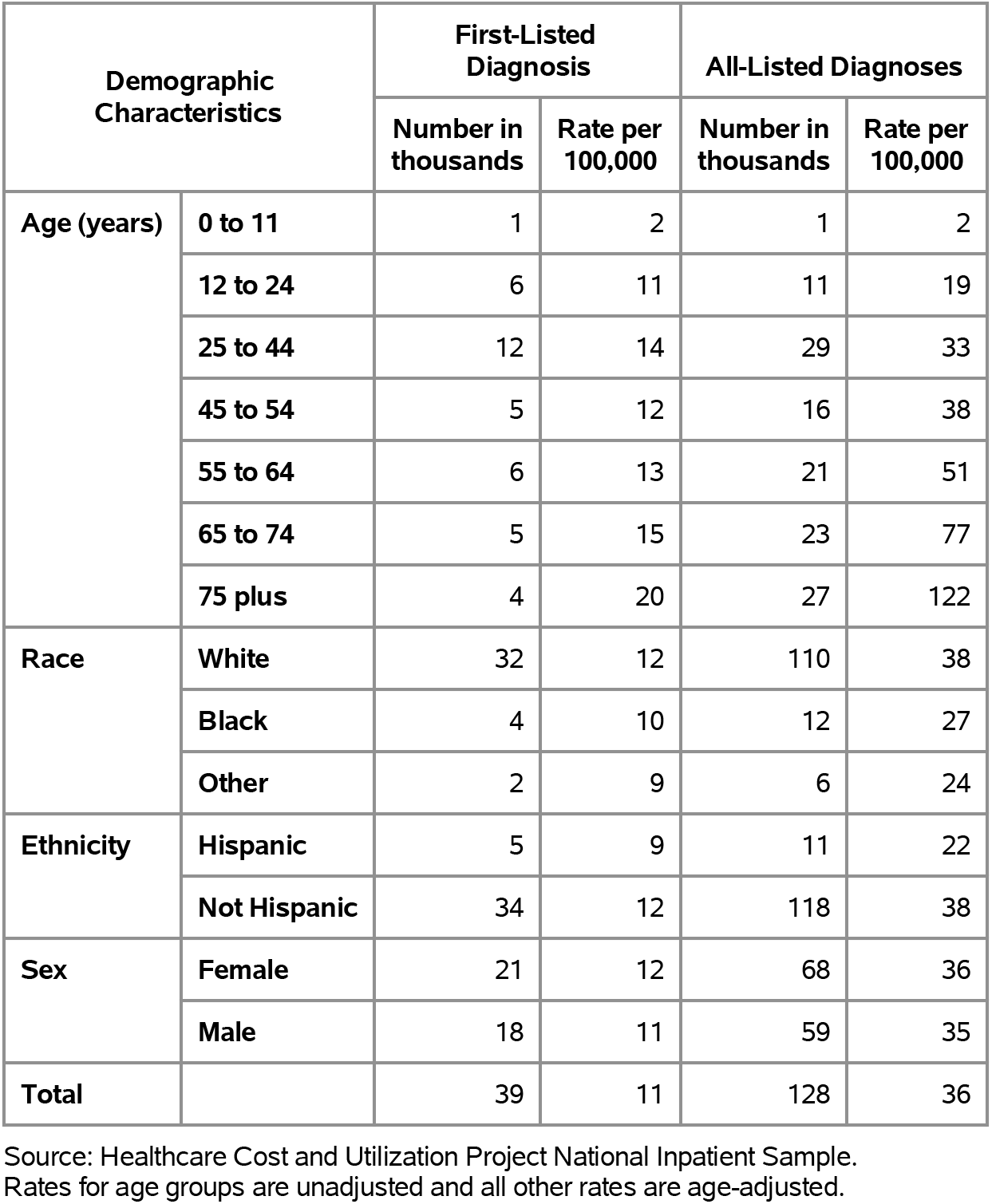
Ulcerative Colitis: Hospital discharges with first-listed and all-listed diagnoses by age, race, ethnicity, and sex in the United States, 2018.

Ulcerative colitis contributed to 1,000 deaths in 2019 (Table 88). Mortality was uncommon among the youngest age groups after which rates (underlying or other cause) increased with age. Age-adjusted mortality rates were higher among men, Whites, and non-Hispanics. Between 2004 and 2019, age-adjusted mortality rates (per 100,000) with ulcerative colitis as underlying or other cause increased by a third from 0.3 to 0.4.(4)

**Table 88:** Ulcerative Colitis: Deaths with underlying or underlying/other cause and lifetime years of life lost by age, race, ethnicity, and sex in the United States, 2019.

| Demographic Characteristics |  | Underlying Cause |  |  | Underlying/Other Cause |  |  |
| --- | --- | --- | --- | --- | --- | --- | --- |
|  |  | Number of Deaths | Rate per 100,000 | Years of potential life lost in thousands | Number of Deaths | Rate per 100,000 | Years of potential life lost in thousands |
| Age (years) | 0 to 11 | . | . | . | . | . | . |
|  | 12 to 24 | 3 | 0.0 | 0.2 | 13 | 0.0 | 0.8 |
|  | 25 to 44 | 15 | 0.0 | 0.6 | 55 | 0.1 | 2.4 |
|  | 45 to 54 | 24 | 0.1 | 0.8 | 83 | 0.2 | 2.6 |
|  | 55 to 64 | 54 | 0.1 | 1.3 | 186 | 0.4 | 4.3 |
|  | 65 to 74 | 104 | 0.3 | 1.6 | 329 | 1.0 | 5.2 |
|  | 75 plus | 224 | 1.0 | 1.6 | 800 | 3.5 | 5.8 |
| Race | White | 377 | 0.1 | 5.2 | 1,346 | 0.4 | 18.7 |
|  | Black | 36 | 0.1 | 0.7 | 92 | 0.2 | 1.8 |
|  | Other | 11 | 0.0 | 0.2 | 28 | 0.1 | 0.6 |
| Ethnicity | Hispanic | 12 | 0.0 | 0.3 | 47 | 0.1 | 1.2 |
|  | Not Hispanic | 412 | 0.1 | 5.8 | 1,419 | 0.4 | 19.8 |
| Sex | Female | 227 | 0.1 | 3.3 | 708 | 0.3 | 10.3 |
|  | Male | 197 | 0.1 | 2.8 | 758 | 0.4 | 10.7 |
| Total |  | 424 | 0.1 | 6.1 | 1,466 | 0.4 | 21.0 |

Source: Vital Statistics of the United States.
Rates for age groups are unadjusted and all other rates are age-adjusted.
Years of potential life lost is based on U.S. life table 2006-2018 estimates of life expectancy at age of death.

Among privately insured enrollees, the claims-based prevalence of ulcerative colitis (based on all-listed diagnoses) was 0.4% (Table 89). Prevalence increased with age until the oldest age group and was higher among women. It was highest among Whites, similar among Blacks and Hispanics, and lowest among Asians. It differed little by region.

**Table 89:**
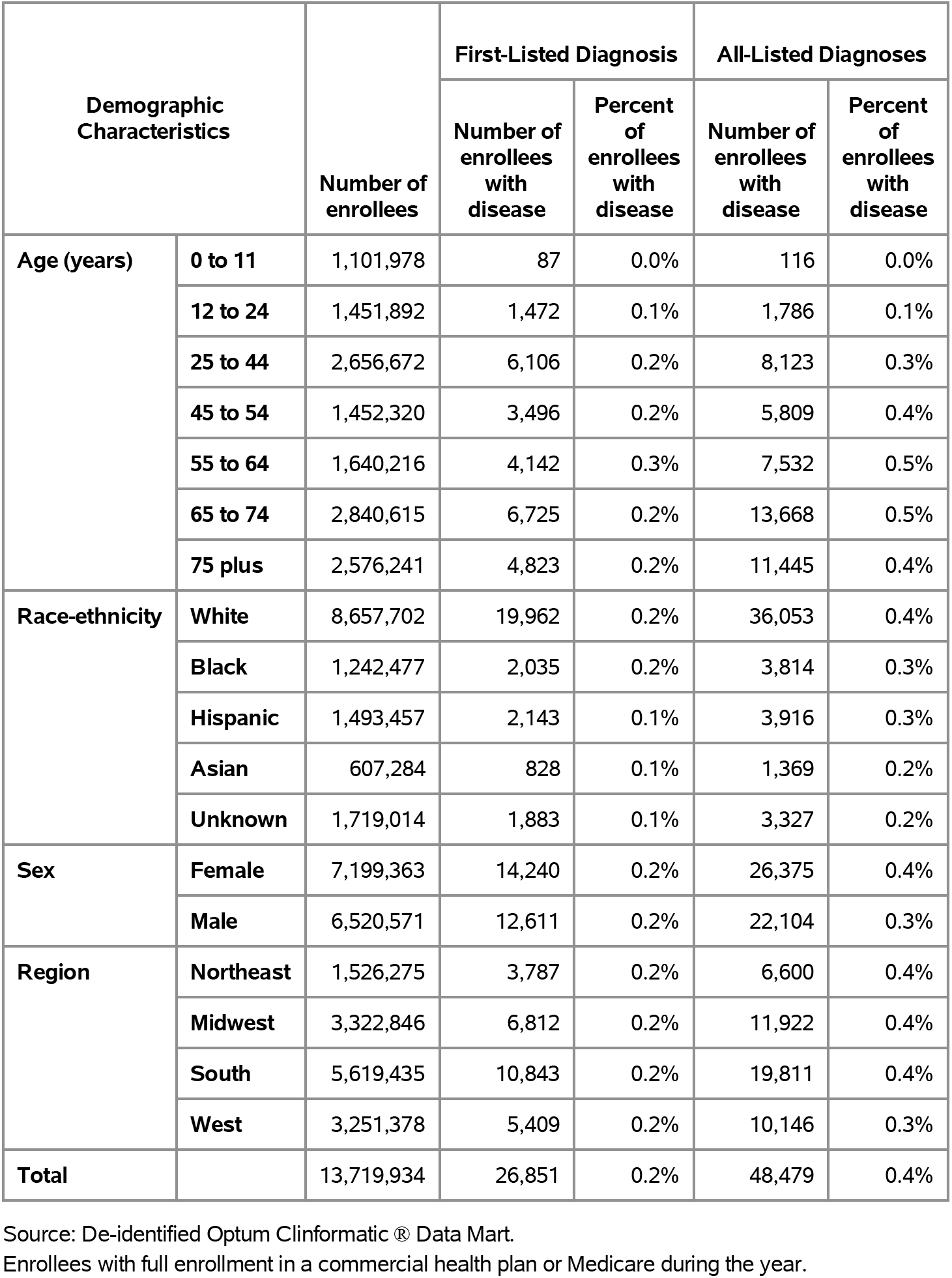
Ulcerative Colitis: Claims-based prevalence with first-listed and all-listed diagnoses by age, race-ethnicity, sex and region among privately insured enrollees, 2020.

| Demographic Characteristics |  | Number of enrollees | First-Listed Diagnosis |  | All-Listed Diagnoses |  |
| --- | --- | --- | --- | --- | --- | --- |
|  |  |  | Number of enrollees with disease | Percent of enrollees with disease | Number of enrollees with disease | Percent of enrollees with disease |
| Age (years) | 0 to 11 | 1,101,978 | 87 | 0.0% | 116 | 0.0% |
|  | 12 to 24 | 1,451,892 | 1,472 | 0.1% | 1,786 | 0.1% |
|  | 25 to 44 | 2,656,672 | 6,106 | 0.2% | 8,123 | 0.3% |
|  | 45 to 54 | 1,452,320 | 3,496 | 0.2% | 5,809 | 0.4% |
|  | 55 to 64 | 1,640,216 | 4,142 | 0.3% | 7,532 | 0.5% |
|  | 65 to 74 | 2,840,615 | 6,725 | 0.2% | 13,668 | 0.5% |
|  | 75 plus | 2,576,241 | 4,823 | 0.2% | 11,445 | 0.4% |
| Race-ethnicity | White | 8,657,702 | 19,962 | 0.2% | 36,053 | 0.4% |
|  | Black | 1,242,477 | 2,035 | 0.2% | 3,814 | 0.3% |
|  | Hispanic | 1,493,457 | 2,143 | 0.1% | 3,916 | 0.3% |
|  | Asian | 607,284 | 828 | 0.1% | 1,369 | 0.2% |
|  | Unknown | 1,719,014 | 1,883 | 0.1% | 3,327 | 0.2% |
| Sex | Female | 7,199,363 | 14,240 | 0.2% | 26,375 | 0.4% |
|  | Male | 6,520,571 | 12,611 | 0.2% | 22,104 | 0.3% |
| Region | Northeast | 1,526,275 | 3,787 | 0.2% | 6,600 | 0.4% |
|  | Midwest | 3,322,846 | 6,812 | 0.2% | 11,922 | 0.4% |
|  | South | 5,619,435 | 10,843 | 0.2% | 19,811 | 0.4% |
|  | West | 3,251,378 | 5,409 | 0.2% | 10,146 | 0.3% |
| Total |  | 13,719,934 | 26,851 | 0.2% | 48,479 | 0.4% |

Source: De-identified Optum Clinformatic® Data Mart.
Enrollees with full enrollment in a commercial health plan or Medicare during the year.

Among commercial insurance enrollees, ambulatory care visit rates with ulcerative colitis (all-listed diagnoses) peaked among persons 55 to 74 years and were higher among women compared with men (Table 90). Among persons with known race-ethnicity, rates were highest among Whites, followed by Blacks, then Hispanics, and lowest among Asians. Rates were highest in the Northeast, followed by the South, then the Midwest, and lowest in the West.

**Table 90:** Ulcerative Colitis: Ambulatory care visits with first-listed and all-listed diagnoses by age, race-ethnicity, sex and region among privately insured enrollees, 2020.

| Demographic Characteristics |  | First-Listed Diagnosis |  | All-Listed Diagnoses |  |
| --- | --- | --- | --- | --- | --- |
|  |  | Number of events | Event rate per 100,000 | Number of events | Event rate per 100,000 |
| Age (years) | 0 to 11 | 215 | 20 | 318 | 29 |
|  | 12 to 24 | 3,718 | 256 | 5,202 | 358 |
|  | 25 to 44 | 12,849 | 484 | 19,957 | 751 |
|  | 45 to 54 | 6,578 | 453 | 12,739 | 877 |
|  | 55 to 64 | 7,490 | 457 | 16,090 | 981 |
|  | 65 to 74 | 11,765 | 414 | 27,889 | 982 |
|  | 75 plus | 8,109 | 315 | 23,147 | 898 |
| Race-ethnicity | White | 37,168 | 429 | 77,927 | 900 |
|  | Black | 3,737 | 301 | 8,049 | 648 |
|  | Hispanic | 4,373 | 293 | 8,916 | 597 |
|  | Asian | 1,728 | 285 | 3,145 | 518 |
|  | Unknown | 3,718 | 216 | 7,305 | 425 |
| Sex | Female | 26,327 | 366 | 56,748 | 788 |
|  | Male | 24,397 | 374 | 48,594 | 745 |
| Region | Northeast | 7,219 | 473 | 14,747 | 966 |
|  | Midwest | 11,940 | 359 | 24,443 | 736 |
|  | South | 21,257 | 378 | 43,552 | 775 |
|  | West | 10,308 | 317 | 22,600 | 695 |
| Total |  | 50,724 | 370 | 105,342 | 768 |

Source: De-identified Optum Clinformatic® Data Mart.
Enrollees with full enrollment in a commercial health plan or Medicare during the year.

Among commercial insurance enrollees, emergency department visit rates with ulcerative colitis (all-listed diagnoses) were highest among persons 75 years and older and were higher among women compared with men (Table 91). Among persons with known race-ethnicity, rates were highest among Whites, followed by Blacks, then Hispanics, and lowest among Asians. Rates were highest in the Northeast, followed by the South, and lowest in the Midwest and West.

**Table 91:** Ulcerative Colitis: Emergency department visits with first-listed and all-listed diagnoses by age, race-ethnicity, sex and region among privately insured enrollees, 2020.

| Demographic Characteristics |  | First-Listed Diagnosis |  | All-Listed Diagnoses |  |
| --- | --- | --- | --- | --- | --- |
|  |  | Number of events | Event rate per 100,000 | Number of events | Event rate per 100,000 |
| Age (years) | 0 to 11 | 2 | 0 | 2 | 0 |
|  | 12 to 24 | 34 | 2 | 63 | 4 |
|  | 25 to 44 | 232 | 9 | 444 | 17 |
|  | 45 to 54 | 89 | 6 | 157 | 11 |
|  | 55 to 64 | 138 | 8 | 261 | 16 |
|  | 65 to 74 | 249 | 9 | 511 | 18 |
|  | 75 plus | 278 | 11 | 628 | 24 |
| Race-ethnicity | White | 727 | 8 | 1,470 | 17 |
|  | Black | 98 | 8 | 191 | 15 |
|  | Hispanic | 86 | 6 | 181 | 12 |
|  | Asian | 24 | 4 | 57 | 9 |
|  | Unknown | 87 | 5 | 167 | 10 |
| Sex | Female | 660 | 9 | 1,343 | 19 |
|  | Male | 362 | 6 | 723 | 11 |
| Region | Northeast | 177 | 12 | 310 | 20 |
|  | Midwest | 228 | 7 | 436 | 13 |
|  | South | 436 | 8 | 918 | 16 |
|  | West | 181 | 6 | 402 | 12 |
| Total |  | 1,022 | 7 | 2,066 | 15 |

Source: De-identified Optum Clinformatic® Data Mart.
Enrollees with full enrollment in a commercial health plan or Medicare during the year.

Among commercial insurance enrollees, hospital discharge rates with ulcerative colitis (all-listed diagnoses) increased with age and were higher among women compared with men (Table 92). Among persons with known race-ethnicity, rates were highest among Whites, followed by Blacks, then Hispanics, and lowest among Asians. Rates were highest in the Northeast, followed by the South, then the Midwest, and lowest in the West.

**Table 92:** Ulcerative Colitis: Hospital discharges with first-listed and all-listed diagnoses by age, race-ethnicity, sex and region among privately insured enrollees, 2020.

| Demographic Characteristics |  | First-Listed Diagnosis |  | All-Listed Diagnoses |  |
| --- | --- | --- | --- | --- | --- |
|  |  | Number of events | Event rate per 100,000 | Number of events | Event rate per 100,000 |
| Age (years) | 0 to 11 | 16 | 1 | 24 | 2 |
|  | 12 to 24 | 125 | 9 | 246 | 17 |
|  | 25 to 44 | 241 | 9 | 873 | 33 |
|  | 45 to 54 | 126 | 9 | 529 | 36 |
|  | 55 to 64 | 181 | 11 | 1,064 | 65 |
|  | 65 to 74 | 332 | 12 | 2,174 | 77 |
|  | 75 plus | 327 | 13 | 2,738 | 106 |
| Race-ethnicity | White | 1,000 | 12 | 5,632 | 65 |
|  | Black | 126 | 10 | 724 | 58 |
|  | Hispanic | 100 | 7 | 588 | 39 |
|  | Asian | 25 | 4 | 140 | 23 |
|  | Unknown | 97 | 6 | 564 | 33 |
| Sex | Female | 789 | 11 | 4,382 | 61 |
|  | Male | 559 | 9 | 3,266 | 50 |
| Region | Northeast | 198 | 13 | 1,130 | 74 |
|  | Midwest | 323 | 10 | 1,834 | 55 |
|  | South | 598 | 11 | 3,235 | 58 |
|  | West | 229 | 7 | 1,449 | 45 |
| Total |  | 1,348 | 10 | 7,648 | 56 |

Source: De-identified Optum Clinformatic® Data Mart.
Enrollees with full enrollment in a commercial health plan or Medicare during the year.

Among Medicare beneficiaries, the claims-based prevalence of ulcerative colitis (based on all-listed diagnoses) was 0.6% (Table 93). Prevalence was highest among persons 65-79 years, did not differ by sex, and was higher among Whites. It was highest in the Northeast and lowest in the South and West.

**Table 93:** Ulcerative Colitis: Claims-based prevalence with first-listed and all-listed diagnoses by age, race, sex and region among fee-for-service, age-eligible Medicare beneficiaries, 2019.

| Demographic Characteristics |  | Number of enrollees | First-Listed Diagnosis |  | All-Listed Diagnoses |  |
| --- | --- | --- | --- | --- | --- | --- |
|  |  |  | Number of enrollees with disease | Percent of enrollees with disease | Number of enrollees with disease | Percent of enrollees with disease |
| Age (years) | 65 to 69 | 8,066,320 | 24,820 | 0.3% | 45,580 | 0.6% |
|  | 70 to 74 | 6,936,940 | 22,580 | 0.3% | 43,040 | 0.6% |
|  | 75 to 79 | 4,894,060 | 15,320 | 0.3% | 30,560 | 0.6% |
|  | 80 to 84 | 3,335,340 | 8,620 | 0.3% | 17,900 | 0.5% |
|  | 85+ | 3,578,120 | 6,700 | 0.2% | 15,300 | 0.4% |
| Race | White | 22,023,800 | 68,360 | 0.3% | 133,360 | 0.6% |
|  | Black | 1,861,660 | 3,260 | 0.2% | 6,920 | 0.4% |
|  | Other | 2,925,320 | 6,420 | 0.2% | 12,100 | 0.4% |
| Sex | Female | 14,975,560 | 43,160 | 0.3% | 85,840 | 0.6% |
|  | Male | 11,835,220 | 34,880 | 0.3% | 66,540 | 0.6% |
| Region | Northeast | 4,748,700 | 17,900 | 0.4% | 34,100 | 0.7% |
|  | Midwest | 6,069,800 | 16,480 | 0.3% | 33,840 | 0.6% |
|  | South | 10,595,900 | 29,640 | 0.3% | 58,060 | 0.5% |
|  | West | 5,396,380 | 14,020 | 0.3% | 26,380 | 0.5% |
| Total |  | 26,810,780 | 78,040 | 0.3% | 152,380 | 0.6% |

Source: Centers for Medicare & Medicaid Services, 5% Denominator and Claims Files. Beneficiaries aged 65+ years with continuous and full Parts A and B enrollment and no Health Maintenance Organization enrollment during the year. Unweighted counts were multiplied by 20 to produce counts for this table.

Among Medicare beneficiaries, ambulatory care visit rates with ulcerative colitis (all-listed diagnoses) peaked among persons 70 to 74 years and were higher among men compared with women and Whites compared with Blacks (Table 94). Rates were highest in the Northeast, followed by the South, then the Midwest, and lowest in the West.

**Table 94:** Ulcerative Colitis: Ambulatory care visits with first-listed and all-listed diagnoses by age, race, sex and region among fee-for-service, age-eligible Medicare beneficiaries, 2019.

| Demographic Characteristics |  | First-Listed Diagnosis |  | All-Listed Diagnoses |  |
| --- | --- | --- | --- | --- | --- |
|  |  | Number of events | Event rate per 100,000 | Number of events | Event rate per 100,000 |
| Age (years) | 65 to 69 | 52,380 | 649 | 110,540 | 1,370 |
|  | 70 to 74 | 47,280 | 682 | 104,680 | 1,509 |
|  | 75 to 79 | 31,500 | 644 | 71,660 | 1,464 |
|  | 80 to 84 | 16,420 | 492 | 42,040 | 1,260 |
|  | 85+ | 11,820 | 330 | 34,120 | 954 |
| Race | White | 140,240 | 637 | 320,340 | 1,455 |
|  | Black | 5,940 | 319 | 14,800 | 795 |
|  | Other | 13,220 | 452 | 27,900 | 954 |
| Sex | Female | 84,080 | 561 | 199,000 | 1,329 |
|  | Male | 75,320 | 636 | 164,040 | 1,386 |
| Region | Northeast | 36,380 | 766 | 82,580 | 1,739 |
|  | Midwest | 31,420 | 518 | 73,800 | 1,216 |
|  | South | 64,120 | 605 | 144,140 | 1,360 |
|  | West | 27,480 | 509 | 62,520 | 1,159 |
| Total |  | 159,400 | 595 | 363,040 | 1,354 |

Source: Centers for Medicare & Medicaid Services, 5% Denominator and Claims Files. Beneficiaries aged 65+ years with continuous and full Parts A and B enrollment and no Health Maintenance Organization enrollment during the year. Unweighted counts were multiplied by 20 to produce counts for this table.

Among Medicare beneficiaries, emergency department visit rates with ulcerative colitis (all-listed diagnoses) increased with age and were higher among women compared with men and Whites compared with Blacks (Table 95). Rates were highest in the Northeast, followed by the Midwest, then the South, and lowest in the West.

**Table 95:** Ulcerative Colitis: Emergency department visits with first-listed and all-listed diagnoses by age, race, sex and region among fee-for-service, age-eligible Medicare beneficiaries, 2019.

| Demographic Characteristics |  | First-Listed Diagnosis |  | All-Listed Diagnoses |  |
| --- | --- | --- | --- | --- | --- |
|  |  | Number of events | Event rate per 100,000 | Number of events | Event rate per 100,000 |
| Age (years) | 65 to 69 | 1,900 | 24 | 7,820 | 97 |
|  | 70 to 74 | 1,760 | 25 | 7,500 | 108 |
|  | 75 to 79 | 1,440 | 29 | 6,980 | 143 |
|  | 80 to 84 | 1,180 | 35 | 5,680 | 170 |
|  | 85+ | 1,160 | 32 | 6,420 | 179 |
| Race | White | 6,400 | 29 | 30,080 | 137 |
|  | Black | 360 | 19 | 1,820 | 98 |
|  | Other | 680 | 23 | 2,500 | 85 |
| Sex | Female | 4,820 | 32 | 20,520 | 137 |
|  | Male | 2,620 | 22 | 13,880 | 117 |
| Region | Northeast | 1,860 | 39 | 8,900 | 187 |
|  | Midwest | 1,620 | 27 | 7,800 | 129 |
|  | South | 2,740 | 26 | 12,020 | 113 |
|  | West | 1,220 | 23 | 5,680 | 105 |
| Total |  | 7,440 | 28 | 34,400 | 128 |

Source: Centers for Medicare & Medicaid Services, 5% Denominator and Claims Files. Beneficiaries aged 65+ years with continuous and full Parts A and B enrollment and no Health Maintenance Organization enrollment during the year. Unweighted counts were multiplied by 20 to produce counts for this table.

Among Medicare beneficiaries, hospital discharge rates with ulcerative colitis (all-listed diagnoses) increased with age until 85 years and were higher among women compared with men and Whites compared with Blacks (Table 96). Rates were highest in the Northeast, followed by the Midwest, then the South, and lowest in the West.

**Table 96:** Ulcerative Colitis: Hospital discharges with first-listed and all-listed diagnoses by age, race, sex and region among fee-for-service, age-eligible Medicare beneficiaries, 2019.

| Demographic Characteristics |  | First-Listed Diagnosis |  | All-Listed Diagnoses |  |
| --- | --- | --- | --- | --- | --- |
|  |  | Number of events | Event rate per 100,000 | Number of events | Event rate per 100,000 |
| Age (years) | 65 to 69 | 1,240 | 15 | 7,300 | 90 |
|  | 70 to 74 | 860 | 12 | 6,420 | 93 |
|  | 75 to 79 | 920 | 19 | 6,900 | 141 |
|  | 80 to 84 | 740 | 22 | 5,080 | 152 |
|  | 85+ | 660 | 18 | 5,380 | 150 |
| Race | White | 3,800 | 17 | 27,340 | 124 |
|  | Black | 300 | 16 | 1,560 | 84 |
|  | Other | 320 | 11 | 2,180 | 75 |
| Sex | Female | 2,720 | 18 | 17,840 | 119 |
|  | Male | 1,700 | 14 | 13,240 | 112 |
| Region | Northeast | 1,000 | 21 | 7,440 | 157 |
|  | Midwest | 1,280 | 21 | 8,100 | 133 |
|  | South | 1,560 | 15 | 10,640 | 100 |
|  | West | 580 | 11 | 4,900 | 91 |
| Total |  | 4,420 | 16 | 31,080 | 116 |

Source: Centers for Medicare & Medicaid Services, 5% Denominator and Claims Files. Beneficiaries aged 65+ years with continuous and full Parts A and B enrollment and no Health Maintenance Organization enrollment during the year. Unweighted counts were multiplied by 20 to produce counts for this table.

Diverticular disease contributed to 3.6 million ambulatory visits (2015) (Table 97). Ambulatory care visits were uncommon among children and then rates (all-listed diagnoses) increased with age. Age-adjusted rates were higher among women compared with men, Whites compared with Blacks, and Hispanics compared with non-Hispanics.

**Table 97:** Diverticular Disease: Ambulatory care visits with first-listed and all-listed diagnoses by age, race, ethnicity, and sex in the United States, 2015.

| Demographic Characteristics |  | First-Listed Diagnosis |  | All-Listed Diagnoses |  |
| --- | --- | --- | --- | --- | --- |
|  |  | Number in thousands | Rate per 100,000 | Number in thousands | Rate per 100,000 |
| Age (years) | 0 to 11 | . | . | . | . |
|  | 12 to 24 | 3 | 6 | 6 | 11 |
|  | 25 to 44 | 71 | 84 | 229 | 271 |
|  | 45 to 54 | 218 | 507 | 482 | 1,119 |
|  | 55 to 64 | 609 | 1,492 | 1,157 | 2,836 |
|  | 65 to 74 | 458 | 1,666 | 985 | 3,579 |
|  | 75 plus | 289 | 1,430 | 721 | 3,567 |
| Race | White | 1,365 | 415 | 3,184 | 1,006 |
|  | Black | 250 | 494 | 313 | 645 |
|  | Other | 33 | 115 | 85 | 331 |
| Ethnicity | Hispanic | 266 | 633 | 529 | 1,485 |
|  | Not Hispanic | 1,382 | 383 | 3,052 | 888 |
| Sex | Female | 1,077 | 498 | 2,232 | 1,074 |
|  | Male | 572 | 311 | 1,349 | 765 |
| Total |  | 1,649 | 412 | 3,581 | 936 |

Source: National Ambulatory Medical Care Survey (3-year average, 2014-2016). Rates for age groups are unadjusted and all other rates are age-adjusted.

Diverticular disease contributed to 1.5 million emergency department visits in 2018 (Table 98). Emergency department visits were uncommon among children and then rates (all-listed diagnoses) increased with age. Age-adjusted rates were higher among women compared with men.

**Table 98:**
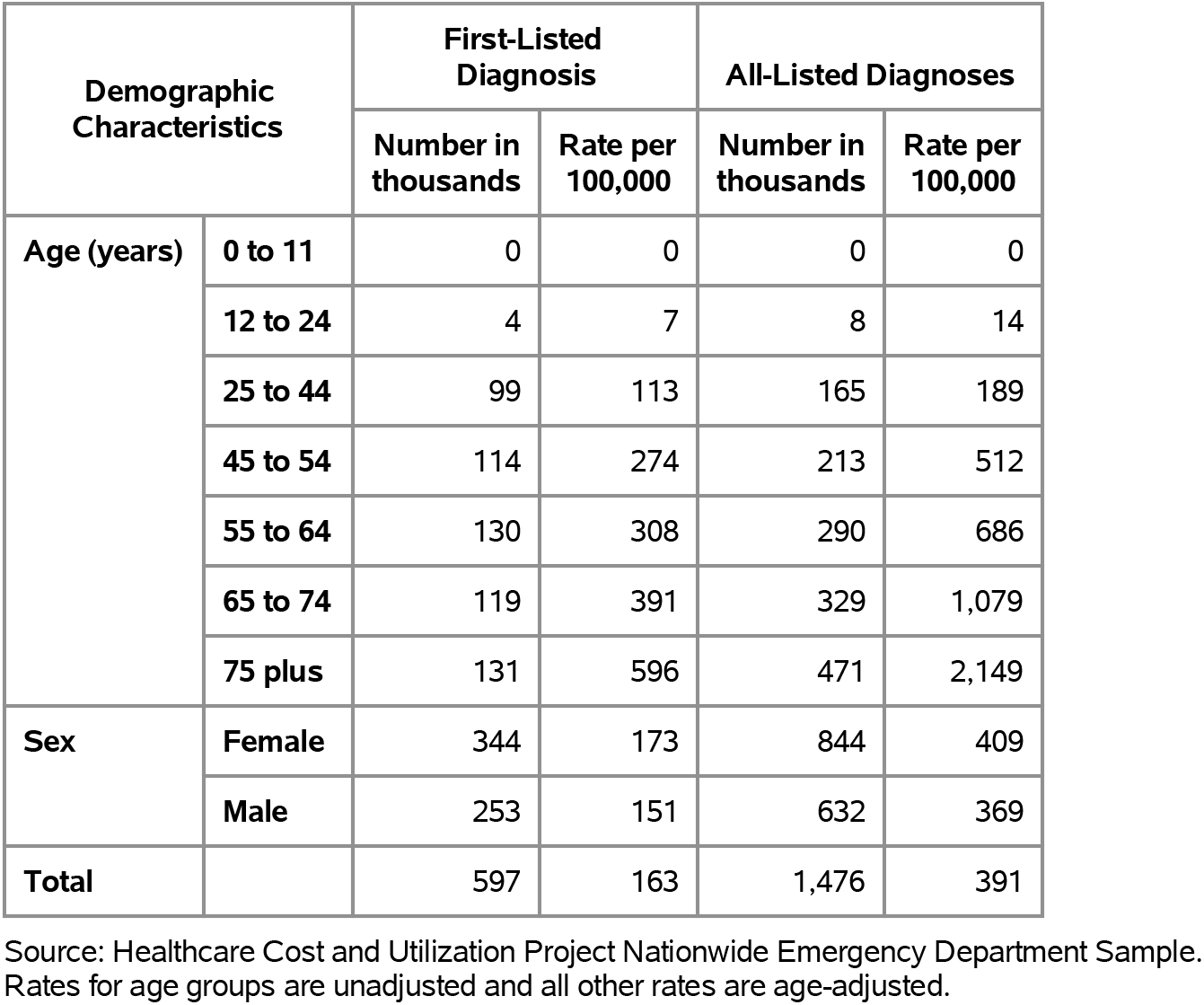
Diverticular Disease: Emergency department visits with first-listed and all-listed diagnoses by age and sex in the United States, 2018.

Diverticular disease contributed to 880,000 hospital discharges in 2018 (Table 99). Hospitalizations were uncommon among children and then hospital discharge rates (all-listed diagnoses) increased with age. Age-adjusted rates were higher among women compared with men, Blacks compared with Whites, and non-Hispanics compared with Hispanics. Between 2004 and 2018, age-adjusted hospital discharge rates (per 100,000) with an all-listed diagnosis decreased by 18% from 278 to 227.(4,6)

**Table 99:** Diverticular Disease: Hospital discharges with first-listed and all-listed diagnoses by age, race, ethnicity, and sex in the United States, 2018.

| Demographic Characteristics |  | First-Listed Diagnosis |  | All-Listed Diagnoses |  |
| --- | --- | --- | --- | --- | --- |
|  |  | Number in thousands | Rate per 100,000 | Number in thousands | Rate per 100,000 |
| Age (years) | 0 to 11 | 0 | 0 | 0 | 0 |
|  | 12 to 24 | 1 | 2 | 2 | 3 |
|  | 25 to 44 | 34 | 39 | 57 | 65 |
|  | 45 to 54 | 44 | 106 | 90 | 217 |
|  | 55 to 64 | 61 | 144 | 160 | 378 |
|  | 65 to 74 | 64 | 211 | 218 | 716 |
|  | 75 plus | 89 | 407 | 353 | 1,608 |
| Race | White | 248 | 81 | 745 | 231 |
|  | Black | 37 | 90 | 106 | 260 |
|  | Other | 13 | 51 | 39 | 164 |
| Ethnicity | Hispanic | 31 | 71 | 91 | 225 |
|  | Not Hispanic | 266 | 80 | 800 | 230 |
| Sex | Female | 165 | 79 | 492 | 228 |
|  | Male | 129 | 76 | 387 | 223 |
| Total |  | 294 | 78 | 880 | 227 |

Source: Healthcare Cost and Utilization Project National Inpatient Sample. Rates for age groups are unadjusted and all other rates are age-adjusted.

Diverticular disease contributed to 5,000 deaths in 2019 (Table 100). Mortality was uncommon among the youngest age groups after which rates (underlying or other cause) increased with age. Age-adjusted mortality rates were higher among women, Whites, and non-Hispanics. Between 2004 and 2019, age-adjusted mortality rates (per 100,000) with diverticular disease as underlying or other cause decreased by 40% from 2.0 to 1.2.(4)

**Table 100:** Diverticular Disease: Deaths with underlying or underlying/other cause and lifetime years of life lost by age, race, ethnicity, and sex in the United States, 2019.

| Demographic Characteristics |  | Underlying Cause |  |  | Underlying/Other Cause |  |  |
| --- | --- | --- | --- | --- | --- | --- | --- |
|  |  | Number of Deaths | Rate per 100,000 | Years of potential life lost in thousands | Number of Deaths | Rate per 100,000 | Years of potential life lost in thousands |
| Age (years) | 0 to 11 | 1 | 0.0 | 0.1 | 1 | 0.0 | 0.1 |
|  | 12 to 24 | . | . | . | . | . | . |
|  | 25 to 44 | 24 | 0.0 | 1.0 | 42 | 0.0 | 1.7 |
|  | 45 to 54 | 72 | 0.2 | 2.2 | 127 | 0.3 | 3.9 |
|  | 55 to 64 | 276 | 0.7 | 6.3 | 445 | 1.0 | 10.2 |
|  | 65 to 74 | 532 | 1.7 | 8.4 | 908 | 2.9 | 14.3 |
|  | 75 plus | 2,014 | 8.9 | 13.6 | 3,262 | 14.4 | 22.0 |
| Race | White | 2,616 | 0.8 | 27.7 | 4,283 | 1.2 | 45.9 |
|  | Black | 234 | 0.6 | 3.2 | 385 | 1.0 | 4.9 |
|  | Other | 69 | 0.3 | 0.8 | 117 | 0.5 | 1.3 |
| Ethnicity | Hispanic | 176 | 0.5 | 2.5 | 299 | 0.8 | 4.2 |
|  | Not Hispanic | 2,743 | 0.7 | 29.1 | 4,486 | 1.2 | 48.0 |
| Sex | Female | 1,932 | 0.8 | 20.4 | 3,043 | 1.3 | 32.0 |
|  | Male | 987 | 0.6 | 11.2 | 1,742 | 1.0 | 20.2 |
| Total |  | 2,919 | 0.7 | 31.6 | 4,785 | 1.2 | 52.1 |

Source: Vital Statistics of the United States.
Rates for age groups are unadjusted and all other rates are age-adjusted.
Years of potential life lost is based on U.S. life table 2006-2018 estimates of life expectancy at age of death.

Among privately insured enrollees, the claims-based prevalence of diverticular disease (based on all-listed diagnoses) was 2.9% (Table 101). Diverticular disease was uncommon in childhood and adolescence and then prevalence increased with age until the oldest age group. Prevalence was higher among women. It was highest among Blacks, followed by Whites and Hispanics, and lowest among Asians. It was highest in the Northeast, followed by the South and Midwest, and lowest in the West.

**Table 101:**
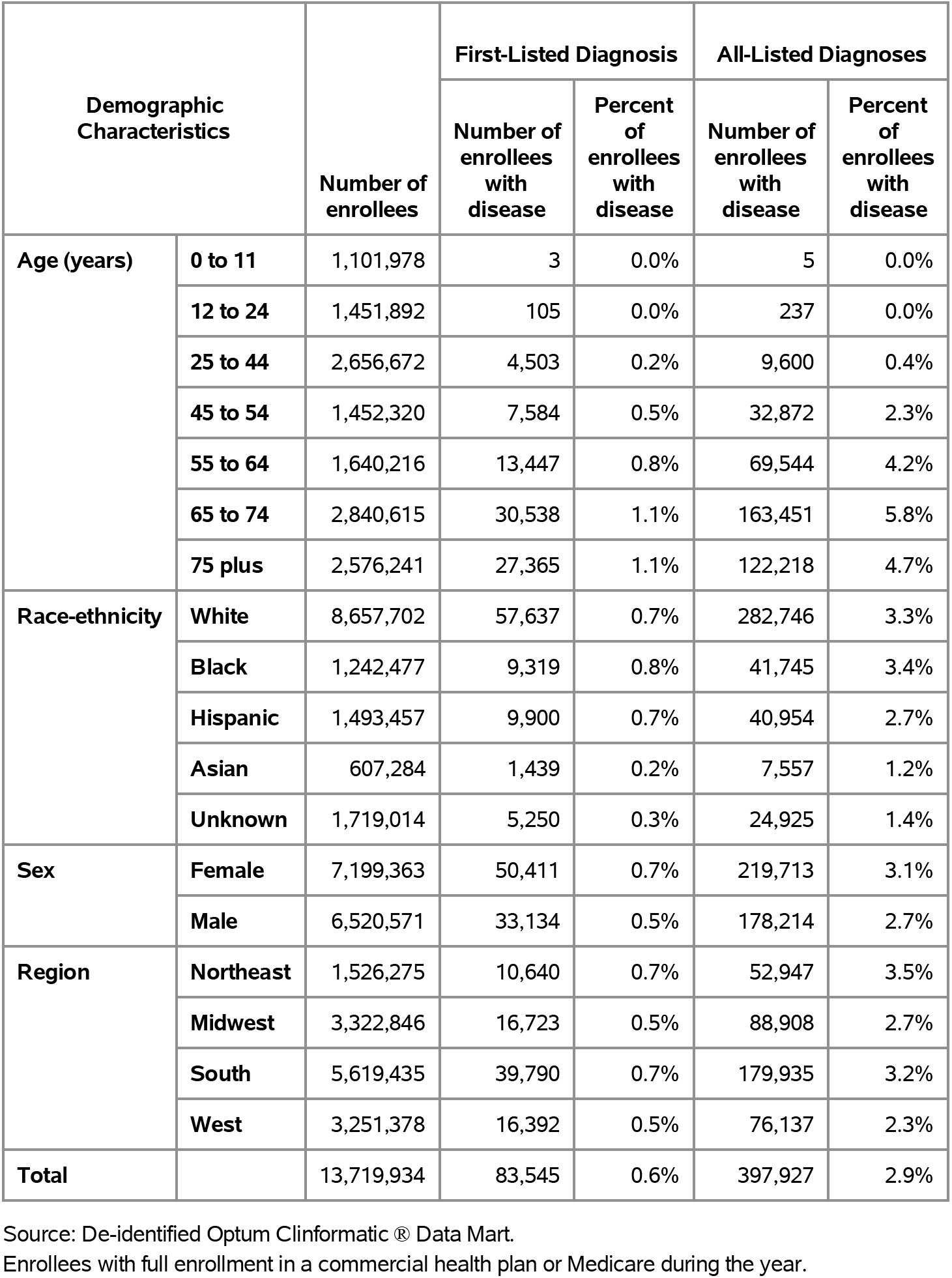
Diverticular Disease: Claims-based prevalence with first-listed and all-listed diagnoses by age, race-ethnicity, sex and region among privately insured enrollees, 2020.

| Demographic Characteristics |  | Number of enrollees | First-Listed Diagnosis |  | All-Listed Diagnoses |  |
| --- | --- | --- | --- | --- | --- | --- |
|  |  |  | Number of enrollees with disease | Percent of enrollees with disease | Number of enrollees with disease | Percent of enrollees with disease |
| Age (years) | 0 to 11 | 1,101,978 | 3 | 0.0% | 5 | 0.0% |
|  | 12 to 24 | 1,451,892 | 105 | 0.0% | 237 | 0.0% |
|  | 25 to 44 | 2,656,672 | 4,503 | 0.2% | 9,600 | 0.4% |
|  | 45 to 54 | 1,452,320 | 7,584 | 0.5% | 32,872 | 2.3% |
|  | 55 to 64 | 1,640,216 | 13,447 | 0.8% | 69,544 | 4.2% |
|  | 65 to 74 | 2,840,615 | 30,538 | 1.1% | 163,451 | 5.8% |
|  | 75 plus | 2,576,241 | 27,365 | 1.1% | 122,218 | 4.7% |
| Race-ethnicity | White | 8,657,702 | 57,637 | 0.7% | 282,746 | 3.3% |
|  | Black | 1,242,477 | 9,319 | 0.8% | 41,745 | 3.4% |
|  | Hispanic | 1,493,457 | 9,900 | 0.7% | 40,954 | 2.7% |
|  | Asian | 607,284 | 1,439 | 0.2% | 7,557 | 1.2% |
|  | Unknown | 1,719,014 | 5,250 | 0.3% | 24,925 | 1.4% |
| Sex | Female | 7,199,363 | 50,411 | 0.7% | 219,713 | 3.1% |
|  | Male | 6,520,571 | 33,134 | 0.5% | 178,214 | 2.7% |
| Region | Northeast | 1,526,275 | 10,640 | 0.7% | 52,947 | 3.5% |
|  | Midwest | 3,322,846 | 16,723 | 0.5% | 88,908 | 2.7% |
|  | South | 5,619,435 | 39,790 | 0.7% | 179,935 | 3.2% |
|  | West | 3,251,378 | 16,392 | 0.5% | 76,137 | 2.3% |
| Total |  | 13,719,934 | 83,545 | 0.6% | 397,927 | 2.9% |

Source: De-identified Optum Clinformatic® Data Mart.
Enrollees with full enrollment in a commercial health plan or Medicare during the year.

Among commercial insurance enrollees, ambulatory care visit rates with diverticular disease (all-listed diagnoses) increased with age until 75 years and were higher among women compared with men (Table 102). Among persons with known race-ethnicity, rates were highest among Blacks, followed by Whites, then Hispanics, and lowest among Asians. Rates were highest in the Northeast, followed by the South, then the Midwest, and lowest in the West.

**Table 102:**
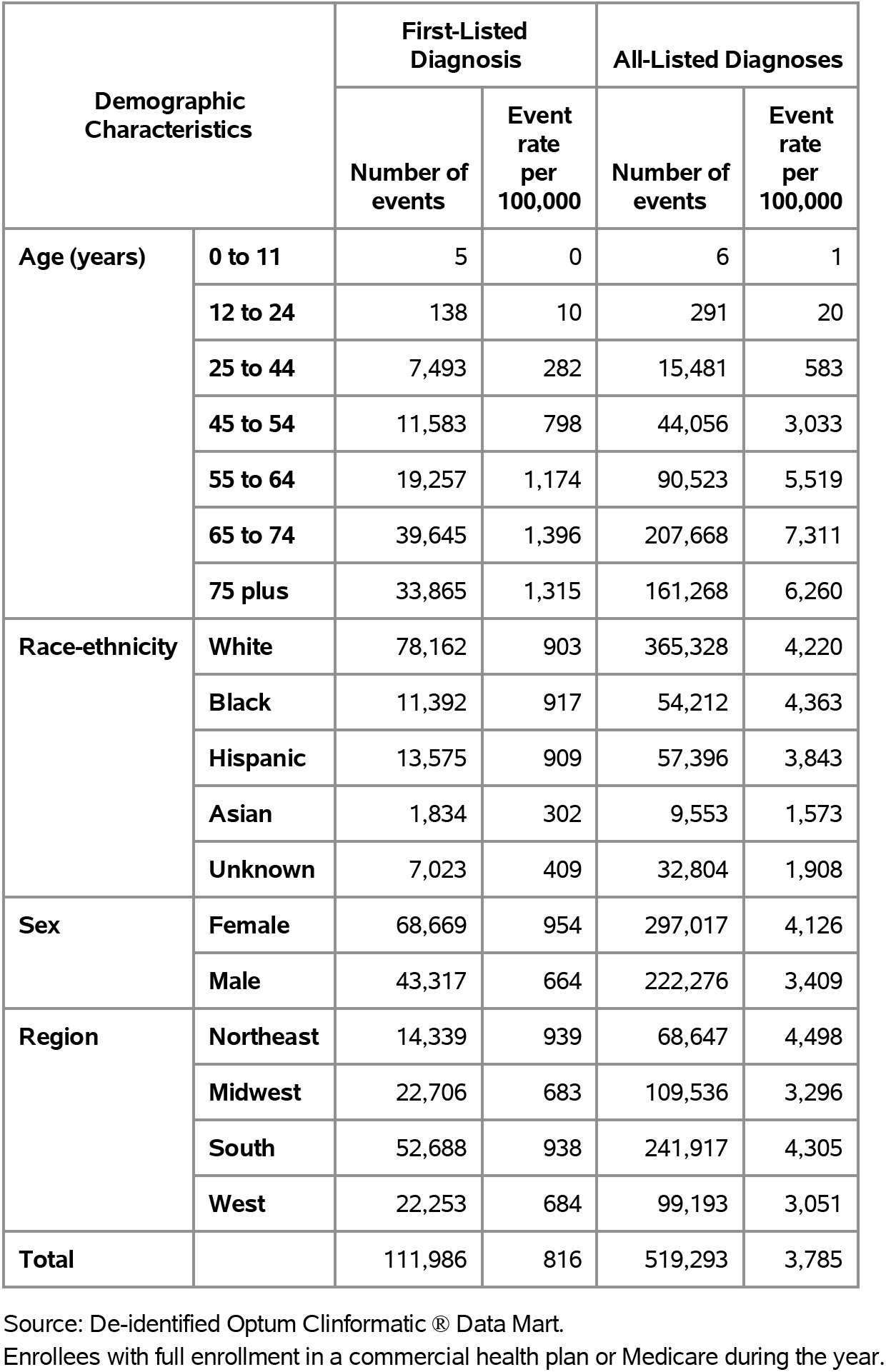
Diverticular Disease: Ambulatory care visits with first-listed and all-listed diagnoses by age, race-ethnicity, sex and region among privately insured enrollees, 2020.

| Demographic Characteristics |  | First-Listed Diagnosis |  | All-Listed Diagnoses |  |
| --- | --- | --- | --- | --- | --- |
|  |  | Number of events | Event rate per 100,000 | Number of events | Event rate per 100,000 |
| Age (years) | 0 to 11 | 5 | 0 | 6 | 1 |
|  | 12 to 24 | 138 | 10 | 291 | 20 |
|  | 25 to 44 | 7,493 | 282 | 15,481 | 583 |
|  | 45 to 54 | 11,583 | 798 | 44,056 | 3,033 |
|  | 55 to 64 | 19,257 | 1,174 | 90,523 | 5,519 |
|  | 65 to 74 | 39,645 | 1,396 | 207,668 | 7,311 |
|  | 75 plus | 33,865 | 1,315 | 161,268 | 6,260 |
| Race-ethnicity | White | 78,162 | 903 | 365,328 | 4,220 |
|  | Black | 11,392 | 917 | 54,212 | 4,363 |
|  | Hispanic | 13,575 | 909 | 57,396 | 3,843 |
|  | Asian | 1,834 | 302 | 9,553 | 1,573 |
|  | Unknown | 7,023 | 409 | 32,804 | 1,908 |
| Sex | Female | 68,669 | 954 | 297,017 | 4,126 |
|  | Male | 43,317 | 664 | 222,276 | 3,409 |
| Region | Northeast | 14,339 | 939 | 68,647 | 4,498 |
|  | Midwest | 22,706 | 683 | 109,536 | 3,296 |
|  | South | 52,688 | 938 | 241,917 | 4,305 |
|  | West | 22,253 | 684 | 99,193 | 3,051 |
| Total |  | 111,986 | 816 | 519,293 | 3,785 |

Source: De-identified Optum Clinformatic® Data Mart.
Enrollees with full enrollment in a commercial health plan or Medicare during the year.

Among commercial insurance enrollees, emergency department visit rates with diverticular disease (all-listed diagnoses) increased with age and were higher among women compared with men (Table 103). Among persons with known race-ethnicity, rates were highest among Blacks, followed by Whites, then Hispanics, and lowest among Asians. Rates were highest in the Northeast, followed by the South, then the West, and lowest in the Midwest.

**Table 103:** Diverticular Disease: Emergency department visits with first-listed and all-listed diagnoses by age, race-ethnicity, sex and region among privately insured enrollees, 2020.

| Demographic Characteristics |  | First-Listed Diagnosis |  | All-Listed Diagnoses |  |
| --- | --- | --- | --- | --- | --- |
|  |  | Number of events | Event rate per 100,000 | Number of events | Event rate per 100,000 |
| Age (years) | 0 to 11 | . | . | . | . |
|  | 12 to 24 | 2 | 0 | 5 | 0 |
|  | 25 to 44 | 218 | 8 | 444 | 17 |
|  | 45 to 54 | 485 | 33 | 1,169 | 80 |
|  | 55 to 64 | 1,277 | 78 | 2,981 | 182 |
|  | 65 to 74 | 5,894 | 207 | 9,379 | 330 |
|  | 75 plus | 6,125 | 238 | 10,503 | 408 |
| Race-ethnicity | White | 9,583 | 111 | 16,888 | 195 |
|  | Black | 1,659 | 134 | 2,816 | 227 |
|  | Hispanic | 1,639 | 110 | 2,792 | 187 |
|  | Asian | 177 | 29 | 357 | 59 |
|  | Unknown | 943 | 55 | 1,628 | 95 |
| Sex | Female | 9,545 | 133 | 15,880 | 221 |
|  | Male | 4,456 | 68 | 8,601 | 132 |
| Region | Northeast | 2,011 | 132 | 3,182 | 208 |
|  | Midwest | 3,025 | 91 | 5,073 | 153 |
|  | South | 6,073 | 108 | 11,059 | 197 |
|  | West | 2,892 | 89 | 5,167 | 159 |
| Total |  | 14,001 | 102 | 24,481 | 178 |

Source: De-identified Optum Clinformatic® Data Mart.
Enrollees with full enrollment in a commercial health plan or Medicare during the year.

Among commercial insurance enrollees, hospital discharge rates with diverticular disease (all-listed diagnoses) increased with age and were higher among women compared with men (Table 104). Among persons with known race-ethnicity, rates were highest among Blacks, followed by Whites, then Hispanics, and lowest among Asians. Rates were highest in the Northeast, followed by the South, then the Midwest, and lowest in the West.

**Table 104:**
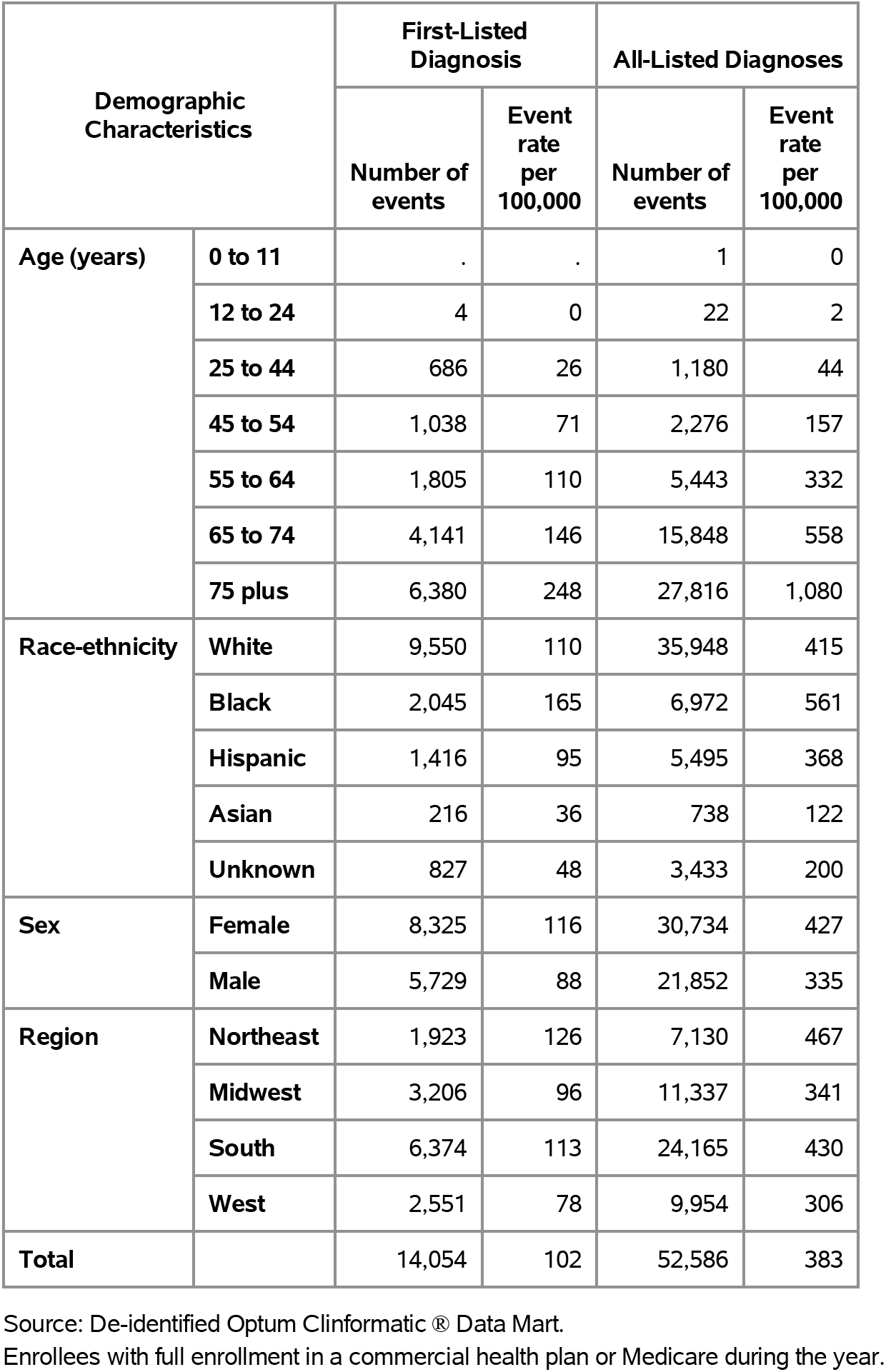
Diverticular Disease: Hospital discharges with first-listed and all-listed diagnoses by age, race-ethnicity, sex and region among privately insured enrollees, 2020.

Among Medicare beneficiaries, the claims-based prevalence of diverticular disease (based on all-listed diagnoses) was 7.1% (Table 105). Prevalence peaked among persons 75-79 years and was higher among women and Whites. It was highest in the South and Northeast, followed by Midwest, and lowest in the West.

**Table 105:** Diverticular Disease: Claims-based prevalence with first-listed and all-listed diagnoses by age, race, sex and region among fee-for-service, age-eligible Medicare beneficiaries, 2019.

| Demographic Characteristics |  | Number of enrollees | First-Listed Diagnosis |  | All-Listed Diagnoses |  |
| --- | --- | --- | --- | --- | --- | --- |
|  |  |  | Number of enrollees with disease | Percent of enrollees with disease | Number of enrollees with disease | Percent of enrollees with disease |
| Age (years) | 65 to 69 | 8,066,320 | 113,280 | 1.4% | 538,180 | 6.7% |
|  | 70 to 74 | 6,936,940 | 122,180 | 1.8% | 546,660 | 7.9% |
|  | 75 to 79 | 4,894,060 | 93,540 | 1.9% | 398,100 | 8.1% |
|  | 80 to 84 | 3,335,340 | 61,800 | 1.9% | 232,840 | 7.0% |
|  | 85+ | 3,578,120 | 55,400 | 1.5% | 196,700 | 5.5% |
| Race | White | 22,023,800 | 375,600 | 1.7% | 1,624,300 | 7.4% |
|  | Black | 1,861,660 | 30,260 | 1.6% | 121,680 | 6.5% |
|  | Other | 2,925,320 | 40,340 | 1.4% | 166,500 | 5.7% |
| Sex | Female | 14,975,560 | 281,640 | 1.9% | 1,088,100 | 7.3% |
|  | Male | 11,835,220 | 164,560 | 1.4% | 824,380 | 7.0% |
| Region | Northeast | 4,748,700 | 86,540 | 1.8% | 367,820 | 7.7% |
|  | Midwest | 6,069,800 | 83,660 | 1.4% | 404,820 | 6.7% |
|  | South | 10,595,900 | 193,460 | 1.8% | 822,700 | 7.8% |
|  | West | 5,396,380 | 82,540 | 1.5% | 317,140 | 5.9% |
| Total |  | 26,810,780 | 446,200 | 1.7% | 1,912,480 | 7.1% |

Source: Centers for Medicare & Medicaid Services, 5% Denominator and Claims Files. Beneficiaries aged 65+ years with continuous and full Parts A and B enrollment and no Health Maintenance Organization enrollment during the year. Unweighted counts were multiplied by 20 to produce counts for this table.

Among Medicare beneficiaries, ambulatory care visit rates with diverticular disease (all-listed diagnoses) peaked among persons 75 to 79 years and were higher among women compared with men and Whites compared with Blacks (Table 106). Rates were higher in the South and Northeast compared with the Midwest and West.

**Table 106:** Diverticular Disease: Ambulatory care visits with first-listed and all-listed diagnoses by age, race, sex and region among fee-for-service, age-eligible Medicare beneficiaries, 2019.

| Demographic Characteristics |  | First-Listed Diagnosis |  | All-Listed Diagnoses |  |
| --- | --- | --- | --- | --- | --- |
|  |  | Number of events | Event rate per 100,000 | Number of events | Event rate per 100,000 |
| Age (years) | 65 to 69 | 134,900 | 1,672 | 675,960 | 8,380 |
|  | 70 to 74 | 143,540 | 2,069 | 680,380 | 9,808 |
|  | 75 to 79 | 101,580 | 2,076 | 487,080 | 9,952 |
|  | 80 to 84 | 63,460 | 1,903 | 268,120 | 8,039 |
|  | 85+ | 45,900 | 1,283 | 190,400 | 5,321 |
| Race | White | 417,160 | 1,894 | 1,951,200 | 8,860 |
|  | Black | 28,760 | 1,545 | 141,860 | 7,620 |
|  | Other | 43,460 | 1,486 | 208,880 | 7,140 |
| Sex | Female | 321,180 | 2,145 | 1,361,240 | 9,090 |
|  | Male | 168,200 | 1,421 | 940,700 | 7,948 |
| Region | Northeast | 97,220 | 2,047 | 449,880 | 9,474 |
|  | Midwest | 89,240 | 1,470 | 453,980 | 7,479 |
|  | South | 210,800 | 1,989 | 1,018,120 | 9,609 |
|  | West | 92,120 | 1,707 | 379,960 | 7,041 |
| Total |  | 489,380 | 1,825 | 2,301,940 | 8,586 |

Source: Centers for Medicare & Medicaid Services, 5% Denominator and Claims Files. Beneficiaries aged 65+ years with continuous and full Parts A and B enrollment and no Health Maintenance Organization enrollment during the year. Unweighted counts were multiplied by 20 to produce counts for this table.

Among Medicare beneficiaries, emergency department visit rates with diverticular disease (all-listed diagnoses) increased with age and were higher among women compared with men and Blacks compared with Whites (Table 107). Rates were highest in the South, followed by the Northeast, then the Midwest, and lowest in the West.

**Table 107:** Diverticular Disease: Emergency department visits with first-listed and all-listed diagnoses by age, race, sex and region among fee-for-service, age-eligible Medicare beneficiaries, 2019.

| Demographic Characteristics |  | First-Listed Diagnosis |  | All-Listed Diagnoses |  |
| --- | --- | --- | --- | --- | --- |
|  |  | Number of events | Event rate per 100,000 | Number of events | Event rate per 100,000 |
| Age (years) | 65 to 69 | 40,420 | 501 | 96,900 | 1,201 |
|  | 70 to 74 | 43,380 | 625 | 116,860 | 1,685 |
|  | 75 to 79 | 38,260 | 782 | 111,600 | 2,280 |
|  | 80 to 84 | 29,660 | 889 | 96,780 | 2,902 |
|  | 85+ | 37,200 | 1,040 | 129,400 | 3,616 |
| Race | White | 155,900 | 708 | 457,240 | 2,076 |
|  | Black | 16,760 | 900 | 47,780 | 2,567 |
|  | Other | 16,260 | 556 | 46,520 | 1,590 |
| Sex | Female | 123,060 | 822 | 338,580 | 2,261 |
|  | Male | 65,860 | 556 | 212,960 | 1,799 |
| Region | Northeast | 36,860 | 776 | 102,940 | 2,168 |
|  | Midwest | 39,420 | 649 | 117,540 | 1,936 |
|  | South | 81,100 | 765 | 237,100 | 2,238 |
|  | West | 31,540 | 584 | 93,960 | 1,741 |
| Total |  | 188,920 | 705 | 551,540 | 2,057 |

Source: Centers for Medicare & Medicaid Services, 5% Denominator and Claims Files. Beneficiaries aged 65+ years with continuous and full Parts A and B enrollment and no Health Maintenance Organization enrollment during the year. Unweighted counts were multiplied by 20 to produce counts for this table.

Among Medicare beneficiaries, hospital discharge rates with diverticular disease (all-listed diagnoses) increased with age and were higher among women compared with men and Blacks compared with Whites (Table 108). Rates were lower in the West compared with the South, Northeast, and Midwest.

**Table 108:**
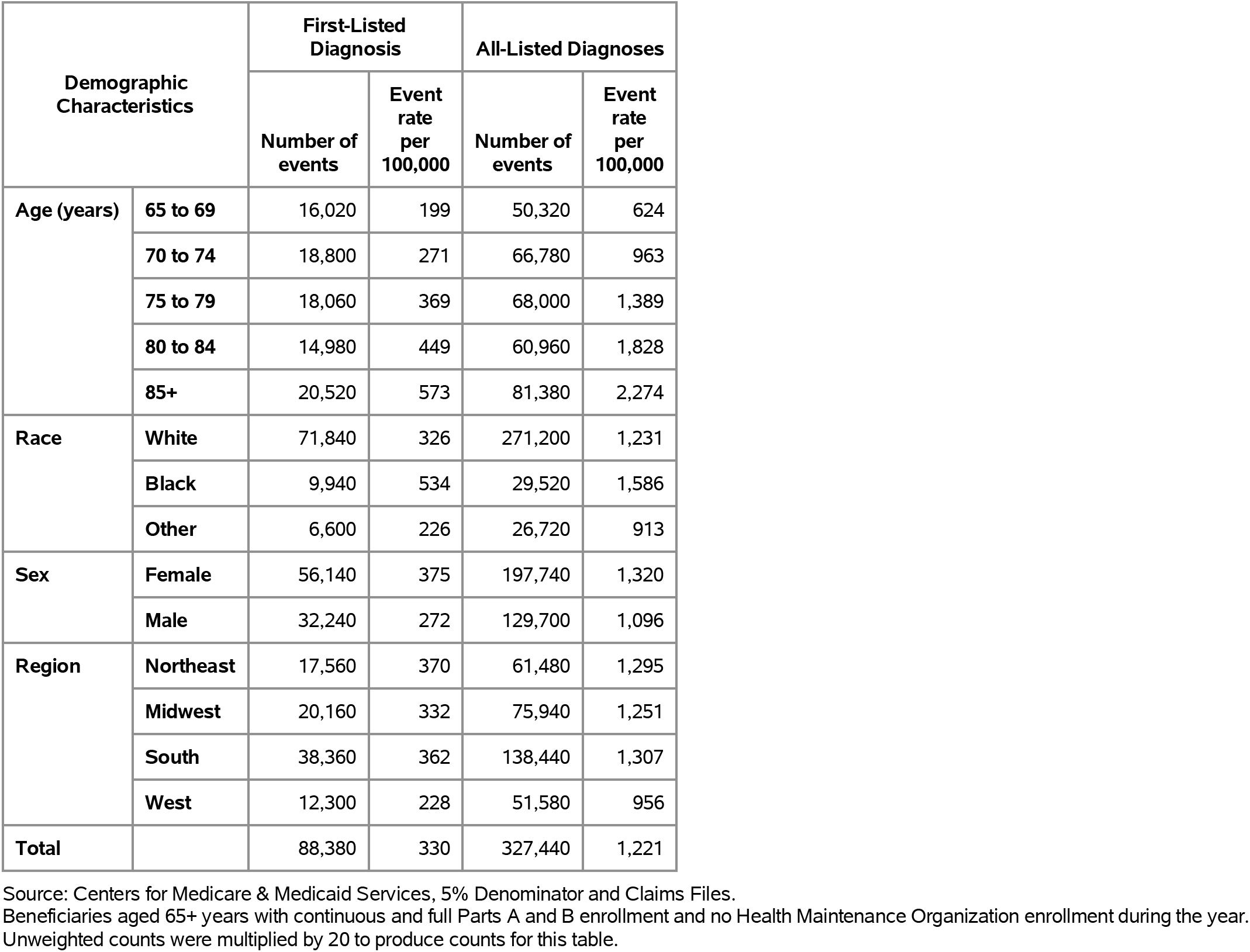
Diverticular Disease: Hospital discharges with first-listed and all-listed diagnoses by age, race, sex and region among fee-for-service, age-eligible Medicare beneficiaries, 2019.

| Demographic Characteristics |  | First-Listed Diagnosis |  | All-Listed Diagnoses |  |
| --- | --- | --- | --- | --- | --- |
|  |  | Number of events | Event rate per 100,000 | Number of events | Event rate per 100,000 |
| Age (years) | 65 to 69 | 16,020 | 199 | 50,320 | 624 |
|  | 70 to 74 | 18,800 | 271 | 66,780 | 963 |
|  | 75 to 79 | 18,060 | 369 | 68,000 | 1,389 |
|  | 80 to 84 | 14,980 | 449 | 60,960 | 1,828 |
|  | 85+ | 20,520 | 573 | 81,380 | 2,274 |
| Race | White | 71,840 | 326 | 271,200 | 1,231 |
|  | Black | 9,940 | 534 | 29,520 | 1,586 |
|  | Other | 6,600 | 226 | 26,720 | 913 |
| Sex | Female | 56,140 | 375 | 197,740 | 1,320 |
|  | Male | 32,240 | 272 | 129,700 | 1,096 |
| Region | Northeast | 17,560 | 370 | 61,480 | 1,295 |
|  | Midwest | 20,160 | 332 | 75,940 | 1,251 |
|  | South | 38,360 | 362 | 138,440 | 1,307 |
|  | West | 12,300 | 228 | 51,580 | 956 |
| Total |  | 88,380 | 330 | 327,440 | 1,221 |

Source: Centers for Medicare & Medicaid Services, 5% Denominator and Claims Files. Beneficiaries aged 65+ years with continuous and full Parts A and B enrollment and no Health Maintenance Organization enrollment during the year. Unweighted counts were multiplied by 20 to produce counts for this table.

Hemorrhoids contributed to 4.0 million ambulatory visits (2015) (Table 109). Ambulatory care visit rates (all-listed diagnoses) were similar among middle-aged and older adults. Age-adjusted rates were higher among men compared with women, Blacks compared with Whites, and Hispanics compared with non-Hispanics.

**Table 109:** Hemorrhoids: Ambulatory care visits with first-listed and all-listed diagnoses by age, race, ethnicity, and sex in the United States, 2015.

| Demographic Characteristics |  | First-Listed Diagnosis |  | All-Listed Diagnoses |  |
| --- | --- | --- | --- | --- | --- |
|  |  | Number in thousands | Rate per 100,000 | Number in thousands | Rate per 100,000 |
| Age (years) | 0 to 11 | . | . | 4 | 8 |
|  | 12 to 24 | 81 | 144 | 112 | 200 |
|  | 25 to 44 | 421 | 498 | 888 | 1,051 |
|  | 45 to 54 | 546 | 1,267 | 1,079 | 2,505 |
|  | 55 to 64 | 418 | 1,025 | 789 | 1,933 |
|  | 65 to 74 | 306 | 1,111 | 660 | 2,397 |
|  | 75 plus | 215 | 1,063 | 504 | 2,492 |
| Race | White | 1,426 | 522 | 2,948 | 1,054 |
|  | Black | 200 | 452 | 619 | 1,392 |
|  | Other | 360 | 1,503 | 469 | 1,966 |
| Ethnicity | Hispanic | 561 | 1,217 | 839 | 1,787 |
|  | Not Hispanic | 1,425 | 483 | 3,197 | 1,064 |
| Sex | Female | 900 | 503 | 2,052 | 1,158 |
|  | Male | 1,086 | 672 | 1,983 | 1,195 |
| Total |  | 1,986 | 584 | 4,036 | 1,176 |

Source: National Ambulatory Medical Care Survey (3-year average, 2014-2016). Rates for age groups are unadjusted and all other rates are age-adjusted.

Hemorrhoids contributed to 603,000 emergency department visits in 2018 (Table 110). Emergency department visit rates (all-listed diagnoses) increased with age. Age-adjusted rates were higher among men compared with women.

**Table 110:** Hemorrhoids: Emergency department visits with first-listed and all-listed diagnoses by age and sex in the United States, 2018.

| Demographic Characteristics |  | First-Listed Diagnosis |  | All-Listed Diagnoses |  |
| --- | --- | --- | --- | --- | --- |
|  |  | Number in thousands | Rate per 100,000 | Number in thousands | Rate per 100,000 |
| Age (years) | 0 to 11 | 1 | 2 | 2 | 4 |
|  | 12 to 24 | 23 | 41 | 36 | 65 |
|  | 25 to 44 | 77 | 89 | 140 | 161 |
|  | 45 to 54 | 30 | 72 | 81 | 194 |
|  | 55 to 64 | 22 | 52 | 94 | 221 |
|  | 65 to 74 | 17 | 55 | 100 | 329 |
|  | 75 plus | 23 | 106 | 149 | 681 |
| Sex | Female | 91 | 53 | 316 | 166 |
|  | Male | 102 | 64 | 287 | 170 |
| Total |  | 193 | 58 | 603 | 169 |

Source: Healthcare Cost and Utilization Project Nationwide Emergency Department Sample. Rates for age groups are unadjusted and all other rates are age-adjusted.

Hemorrhoids contributed to 347,000 hospital discharges in 2018 (Table 111). Hospital discharge rates (all-listed diagnoses) increased with age. Age-adjusted rates were higher among men compared with women, Blacks compared with Whites, and Hispanics compared with non-Hispanics. Between 2004 and 2018, age-adjusted hospital discharge rates (per 100,000) with an all-listed diagnosis decreased by 12% from 104 to 91.(4,6)

**Table 111:** Hemorrhoids: Hospital discharges with first-listed and all-listed diagnoses by age, race, ethnicity, and sex in the United States, 2018.

| Demographic Characteristics |  | First-Listed Diagnosis |  | All-Listed Diagnoses |  |
| --- | --- | --- | --- | --- | --- |
|  |  | Number in thousands | Rate per 100,000 | Number in thousands | Rate per 100,000 |
| Age (years) | 0 to 11 | 0 | 0 | 1 | 1 |
|  | 12 to 24 | 0 | 0 | 4 | 8 |
|  | 25 to 44 | 4 | 4 | 35 | 40 |
|  | 45 to 54 | 3 | 8 | 40 | 95 |
|  | 55 to 64 | 4 | 10 | 65 | 153 |
|  | 65 to 74 | 5 | 15 | 82 | 268 |
|  | 75 plus | 9 | 40 | 121 | 552 |
| Race | White | 19 | 6 | 270 | 86 |
|  | Black | 5 | 11 | 58 | 137 |
|  | Other | 2 | 9 | 24 | 97 |
| Ethnicity | Hispanic | 3 | 8 | 40 | 96 |
|  | Not Hispanic | 22 | 7 | 311 | 92 |
| Sex | Female | 13 | 6 | 184 | 89 |
|  | Male | 12 | 7 | 163 | 93 |
| Total |  | 25 | 7 | 347 | 91 |

Source: Healthcare Cost and Utilization Project National Inpatient Sample. Rates for age groups are unadjusted and all other rates are age-adjusted.

Hemorrhoids contributed to <1,000 deaths in 2019 (Table 112). Mortality from hemorrhoids was rare, but occasionally occurred among older adults. Between 2004 and 2019, mortality with hemorrhoids as underlying or other cause remained uncommon.(4)

**Table 112:** Hemorrhoids: Deaths with underlying or underlying/other cause and lifetime years of life lost by age, race, ethnicity, and sex in the United States, 2019.

| Demographic Characteristics |  | Underlying Cause |  |  | Underlying/Other Cause |  |  |
| --- | --- | --- | --- | --- | --- | --- | --- |
|  |  | Number of Deaths | Rate per 100,000 | Years of potential life lost in thousands | Number of Deaths | Rate per 100,000 | Years of potential life lost in thousands |
| Age (years) | 0 to 11 | . | . | . | . | . | . |
|  | 12 to 24 | . | . | . | . | . | . |
|  | 25 to 44 | 4 | 0.0 | 0.2 | 7 | 0.0 | 0.3 |
|  | 45 to 54 | 1 | 0.0 | 0.0 | 5 | 0.0 | 0.1 |
|  | 55 to 64 | 2 | 0.0 | 0.0 | 20 | 0.0 | 0.5 |
|  | 65 to 74 | 6 | 0.0 | 0.1 | 18 | 0.1 | 0.3 |
|  | 75 plus | 11 | 0.0 | 0.1 | 41 | 0.2 | 0.2 |
| Race | White | 19 | 0.0 | 0.3 | 69 | 0.0 | 1.0 |
|  | Black | 2 | 0.0 | 0.0 | 12 | 0.0 | 0.2 |
|  | Other | 3 | 0.0 | 0.1 | 10 | 0.0 | 0.2 |
| Ethnicity | Hispanic | 4 | 0.0 | 0.1 | 11 | 0.0 | 0.2 |
|  | Not Hispanic | 20 | 0.0 | 0.3 | 80 | 0.0 | 1.2 |
| Sex | Female | 11 | 0.0 | 0.2 | 47 | 0.0 | 0.7 |
|  | Male | 13 | 0.0 | 0.2 | 44 | 0.0 | 0.7 |
| Total |  | 24 | 0.0 | 0.4 | 91 | 0.0 | 1.4 |

Source: Vital Statistics of the United States.
Rates for age groups are unadjusted and all other rates are age-adjusted.
Years of potential life lost is based on U.S. life table 2006-2018 estimates of life expectancy at age of death.

Among privately insured enrollees, the claims-based prevalence of hemorrhoids (based on all-listed diagnoses) was 2.7% (Table 113). Prevalence increased with age until the oldest age group and was higher among women. It was highest among Blacks, followed by Whites and Hispanics, and lowest among Asians. It was highest in the Northeast, followed by the South, and lowest in the Midwest and West.

**Table 113:** Hemorrhoids: Claims-based prevalence with first-listed and all-listed diagnoses by age, race-ethnicity, sex and region among privately insured enrollees, 2020.

| Demographic Characteristics |  | Number of enrollees | First-Listed Diagnosis |  | All-Listed Diagnoses |  |
| --- | --- | --- | --- | --- | --- | --- |
|  |  |  | Number of enrollees with disease | Percent of enrollees with disease | Number of enrollees with disease | Percent of enrollees with disease |
| Age (years) | 0 to 11 | 1,101,978 | 188 | 0.0% | 545 | 0.0% |
|  | 12 to 24 | 1,451,892 | 2,079 | 0.1% | 4,756 | 0.3% |
|  | 25 to 44 | 2,656,672 | 14,761 | 0.6% | 36,906 | 1.4% |
|  | 45 to 54 | 1,452,320 | 9,720 | 0.7% | 50,138 | 3.5% |
|  | 55 to 64 | 1,640,216 | 10,428 | 0.6% | 70,544 | 4.3% |
|  | 65 to 74 | 2,840,615 | 16,936 | 0.6% | 130,407 | 4.6% |
|  | 75 plus | 2,576,241 | 13,195 | 0.5% | 76,797 | 3.0% |
| Race-ethnicity | White | 8,657,702 | 43,330 | 0.5% | 249,109 | 2.9% |
|  | Black | 1,242,477 | 6,838 | 0.6% | 37,975 | 3.1% |
|  | Hispanic | 1,493,457 | 8,416 | 0.6% | 42,185 | 2.8% |
|  | Asian | 607,284 | 3,201 | 0.5% | 14,031 | 2.3% |
|  | Unknown | 1,719,014 | 5,522 | 0.3% | 26,793 | 1.6% |
| Sex | Female | 7,199,363 | 35,481 | 0.5% | 199,197 | 2.8% |
|  | Male | 6,520,571 | 31,826 | 0.5% | 170,896 | 2.6% |
| Region | Northeast | 1,526,275 | 8,670 | 0.6% | 49,981 | 3.3% |
|  | Midwest | 3,322,846 | 13,463 | 0.4% | 81,198 | 2.4% |
|  | South | 5,619,435 | 29,700 | 0.5% | 160,730 | 2.9% |
|  | West | 3,251,378 | 15,474 | 0.5% | 78,184 | 2.4% |
| Total |  | 13,719,934 | 67,307 | 0.5% | 370,093 | 2.7% |

Source: De-identified Optum Clinformatic® Data Mart.
Enrollees with full enrollment in a commercial health plan or Medicare during the year.

Among commercial insurance enrollees, ambulatory care visit rates with hemorrhoids (all-listed diagnoses) increased with age until 75 years and were higher among women compared with men (Table 114). Among persons with known race-ethnicity, rates were highest among Blacks, followed by Hispanics, then Whites, and lowest among Asians. Rates were highest in the Northeast, followed by the South, then the West, and lowest in the Midwest.

**Table 114:** Hemorrhoids: Ambulatory care visits with first-listed and all-listed diagnoses by age, race-ethnicity, sex and region among privately insured enrollees, 2020.

| Demographic Characteristics |  | First-Listed Diagnosis |  | All-Listed Diagnoses |  |
| --- | --- | --- | --- | --- | --- |
|  |  | Number of events | Event rate per 100,000 | Number of events | Event rate per 100,000 |
| Age (years) | 0 to 11 | 210 | 19 | 622 | 56 |
|  | 12 to 24 | 2,522 | 174 | 6,006 | 414 |
|  | 25 to 44 | 20,127 | 758 | 50,760 | 1,911 |
|  | 45 to 54 | 13,584 | 935 | 62,938 | 4,334 |
|  | 55 to 64 | 14,470 | 882 | 85,964 | 5,241 |
|  | 65 to 74 | 23,232 | 818 | 153,968 | 5,420 |
|  | 75 plus | 17,528 | 680 | 91,249 | 3,542 |
| Race-ethnicity | White | 59,473 | 687 | 299,544 | 3,460 |
|  | Black | 9,000 | 724 | 46,033 | 3,705 |
|  | Hispanic | 11,336 | 759 | 54,596 | 3,656 |
|  | Asian | 4,478 | 737 | 18,241 | 3,004 |
|  | Unknown | 7,386 | 430 | 33,093 | 1,925 |
| Sex | Female | 47,481 | 660 | 242,347 | 3,366 |
|  | Male | 44,192 | 678 | 209,160 | 3,208 |
| Region | Northeast | 11,352 | 744 | 59,390 | 3,891 |
|  | Midwest | 17,992 | 541 | 93,257 | 2,807 |
|  | South | 41,273 | 734 | 202,711 | 3,607 |
|  | West | 21,056 | 648 | 96,149 | 2,957 |
| Total |  | 91,673 | 668 | 451,507 | 3,291 |

Source: De-identified Optum Clinformatic® Data Mart.
Enrollees with full enrollment in a commercial health plan or Medicare during the year.

Among commercial insurance enrollees, emergency department visit rates with hemorrhoids (all-listed diagnoses) peaked among persons 55 to 64 years and were higher among women compared with men (Table 115). Among persons with known race-ethnicity, rates were highest among Blacks, followed by Whites, then Hispanics, and lowest among Asians. Rates were highest in the Northeast, followed by the South, then the Midwest, and lowest in the West.

**Table 115:** Hemorrhoids: Emergency department visits with first-listed and all-listed diagnoses by age, race-ethnicity, sex and region among privately insured enrollees, 2020.

| Demographic Characteristics |  | First-Listed Diagnosis |  | All-Listed Diagnoses |  |
| --- | --- | --- | --- | --- | --- |
|  |  | Number of events | Event rate per 100,000 | Number of events | Event rate per 100,000 |
| Age (years) | 0 to 11 | . | . | . | . |
|  | 12 to 24 | 29 | 2 | 110 | 8 |
|  | 25 to 44 | 345 | 13 | 1,135 | 43 |
|  | 45 to 54 | 231 | 16 | 1,098 | 76 |
|  | 55 to 64 | 346 | 21 | 1,600 | 98 |
|  | 65 to 74 | 665 | 23 | 1,470 | 52 |
|  | 75 plus | 925 | 36 | 1,990 | 77 |
| Race-ethnicity | White | 1,551 | 18 | 4,879 | 56 |
|  | Black | 399 | 32 | 946 | 76 |
|  | Hispanic | 305 | 20 | 778 | 52 |
|  | Asian | 66 | 11 | 232 | 38 |
|  | Unknown | 220 | 13 | 568 | 33 |
| Sex | Female | 1,459 | 20 | 4,222 | 59 |
|  | Male | 1,082 | 17 | 3,181 | 49 |
| Region | Northeast | 362 | 24 | 1,030 | 67 |
|  | Midwest | 513 | 15 | 1,602 | 48 |
|  | South | 1,216 | 22 | 3,483 | 62 |
|  | West | 450 | 14 | 1,288 | 40 |
| Total |  | 2,541 | 19 | 7,403 | 54 |

Source: De-identified Optum Clinformatic® Data Mart.
Enrollees with full enrollment in a commercial health plan or Medicare during the year.

Among commercial insurance enrollees, hospital discharge rates with hemorrhoids (all-listed diagnoses) increased with age and were higher among women compared with men (Table 116). Among persons with known race-ethnicity, rates were highest among Blacks, followed by Whites and Hispanics, and lowest among Asians. Rates were highest in the Northeast, followed by the South, then the Midwest, and lowest in the West.

**Table 116:** Hemorrhoids: Hospital discharges with first-listed and all-listed diagnoses by age, race-ethnicity, sex and region among privately insured enrollees, 2020.

| Demographic Characteristics |  | First-Listed Diagnosis |  | All-Listed Diagnoses |  |
| --- | --- | --- | --- | --- | --- |
|  |  | Number of events | Event rate per 100,000 | Number of events | Event rate per 100,000 |
| Age (years) | 0 to 11 | . | . | 18 | 2 |
|  | 12 to 24 | 2 | 0 | 82 | 6 |
|  | 25 to 44 | 45 | 2 | 648 | 24 |
|  | 45 to 54 | 65 | 4 | 909 | 63 |
|  | 55 to 64 | 91 | 6 | 2,225 | 136 |
|  | 65 to 74 | 264 | 9 | 6,183 | 218 |
|  | 75 plus | 516 | 20 | 10,050 | 390 |
| Race-ethnicity | White | 600 | 7 | 13,006 | 150 |
|  | Black | 154 | 12 | 2,988 | 240 |
|  | Hispanic | 142 | 10 | 2,209 | 148 |
|  | Asian | 28 | 5 | 454 | 75 |
|  | Unknown | 59 | 3 | 1,458 | 85 |
| Sex | Female | 507 | 7 | 11,142 | 155 |
|  | Male | 476 | 7 | 8,973 | 138 |
| Region | Northeast | 156 | 10 | 2,860 | 187 |
|  | Midwest | 189 | 6 | 4,323 | 130 |
|  | South | 427 | 8 | 9,160 | 163 |
|  | West | 211 | 6 | 3,772 | 116 |
| Total |  | 983 | 7 | 20,115 | 147 |

Source: De-identified Optum Clinformatic® Data Mart.
Enrollees with full enrollment in a commercial health plan or Medicare during the year.

Among Medicare beneficiaries, the claims-based prevalence of hemorrhoids (based on all-listed diagnoses) was 4.2% (Table 117). Prevalence peaked among persons 70-74 years and was higher among women and Blacks. It was highest in the Northeast, followed by the South, then the West, and lowest in the Midwest.

**Table 117:** Hemorrhoids: Claims-based prevalence with first-listed and all-listed diagnoses by age, race, sex and region among fee-for-service, age-eligible Medicare beneficiaries, 2019.

| Demographic Characteristics |  | Number of enrollees | First-Listed Diagnosis |  | All-Listed Diagnoses |  |
| --- | --- | --- | --- | --- | --- | --- |
|  |  |  | Number of enrollees with disease | Percent of enrollees with disease | Number of enrollees with disease | Percent of enrollees with disease |
| Age (years) | 65 to 69 | 8,066,320 | 60,360 | 0.7% | 371,360 | 4.6% |
|  | 70 to 74 | 6,936,940 | 54,280 | 0.8% | 328,060 | 4.7% |
|  | 75 to 79 | 4,894,060 | 39,600 | 0.8% | 214,980 | 4.4% |
|  | 80 to 84 | 3,335,340 | 27,400 | 0.8% | 116,980 | 3.5% |
|  | 85+ | 3,578,120 | 27,880 | 0.8% | 94,840 | 2.7% |
| Race | White | 22,023,800 | 168,740 | 0.8% | 912,080 | 4.1% |
|  | Black | 1,861,660 | 14,040 | 0.8% | 80,880 | 4.3% |
|  | Other | 2,925,320 | 26,740 | 0.9% | 133,260 | 4.6% |
| Sex | Female | 14,975,560 | 123,040 | 0.8% | 637,020 | 4.3% |
|  | Male | 11,835,220 | 86,480 | 0.7% | 489,200 | 4.1% |
| Region | Northeast | 4,748,700 | 40,860 | 0.9% | 227,920 | 4.8% |
|  | Midwest | 6,069,800 | 38,780 | 0.6% | 225,160 | 3.7% |
|  | South | 10,595,900 | 83,420 | 0.8% | 460,680 | 4.3% |
|  | West | 5,396,380 | 46,460 | 0.9% | 212,460 | 3.9% |
| Total |  | 26,810,780 | 209,520 | 0.8% | 1,126,220 | 4.2% |

Source: Centers for Medicare & Medicaid Services, 5% Denominator and Claims Files.
Beneficiaries aged 65+ years with continuous and full Parts A and B enrollment and no Health Maintenance Organization enrollment during the year. Unweighted counts were multiplied by 20 to produce counts for this table.

Among Medicare beneficiaries, ambulatory care visit rates with hemorrhoids (all-listed diagnoses) peaked among persons 70 to 74 years, were higher among women compared with men, and did not differ by race (Table 118). Rates were highest in the Northeast, followed by the South, then the West, and lowest in the Midwest.

**Table 118:** Hemorrhoids: Ambulatory care visits with first-listed and all-listed diagnoses by age, race, sex and region among fee-for-service, age-eligible Medicare beneficiaries, 2019.

| Demographic Characteristics |  | First-Listed Diagnosis |  | All-Listed Diagnoses |  |
| --- | --- | --- | --- | --- | --- |
|  |  | Number of events | Event rate per 100,000 | Number of events | Event rate per 100,000 |
| Age (years) | 65 to 69 | 84,720 | 1,050 | 452,640 | 5,611 |
|  | 70 to 74 | 75,080 | 1,082 | 397,100 | 5,724 |
|  | 75 to 79 | 52,700 | 1,077 | 254,780 | 5,206 |
|  | 80 to 84 | 36,140 | 1,084 | 140,780 | 4,221 |
|  | 85+ | 31,780 | 888 | 110,980 | 3,102 |
| Race | White | 226,420 | 1,028 | 1,090,980 | 4,954 |
|  | Black | 16,600 | 892 | 92,220 | 4,954 |
|  | Other | 37,400 | 1,278 | 173,080 | 5,917 |
| Sex | Female | 160,360 | 1,071 | 772,560 | 5,159 |
|  | Male | 120,060 | 1,014 | 583,720 | 4,932 |
| Region | Northeast | 53,140 | 1,119 | 268,380 | 5,652 |
|  | Midwest | 50,780 | 837 | 254,440 | 4,192 |
|  | South | 114,000 | 1,076 | 566,500 | 5,346 |
|  | West | 62,500 | 1,158 | 266,960 | 4,947 |
| Total |  | 280,420 | 1,046 | 1,356,280 | 5,059 |

Source: Centers for Medicare & Medicaid Services, 5% Denominator and Claims Files. Beneficiaries aged 65+ years with continuous and full Parts A and B enrollment and no Health Maintenance Organization enrollment during the year. Unweighted counts were multiplied by 20 to produce counts for this table.

Among Medicare beneficiaries, emergency department visit rates with hemorrhoids (all-listed diagnoses) increased with age and were higher among women compared with men and Blacks compared with Whites (Table 119). Rates were highest in the Northeast, followed by the South, then the Midwest, and lowest in the West.

**Table 119:**
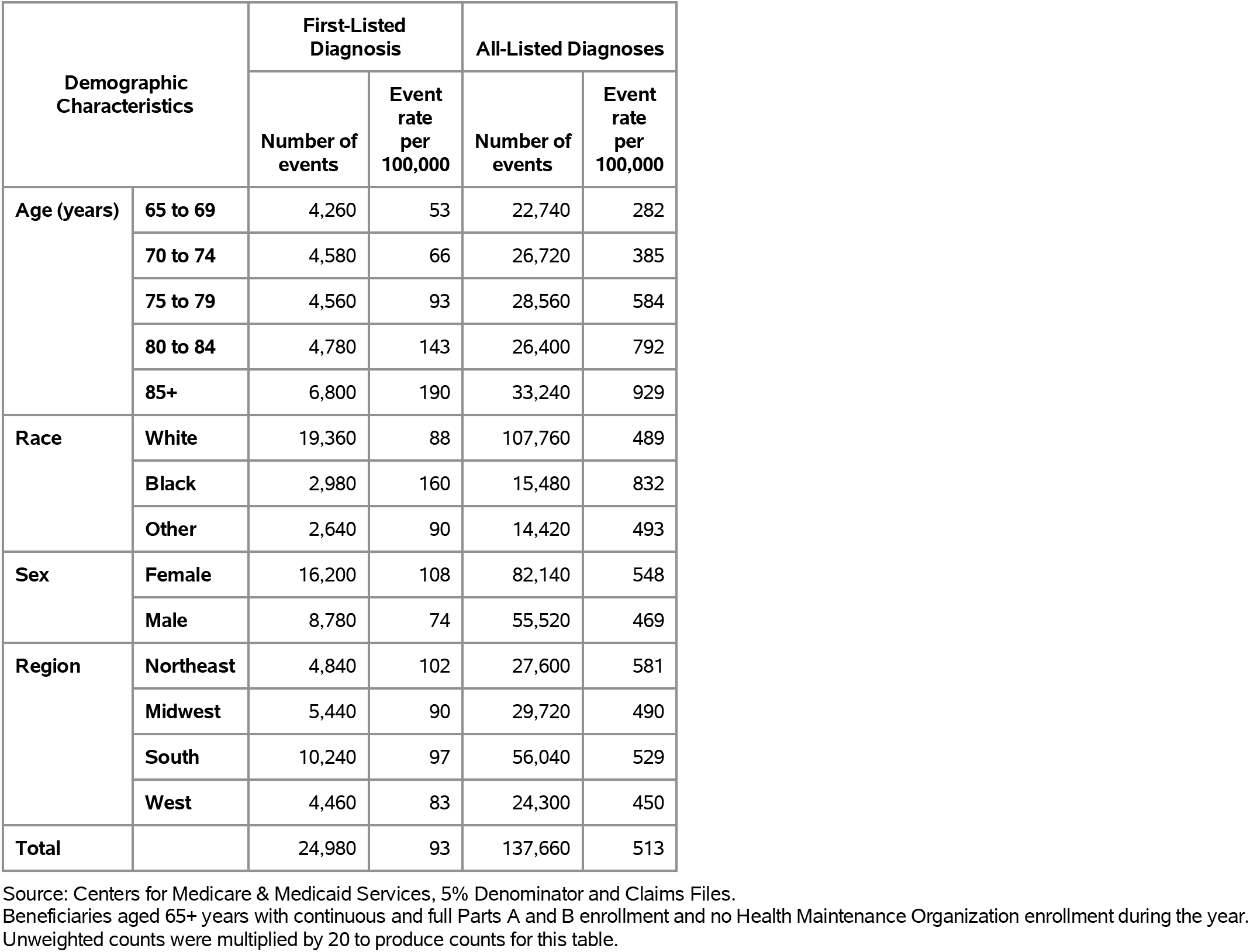
Hemorrhoids: Emergency department visits with first-listed and all-listed diagnoses by age, race, sex and region among fee-for-service, age-eligible Medicare beneficiaries, 2019.

| Demographic Characteristics |  | First-Listed Diagnosis |  | All-Listed Diagnoses |  |
| --- | --- | --- | --- | --- | --- |
|  |  | Number of events | Event rate per 100,000 | Number of events | Event rate per 100,000 |
| Age (years) | 65 to 69 | 4,260 | 53 | 22,740 | 282 |
|  | 70 to 74 | 4,580 | 66 | 26,720 | 385 |
|  | 75 to 79 | 4,560 | 93 | 28,560 | 584 |
|  | 80 to 84 | 4,780 | 143 | 26,400 | 792 |
|  | 85+ | 6,800 | 190 | 33,240 | 929 |
| Race | White | 19,360 | 88 | 107,760 | 489 |
|  | Black | 2,980 | 160 | 15,480 | 832 |
|  | Other | 2,640 | 90 | 14,420 | 493 |
| Sex | Female | 16,200 | 108 | 82,140 | 548 |
|  | Male | 8,780 | 74 | 55,520 | 469 |
| Region | Northeast | 4,840 | 102 | 27,600 | 581 |
|  | Midwest | 5,440 | 90 | 29,720 | 490 |
|  | South | 10,240 | 97 | 56,040 | 529 |
|  | West | 4,460 | 83 | 24,300 | 450 |
| Total |  | 24,980 | 93 | 137,660 | 513 |

Source: Centers for Medicare & Medicaid Services, 5% Denominator and Claims Files. Beneficiaries aged 65+ years with continuous and full Parts A and B enrollment and no Health Maintenance Organization enrollment during the year. Unweighted counts were multiplied by 20 to produce counts for this table.

Among Medicare beneficiaries, hospital discharge rates with hemorrhoids (all-listed diagnoses) increased with age and were higher among women compared with men and Blacks compared with Whites (Table 120). Rates were highest in the Northeast, followed by the South, then the Midwest, and lowest in the West.

**Table 120:** Hemorrhoids: Hospital discharges with first-listed and all-listed diagnoses by age, race, sex and region among fee-for-service, age-eligible Medicare beneficiaries, 2019.

| Demographic Characteristics |  | First-Listed Diagnosis |  | All-Listed Diagnoses |  |
| --- | --- | --- | --- | --- | --- |
|  |  | Number of events | Event rate per 100,000 | Number of events | Event rate per 100,000 |
| Age (years) | 65 to 69 | 1,200 | 15 | 18,860 | 234 |
|  | 70 to 74 | 1,400 | 20 | 22,600 | 326 |
|  | 75 to 79 | 1,500 | 31 | 25,040 | 512 |
|  | 80 to 84 | 1,820 | 55 | 22,800 | 684 |
|  | 85+ | 2,220 | 62 | 26,520 | 741 |
| Race | White | 6,400 | 29 | 91,580 | 416 |
|  | Black | 1,040 | 56 | 12,680 | 681 |
|  | Other | 700 | 24 | 11,560 | 395 |
| Sex | Female | 5,140 | 34 | 67,460 | 450 |
|  | Male | 3,000 | 25 | 48,360 | 409 |
| Region | Northeast | 1,840 | 39 | 22,880 | 482 |
|  | Midwest | 1,780 | 29 | 25,580 | 421 |
|  | South | 3,420 | 32 | 48,020 | 453 |
|  | West | 1,100 | 20 | 19,340 | 358 |
| Total |  | 8,140 | 30 | 115,820 | 432 |

Source: Centers for Medicare & Medicaid Services, 5% Denominator and Claims Files. Beneficiaries aged 65+ years with continuous and full Parts A and B enrollment and no Health Maintenance Organization enrollment during the year. Unweighted counts were multiplied by 20 to produce counts for this table.

Liver disease (acute and chronic) contributed to 4.2 million ambulatory visits (2015) (Table 121). Ambulatory care visit rates (all-listed diagnoses) were highest among persons 55-64 years. Age-adjusted ambulatory care visit rates were higher among women compared with men, Blacks compared with Whites, and non-Hispanics compared with Hispanics.

**Table 121:** Liver Disease: Ambulatory care visits with first-listed and all-listed diagnoses by age, race, ethnicity, and sex in the United States, 2015.

| Demographic Characteristics |  | First-Listed Diagnosis |  | All-Listed Diagnoses |  |
| --- | --- | --- | --- | --- | --- |
|  |  | Number in thousands | Rate per 100,000 | Number in thousands | Rate per 100,000 |
| Age (years) | 0 to 11 | . | . | 6 | 12 |
|  | 12 to 24 | 19 | 34 | 88 | 157 |
|  | 25 to 44 | 212 | 251 | 729 | 862 |
|  | 45 to 54 | 356 | 826 | 1,111 | 2,578 |
|  | 55 to 64 | 214 | 523 | 1,174 | 2,878 |
|  | 65 to 74 | 145 | 527 | 501 | 1,819 |
|  | 75 plus | 195 | 963 | 558 | 2,759 |
| Race | White | 875 | 332 | 3,145 | 1,112 |
|  | Black | 203 | 466 | 670 | 1,602 |
|  | Other | 62 | 238 | 351 | 1,440 |
| Ethnicity | Hispanic | 78 | 172 | 530 | 1,149 |
|  | Not Hispanic | 1,062 | 366 | 3,636 | 1,195 |
| Sex | Female | 627 | 370 | 2,441 | 1,362 |
|  | Male | 513 | 312 | 1,725 | 1,023 |
| Total |  | 1,140 | 340 | 4,166 | 1,196 |

Source: National Ambulatory Medical Care Survey (3-year average, 2014-2016). Rates for age groups are unadjusted and all other rates are age-adjusted.

Liver disease (acute and chronic) contributed to 2.2 million emergency department visits in 2018 (Table 122). Emergency department visit rates (all-listed diagnoses) were highest among persons 55-64 years. Age-adjusted emergency department visit rates were higher among men compared with women.

**Table 122:** Liver Disease: Emergency department visits with first-listed and all-listed diagnoses by age and sex in the United States, 2018.

| Demographic Characteristics |  | First-Listed Diagnosis |  | All-Listed Diagnoses |  |
| --- | --- | --- | --- | --- | --- |
|  |  | Number in thousands | Rate per 100,000 | Number in thousands | Rate per 100,000 |
| Age (years) | 0 to 11 | 1 | 2 | 7 | 14 |
|  | 12 to 24 | 6 | 11 | 53 | 96 |
|  | 25 to 44 | 58 | 66 | 428 | 492 |
|  | 45 to 54 | 76 | 182 | 453 | 1,089 |
|  | 55 to 64 | 99 | 234 | 593 | 1,404 |
|  | 65 to 74 | 55 | 181 | 398 | 1,305 |
|  | 75 plus | 26 | 117 | 265 | 1,208 |
| Sex | Female | 133 | 70 | 1,011 | 536 |
|  | Male | 187 | 105 | 1,187 | 672 |
| Total |  | 320 | 87 | 2,198 | 601 |

Source: Healthcare Cost and Utilization Project Nationwide Emergency Department Sample. Rates for age groups are unadjusted and all other rates are age-adjusted.

Liver disease (acute and chronic) contributed to 1.7 million hospital discharges in 2018 (Table 123). Hospital discharge rates (all-listed diagnoses) increased with age until the oldest age group. Age-adjusted hospital discharge rates were higher among men compared with women, Whites compared with Blacks, and Hispanics compared with non-Hispanics. Between 2004 and 2018, age-adjusted hospital discharge rates (per 100,000) with an all-listed diagnosis increased by 72% from 259 to 446.(7)

**Table 123:** Liver Disease: Hospital discharges with first-listed and all-listed diagnoses by age, race, ethnicity, and sex in the United States, 2018.

| Demographic Characteristics |  | First-Listed Diagnosis |  | All-Listed Diagnoses |  |
| --- | --- | --- | --- | --- | --- |
|  |  | Number in thousands | Rate per 100,000 | Number in thousands | Rate per 100,000 |
| Age (years) | 0 to 11 | 1 | 2 | 10 | 20 |
|  | 12 to 24 | 2 | 4 | 29 | 53 |
|  | 25 to 44 | 37 | 42 | 262 | 301 |
|  | 45 to 54 | 56 | 134 | 309 | 741 |
|  | 55 to 64 | 81 | 193 | 460 | 1,089 |
|  | 65 to 74 | 52 | 170 | 357 | 1,171 |
|  | 75 plus | 24 | 107 | 242 | 1,105 |
| Race | White | 214 | 71 | 1,370 | 455 |
|  | Black | 23 | 48 | 202 | 439 |
|  | Other | 18 | 68 | 115 | 442 |
| Ethnicity | Hispanic | 45 | 94 | 252 | 528 |
|  | Not Hispanic | 210 | 65 | 1,435 | 442 |
| Sex | Female | 105 | 53 | 742 | 381 |
|  | Male | 148 | 82 | 928 | 517 |
| Total |  | 253 | 67 | 1,670 | 446 |

Source: Healthcare Cost and Utilization Project National Inpatient Sample. Rates for age groups are unadjusted and all other rates are age-adjusted.

Liver disease (acute and chronic) contributed to 107,000 deaths in 2019 (Table 124). Mortality rates (underlying or other cause) increased with age. Age-adjusted mortality rates were higher among men, Whites, and Hispanics. Between 2004 and 2019, age-adjusted mortality rates (per 100,000) with liver disease as underlying or other cause increased by 7% from 24.9 to 26.8.(7)

**Table 124:** Liver Disease: Deaths with underlying or underlying/other cause and lifetime years of life lost by age, race, ethnicity, and sex in the United States, 2019.

| Demographic Characteristics |  | Underlying Cause |  |  | Underlying/Other Cause |  |  |
| --- | --- | --- | --- | --- | --- | --- | --- |
|  |  | Number of Deaths | Rate per 100,000 | Years of potential life lost in thousands | Number of Deaths | Rate per 100,000 | Years of potential life lost in thousands |
| Age (years) | 0 to 11 | 58 | 0.1 | 4.4 | 284 | 0.6 | 21.8 |
|  | 12 to 24 | 80 | 0.1 | 4.7 | 274 | 0.5 | 16.2 |
|  | 25 to 44 | 5,395 | 6.2 | 229.1 | 8,317 | 9.5 | 353.7 |
|  | 45 to 54 | 9,669 | 23.7 | 300.3 | 15,314 | 37.5 | 474.3 |
|  | 55 to 64 | 17,669 | 41.6 | 409.6 | 31,960 | 75.3 | 735.4 |
|  | 65 to 74 | 14,466 | 45.9 | 233.1 | 29,011 | 92.1 | 464.9 |
|  | 75 plus | 10,620 | 47.0 | 90.7 | 22,318 | 98.9 | 187.9 |
| Race | White | 50,610 | 15.8 | 1,102.0 | 91,676 | 28.1 | 1,911.8 |
|  | Black | 4,771 | 10.1 | 102.7 | 10,688 | 22.6 | 222.3 |
|  | Other | 2,576 | 9.6 | 67.1 | 5,114 | 19.3 | 120.1 |
| Ethnicity | Hispanic | 7,093 | 15.3 | 178.7 | 12,662 | 27.7 | 310.3 |
|  | Not Hispanic | 50,864 | 14.8 | 1,093.1 | 94,816 | 27.0 | 1,943.9 |
| Sex | Female | 23,008 | 11.1 | 521.6 | 41,890 | 19.8 | 914.5 |
|  | Male | 34,949 | 18.7 | 750.3 | 65,588 | 34.7 | 1,339.7 |
| Total |  | 57,957 | 14.7 | 1,271.8 | 107,478 | 26.8 | 2,254.1 |

Source: Vital Statistics of the United States.
Rates for age groups are unadjusted and all other rates are age-adjusted.
Years of potential life lost is based on U.S. life table 2006-2018 estimates of life expectancy at age of death.

Among privately insured enrollees, the claims-based prevalence of liver disease (acute and chronic) (based on all-listed diagnoses) was 1.9% (Table 125). Prevalence increased with age until the oldest age groups and did not differ by sex. It was highest among Hispanics, followed by Blacks, Whites, and Asians. It was highest in the South, followed by the Northeast, then the West, and lowest in the Midwest.

**Table 125:** Liver Disease: Claims-based prevalence with first-listed and all-listed diagnoses by age, race-ethnicity, sex and region among privately insured enrollees, 2020.

| Demographic Characteristics |  | Number of enrollees | First-Listed Diagnosis |  | All-Listed Diagnoses |  |
| --- | --- | --- | --- | --- | --- | --- |
|  |  |  | Number of enrollees with disease | Percent of enrollees with disease | Number of enrollees with disease | Percent of enrollees with disease |
| Age (years) | 0 to 11 | 1,101,978 | 123 | 0.0% | 386 | 0.0% |
|  | 12 to 24 | 1,451,892 | 852 | 0.1% | 2,888 | 0.2% |
|  | 25 to 44 | 2,656,672 | 6,458 | 0.2% | 26,241 | 1.0% |
|  | 45 to 54 | 1,452,320 | 7,930 | 0.5% | 32,265 | 2.2% |
|  | 55 to 64 | 1,640,216 | 15,046 | 0.9% | 52,772 | 3.2% |
|  | 65 to 74 | 2,840,615 | 27,402 | 1.0% | 90,425 | 3.2% |
|  | 75 plus | 2,576,241 | 14,230 | 0.6% | 55,117 | 2.1% |
| Race-ethnicity | White | 8,657,702 | 44,576 | 0.5% | 160,260 | 1.9% |
|  | Black | 1,242,477 | 6,693 | 0.5% | 25,368 | 2.0% |
|  | Hispanic | 1,493,457 | 12,177 | 0.8% | 42,511 | 2.8% |
|  | Asian | 607,284 | 2,683 | 0.4% | 10,887 | 1.8% |
|  | Unknown | 1,719,014 | 5,912 | 0.3% | 21,068 | 1.2% |
| Sex | Female | 7,199,363 | 39,176 | 0.5% | 137,573 | 1.9% |
|  | Male | 6,520,571 | 32,865 | 0.5% | 122,521 | 1.9% |
| Region | Northeast | 1,526,275 | 8,884 | 0.6% | 30,224 | 2.0% |
|  | Midwest | 3,322,846 | 13,224 | 0.4% | 47,748 | 1.4% |
|  | South | 5,619,435 | 34,360 | 0.6% | 125,392 | 2.2% |
|  | West | 3,251,378 | 15,573 | 0.5% | 56,730 | 1.7% |
| Total |  | 13,719,934 | 72,041 | 0.5% | 260,094 | 1.9% |

Source: De-identified Optum Clinformatic® Data Mart.
Enrollees with full enrollment in a commercial health plan or Medicare during the year.

Among commercial insurance enrollees, ambulatory care visit rates with liver disease (acute and chronic) (all-listed diagnoses) peaked among persons 55 to 64 years and were higher among women compared with men (Table 126). Among persons with known race-ethnicity, rates were much higher among Hispanics, followed by Blacks and Whites, and lowest among Asians. Rates were highest in the South, followed by the Northeast, then the West, and lowest in the Midwest.

**Table 126:** Liver Disease: Ambulatory care visits with first-listed and all-listed diagnoses by age, race-ethnicity, sex and region among privately insured enrollees, 2020.

| Demographic Characteristics |  | First-Listed Diagnosis |  | All-Listed Diagnoses |  |
| --- | --- | --- | --- | --- | --- |
|  |  | Number of events | Event rate per 100,000 | Number of events | Event rate per 100,000 |
| Age (years) | 0 to 11 | 187 | 17 | 566 | 51 |
|  | 12 to 24 | 1,236 | 85 | 4,298 | 296 |
|  | 25 to 44 | 10,065 | 379 | 42,212 | 1,589 |
|  | 45 to 54 | 14,285 | 984 | 59,571 | 4,102 |
|  | 55 to 64 | 30,013 | 1,830 | 109,754 | 6,691 |
|  | 65 to 74 | 52,917 | 1,863 | 187,580 | 6,603 |
|  | 75 plus | 26,787 | 1,040 | 100,440 | 3,899 |
| Race-ethnicity | White | 85,369 | 986 | 309,883 | 3,579 |
|  | Black | 11,445 | 921 | 44,891 | 3,613 |
|  | Hispanic | 22,988 | 1,539 | 88,304 | 5,913 |
|  | Asian | 4,117 | 678 | 19,051 | 3,137 |
|  | Unknown | 11,571 | 673 | 42,292 | 2,460 |
| Sex | Female | 71,955 | 999 | 269,807 | 3,748 |
|  | Male | 63,535 | 974 | 234,614 | 3,598 |
| Region | Northeast | 16,417 | 1,076 | 57,007 | 3,735 |
|  | Midwest | 25,280 | 761 | 88,765 | 2,671 |
|  | South | 64,350 | 1,145 | 247,770 | 4,409 |
|  | West | 29,443 | 906 | 110,879 | 3,410 |
| Total |  | 135,490 | 988 | 504,421 | 3,677 |

Source: De-identified Optum Clinformatic® Data Mart.
Enrollees with full enrollment in a commercial health plan or Medicare during the year.

Among commercial insurance enrollees, emergency department visit rates with liver disease (acute and chronic) (all-listed diagnoses) increased with age until 75 years and were higher among men compared with women (Table 127). Among persons with known race-ethnicity, rates were highest among Hispanics, followed by Blacks, then Whites, and much lower among Asians. Rates were highest in the South, followed by the Northeast, then the West, and lowest in the Midwest.

**Table 127:** Liver Disease: Emergency department visits with first-listed and all-listed diagnoses by age, race-ethnicity, sex and region among privately insured enrollees, 2020.

| Demographic Characteristics |  | First-Listed Diagnosis |  | All-Listed Diagnoses |  |
| --- | --- | --- | --- | --- | --- |
|  |  | Number of events | Event rate per 100,000 | Number of events | Event rate per 100,000 |
| Age (years) | 0 to 11 | . | . | 2 | 0 |
|  | 12 to 24 | 4 | 0 | 41 | 3 |
|  | 25 to 44 | 129 | 5 | 492 | 19 |
|  | 45 to 54 | 387 | 27 | 1,100 | 76 |
|  | 55 to 64 | 1,222 | 75 | 3,291 | 201 |
|  | 65 to 74 | 2,208 | 78 | 6,410 | 226 |
|  | 75 plus | 1,146 | 44 | 3,909 | 152 |
| Race-ethnicity | White | 3,031 | 35 | 9,169 | 106 |
|  | Black | 516 | 42 | 1,592 | 128 |
|  | Hispanic | 861 | 58 | 2,550 | 171 |
|  | Asian | 93 | 15 | 258 | 42 |
|  | Unknown | 595 | 35 | 1,676 | 97 |
| Sex | Female | 2,419 | 34 | 7,529 | 105 |
|  | Male | 2,677 | 41 | 7,716 | 118 |
| Region | Northeast | 613 | 40 | 1,698 | 111 |
|  | Midwest | 911 | 27 | 2,749 | 83 |
|  | South | 2,502 | 45 | 7,429 | 132 |
|  | West | 1,070 | 33 | 3,369 | 104 |
| Total |  | 5,096 | 37 | 15,245 | 111 |

Source: De-identified Optum Clinformatic ® Data Mart.
Enrollees with full enrollment in a commercial health plan or Medicare during the year.

Among commercial insurance enrollees, hospital discharge rates with liver disease (acute and chronic) (all-listed diagnoses) increased with age until 75 years were higher among men compared with women (Table 128). Among persons with known race-ethnicity, rates were highest among Blacks, followed by Hispanics, then Whites, and much lower among Asians. Rates were highest in the South, followed by the Northeast, then the Midwest, and lowest in the West.

**Table 128:**
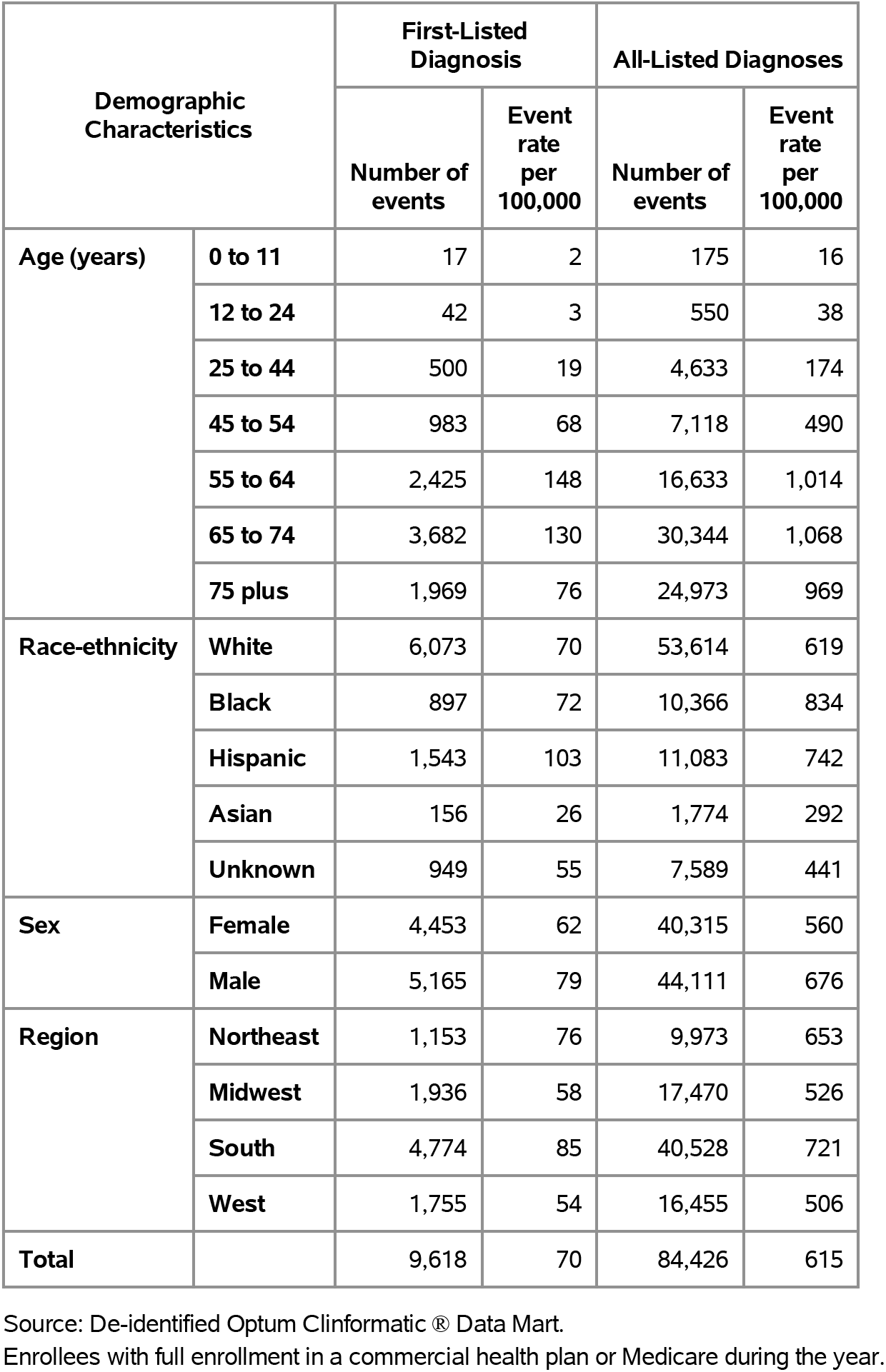
Liver Disease: Hospital discharges with first-listed and all-listed diagnoses by age, race-ethnicity, sex and region among privately insured enrollees, 2020.

| Demographic Characteristics |  | First-Listed Diagnosis |  | All-Listed Diagnoses |  |
| --- | --- | --- | --- | --- | --- |
|  |  | Number of events | Event rate per 100,000 | Number of events | Event rate per 100,000 |
| Age (years) | 0 to 11 | 17 | 2 | 175 | 16 |
|  | 12 to 24 | 42 | 3 | 550 | 38 |
|  | 25 to 44 | 500 | 19 | 4,633 | 174 |
|  | 45 to 54 | 983 | 68 | 7,118 | 490 |
|  | 55 to 64 | 2,425 | 148 | 16,633 | 1,014 |
|  | 65 to 74 | 3,682 | 130 | 30,344 | 1,068 |
|  | 75 plus | 1,969 | 76 | 24,973 | 969 |
| Race-ethnicity | White | 6,073 | 70 | 53,614 | 619 |
|  | Black | 897 | 72 | 10,366 | 834 |
|  | Hispanic | 1,543 | 103 | 11,083 | 742 |
|  | Asian | 156 | 26 | 1,774 | 292 |
|  | Unknown | 949 | 55 | 7,589 | 441 |
| Sex | Female | 4,453 | 62 | 40,315 | 560 |
|  | Male | 5,165 | 79 | 44,111 | 676 |
| Region | Northeast | 1,153 | 76 | 9,973 | 653 |
|  | Midwest | 1,936 | 58 | 17,470 | 526 |
|  | South | 4,774 | 85 | 40,528 | 721 |
|  | West | 1,755 | 54 | 16,455 | 506 |
| Total |  | 9,618 | 70 | 84,426 | 615 |

Source: De-identified Optum Clinformatic® Data Mart.
Enrollees with full enrollment in a commercial health plan or Medicare during the year.

Among Medicare beneficiaries, the claims-based prevalence of liver disease (acute and chronic) (based on all-listed diagnoses) was 3.6% (Table 129). Prevalence decreased with age and did not differ by sex or race. It was highest in the Northeast and South, followed by the West, and lowest in the Midwest.

**Table 129:** Liver Disease: Claims-based prevalence with first-listed and all-listed diagnoses by age, race, sex and region among fee-for-service, age-eligible Medicare beneficiaries, 2019.

| Demographic Characteristics |  | Number of enrollees | First-Listed Diagnosis |  | All-Listed Diagnoses |  |
| --- | --- | --- | --- | --- | --- | --- |
|  |  |  | Number of enrollees with disease | Percent of enrollees with disease | Number of enrollees with disease | Percent of enrollees with disease |
| Age (years) | 65 to 69 | 8,066,320 | 114,920 | 1.4% | 336,660 | 4.2% |
|  | 70 to 74 | 6,936,940 | 89,120 | 1.3% | 272,700 | 3.9% |
|  | 75 to 79 | 4,894,060 | 52,720 | 1.1% | 173,560 | 3.5% |
|  | 80 to 84 | 3,335,340 | 28,440 | 0.9% | 101,500 | 3.0% |
|  | 85+ | 3,578,120 | 18,160 | 0.5% | 78,280 | 2.2% |
| Race | White | 22,023,800 | 237,340 | 1.1% | 761,080 | 3.5% |
|  | Black | 1,861,660 | 19,540 | 1.0% | 65,800 | 3.5% |
|  | Other | 2,925,320 | 46,480 | 1.6% | 135,820 | 4.6% |
| Sex | Female | 14,975,560 | 172,140 | 1.1% | 534,800 | 3.6% |
|  | Male | 11,835,220 | 131,220 | 1.1% | 427,900 | 3.6% |
| Region | Northeast | 4,748,700 | 58,360 | 1.2% | 179,960 | 3.8% |
|  | Midwest | 6,069,800 | 56,460 | 0.9% | 183,960 | 3.0% |
|  | South | 10,595,900 | 125,600 | 1.2% | 402,560 | 3.8% |
|  | West | 5,396,380 | 62,940 | 1.2% | 196,220 | 3.6% |
| Total |  | 26,810,780 | 303,360 | 1.1% | 962,700 | 3.6% |

Source: Centers for Medicare & Medicaid Services, 5% Denominator and Claims Files. Beneficiaries aged 65+ years with continuous and full Parts A and B enrollment and no Health Maintenance Organization enrollment during the year. Unweighted counts were multiplied by 20 to produce counts for this table.

Among Medicare beneficiaries, ambulatory care visit rates with liver disease (acute and chronic) (all-listed diagnoses) decreased with age, differed little by sex, and were higher among Whites compared with Blacks (Table 130). Rates were lower in the Midwest compared with the Northeast, South, and West.

**Table 130:** Liver Disease: Ambulatory care visits with first-listed and all-listed diagnoses by age, race, sex and region among fee-for-service, age-eligible Medicare beneficiaries, 2019.

| Demographic Characteristics |  | First-Listed Diagnosis |  | All-Listed Diagnoses |  |
| --- | --- | --- | --- | --- | --- |
|  |  | Number of events | Event rate per 100,000 | Number of events | Event rate per 100,000 |
| Age (years) | 65 to 69 | 221,020 | 2,740 | 710,440 | 8,807 |
|  | 70 to 74 | 165,860 | 2,391 | 567,200 | 8,177 |
|  | 75 to 79 | 93,380 | 1,908 | 324,020 | 6,621 |
|  | 80 to 84 | 52,540 | 1,575 | 177,080 | 5,309 |
|  | 85+ | 28,500 | 797 | 105,940 | 2,961 |
| Race | White | 439,940 | 1,998 | 1,470,640 | 6,678 |
|  | Black | 30,620 | 1,645 | 108,720 | 5,840 |
|  | Other | 90,740 | 3,102 | 305,320 | 10,437 |
| Sex | Female | 314,120 | 2,098 | 1,050,940 | 7,018 |
|  | Male | 247,180 | 2,089 | 833,740 | 7,045 |
| Region | Northeast | 105,260 | 2,217 | 354,740 | 7,470 |
|  | Midwest | 109,520 | 1,804 | 354,700 | 5,844 |
|  | South | 229,600 | 2,167 | 786,040 | 7,418 |
|  | West | 116,920 | 2,167 | 389,200 | 7,212 |
| Total |  | 561,300 | 2,094 | 1,884,680 | 7,030 |

Source: Centers for Medicare & Medicaid Services, 5% Denominator and Claims Files. Beneficiaries aged 65+ years with continuous and full Parts A and B enrollment and no Health Maintenance Organization enrollment during the year. Unweighted counts were multiplied by 20 to produce counts for this table.

Among Medicare beneficiaries, emergency department visit rates with liver disease (acute and chronic) (all-listed diagnoses) were lowest among persons 85 years and over and were higher among men compared with women and Blacks compared with Whites (Table 131). Rates were highest in the South, followed by the West, then the Northeast, and lowest in the Midwest.

**Table 131:** Liver Disease: Emergency department visits with first-listed and all-listed diagnoses by age, race, sex and region among fee-for-service, age-eligible Medicare beneficiaries, 2019.

| Demographic Characteristics |  | First-Listed Diagnosis |  | All-Listed Diagnoses |  |
| --- | --- | --- | --- | --- | --- |
|  |  | Number of events | Event rate per 100,000 | Number of events | Event rate per 100,000 |
| Age (years) | 65 to 69 | 25,540 | 317 | 135,220 | 1,676 |
|  | 70 to 74 | 16,360 | 236 | 109,040 | 1,572 |
|  | 75 to 79 | 11,300 | 231 | 78,760 | 1,609 |
|  | 80 to 84 | 6,900 | 207 | 54,220 | 1,626 |
|  | 85+ | 5,340 | 149 | 50,800 | 1,420 |
| Race | White | 48,600 | 221 | 328,820 | 1,493 |
|  | Black | 4,380 | 235 | 37,980 | 2,040 |
|  | Other | 12,460 | 426 | 61,240 | 2,093 |
| Sex | Female | 32,980 | 220 | 221,960 | 1,482 |
|  | Male | 32,460 | 274 | 206,080 | 1,741 |
| Region | Northeast | 11,160 | 235 | 73,620 | 1,550 |
|  | Midwest | 11,960 | 197 | 83,540 | 1,376 |
|  | South | 27,540 | 260 | 183,360 | 1,730 |
|  | West | 14,780 | 274 | 87,520 | 1,622 |
| Total |  | 65,440 | 244 | 428,040 | 1,597 |

Source: Centers for Medicare & Medicaid Services, 5% Denominator and Claims Files. Beneficiaries aged 65+ years with continuous and full Parts A and B enrollment and no Health Maintenance Organization enrollment during the year. Unweighted counts were multiplied by 20 to produce counts for this table.

Among Medicare beneficiaries, hospital discharge rates with liver disease (acute and chronic) (all-listed diagnoses) were lowest among persons 85 years and over and were higher among men compared with women and Blacks compared with Whites (Table 132). Rates were higher in the South compared with the Northeast, Midwest, and West.

**Table 132:** Liver Disease: Hospital discharges with first-listed and all-listed diagnoses by age, race, sex and region among fee-for-service, age-eligible Medicare beneficiaries, 2019.

| Demographic Characteristics |  | First-Listed Diagnosis |  | All-Listed Diagnoses |  |
| --- | --- | --- | --- | --- | --- |
|  |  | Number of events | Event rate per 100,000 | Number of events | Event rate per 100,000 |
| Age (years) | 65 to 69 | 17,200 | 213 | 111,100 | 1,377 |
|  | 70 to 74 | 10,600 | 153 | 88,960 | 1,282 |
|  | 75 to 79 | 7,040 | 144 | 64,720 | 1,322 |
|  | 80 to 84 | 4,100 | 123 | 44,620 | 1,338 |
|  | 85+ | 2,840 | 79 | 39,600 | 1,107 |
| Race | White | 31,240 | 142 | 269,620 | 1,224 |
|  | Black | 2,300 | 124 | 30,060 | 1,615 |
|  | Other | 8,240 | 282 | 49,320 | 1,686 |
| Sex | Female | 20,220 | 135 | 172,760 | 1,154 |
|  | Male | 21,560 | 182 | 176,240 | 1,489 |
| Region | Northeast | 6,300 | 133 | 58,420 | 1,230 |
|  | Midwest | 9,280 | 153 | 75,620 | 1,246 |
|  | South | 17,400 | 164 | 145,820 | 1,376 |
|  | West | 8,800 | 163 | 69,140 | 1,281 |
| Total |  | 41,780 | 156 | 349,000 | 1,302 |

Source: Centers for Medicare & Medicaid Services, 5% Denominator and Claims Files. Beneficiaries aged 65+ years with continuous and full Parts A and B enrollment and no Health Maintenance Organization enrollment during the year. Unweighted counts were multiplied by 20 to produce counts for this table.

Hepatitis A contributed to 12,000 ambulatory visits (2015) (Table 133). Ambulatory care visits were uncommon, and rates (all-listed diagnoses) were highest among persons 55-64 years. Age-adjusted ambulatory care visit rates were higher among women compared with men, Whites compared with Blacks, and Hispanics compared with non-Hispanics.

**Table 133:** Hepatitis A: Ambulatory care visits with first-listed and all-listed diagnoses by age, race, ethnicity, and sex in the United States, 2015.

| Demographic Characteristics |  | All-Listed Diagnoses |  |
| --- | --- | --- | --- |
|  |  | Number in thousands | Rate per 100,000 |
| Age (years) | 0 to 11 | . | . |
|  | 12 to 24 | . | . |
|  | 25 to 44 | . | . |
|  | 45 to 54 | . | . |
|  | 55 to 64 | 10 | 25 |
|  | 65 to 74 | . | . |
|  | 75 plus | 2 | 8 |
| Race | White | 11 | 3 |
|  | Black | 0 | 1 |
|  | Other | . | . |
| Ethnicity | Hispanic | 5 | 10 |
|  | Not Hispanic | 7 | 2 |
| Sex | Female | 10 | 4 |
|  | Male | 2 | 1 |
| Total |  | 12 | 3 |

Source: National Ambulatory Medical Care Survey (3-year average, 2014-2016). Rates for age groups are unadjusted and all other rates are age-adjusted.

Hepatitis A contributed to 19,000 emergency department visits in 2018 (Table 134). Emergency department visit rates (all-listed diagnoses) were uncommon among children and highest among persons 25-44 years. Age-adjusted emergency department visit rates were higher among men compared with women.

**Table 134:** Hepatitis A: Emergency department visits with first-listed and all-listed diagnoses by age and sex in the United States, 2018.

| Demographic Characteristics |  | First-Listed Diagnosis |  | All-Listed Diagnoses |  |
| --- | --- | --- | --- | --- | --- |
|  |  | Number in thousands | Rate per 100,000 | Number in thousands | Rate per 100,000 |
| Age (years) | 0 to 11 | 0 | 0 | 0 | 0 |
|  | 12 to 24 | 1 | 1 | 1 | 2 |
|  | 25 to 44 | 4 | 5 | 8 | 9 |
|  | 45 to 54 | 1 | 2 | 3 | 7 |
|  | 55 to 64 | 1 | 1 | 3 | 7 |
|  | 65 to 74 | 0 | 1 | 2 | 6 |
|  | 75 plus | 0 | 1 | 2 | 7 |
| Sex | Female | 3 | 2 | 8 | 5 |
|  | Male | 4 | 3 | 10 | 6 |
| Total |  | 7 | 2 | 19 | 6 |

Source: Healthcare Cost and Utilization Project Nationwide Emergency Department Sample. Rates for age groups are unadjusted and all other rates are age-adjusted.

Hepatitis A contributed to 15,000 hospital discharges in 2018 (Table 135). Hospital discharges were uncommon among children and rates (all-listed diagnoses) were highest among persons 25-44 years. Age-adjusted hospital discharge rates were higher among men compared with women, Whites compared with Blacks, and non-Hispanics compared with Hispanics. Between 2004 and 2018, age-adjusted hospital discharge rates (per 100,000) with an all-listed diagnosis increased by a third from 3 to 4.(4)

**Table 135:**
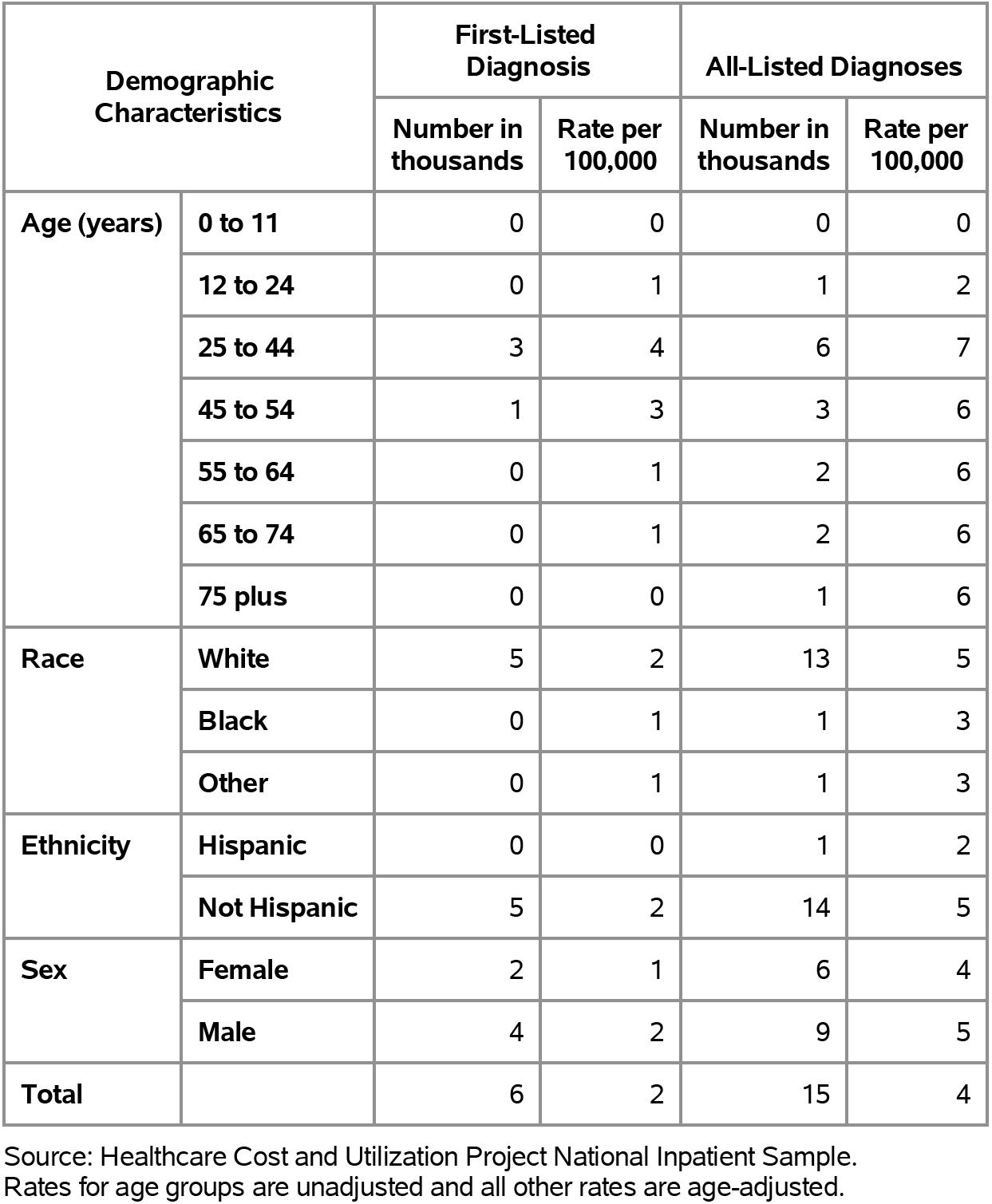
Hepatitis A: Hospital discharges with first-listed and all-listed diagnoses by age, race, ethnicity, and sex in the United States, 2018.

| Demographic Characteristics |  | First-Listed Diagnosis |  | All-Listed Diagnoses |  |
| --- | --- | --- | --- | --- | --- |
|  |  | Number in thousands | Rate per 100,000 | Number in thousands | Rate per 100,000 |
| Age (years) | 0 to 11 | 0 | 0 | 0 | 0 |
|  | 12 to 24 | 0 | 1 | 1 | 2 |
|  | 25 to 44 | 3 | 4 | 6 | 7 |
|  | 45 to 54 | 1 | 3 | 3 | 6 |
|  | 55 to 64 | 0 | 1 | 2 | 6 |
|  | 65 to 74 | 0 | 1 | 2 | 6 |
|  | 75 plus | 0 | 0 | 1 | 6 |
| Race | White | 5 | 2 | 13 | 5 |
|  | Black | 0 | 1 | 1 | 3 |
|  | Other | 0 | 1 | 1 | 3 |
| Ethnicity | Hispanic | 0 | 0 | 1 | 2 |
|  | Not Hispanic | 5 | 2 | 14 | 5 |
| Sex | Female | 2 | 1 | 6 | 4 |
|  | Male | 4 | 2 | 9 | 5 |
| Total |  | 6 | 2 | 15 | 4 |

Source: Healthcare Cost and Utilization Project National Inpatient Sample. Rates for age groups are unadjusted and all other rates are age-adjusted.

Hepatitis A contributed to <1,000 deaths in 2019 (Table 136). Mortality was uncommon among children and young adults and rates (underlying or other cause) were highest among persons 75 years and over. Age-adjusted mortality rates were higher among men, Whites, and non-Hispanics. Between 2004 and 2019, mortality with hepatitis A as underlying or other cause remained uncommon.(4)

**Table 136:** Hepatitis A: Deaths with underlying or underlying/other cause and lifetime years of life lost by age, race, ethnicity, and sex in the United States, 2019.

| Demographic Characteristics |  | Underlying Cause |  |  | Underlying/Other Cause |  |  |
| --- | --- | --- | --- | --- | --- | --- | --- |
|  |  | Number of Deaths | Rate per 100,000 | Years of potential life lost in thousands | Number of Deaths | Rate per 100,000 | Years of potential life lost in thousands |
| Age (years) | 0 to 11 | . | . | . | . | . | . |
|  | 12 to 24 | . | . | . | . | . | . |
|  | 25 to 44 | 14 | 0.0 | 0.6 | 25 | 0.0 | 1.0 |
|  | 45 to 54 | 30 | 0.1 | 0.9 | 56 | 0.1 | 1.7 |
|  | 55 to 64 | 40 | 0.1 | 0.9 | 62 | 0.1 | 1.4 |
|  | 65 to 74 | 26 | 0.1 | 0.4 | 46 | 0.1 | 0.7 |
|  | 75 plus | 29 | 0.1 | 0.2 | 37 | 0.2 | 0.3 |
| Race | White | 131 | 0.0 | 2.8 | 206 | 0.1 | 4.7 |
|  | Black | 5 | 0.0 | 0.2 | 13 | 0.0 | 0.3 |
|  | Other | 3 | 0.0 | 0.1 | 7 | 0.0 | 0.2 |
| Ethnicity | Hispanic | 4 | 0.0 | 0.1 | 11 | 0.0 | 0.3 |
|  | Not Hispanic | 135 | 0.0 | 3.0 | 215 | 0.1 | 4.9 |
| Sex | Female | 38 | 0.0 | 0.9 | 66 | 0.0 | 1.6 |
|  | Male | 101 | 0.1 | 2.2 | 160 | 0.1 | 3.6 |
| Total |  | 139 | 0.0 | 3.1 | 226 | 0.1 | 5.2 |

Source: Vital Statistics of the United States.
Rates for age groups are unadjusted and all other rates are age-adjusted.
Years of potential life lost is based on U.S. life table 2006-2018 estimates of life expectancy at age of death.

Among commercial insurance enrollees, the claims-based prevalence of hepatitis A (based on all-listed diagnoses) was <0.1% (Table 137).

**Table 137:** Hepatitis A: Claims-based prevalence with first-listed and all-listed diagnoses by age, race-ethnicity, sex and region among privately insured enrollees, 2020.

| Demographic Characteristics |  | Number of enrollees | First-Listed Diagnosis |  | All-Listed Diagnoses |  |
| --- | --- | --- | --- | --- | --- | --- |
|  |  |  | Number of enrollees with disease | Percent of enrollees with disease | Number of enrollees with disease | Percent of enrollees with disease |
| Age (years) | 0 to 11 | 1,101,978 | 3 | 0.0% | 6 | 0.0% |
|  | 12 to 24 | 1,451,892 | 18 | 0.0% | 36 | 0.0% |
|  | 25 to 44 | 2,656,672 | 57 | 0.0% | 126 | 0.0% |
|  | 45 to 54 | 1,452,320 | 33 | 0.0% | 122 | 0.0% |
|  | 55 to 64 | 1,640,216 | 73 | 0.0% | 240 | 0.0% |
|  | 65 to 74 | 2,840,615 | 80 | 0.0% | 492 | 0.0% |
|  | 75 plus | 2,576,241 | 54 | 0.0% | 391 | 0.0% |
| Race-ethnicity | White | 8,657,702 | 190 | 0.0% | 863 | 0.0% |
|  | Black | 1,242,477 | 37 | 0.0% | 156 | 0.0% |
|  | Hispanic | 1,493,457 | 41 | 0.0% | 199 | 0.0% |
|  | Asian | 607,284 | 24 | 0.0% | 72 | 0.0% |
|  | Unknown | 1,719,014 | 26 | 0.0% | 123 | 0.0% |
| Sex | Female | 7,199,363 | 161 | 0.0% | 717 | 0.0% |
|  | Male | 6,520,571 | 157 | 0.0% | 696 | 0.0% |
| Region | Northeast | 1,526,275 | 48 | 0.0% | 209 | 0.0% |
|  | Midwest | 3,322,846 | 41 | 0.0% | 203 | 0.0% |
|  | South | 5,619,435 | 177 | 0.0% | 734 | 0.0% |
|  | West | 3,251,378 | 52 | 0.0% | 267 | 0.0% |
| Total |  | 13,719,934 | 318 | 0.0% | 1,413 | 0.0% |

Source: De-identified Optum Clinformatic® Data Mart.
Enrollees with full enrollment in a commercial health plan or Medicare during the year.

Among commercial insurance enrollees, ambulatory care visit rates with hepatitis A (all-listed diagnoses) increased with age until 75 years and did not differ by sex (Table 138). Among persons with known race-ethnicity, rates were highest among Hispanics, followed by Asians, Blacks, and Whites. Rates were highest in the Northeast, followed by the South, then the West, and lowest in the Midwest.

**Table 138:** Hepatitis A: Ambulatory care visits with first-listed and all-listed diagnoses by age, race-ethnicity, sex and region among privately insured enrollees, 2020.

| Demographic Characteristics |  | First-Listed Diagnosis |  | All-Listed Diagnoses |  |
| --- | --- | --- | --- | --- | --- |
|  |  | Number of events | Event rate per 100,000 | Number of events | Event rate per 100,000 |
| Age (years) | 0 to 11 | 3 | 0 | 5 | 0 |
|  | 12 to 24 | 20 | 1 | 37 | 3 |
|  | 25 to 44 | 70 | 3 | 158 | 6 |
|  | 45 to 54 | 47 | 3 | 155 | 11 |
|  | 55 to 64 | 89 | 5 | 294 | 18 |
|  | 65 to 74 | 103 | 4 | 607 | 21 |
|  | 75 plus | 66 | 3 | 507 | 20 |
| Race-ethnicity | White | 256 | 3 | 1,111 | 13 |
|  | Black | 38 | 3 | 169 | 14 |
|  | Hispanic | 44 | 3 | 263 | 18 |
|  | Asian | 31 | 5 | 91 | 15 |
|  | Unknown | 29 | 2 | 129 | 8 |
| Sex | Female | 198 | 3 | 926 | 13 |
|  | Male | 200 | 3 | 837 | 13 |
| Region | Northeast | 67 | 4 | 310 | 20 |
|  | Midwest | 61 | 2 | 231 | 7 |
|  | South | 207 | 4 | 879 | 16 |
|  | West | 63 | 2 | 343 | 11 |
| Total |  | 398 | 3 | 1,763 | 13 |

Source: De-identified Optum Clinformatic® Data Mart.
Enrollees with full enrollment in a commercial health plan or Medicare during the year.

Among commercial insurance enrollees, the emergency department visit rate with hepatitis A (all-listed diagnoses) was less than 1 per 100,000 (Table 139).

**Table 139:** Hepatitis A: Emergency department visits with first-listed and all-listed diagnoses by age, race-ethnicity, sex and region among privately insured enrollees, 2020.

| Demographic Characteristics |  | First-Listed Diagnosis |  | All-Listed Diagnoses |  |
| --- | --- | --- | --- | --- | --- |
|  |  | Number of events | Event rate per 100,000 | Number of events | Event rate per 100,000 |
| Age (years) | 0 to 11 | . | . | . | . |
|  | 12 to 24 | . | . | . | . |
|  | 25 to 44 | 2 | 0 | 6 | 0 |
|  | 45 to 54 | . | . | 3 | 0 |
|  | 55 to 64 | 5 | 0 | 10 | 1 |
|  | 65 to 74 | 5 | 0 | 14 | 0 |
|  | 75 plus | 1 | 0 | 9 | 0 |
| Race-ethnicity | White | 8 | 0 | 29 | 0 |
|  | Black | . | . | 2 | 0 |
|  | Hispanic | 1 | 0 | 3 | 0 |
|  | Asian | . | . | 1 | 0 |
|  | Unknown | 4 | 0 | 7 | 0 |
| Sex | Female | 5 | 0 | 16 | 0 |
|  | Male | 8 | 0 | 26 | 0 |
| Region | Northeast | 1 | 0 | 7 | 0 |
|  | Midwest | 1 | 0 | 8 | 0 |
|  | South | 9 | 0 | 20 | 0 |
|  | West | 2 | 0 | 7 | 0 |
| Total |  | 13 | 0 | 42 | 0 |

Source: De-identified Optum Clinformatics® Data Mart.
Enrollees with full enrollment in a commercial health plan or Medicare during the year.

Among commercial insurance enrollees, hospital discharge rates with hepatitis A (all-listed diagnoses) increased with age and differed little by sex (Table 140). Among persons with known race-ethnicity, rates were highest among Blacks, followed by Whites and Hispanics, and lowest among Asians. Rates were highest in the South, followed by the Northeast, then the West, and lowest in the Midwest.

**Table 140:** Hepatitis A: Hospital discharges with first-listed and all-listed diagnoses by age, race-ethnicity, sex and region among privately insured enrollees, 2020.

| Demographic Characteristics |  | First-Listed Diagnosis |  | All-Listed Diagnoses |  |
| --- | --- | --- | --- | --- | --- |
|  |  | Number of events | Event rate per 100,000 | Number of events | Event rate per 100,000 |
| Age (years) | 0 to 11 | . | . | 1 | 0 |
|  | 12 to 24 | . | . | 11 | 1 |
|  | 25 to 44 | 15 | 1 | 32 | 1 |
|  | 45 to 54 | 12 | 1 | 39 | 3 |
|  | 55 to 64 | 26 | 2 | 90 | 5 |
|  | 65 to 74 | 18 | 1 | 177 | 6 |
|  | 75 plus | 11 | 0 | 145 | 6 |
| Race-ethnicity | White | 55 | 1 | 321 | 4 |
|  | Black | 13 | 1 | 59 | 5 |
|  | Hispanic | 6 | 0 | 58 | 4 |
|  | Asian | 2 | 0 | 17 | 3 |
|  | Unknown | 6 | 0 | 40 | 2 |
| Sex | Female | 37 | 1 | 235 | 3 |
|  | Male | 45 | 1 | 260 | 4 |
| Region | Northeast | 9 | 1 | 56 | 4 |
|  | Midwest | 11 | 0 | 82 | 2 |
|  | South | 48 | 1 | 260 | 5 |
|  | West | 14 | 0 | 97 | 3 |
| Total |  | 82 | 1 | 495 | 4 |

Source: De-identified Optum Clinformatic® Data Mart.
Enrollees with full enrollment in a commercial health plan or Medicare during the year.

Among Medicare beneficiaries, the claims-based prevalence of hepatitis A (based on all-listed diagnoses) was <0.1% (Table 141).

**Table 141:** Hepatitis A: Claims-based prevalence with first-listed and all-listed diagnoses by age, race, sex and region among fee-for-service, age-eligible Medicare beneficiaries, 2019.

| Demographic Characteristics |  | Number of enrollees | First-Listed Diagnosis |  | All-Listed Diagnoses |  |
| --- | --- | --- | --- | --- | --- | --- |
|  |  |  | Number of enrollees with disease | Percent of enrollees with disease | Number of enrollees with disease | Percent of enrollees with disease |
| Age (years) | 65 to 69 | 8,066,320 | 300 | 0.0% | 1,660 | 0.0% |
|  | 70 to 74 | 6,936,940 | . | . | 1,120 | 0.0% |
|  | 75 to 79 | 4,894,060 | . | . | 880 | 0.0% |
|  | 80 to 84 | 3,335,340 | . | . | 580 | 0.0% |
|  | 85+ | 3,578,120 | . | . | 400 | 0.0% |
| Race | White | 22,023,800 | 640 | 0.0% | 3,720 | 0.0% |
|  | Black | 1,861,660 | . | . | 220 | 0.0% |
|  | Other | 2,925,320 | . | . | 700 | 0.0% |
| Sex | Female | 14,975,560 | 500 | 0.0% | 2,540 | 0.0% |
|  | Male | 11,835,220 | 240 | 0.0% | 2,100 | 0.0% |
| Region | Northeast | 4,748,700 | . | . | 620 | 0.0% |
|  | Midwest | 6,069,800 | . | . | 780 | 0.0% |
|  | South | 10,595,900 | 360 | 0.0% | 2,280 | 0.0% |
|  | West | 5,396,380 | . | . | 960 | 0.0% |
| Total |  | 26,810,780 | 740 | 0.0% | 4,640 | 0.0% |

Source: Centers for Medicare & Medicaid Services, 5% Denominator and Claims Files.
Beneficiaries aged 65+ years with continuous and full Parts A and B enrollment and no Health Maintenance Organization enrollment during the year. Unweighted counts were multiplied by 20 to produce counts for this table.

Among Medicare beneficiaries, ambulatory care visits with hepatitis A were uncommon among persons over 80 years and Blacks (Table 142). Rates (all-listed diagnoses) were higher among men compared with women. Rates were much higher in the South, intermediate in the Northeast and West, and lowest in the Midwest.

**Table 142:** Hepatitis A: Ambulatory care visits with first-listed and all-listed diagnoses by age, race, sex and region among fee-for-service, age-eligible Medicare beneficiaries, 2019.

| Demographic Characteristics |  | First-Listed Diagnosis |  | All-Listed Diagnoses |  |
| --- | --- | --- | --- | --- | --- |
|  |  | Number of events | Event rate per 100,000 | Number of events | Event rate per 100,000 |
| Age (years) | 65 to 69 | 400 | 5 | 2,060 | 26 |
|  | 70 to 74 | . | . | 1,000 | 14 |
|  | 75 to 79 | . | . | 1,700 | 35 |
|  | 80 to 84 | . | . | . | . |
|  | 85+ | . | . | . | . |
| Race | White | 840 | 4 | 4,700 | 21 |
|  | Black | . | . | . | . |
|  | Other | . | . | . | . |
| Sex | Female | . | . | 2,740 | 18 |
|  | Male | . | . | 2,880 | 24 |
| Region | Northeast | . | . | 720 | 15 |
|  | Midwest | . | . | 480 | 8 |
|  | South | 520 | 5 | 3,780 | 36 |
|  | West | . | . | 640 | 12 |
| Total |  | 960 | 4 | 5,620 | 21 |

Source: Centers for Medicare & Medicaid Services, 5% Denominator and Claims Files. Beneficiaries aged 65+ years with continuous and full Parts A and B enrollment and no Health Maintenance Organization enrollment during the year. Unweighted counts were multiplied by 20 to produce counts for this table.

Among Medicare beneficiaries, emergency department visits with hepatitis A were uncommon among Blacks (Table 143). Rates (all-listed diagnoses) were highest among persons 80 to 84 years and higher among men compared with women. Rates were higher in the South compared with Northeast, West, and Midwest.

**Table 143:** Hepatitis A: Emergency department visits with first-listed and all-listed diagnoses by age, race, sex and region among fee-for-service, age-eligible Medicare beneficiaries, 2019.

| Demographic Characteristics |  | First-Listed Diagnosis |  | All-Listed Diagnoses |  |
| --- | --- | --- | --- | --- | --- |
|  |  | Number of events | Event rate per 100,000 | Number of events | Event rate per 100,000 |
| Age (years) | 65 to 69 | . | . | 600 | 7 |
|  | 70 to 74 | . | . | 480 | 7 |
|  | 75 to 79 | . | . | 400 | 8 |
|  | 80 to 84 | . | . | 320 | 10 |
|  | 85+ | . | . | 260 | 7 |
| Race | White | 280 | 1 | 1,780 | 8 |
|  | Black | . | . | . | . |
|  | Other | . | . | . | . |
| Sex | Female | . | . | 1,020 | 7 |
|  | Male | . | . | 1,040 | 9 |
| Region | Northeast | . | . | 260 | 5 |
|  | Midwest | . | . | 420 | 7 |
|  | South | . | . | 1,040 | 10 |
|  | West | . | . | 340 | 6 |
| Total |  | 280 | 1 | 2,060 | 8 |

Source: Centers for Medicare & Medicaid Services, 5% Denominator and Claims Files. Beneficiaries aged 65+ years with continuous and full Parts A and B enrollment and no Health Maintenance Organization enrollment during the year. Unweighted counts were multiplied by 20 to produce counts for this table.

Among Medicare beneficiaries, hospital discharges with hepatitis A were uncommon among persons over 80 years and Blacks (Table 144). Rates (all-listed diagnoses) were higher among men compared with women. Rates were higher in the South and West compared with the Northeast and Midwest.

**Table 144:**
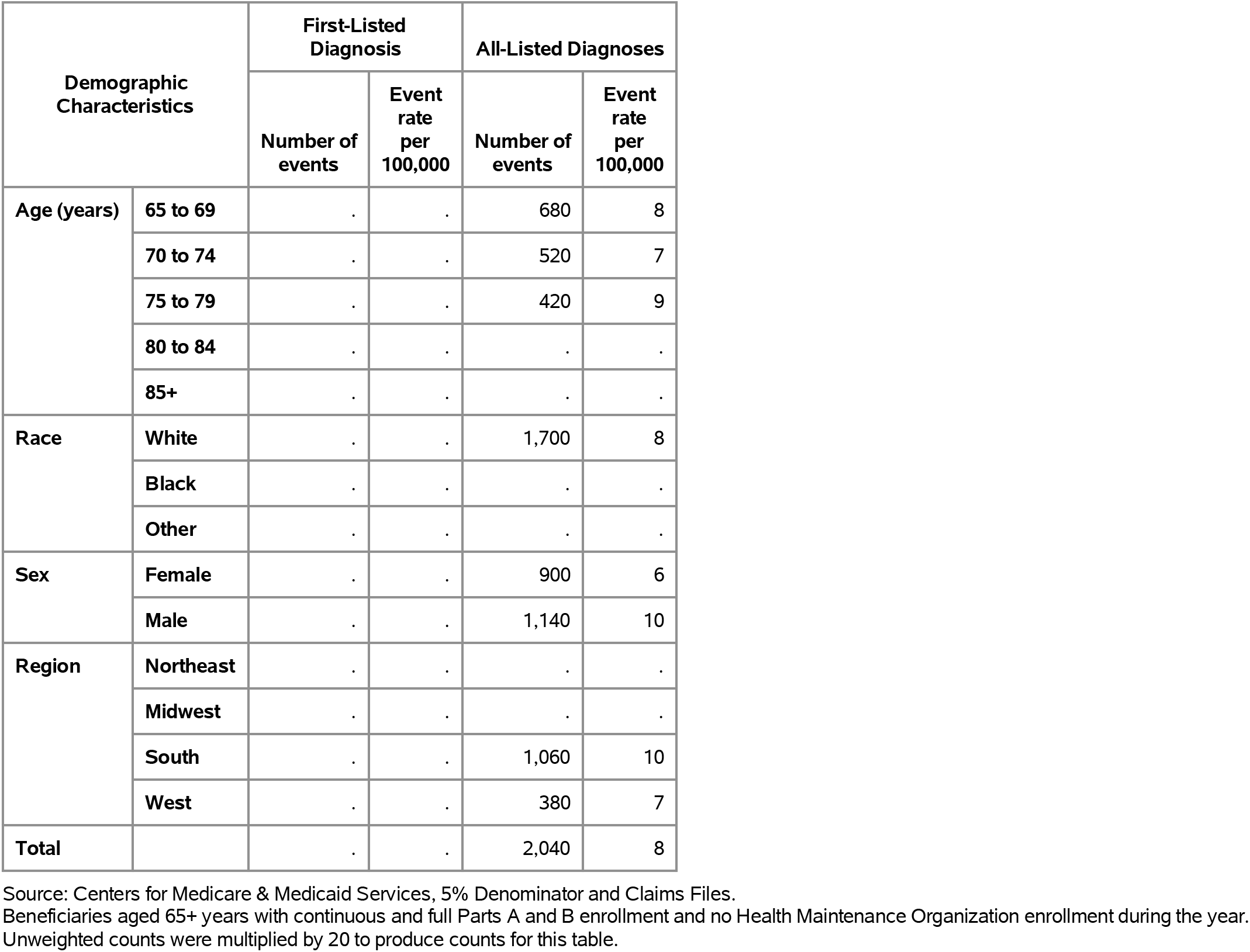
Hepatitis A: Hospital discharges with first-listed and all-listed diagnoses by age, race, sex and region among fee-for-service, age-eligible Medicare beneficiaries, 2019.

Hepatitis B contributed to 764,000 ambulatory visits (2015) (Table 145). Ambulatory care visits were uncommon among children and rates (all-listed diagnoses) were much higher among persons 75 years and over compared to younger age groups. Age-adjusted ambulatory care visit rates were higher among men compared with women, Blacks compared with Whites, and non-Hispanics compared with Hispanics.

**Table 145:** Hepatitis B: Ambulatory care visits with first-listed and all-listed diagnoses by age, race, ethnicity, and sex in the United States, 2015.

| Demographic Characteristics |  | First-Listed Diagnosis |  | All-Listed Diagnoses |  |
| --- | --- | --- | --- | --- | --- |
|  |  | Number in thousands | Rate per 100,000 | Number in thousands | Rate per 100,000 |
| Age (years) | 0 to 11 | . | . | . | . |
|  | 12 to 24 | 31 | 56 | 31 | 56 |
|  | 25 to 44 | 2 | 2 | 79 | 94 |
|  | 45 to 54 | 100 | 231 | 191 | 444 |
|  | 55 to 64 | 5 | 13 | 111 | 273 |
|  | 65 to 74 | 2 | 9 | 62 | 224 |
|  | 75 plus | 0 | 1 | 289 | 1,431 |
| Race | White | 6 | 2 | 148 | 45 |
|  | Black | 46 | 98 | 64 | 131 |
|  | Other | 88 | 350 | 553 | 2,863 |
| Ethnicity | Hispanic | . | . | 17 | 34 |
|  | Not Hispanic | 141 | 49 | 747 | 255 |
| Sex | Female | 4 | 2 | 196 | 123 |
|  | Male | 136 | 81 | 568 | 362 |
| Total |  | 141 | 41 | 764 | 228 |

Source: National Ambulatory Medical Care Survey (3-year average, 2014-2016). Rates for age groups are unadjusted and all other rates are age-adjusted.

Hepatitis B contributed to 89,000 emergency department visits in 2018 (Table 146). Emergency department visits were uncommon among children and rates (all-listed diagnoses) peaked at 55-64 years. Age-adjusted rates were higher among men compared with women.

**Table 146:** Hepatitis B: Emergency department visits with first-listed and all-listed diagnoses by age and sex in the United States, 2018.

| Demographic Characteristics |  | First-Listed Diagnosis |  | All-Listed Diagnoses |  |
| --- | --- | --- | --- | --- | --- |
|  |  | Number in thousands | Rate per 100,000 | Number in thousands | Rate per 100,000 |
| Age (years) | 0 to 11 | 0 | 0 | 0 | 0 |
|  | 12 to 24 | 0 | 0 | 1 | 3 |
|  | 25 to 44 | 1 | 2 | 20 | 23 |
|  | 45 to 54 | 1 | 2 | 20 | 48 |
|  | 55 to 64 | 0 | 1 | 25 | 59 |
|  | 65 to 74 | 0 | 1 | 14 | 47 |
|  | 75 plus | 0 | 0 | 8 | 35 |
| Sex | Female | 1 | 1 | 34 | 19 |
|  | Male | 2 | 1 | 55 | 31 |
| Total |  | 3 | 1 | 89 | 25 |

Source: Healthcare Cost and Utilization Project Nationwide Emergency Department Sample. Rates for age groups are unadjusted and all other rates are age-adjusted.

Hepatitis B contributed to 82,000 hospital discharges in 2018 (Table 147). Hospital discharges were uncommon among children and rates (all-listed diagnoses) peaked at 55-64 years. Age-adjusted rates were higher among men compared with women, Blacks compared with Whites, and non-Hispanics compared with Hispanics. Between 2004 and 2018, age-adjusted hospital discharge rates (per 100,000) with an all-listed diagnosis remained stable at 23.(4)

**Table 147:**
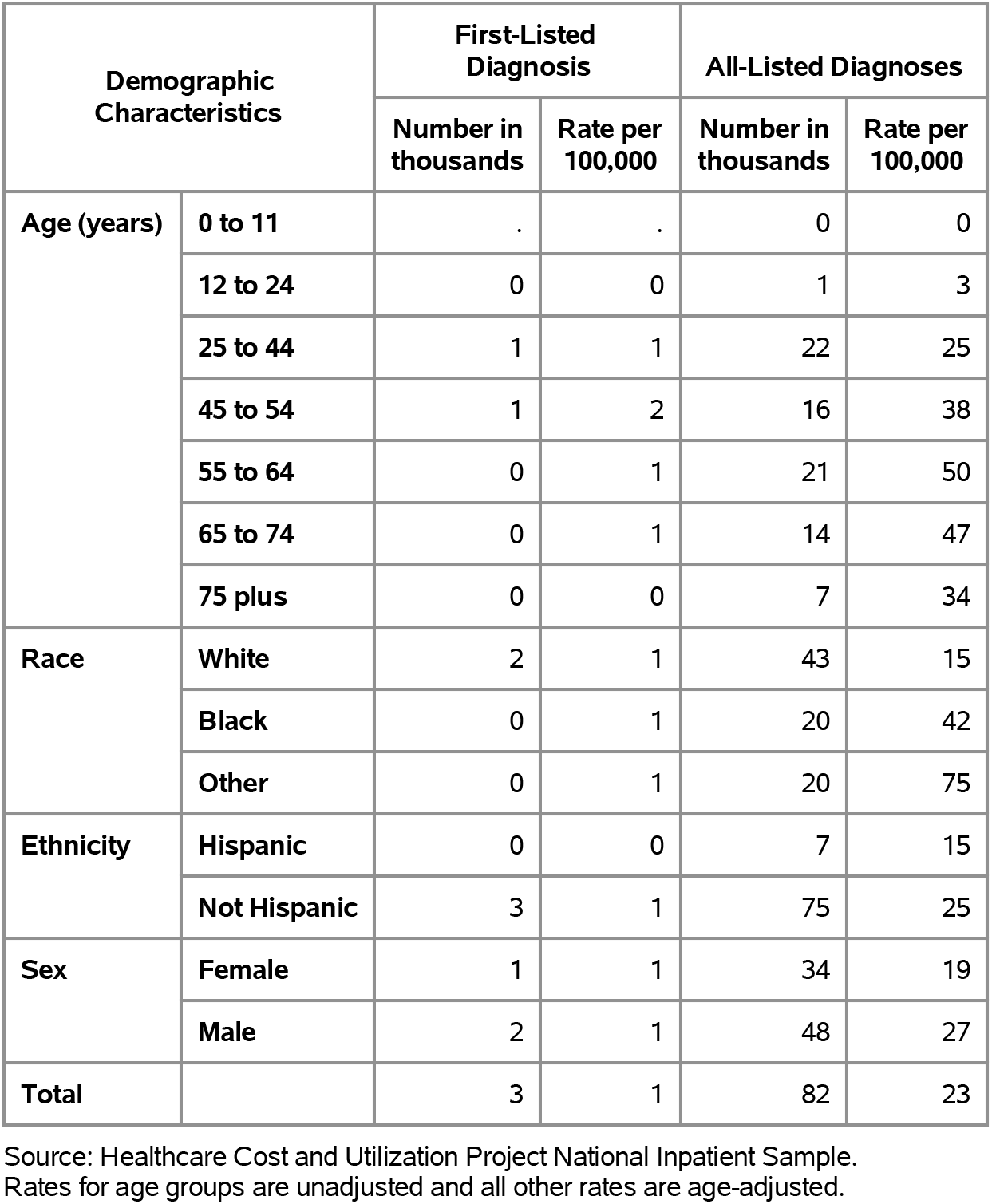
Hepatitis B: Hospital discharges with first-listed and all-listed diagnoses by age, race, ethnicity, and sex in the United States, 2018.

| Demographic Characteristics |  | First-Listed Diagnosis |  | All-Listed Diagnoses |  |
| --- | --- | --- | --- | --- | --- |
|  |  | Number in thousands | Rate per 100,000 | Number in thousands | Rate per 100,000 |
| Age (years) | 0 to 11 | . | . | 0 | 0 |
|  | 12 to 24 | 0 | 0 | 1 | 3 |
|  | 25 to 44 | 1 | 1 | 22 | 25 |
|  | 45 to 54 | 1 | 2 | 16 | 38 |
|  | 55 to 64 | 0 | 1 | 21 | 50 |
|  | 65 to 74 | 0 | 1 | 14 | 47 |
|  | 75 plus | 0 | 0 | 7 | 34 |
| Race | White | 2 | 1 | 43 | 15 |
|  | Black | 0 | 1 | 20 | 42 |
|  | Other | 0 | 1 | 20 | 75 |
| Ethnicity | Hispanic | 0 | 0 | 7 | 15 |
|  | Not Hispanic | 3 | 1 | 75 | 25 |
| Sex | Female | 1 | 1 | 34 | 19 |
|  | Male | 2 | 1 | 48 | 27 |
| Total |  | 3 | 1 | 82 | 23 |

Source: Healthcare Cost and Utilization Project National Inpatient Sample. Rates for age groups are unadjusted and all other rates are age-adjusted.

Hepatitis B contributed to 2,000 deaths in 2019 (Table 148). Mortality was uncommon among the youngest age groups after which rates (underlying or other cause) increased with age until the oldest age group. Like health care use rates, age-adjusted mortality rates were higher among men, Blacks, and non-Hispanics. Between 2004 and 2019, age-adjusted mortality rates (per 100,000) with hepatitis B as underlying or other cause decreased by a third from 0.6 to 0.4.(4)

**Table 148:** Hepatitis B: Deaths with underlying or underlying/other cause and lifetime years of life lost by age, race, ethnicity, and sex in the United States, 2019.

| Demographic Characteristics |  | Underlying Cause |  |  | Underlying/Other Cause |  |  |
| --- | --- | --- | --- | --- | --- | --- | --- |
|  |  | Number of Deaths | Rate per 100,000 | Years of potential life lost in thousands | Number of Deaths | Rate per 100,000 | Years of potential life lost in thousands |
| Age (years) | 0 to 11 | . | . | . | . | . | . |
|  | 12 to 24 | 1 | 0.0 | 0.0 | 4 | 0.0 | 0.2 |
|  | 25 to 44 | 42 | 0.0 | 1.7 | 153 | 0.2 | 6.3 |
|  | 45 to 54 | 86 | 0.2 | 2.6 | 256 | 0.6 | 7.7 |
|  | 55 to 64 | 118 | 0.3 | 2.7 | 506 | 1.2 | 11.3 |
|  | 65 to 74 | 113 | 0.4 | 1.8 | 486 | 1.5 | 7.7 |
|  | 75 plus | 77 | 0.3 | 0.7 | 267 | 1.2 | 2.2 |
| Race | White | 249 | 0.1 | 5.7 | 879 | 0.3 | 19.0 |
|  | Black | 59 | 0.1 | 1.3 | 299 | 0.6 | 6.6 |
|  | Other | 129 | 0.5 | 2.6 | 494 | 1.8 | 9.8 |
| Ethnicity | Hispanic | 33 | 0.1 | 0.8 | 84 | 0.2 | 1.9 |
|  | Not Hispanic | 404 | 0.1 | 8.8 | 1,588 | 0.5 | 33.5 |
| Sex | Female | 128 | 0.1 | 2.8 | 416 | 0.2 | 9.4 |
|  | Male | 309 | 0.2 | 6.7 | 1,256 | 0.7 | 26.0 |
| Total |  | 437 | 0.1 | 9.6 | 1,672 | 0.4 | 35.4 |

Source: Vital Statistics of the United States.
Rates for age groups are unadjusted and all other rates are age-adjusted.
Years of potential life lost is based on U.S. life table 2006-2018 estimates of life expectancy at age of death.

Among privately insured enrollees, the claims-based prevalence of hepatitis B (based on all-listed diagnoses) was 0.1% (Table 149). Hepatitis B was uncommon among children and adolescents and the youngest adults and prevalence was highest among persons 55-74 years. Prevalence did not differ by sex and was highest among Asians, similar among Blacks and Hispanics, and lowest among Whites. It was highest in the Northeast and lowest in the Midwest.

**Table 149:** Hepatitis B: Claims-based prevalence with first-listed and all-listed diagnoses by age, race-ethnicity, sex and region among privately insured enrollees, 2020.

| Demographic Characteristics |  | Number of enrollees | First-Listed Diagnosis |  | All-Listed Diagnoses |  |
| --- | --- | --- | --- | --- | --- | --- |
|  |  |  | Number of enrollees with disease | Percent of enrollees with disease | Number of enrollees with disease | Percent of enrollees with disease |
| Age (years) | 0 to 11 | 1,101,978 | 8 | 0.0% | 27 | 0.0% |
|  | 12 to 24 | 1,451,892 | 83 | 0.0% | 114 | 0.0% |
|  | 25 to 44 | 2,656,672 | 1,178 | 0.0% | 1,911 | 0.1% |
|  | 45 to 54 | 1,452,320 | 1,146 | 0.1% | 2,105 | 0.1% |
|  | 55 to 64 | 1,640,216 | 1,035 | 0.1% | 2,563 | 0.2% |
|  | 65 to 74 | 2,840,615 | 1,996 | 0.1% | 5,001 | 0.2% |
|  | 75 plus | 2,576,241 | 732 | 0.0% | 2,180 | 0.1% |
| Race-ethnicity | White | 8,657,702 | 1,325 | 0.0% | 3,977 | 0.0% |
|  | Black | 1,242,477 | 614 | 0.0% | 1,617 | 0.1% |
|  | Hispanic | 1,493,457 | 344 | 0.0% | 991 | 0.1% |
|  | Asian | 607,284 | 3,215 | 0.5% | 5,828 | 1.0% |
|  | Unknown | 1,719,014 | 680 | 0.0% | 1,488 | 0.1% |
| Sex | Female | 7,199,363 | 2,806 | 0.0% | 6,211 | 0.1% |
|  | Male | 6,520,571 | 3,372 | 0.1% | 7,690 | 0.1% |
| Region | Northeast | 1,526,275 | 1,253 | 0.1% | 2,465 | 0.2% |
|  | Midwest | 3,322,846 | 736 | 0.0% | 1,621 | 0.0% |
|  | South | 5,619,435 | 2,166 | 0.0% | 5,480 | 0.1% |
|  | West | 3,251,378 | 2,023 | 0.1% | 4,335 | 0.1% |
| Total |  | 13,719,934 | 6,178 | 0.0% | 13,901 | 0.1% |

Source: De-identified Optum Clinformatic® Data Mart.
Enrollees with full enrollment in a commercial health plan or Medicare during the year.

Among commercial insurance enrollees, ambulatory care visit rates with hepatitis B (all-listed diagnoses) increased with age until 75 years and were higher among men compared with women (Table 150). Among persons with known race-ethnicity, rates were much higher among Asians, followed by Blacks, then Hispanics, and lowest among Whites. Rates were highest in the Northeast, followed by the West, then the South, and lowest in the Midwest.

**Table 150:**
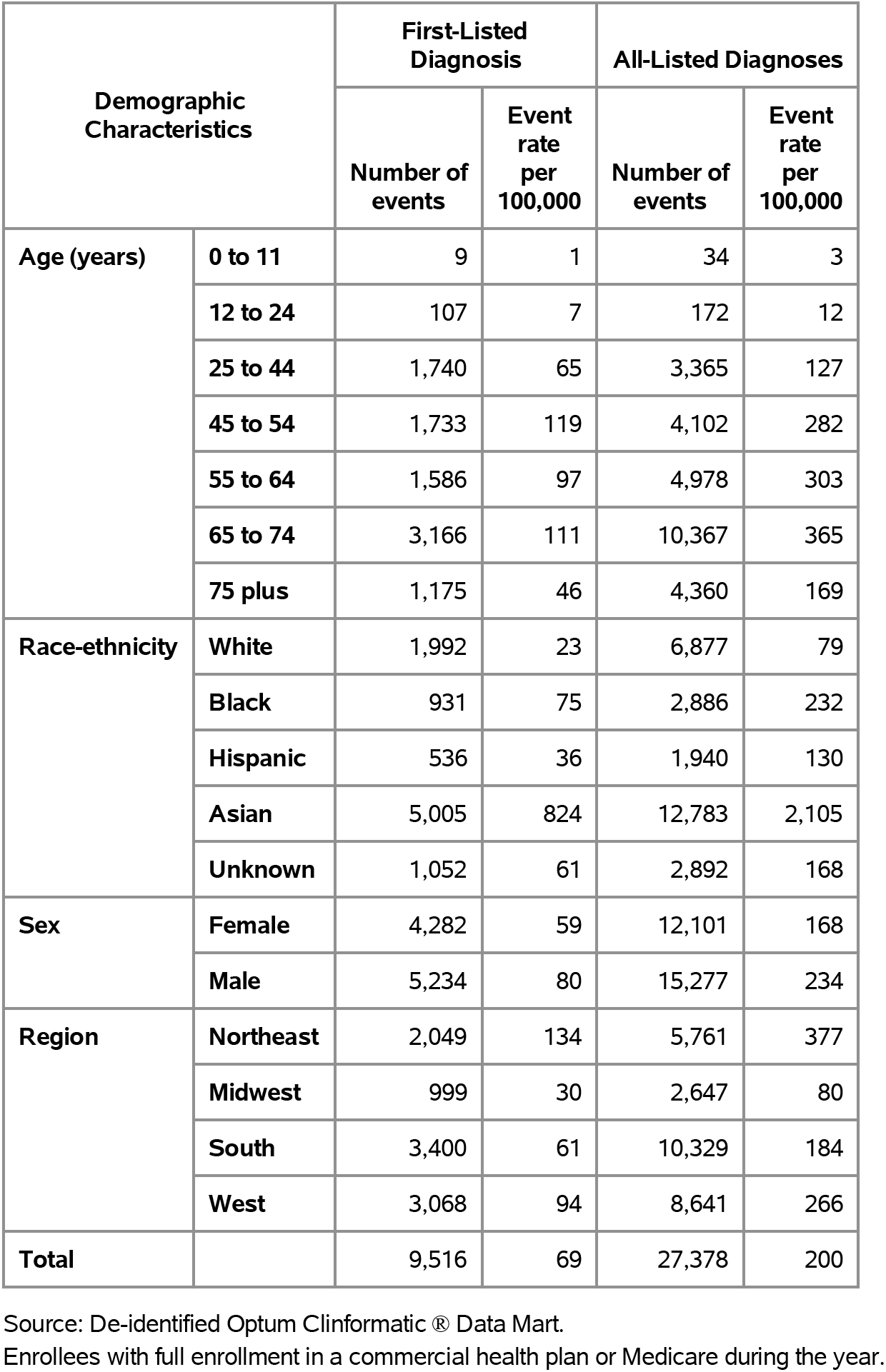
Hepatitis B: Ambulatory care visits with first-listed and all-listed diagnoses by age, race-ethnicity, sex and region among privately insured enrollees, 2020.

Among commercial insurance enrollees, emergency department visit rates with hepatitis B (all-listed diagnoses) were highest among persons 25 to 44 years and differed little by sex (Table 151). Among persons with known race-ethnicity, rates were much higher among Asians compared with other race-ethnicities. Rates were higher in the Northeast and South compared with the Midwest and West.

**Table 151:**
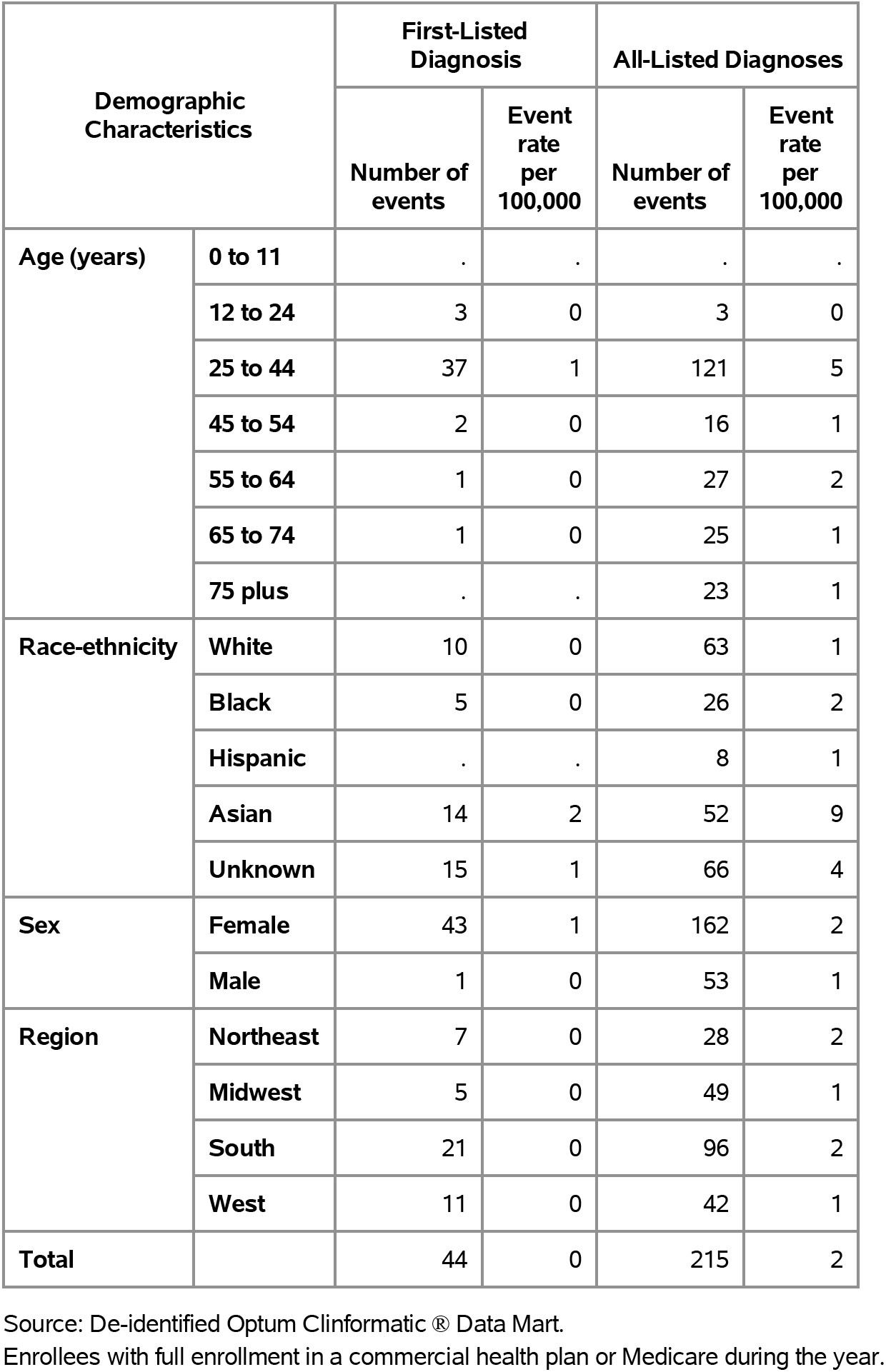
Hepatitis B: Emergency department visits with first-listed and all-listed diagnoses by age, race-ethnicity, sex and region among privately insured enrollees, 2020.

| Demographic Characteristics |  | First-Listed Diagnosis |  | All-Listed Diagnoses |  |
| --- | --- | --- | --- | --- | --- |
|  |  | Number of events | Event rate per 100,000 | Number of events | Event rate per 100,000 |
| Age (years) | 0 to 11 | . | . | . | . |
|  | 12 to 24 | 3 | 0 | 3 | 0 |
|  | 25 to 44 | 37 | 1 | 121 | 5 |
|  | 45 to 54 | 2 | 0 | 16 | 1 |
|  | 55 to 64 | 1 | 0 | 27 | 2 |
|  | 65 to 74 | 1 | 0 | 25 | 1 |
|  | 75 plus | . | . | 23 | 1 |
| Race-ethnicity | White | 10 | 0 | 63 | 1 |
|  | Black | 5 | 0 | 26 | 2 |
|  | Hispanic | . | . | 8 | 1 |
|  | Asian | 14 | 2 | 52 | 9 |
|  | Unknown | 15 | 1 | 66 | 4 |
| Sex | Female | 43 | 1 | 162 | 2 |
|  | Male | 1 | 0 | 53 | 1 |
| Region | Northeast | 7 | 0 | 28 | 2 |
|  | Midwest | 5 | 0 | 49 | 1 |
|  | South | 21 | 0 | 96 | 2 |
|  | West | 11 | 0 | 42 | 1 |
| Total |  | 44 | 0 | 215 | 2 |

Source: De-identified Optum Clinformatics® Data Mart.
Enrollees with full enrollment in a commercial health plan or Medicare during the year.

Among commercial insurance enrollees, hospital discharge rates with hepatitis B (all-listed diagnoses) increased with age until 75 years and were higher among men compared with women (Table 152). Among persons with known race-ethnicity, rates were highest among Asians, followed by Blacks, and lowest among Whites and Hispanics. Rates were highest in the Northeast, followed by the South, then the West, and lowest in the Midwest.

**Table 152:** Hepatitis B: Hospital discharges with first-listed and all-listed diagnoses by age, race-ethnicity, sex and region among privately insured enrollees, 2020.

| Demographic Characteristics |  | First-Listed Diagnosis |  | All-Listed Diagnoses |  |
| --- | --- | --- | --- | --- | --- |
|  |  | Number of events | Event rate per 100,000 | Number of events | Event rate per 100,000 |
| Age (years) | 0 to 11 | . | . | 4 | 0 |
|  | 12 to 24 | . | . | 14 | 1 |
|  | 25 to 44 | 8 | 0 | 211 | 8 |
|  | 45 to 54 | 6 | 0 | 225 | 15 |
|  | 55 to 64 | 6 | 0 | 457 | 28 |
|  | 65 to 74 | 7 | 0 | 870 | 31 |
|  | 75 plus | 4 | 0 | 504 | 20 |
| Race-ethnicity | White | 17 | 0 | 1,030 | 12 |
|  | Black | 4 | 0 | 434 | 35 |
|  | Hispanic | 7 | 0 | 189 | 13 |
|  | Asian | 2 | 0 | 414 | 68 |
|  | Unknown | 1 | 0 | 218 | 13 |
| Sex | Female | 12 | 0 | 900 | 13 |
|  | Male | 19 | 0 | 1,385 | 21 |
| Region | Northeast | 3 | 0 | 354 | 23 |
|  | Midwest | 3 | 0 | 327 | 10 |
|  | South | 23 | 0 | 1,110 | 20 |
|  | West | 2 | 0 | 494 | 15 |
| Total |  | 31 | 0 | 2,285 | 17 |

Source: De-identified Optum Clinformatic® Data Mart.
Enrollees with full enrollment in a commercial health plan or Medicare during the year.

Among Medicare beneficiaries, the claims-based prevalence of hepatitis B (based on all-listed diagnoses) was 0.1% (Table 153). Prevalence was highest among persons 65-74 years and was higher among men and Blacks. It was highest in the West, and lowest in the Midwest and South.

**Table 153:** Hepatitis B: Claims-based prevalence with first-listed and all-listed diagnoses by age, race, sex and region among fee-for-service, age-eligible Medicare beneficiaries, 2019.

| Demographic Characteristics |  | Number of enrollees | First-Listed Diagnosis |  | All-Listed Diagnoses |  |
| --- | --- | --- | --- | --- | --- | --- |
|  |  |  | Number of enrollees with disease | Percent of enrollees with disease | Number of enrollees with disease | Percent of enrollees with disease |
| Age (years) | 65 to 69 | 8,066,320 | 6,500 | 0.1% | 15,680 | 0.2% |
|  | 70 to 74 | 6,936,940 | 4,720 | 0.1% | 11,560 | 0.2% |
|  | 75 to 79 | 4,894,060 | 2,140 | 0.0% | 5,540 | 0.1% |
|  | 80 to 84 | 3,335,340 | 1,580 | 0.0% | 3,900 | 0.1% |
|  | 85+ | 3,578,120 | 740 | 0.0% | 2,280 | 0.1% |
| Race | White | 22,023,800 | 4,120 | 0.0% | 14,740 | 0.1% |
|  | Black | 1,861,660 | 1,480 | 0.1% | 4,560 | 0.2% |
|  | Other | 2,925,320 | 10,080 | 0.3% | 19,660 | 0.7% |
| Sex | Female | 14,975,560 | 8,060 | 0.1% | 18,960 | 0.1% |
|  | Male | 11,835,220 | 7,620 | 0.1% | 20,000 | 0.2% |
| Region | Northeast | 4,748,700 | 2,980 | 0.1% | 7,740 | 0.2% |
|  | Midwest | 6,069,800 | 1,780 | 0.0% | 5,080 | 0.1% |
|  | South | 10,595,900 | 4,700 | 0.0% | 12,300 | 0.1% |
|  | West | 5,396,380 | 6,220 | 0.1% | 13,840 | 0.3% |
| Total |  | 26,810,780 | 15,680 | 0.1% | 38,960 | 0.1% |

Source: Centers for Medicare & Medicaid Services, 5% Denominator and Claims Files.
Beneficiaries aged 65+ years with continuous and full Parts A and B enrollment and no Health Maintenance Organization enrollment during the year. Unweighted counts were multiplied by 20 to produce counts for this table.

Among Medicare beneficiaries, ambulatory care visit rates with hepatitis B (all-listed diagnoses) generally decreased with age and were higher among men compared with women and over four times higher among Blacks compared with Whites (Table 154). Rates were highest in the West, followed by the Northeast, then the South, and lowest in the Midwest.

**Table 154:** Hepatitis B: Ambulatory care visits with first-listed and all-listed diagnoses by age, race, sex and region among fee-for-service, age-eligible Medicare beneficiaries, 2019.

| Demographic Characteristics |  | First-Listed Diagnosis |  | All-Listed Diagnoses |  |
| --- | --- | --- | --- | --- | --- |
|  |  | Number of events | Event rate per 100,000 | Number of events | Event rate per 100,000 |
| Age (years) | 65 to 69 | 12,000 | 149 | 36,920 | 458 |
|  | 70 to 74 | 8,980 | 129 | 30,120 | 434 |
|  | 75 to 79 | 3,700 | 76 | 14,260 | 291 |
|  | 80 to 84 | 3,060 | 92 | 11,340 | 340 |
|  | 85+ | 1,180 | 33 | 5,700 | 159 |
| Race | White | 6,900 | 31 | 25,840 | 117 |
|  | Black | 2,660 | 143 | 9,540 | 512 |
|  | Other | 19,360 | 662 | 62,960 | 2,152 |
| Sex | Female | 14,860 | 99 | 48,540 | 324 |
|  | Male | 14,060 | 119 | 49,800 | 421 |
| Region | Northeast | 5,540 | 117 | 22,660 | 477 |
|  | Midwest | 2,960 | 49 | 9,700 | 160 |
|  | South | 8,500 | 80 | 26,100 | 246 |
|  | West | 11,920 | 221 | 39,880 | 739 |
| Total |  | 28,920 | 108 | 98,340 | 367 |

Source: Centers for Medicare & Medicaid Services, 5% Denominator and Claims Files. Beneficiaries aged 65+ years with continuous and full Parts A and B enrollment and no Health Maintenance Organization enrollment during the year. Unweighted counts were multiplied by 20 to produce counts for this table.

Among Medicare beneficiaries, emergency department visit rates with hepatitis B (all-listed diagnoses) were highest among persons 65 to 69 years and were higher among men compared with women and over six times higher among Blacks compared with Whites (Table 155). Rates were higher in the West compared with the South, Midwest, and Northeast.

**Table 155:** Hepatitis B: Emergency department visits with first-listed and all-listed diagnoses by age, race, sex and region among fee-for-service, age-eligible Medicare beneficiaries, 2019.

| Demographic Characteristics |  | First-Listed Diagnosis |  | All-Listed Diagnoses |  |
| --- | --- | --- | --- | --- | --- |
|  |  | Number of events | Event rate per 100,000 | Number of events | Event rate per 100,000 |
| Age (years) | 65 to 69 | . | . | 3,920 | 49 |
|  | 70 to 74 | . | . | 2,860 | 41 |
|  | 75 to 79 | . | . | 1,240 | 25 |
|  | 80 to 84 | . | . | 1,460 | 44 |
|  | 85+ | . | . | 1,020 | 29 |
| Race | White | . | . | 4,640 | 21 |
|  | Black | . | . | 2,540 | 136 |
|  | Other | . | . | 3,320 | 113 |
| Sex | Female | . | . | 4,400 | 29 |
|  | Male | . | . | 6,100 | 52 |
| Region | Northeast | . | . | 1,820 | 38 |
|  | Midwest | . | . | 2,160 | 36 |
|  | South | . | . | 3,700 | 35 |
|  | West | . | . | 2,820 | 52 |
| Total |  | . | . | 10,500 | 39 |

Source: Centers for Medicare & Medicaid Services, 5% Denominator and Claims Files. Beneficiaries aged 65+ years with continuous and full Parts A and B enrollment and no Health Maintenance Organization enrollment during the year. Unweighted counts were multiplied by 20 to produce counts for this table.

Among Medicare beneficiaries, hospital discharge rates with hepatitis B (all-listed diagnoses) were highest among persons 65 to 69 years and were higher among men compared with women and five times higher among Blacks compared with Whites (Table 156). Rates were highest in the West, followed by the Northeast, then the South, and lowest in the Midwest.

**Table 156:** Hepatitis B: Hospital discharges with first-listed and all-listed diagnoses by age, race, sex and region among fee-for-service, age-eligible Medicare beneficiaries, 2019.

| Demographic Characteristics |  | First-Listed Diagnosis |  | All-Listed Diagnoses |  |
| --- | --- | --- | --- | --- | --- |
|  |  | Number of events | Event rate per 100,000 | Number of events | Event rate per 100,000 |
| Age (years) | 65 to 69 | . | . | 3,940 | 49 |
|  | 70 to 74 | . | . | 3,040 | 44 |
|  | 75 to 79 | . | . | 1,440 | 29 |
|  | 80 to 84 | . | . | 1,440 | 43 |
|  | 85+ | . | . | 880 | 25 |
| Race | White | . | . | 5,240 | 24 |
|  | Black | . | . | 2,320 | 125 |
|  | Other | . | . | 3,180 | 109 |
| Sex | Female | . | . | 4,500 | 30 |
|  | Male | . | . | 6,240 | 53 |
| Region | Northeast | . | . | 2,080 | 44 |
|  | Midwest | . | . | 1,960 | 32 |
|  | South | . | . | 3,920 | 37 |
|  | West | . | . | 2,780 | 52 |
| Total |  | . | . | 10,740 | 40 |

Source: Centers for Medicare & Medicaid Services, 5% Denominator and Claims Files.
Beneficiaries aged 65+ years with continuous and full Parts A and B enrollment and no Health Maintenance Organization enrollment during the year. Unweighted counts were multiplied by 20 to produce counts for this table.

Hepatitis C contributed to 2.1 million ambulatory visits (2015) (Table 157). Ambulatory care visit rates (all-listed diagnoses) peaked among persons 55-64 years. Age-adjusted rates were higher among men compared with women, Blacks compared with Whites, and Hispanics compared with non-Hispanics.

**Table 157:** Hepatitis C: Ambulatory care visits with first-listed and all-listed diagnoses by age, race, ethnicity, and sex in the United States, 2015.

| Demographic Characteristics |  | First-Listed Diagnosis |  | All-Listed Diagnoses |  |
| --- | --- | --- | --- | --- | --- |
|  |  | Number in thousands | Rate per 100,000 | Number in thousands | Rate per 100,000 |
| Age (years) | 0 to 11 | . | . | 1 | 2 |
|  | 12 to 24 | 31 | 56 | 48 | 85 |
|  | 25 to 44 | 95 | 112 | 314 | 372 |
|  | 45 to 54 | 151 | 349 | 371 | 861 |
|  | 55 to 64 | 242 | 592 | 831 | 2,037 |
|  | 65 to 74 | 61 | 223 | 392 | 1,425 |
|  | 75 plus | . | . | 114 | 565 |
| Race | White | 478 | 165 | 1,719 | 561 |
|  | Black | 97 | 203 | 271 | 567 |
|  | Other | 5 | 21 | 82 | 309 |
| Ethnicity | Hispanic | 15 | 29 | 352 | 818 |
|  | Not Hispanic | 565 | 176 | 1,719 | 525 |
| Sex | Female | 280 | 138 | 888 | 434 |
|  | Male | 300 | 174 | 1,184 | 665 |
| Total |  | 580 | 156 | 2,072 | 542 |

Source: National Ambulatory Medical Care Survey (3-year average, 2014-2016). Rates for age groups are unadjusted and all other rates are age-adjusted.

Hepatitis C contributed to 865,000 emergency department visits in 2018 (Table 158). Emergency department visit rates (all-listed diagnoses) peaked among persons 55-64 years. Age-adjusted rates were higher among men compared with women.

**Table 158:** Hepatitis C: Emergency department visits with first-listed and all-listed diagnoses by age and sex in the United States, 2018.

| Demographic Characteristics |  | First-Listed Diagnosis |  | All-Listed Diagnoses |  |
| --- | --- | --- | --- | --- | --- |
|  |  | Number in thousands | Rate per 100,000 | Number in thousands | Rate per 100,000 |
| Age (years) | 0 to 11 | 0 | 0 | 0 | 1 |
|  | 12 to 24 | 1 | 1 | 15 | 27 |
|  | 25 to 44 | 4 | 4 | 217 | 249 |
|  | 45 to 54 | 1 | 3 | 182 | 436 |
|  | 55 to 64 | 1 | 4 | 300 | 710 |
|  | 65 to 74 | 1 | 2 | 124 | 407 |
|  | 75 plus | 0 | 1 | 26 | 120 |
| Sex | Female | 3 | 2 | 333 | 181 |
|  | Male | 5 | 3 | 531 | 290 |
| Total |  | 8 | 2 | 865 | 234 |

Source: Healthcare Cost and Utilization Project Nationwide Emergency Department Sample. Rates for age groups are unadjusted and all other rates are age-adjusted.

Hepatitis C contributed to 600,000 hospital discharges in 2018 (Table 159). Hospital discharge rates (all-listed diagnoses) peaked among persons 55-64 years. Age-adjusted rates were higher among men compared with women, Blacks compared with Whites, and non-Hispanics compared with Hispanics. Between 2004 and 2018, age-adjusted hospital discharge rates (per 100,000) with an all-listed diagnosis increased by 12% from 143 to 160.(7)

**Table 159:**
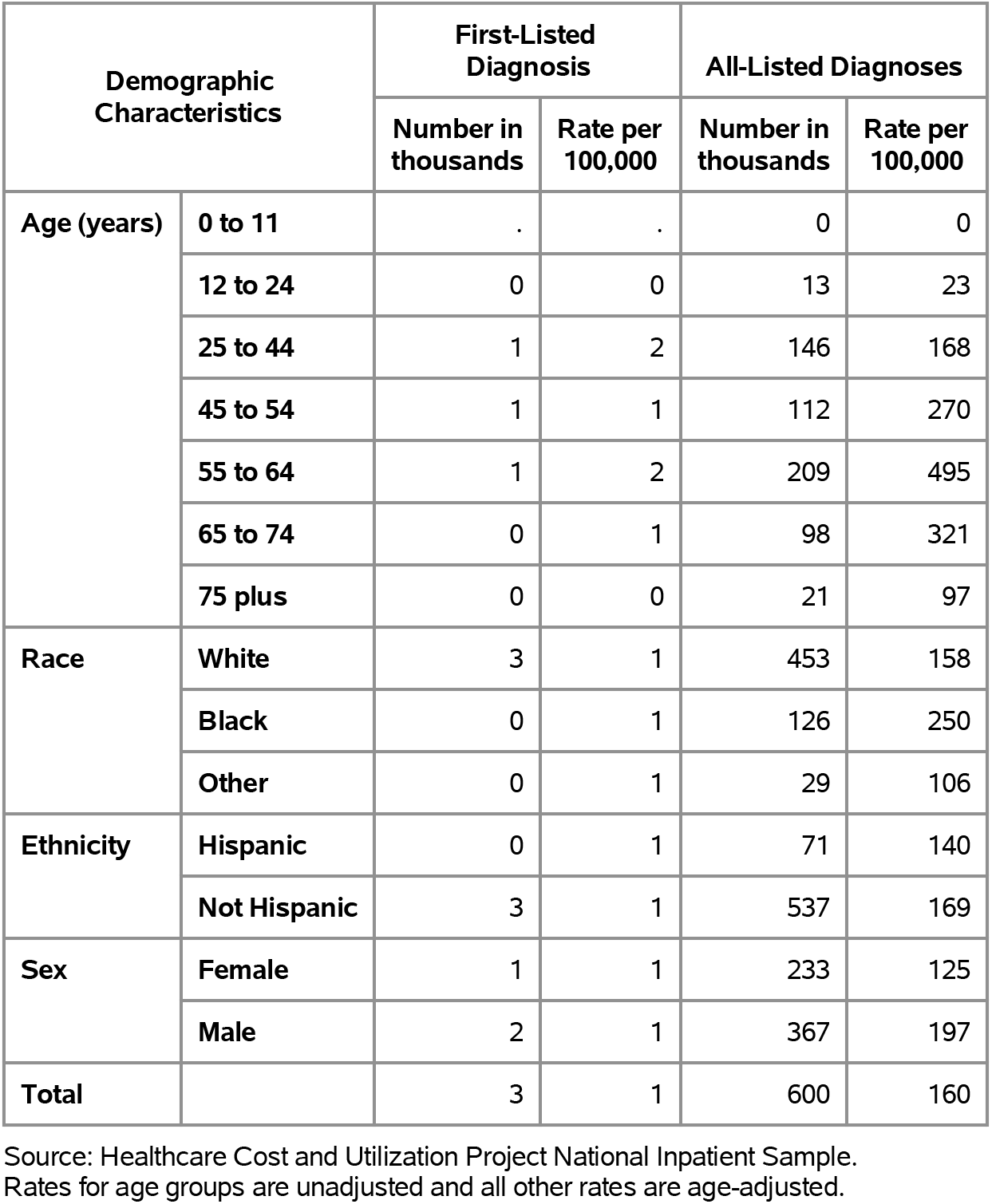
Hepatitis C: Hospital discharges with first-listed and all-listed diagnoses by age, race, ethnicity, and sex in the United States, 2018.

| Demographic Characteristics |  | First-Listed Diagnosis |  | All-Listed Diagnoses |  |
| --- | --- | --- | --- | --- | --- |
|  |  | Number in thousands | Rate per 100,000 | Number in thousands | Rate per 100,000 |
| Age (years) | 0 to 11 | . | . | 0 | 0 |
|  | 12 to 24 | 0 | 0 | 13 | 23 |
|  | 25 to 44 | 1 | 2 | 146 | 168 |
|  | 45 to 54 | 1 | 1 | 112 | 270 |
|  | 55 to 64 | 1 | 2 | 209 | 495 |
|  | 65 to 74 | 0 | 1 | 98 | 321 |
|  | 75 plus | 0 | 0 | 21 | 97 |
| Race | White | 3 | 1 | 453 | 158 |
|  | Black | 0 | 1 | 126 | 250 |
|  | Other | 0 | 1 | 29 | 106 |
| Ethnicity | Hispanic | 0 | 1 | 71 | 140 |
|  | Not Hispanic | 3 | 1 | 537 | 169 |
| Sex | Female | 1 | 1 | 233 | 125 |
|  | Male | 2 | 1 | 367 | 197 |
| Total |  | 3 | 1 | 600 | 160 |

Source: Healthcare Cost and Utilization Project National Inpatient Sample. Rates for age groups are unadjusted and all other rates are age-adjusted.

Hepatitis C contributed to 14,000 deaths in 2019 (Table 160). Mortality rates (underlying or other cause) peaked among persons 55-64 years. Age-adjusted mortality rates were higher among men, Blacks, and non-Hispanics. Between 2004 and 2019, age-adjusted mortality rates (per 100,000) with hepatitis C as underlying or other cause decreased by 13% from 3.8 to 3.3. (4)

**Table 160:** Hepatitis C: Deaths with underlying or underlying/other cause and lifetime years of life lost by age, race, ethnicity, and sex in the United States, 2019.

| Demographic Characteristics |  | Underlying Cause |  |  | Underlying/Other Cause |  |  |
| --- | --- | --- | --- | --- | --- | --- | --- |
|  |  | Number of Deaths | Rate per 100,000 | Years of potential life lost in thousands | Number of Deaths | Rate per 100,000 | Years of potential life lost in thousands |
| Age (years) | 0 to 11 | . | . | . | . | . | . |
|  | 12 to 24 | 2 | 0.0 | 0.1 | 7 | 0.0 | 0.4 |
|  | 25 to 44 | 144 | 0.2 | 6.0 | 643 | 0.7 | 27.2 |
|  | 45 to 54 | 478 | 1.2 | 14.6 | 1,685 | 4.1 | 50.9 |
|  | 55 to 64 | 1,525 | 3.6 | 34.8 | 6,343 | 14.9 | 141.7 |
|  | 65 to 74 | 1,045 | 3.3 | 17.1 | 4,515 | 14.3 | 72.6 |
|  | 75 plus | 327 | 1.4 | 2.8 | 1,120 | 5.0 | 9.6 |
| Race | White | 2,826 | 0.8 | 62.4 | 10,999 | 3.2 | 241.5 |
|  | Black | 550 | 1.1 | 10.1 | 2,693 | 5.2 | 48.1 |
|  | Other | 145 | 0.5 | 2.9 | 621 | 2.3 | 12.9 |
| Ethnicity | Hispanic | 451 | 0.9 | 10.7 | 1,457 | 3.0 | 34.1 |
|  | Not Hispanic | 3,070 | 0.8 | 64.7 | 12,856 | 3.4 | 268.4 |
| Sex | Female | 1,258 | 0.6 | 28.6 | 4,030 | 1.8 | 93.6 |
|  | Male | 2,263 | 1.1 | 46.8 | 10,283 | 4.9 | 208.9 |
| Total |  | 3,521 | 0.8 | 75.4 | 14,313 | 3.3 | 302.5 |

Source: Vital Statistics of the United States.
Rates for age groups are unadjusted and all other rates are age-adjusted.
Years of potential life lost is based on U.S. life table 2006-2018 estimates of life expectancy at age of death.

Among privately insured enrollees, the claims-based prevalence of hepatitis C (based on all-listed diagnoses) was 0.3% (Table 161). Prevalence peaked among persons 55-74 years and was higher among men. It was highest among Blacks, followed by Hispanics, and lowest among Whites and Asians. It was highest in the South and lowest in the Midwest.

**Table 161:** Hepatitis C: Claims-based prevalence with first-listed and all-listed diagnoses by age, race-ethnicity, sex and region among privately insured enrollees, 2020.

| Demographic Characteristics |  | Number of enrollees | First-Listed Diagnosis |  | All-Listed Diagnoses |  |
| --- | --- | --- | --- | --- | --- | --- |
|  |  |  | Number of enrollees with disease | Percent of enrollees with disease | Number of enrollees with disease | Percent of enrollees with disease |
| Age (years) | 0 to 11 | 1,101,978 | 9 | 0.0% | 23 | 0.0% |
|  | 12 to 24 | 1,451,892 | 78 | 0.0% | 143 | 0.0% |
|  | 25 to 44 | 2,656,672 | 855 | 0.0% | 1,904 | 0.1% |
|  | 45 to 54 | 1,452,320 | 788 | 0.1% | 2,502 | 0.2% |
|  | 55 to 64 | 1,640,216 | 2,851 | 0.2% | 10,497 | 0.6% |
|  | 65 to 74 | 2,840,615 | 4,838 | 0.2% | 18,032 | 0.6% |
|  | 75 plus | 2,576,241 | 923 | 0.0% | 4,272 | 0.2% |
| Race-ethnicity | White | 8,657,702 | 5,560 | 0.1% | 20,564 | 0.2% |
|  | Black | 1,242,477 | 2,102 | 0.2% | 7,171 | 0.6% |
|  | Hispanic | 1,493,457 | 1,197 | 0.1% | 4,562 | 0.3% |
|  | Asian | 607,284 | 276 | 0.0% | 1,047 | 0.2% |
|  | Unknown | 1,719,014 | 1,207 | 0.1% | 4,029 | 0.2% |
| Sex | Female | 7,199,363 | 4,044 | 0.1% | 14,997 | 0.2% |
|  | Male | 6,520,571 | 6,298 | 0.1% | 22,376 | 0.3% |
| Region | Northeast | 1,526,275 | 1,188 | 0.1% | 4,269 | 0.3% |
|  | Midwest | 3,322,846 | 1,356 | 0.0% | 4,930 | 0.1% |
|  | South | 5,619,435 | 5,668 | 0.1% | 20,093 | 0.4% |
|  | West | 3,251,378 | 2,130 | 0.1% | 8,081 | 0.2% |
| Total |  | 13,719,934 | 10,342 | 0.1% | 37,373 | 0.3% |

Source: De-identified Optum Clinformatic® Data Mart.
Enrollees with full enrollment in a commercial health plan or Medicare during the year.

Among commercial insurance enrollees, ambulatory care visit rates with hepatitis C (all-listed diagnoses) peaked among persons 55 to 64 years and were higher among men compared with women (Table 162). Among persons with known race-ethnicity, rates were much higher among Blacks, followed by Hispanics, then Whites, and lowest among Asians. Rates were highest in the South, followed by the Northeast, then the West, and lowest in the Midwest.

**Table 162:** Hepatitis C: Ambulatory care visits with first-listed and all-listed diagnoses by age, race-ethnicity, sex and region among privately insured enrollees, 2020.

| Demographic Characteristics |  | First-Listed Diagnosis |  | All-Listed Diagnoses |  |
| --- | --- | --- | --- | --- | --- |
|  |  | Number of events | Event rate per 100,000 | Number of events | Event rate per 100,000 |
| Age (years) | 0 to 11 | 16 | 1 | 45 | 4 |
|  | 12 to 24 | 126 | 9 | 271 | 19 |
|  | 25 to 44 | 1,457 | 55 | 3,692 | 139 |
|  | 45 to 54 | 1,606 | 111 | 5,314 | 366 |
|  | 55 to 64 | 4,825 | 294 | 21,782 | 1,328 |
|  | 65 to 74 | 7,997 | 282 | 36,232 | 1,275 |
|  | 75 plus | 1,459 | 57 | 8,009 | 311 |
| Race-ethnicity | White | 9,257 | 107 | 40,491 | 468 |
|  | Black | 3,514 | 283 | 14,905 | 1,200 |
|  | Hispanic | 1,946 | 130 | 9,346 | 626 |
|  | Asian | 408 | 67 | 1,923 | 317 |
|  | Unknown | 2,361 | 137 | 8,680 | 505 |
| Sex | Female | 6,530 | 91 | 29,672 | 412 |
|  | Male | 10,956 | 168 | 45,673 | 700 |
| Region | Northeast | 1,902 | 125 | 8,348 | 547 |
|  | Midwest | 2,427 | 73 | 9,366 | 282 |
|  | South | 9,675 | 172 | 41,631 | 741 |
|  | West | 3,482 | 107 | 16,000 | 492 |
| Total |  | 17,486 | 127 | 75,345 | 549 |

Source: De-identified Optum Clinformatic® Data Mart.
Enrollees with full enrollment in a commercial health plan or Medicare during the year.

Among commercial insurance enrollees, emergency department visit rates with hepatitis C (all-listed diagnoses) peaked among persons 55 to 64 years and were higher among men compared with women (Table 163). Among persons with known race-ethnicity, rates were highest among Blacks, followed by Whites and Hispanics, and lowest among Asians. Rates were higher in the Northeast and South compared with the West and Midwest.

**Table 163:** Hepatitis C: Emergency department visits with first-listed and all-listed diagnoses by age, race-ethnicity, sex and region among privately insured enrollees, 2020.

| Demographic Characteristics |  | First-Listed Diagnosis |  | All-Listed Diagnoses |  |
| --- | --- | --- | --- | --- | --- |
|  |  | Number of events | Event rate per 100,000 | Number of events | Event rate per 100,000 |
| Age (years) | 0 to 11 | . | . | . | . |
|  | 12 to 24 | . | . | 4 | 0 |
|  | 25 to 44 | 5 | 0 | 65 | 2 |
|  | 45 to 54 | 3 | 0 | 55 | 4 |
|  | 55 to 64 | 19 | 1 | 213 | 13 |
|  | 65 to 74 | 10 | 0 | 234 | 8 |
|  | 75 plus | 3 | 0 | 56 | 2 |
| Race-ethnicity | White | 24 | 0 | 347 | 4 |
|  | Black | 12 | 1 | 114 | 9 |
|  | Hispanic | 2 | 0 | 60 | 4 |
|  | Asian | . | . | 11 | 2 |
|  | Unknown | 2 | 0 | 95 | 6 |
| Sex | Female | 17 | 0 | 228 | 3 |
|  | Male | 23 | 0 | 399 | 6 |
| Region | Northeast | 1 | 0 | 95 | 6 |
|  | Midwest | 7 | 0 | 93 | 3 |
|  | South | 27 | 0 | 315 | 6 |
|  | West | 5 | 0 | 124 | 4 |
| Total |  | 40 | 0 | 627 | 5 |

Source: De-identified Optum Clinformatic® Data Mart.
Enrollees with full enrollment in a commercial health plan or Medicare during the year.

Among commercial insurance enrollees, hospital discharge rates with hepatitis C (all-listed diagnoses) peaked among persons 55 to 64 years and were higher among men compared with women (Table 164). Among persons with known race-ethnicity, rates were highest among Blacks, followed by Hispanics, then Whites, and lowest among Asians. Rates were highest in the South, followed by the Northeast, then the West, and lowest in the Midwest.

**Table 164:** Hepatitis C: Hospital discharges with first-listed and all-listed diagnoses by age, race-ethnicity, sex and region among privately insured enrollees, 2020.

| Demographic Characteristics |  | First-Listed Diagnosis |  | All-Listed Diagnoses |  |
| --- | --- | --- | --- | --- | --- |
|  |  | Number of events | Event rate per 100,000 | Number of events | Event rate per 100,000 |
| Age (years) | 0 to 11 | . | . | 1 | 0 |
|  | 12 to 24 | . | . | 41 | 3 |
|  | 25 to 44 | 1 | 0 | 436 | 16 |
|  | 45 to 54 | 2 | 0 | 924 | 64 |
|  | 55 to 64 | 6 | 0 | 3,968 | 242 |
|  | 65 to 74 | 18 | 1 | 5,690 | 200 |
|  | 75 plus | 3 | 0 | 1,480 | 57 |
| Race-ethnicity | White | 16 | 0 | 6,740 | 78 |
|  | Black | 5 | 0 | 2,771 | 223 |
|  | Hispanic | 2 | 0 | 1,281 | 86 |
|  | Asian | . | . | 196 | 32 |
|  | Unknown | 7 | 0 | 1,552 | 90 |
| Sex | Female | 14 | 0 | 4,904 | 68 |
|  | Male | 16 | 0 | 7,636 | 117 |
| Region | Northeast | 1 | 0 | 1,405 | 92 |
|  | Midwest | 5 | 0 | 1,764 | 53 |
|  | South | 18 | 0 | 7,127 | 127 |
|  | West | 6 | 0 | 2,244 | 69 |
| Total |  | 30 | 0 | 12,540 | 91 |

Source: De-identified Optum Clinformatic® Data Mart.
Enrollees with full enrollment in a commercial health plan or Medicare during the year.

Among Medicare beneficiaries, the claims-based prevalence of hepatitis C (based on all-listed diagnoses) was 0.5% (Table 165). Prevalence decreased with age and was higher among men and Blacks. It was highest in the West, and lowest in the Midwest.

**Table 165:**
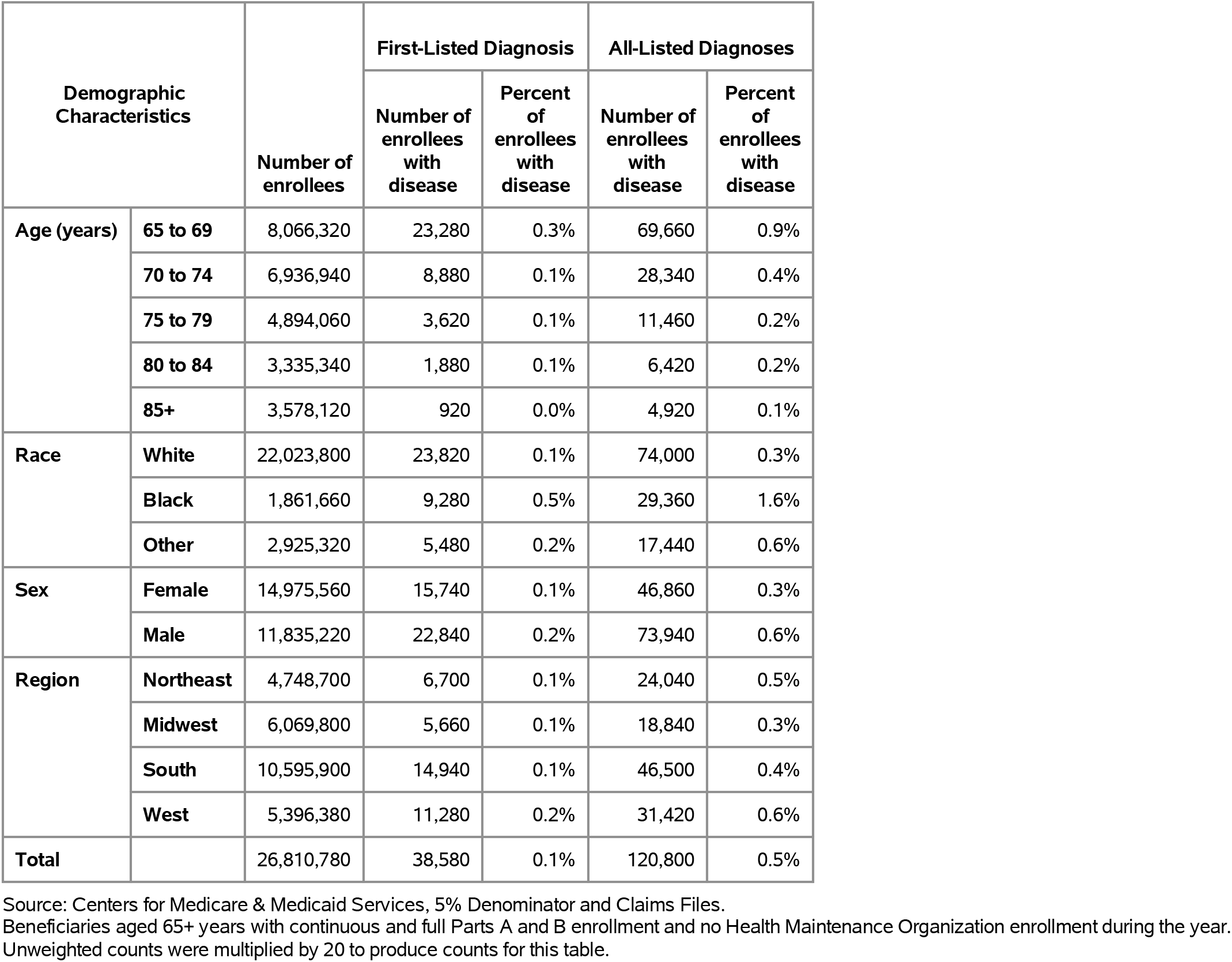
Hepatitis C: Claims-based prevalence with first-listed and all-listed diagnoses by age, race, sex and region among fee-for-service, age-eligible Medicare beneficiaries, 2019.

| Demographic Characteristics |  | Number of enrollees | First-Listed Diagnosis |  | All-Listed Diagnoses |  |
| --- | --- | --- | --- | --- | --- | --- |
|  |  |  | Number of enrollees with disease | Percent of enrollees with disease | Number of enrollees with disease | Percent of enrollees with disease |
| Age (years) | 65 to 69 | 8,066,320 | 23,280 | 0.3% | 69,660 | 0.9% |
|  | 70 to 74 | 6,936,940 | 8,880 | 0.1% | 28,340 | 0.4% |
|  | 75 to 79 | 4,894,060 | 3,620 | 0.1% | 11,460 | 0.2% |
|  | 80 to 84 | 3,335,340 | 1,880 | 0.1% | 6,420 | 0.2% |
|  | 85+ | 3,578,120 | 920 | 0.0% | 4,920 | 0.1% |
| Race | White | 22,023,800 | 23,820 | 0.1% | 74,000 | 0.3% |
|  | Black | 1,861,660 | 9,280 | 0.5% | 29,360 | 1.6% |
|  | Other | 2,925,320 | 5,480 | 0.2% | 17,440 | 0.6% |
| Sex | Female | 14,975,560 | 15,740 | 0.1% | 46,860 | 0.3% |
|  | Male | 11,835,220 | 22,840 | 0.2% | 73,940 | 0.6% |
| Region | Northeast | 4,748,700 | 6,700 | 0.1% | 24,040 | 0.5% |
|  | Midwest | 6,069,800 | 5,660 | 0.1% | 18,840 | 0.3% |
|  | South | 10,595,900 | 14,940 | 0.1% | 46,500 | 0.4% |
|  | West | 5,396,380 | 11,280 | 0.2% | 31,420 | 0.6% |
| Total |  | 26,810,780 | 38,580 | 0.1% | 120,800 | 0.5% |

Source: Centers for Medicare & Medicaid Services, 5% Denominator and Claims Files.
Beneficiaries aged 65+ years with continuous and full Parts A and B enrollment and no Health Maintenance Organization enrollment during the year. Unweighted counts were multiplied by 20 to produce counts for this table.

Among Medicare beneficiaries, ambulatory care visit rates with hepatitis C (all-listed diagnoses) decreased with age and were twice as high among men compared with women and over five times higher among Blacks compared with Whites (Table 166). Rates were highest in the West, followed by the Northeast and South, and lowest in the Midwest.

**Table 166:** Hepatitis C: Ambulatory care visits with first-listed and all-listed diagnoses by age, race, sex and region among fee-for-service, age-eligible Medicare beneficiaries, 2019.

| Demographic Characteristics |  | First-Listed Diagnosis |  | All-Listed Diagnoses |  |
| --- | --- | --- | --- | --- | --- |
|  |  | Number of events | Event rate per 100,000 | Number of events | Event rate per 100,000 |
| Age (years) | 65 to 69 | 42,580 | 528 | 154,180 | 1,911 |
|  | 70 to 74 | 16,240 | 234 | 63,940 | 922 |
|  | 75 to 79 | 6,720 | 137 | 25,720 | 526 |
|  | 80 to 84 | 4,420 | 133 | 14,380 | 431 |
|  | 85+ | 1,840 | 51 | 9,300 | 260 |
| Race | White | 41,560 | 189 | 154,420 | 701 |
|  | Black | 18,460 | 992 | 70,440 | 3,784 |
|  | Other | 11,780 | 403 | 42,660 | 1,458 |
| Sex | Female | 29,940 | 200 | 102,340 | 683 |
|  | Male | 41,860 | 354 | 165,180 | 1,396 |
| Region | Northeast | 12,240 | 258 | 51,280 | 1,080 |
|  | Midwest | 9,460 | 156 | 38,660 | 637 |
|  | South | 28,120 | 265 | 107,020 | 1,010 |
|  | West | 21,980 | 407 | 70,560 | 1,308 |
| Total |  | 71,800 | 268 | 267,520 | 998 |

Source: Centers for Medicare & Medicaid Services, 5% Denominator and Claims Files. Beneficiaries aged 65+ years with continuous and full Parts A and B enrollment and no Health Maintenance Organization enrollment during the year. Unweighted counts were multiplied by 20 to produce counts for this table.

Among Medicare beneficiaries, emergency department visit rates with hepatitis C (all-listed diagnoses) decreased with age and were over twice as high among men compared with women and seven times higher among Blacks compared with Whites (Table 167). Rates were highest in the West, followed by the Northeast, then the South, and lowest in the Midwest.

**Table 167:** Hepatitis C: Emergency department visits with first-listed and all-listed diagnoses by age, race, sex and region among fee-for-service, age-eligible Medicare beneficiaries, 2019.

| Demographic Characteristics |  | First-Listed Diagnosis |  | All-Listed Diagnoses |  |
| --- | --- | --- | --- | --- | --- |
|  |  | Number of events | Event rate per 100,000 | Number of events | Event rate per 100,000 |
| Age (years) | 65 to 69 | 240 | 3 | 36,740 | 455 |
|  | 70 to 74 | . | . | 14,120 | 204 |
|  | 75 to 79 | . | . | 6,280 | 128 |
|  | 80 to 84 | . | . | 2,920 | 88 |
|  | 85+ | . | . | 2,940 | 82 |
| Race | White | . | . | 34,500 | 157 |
|  | Black | . | . | 20,620 | 1,108 |
|  | Other | . | . | 7,880 | 269 |
| Sex | Female | . | . | 23,300 | 156 |
|  | Male | . | . | 39,700 | 335 |
| Region | Northeast | . | . | 12,780 | 269 |
|  | Midwest | . | . | 10,660 | 176 |
|  | South | . | . | 23,600 | 223 |
|  | West | . | . | 15,960 | 296 |
| Total |  | 400 | 1 | 63,000 | 235 |

Source: Centers for Medicare & Medicaid Services, 5% Denominator and Claims Files. Beneficiaries aged 65+ years with continuous and full Parts A and B enrollment and no Health Maintenance Organization enrollment during the year. Unweighted counts were multiplied by 20 to produce counts for this table.

Among Medicare beneficiaries, hospital discharge rates with hepatitis C (all-listed diagnoses) decreased with age and were over twice as high among men compared with women and over six times higher among Blacks compared with Whites (Table 168). Rates were highest in the Northeast, followed by the West, then the South, and lowest in the Midwest.

**Table 168:** Hepatitis C: Hospital discharges with first-listed and all-listed diagnoses by age, race, sex and region among fee-for-service, age-eligible Medicare beneficiaries, 2019.

| Demographic Characteristics |  | First-Listed Diagnosis |  | All-Listed Diagnoses |  |
| --- | --- | --- | --- | --- | --- |
|  |  | Number of events | Event rate per 100,000 | Number of events | Event rate per 100,000 |
| Age (years) | 65 to 69 | . | . | 30,660 | 380 |
|  | 70 to 74 | . | . | 11,920 | 172 |
|  | 75 to 79 | . | . | 5,460 | 112 |
|  | 80 to 84 | . | . | 2,500 | 75 |
|  | 85+ | . | . | 2,520 | 70 |
| Race | White | . | . | 30,080 | 137 |
|  | Black | . | . | 16,160 | 868 |
|  | Other | . | . | 6,820 | 233 |
| Sex | Female | . | . | 19,280 | 129 |
|  | Male | . | . | 33,780 | 285 |
| Region | Northeast | . | . | 11,400 | 240 |
|  | Midwest | . | . | 8,940 | 147 |
|  | South | . | . | 20,100 | 190 |
|  | West | . | . | 12,620 | 234 |
| Total |  | . | . | 53,060 | 198 |

Source: Centers for Medicare & Medicaid Services, 5% Denominator and Claims Files. Beneficiaries aged 65+ years with continuous and full Parts A and B enrollment and no Health Maintenance Organization enrollment during the year. Unweighted counts were multiplied by 20 to produce counts for this table.

Gallstones contributed to an estimated 2.2 million ambulatory visits (2015) (Table 169). Ambulatory care visits rates (all-listed diagnoses) were highest among persons 65-74 years. Age-adjusted rates were higher among women compared with men, Whites compared with Blacks, and Hispanics compared with non-Hispanics.

**Table 169:** Gallstones: Ambulatory care visits with first-listed and all-listed diagnoses by age, race, ethnicity, and sex in the United States, 2015.

| Demographic Characteristics |  | First-Listed Diagnosis |  | All-Listed Diagnoses |  |
| --- | --- | --- | --- | --- | --- |
|  |  | Number in thousands | Rate per 100,000 | Number in thousands | Rate per 100,000 |
| Age (years) | 0 to 11 | 2 | 4 | 2 | 4 |
|  | 12 to 24 | 123 | 219 | 148 | 263 |
|  | 25 to 44 | 383 | 453 | 565 | 669 |
|  | 45 to 54 | 233 | 542 | 467 | 1,084 |
|  | 55 to 64 | 328 | 804 | 414 | 1,015 |
|  | 65 to 74 | 172 | 624 | 389 | 1,415 |
|  | 75 plus | 113 | 558 | 218 | 1,080 |
| Race | White | 1,139 | 426 | 1,901 | 691 |
|  | Black | 147 | 313 | 172 | 380 |
|  | Other | 67 | 235 | 131 | 519 |
| Ethnicity | Hispanic | 491 | 1,029 | 600 | 1,254 |
|  | Not Hispanic | 863 | 305 | 1,604 | 554 |
| Sex | Female | 899 | 546 | 1,466 | 870 |
|  | Male | 454 | 255 | 738 | 418 |
| Total |  | 1,354 | 397 | 2,204 | 642 |

Source: National Ambulatory Medical Care Survey (3-year average, 2014-2016). Rates for age groups are unadjusted and all other rates are age-adjusted.

Gallstones contributed to an estimated 1.1 million emergency department visits in 2018 (Table 170). Emergency department visit rates (all-listed diagnoses) increased with age. Age-adjusted rates were higher among women compared with men.

**Table 170:** Gallstones: Emergency department visits with first-listed and all-listed diagnoses by age and sex in the United States, 2018.

| Demographic Characteristics |  | First-Listed Diagnosis |  | All-Listed Diagnoses |  |
| --- | --- | --- | --- | --- | --- |
|  |  | Number in thousands | Rate per 100,000 | Number in thousands | Rate per 100,000 |
| Age (years) | 0 to 11 | 1 | 2 | 2 | 5 |
|  | 12 to 24 | 71 | 128 | 97 | 175 |
|  | 25 to 44 | 238 | 273 | 335 | 385 |
|  | 45 to 54 | 97 | 234 | 163 | 390 |
|  | 55 to 64 | 88 | 208 | 179 | 423 |
|  | 65 to 74 | 71 | 234 | 169 | 553 |
|  | 75 plus | 67 | 305 | 205 | 935 |
| Sex | Female | 437 | 260 | 734 | 417 |
|  | Male | 196 | 117 | 416 | 243 |
| Total |  | 633 | 188 | 1,149 | 328 |

Source: Healthcare Cost and Utilization Project Nationwide Emergency Department Sample. Rates for age groups are unadjusted and all other rates are age-adjusted.

Gallstones contributed to an estimated 615,000 hospital discharges in 2018 (Table 171). Hospital discharge rates (all-listed diagnoses) increased with age. Age-adjusted rates were higher among women compared with men, Whites compared with Blacks, and Hispanics compared with non-Hispanics. Between 2004 and 2018, age-adjusted hospital discharge rates (per 100,000) with an all-listed diagnosis decreased by 21% from 212 to 167.(7) Hospital discharge rates underestimate the actual burden because most hospitalizations with gallstones were for cholecystectomy and a high proportion of cholecystectomies were performed laparoscopically without an overnight stay, and therefore, were not included in hospitalization statistics.

**Table 171:** Gallstones: Hospital discharges with first-listed and all-listed diagnoses by age, race, ethnicity, and sex in the United States, 2018.

| Demographic Characteristics |  | First-Listed Diagnosis |  | All-Listed Diagnoses |  |
| --- | --- | --- | --- | --- | --- |
|  |  | Number in thousands | Rate per 100,000 | Number in thousands | Rate per 100,000 |
| Age (years) | 0 to 11 | 1 | 1 | 2 | 4 |
|  | 12 to 24 | 18 | 32 | 29 | 52 |
|  | 25 to 44 | 67 | 78 | 114 | 131 |
|  | 45 to 54 | 38 | 91 | 77 | 186 |
|  | 55 to 64 | 45 | 107 | 108 | 256 |
|  | 65 to 74 | 46 | 149 | 121 | 397 |
|  | 75 plus | 53 | 243 | 164 | 746 |
| Race | White | 222 | 78 | 502 | 169 |
|  | Black | 27 | 61 | 72 | 165 |
|  | Other | 21 | 82 | 48 | 188 |
| Ethnicity | Hispanic | 58 | 112 | 112 | 233 |
|  | Not Hispanic | 213 | 69 | 510 | 158 |
| Sex | Female | 166 | 92 | 353 | 186 |
|  | Male | 102 | 59 | 263 | 151 |
| Total |  | 268 | 75 | 615 | 167 |

Source: Healthcare Cost and Utilization Project National Inpatient Sample. Rates for age groups are unadjusted and all other rates are age-adjusted.

Gallstones contributed to an estimated 2,000 deaths in 2019 (Table 172). Mortality was uncommon among the youngest age groups after which rates (underlying or other cause) increased with age. Age-adjusted mortality rates were higher among men and did not differ by race or ethnicity. Between 2004 and 2019, age-adjusted mortality rates (per 100,000) with gallstone disease as underlying or other cause decreased by 43% from 0.7 to 0.4.(4)

**Table 172:** Gallstones: Deaths with underlying or underlying/other cause and lifetime years of life lost by age, race, ethnicity, and sex in the United States, 2019.

| Demographic Characteristics |  | Underlying Cause |  |  | Underlying/Other Cause |  |  |
| --- | --- | --- | --- | --- | --- | --- | --- |
|  |  | Number of Deaths | Rate per 100,000 | Years of potential life lost in thousands | Number of Deaths | Rate per 100,000 | Years of potential life lost in thousands |
| Age (years) | 0 to 11 | . | . | . | . | . | . |
|  | 12 to 24 | 1 | 0.0 | 0.1 | 3 | 0.0 | 0.2 |
|  | 25 to 44 | 21 | 0.0 | 0.9 | 61 | 0.1 | 2.6 |
|  | 45 to 54 | 38 | 0.1 | 1.2 | 66 | 0.2 | 2.1 |
|  | 55 to 64 | 99 | 0.2 | 2.3 | 190 | 0.4 | 4.4 |
|  | 65 to 74 | 181 | 0.6 | 2.8 | 307 | 1.0 | 4.8 |
|  | 75 plus | 710 | 3.1 | 4.4 | 1,169 | 5.2 | 7.4 |
| Race | White | 892 | 0.3 | 9.8 | 1,517 | 0.4 | 17.6 |
|  | Black | 77 | 0.2 | 1.0 | 154 | 0.4 | 2.4 |
|  | Other | 81 | 0.4 | 0.9 | 125 | 0.5 | 1.5 |
| Ethnicity | Hispanic | 99 | 0.3 | 1.4 | 157 | 0.4 | 2.5 |
|  | Not Hispanic | 951 | 0.3 | 10.3 | 1,639 | 0.4 | 19.0 |
| Sex | Female | 564 | 0.2 | 5.9 | 959 | 0.4 | 10.9 |
|  | Male | 486 | 0.3 | 5.7 | 837 | 0.5 | 10.5 |
| Total |  | 1,050 | 0.3 | 11.7 | 1,796 | 0.4 | 21.5 |

Source: Vital Statistics of the United States.
Rates for age groups are unadjusted and all other rates are age-adjusted.
Years of potential life lost is based on U.S. life table 2006-2018 estimates of life expectancy at age of death.

Among privately insured enrollees, the claims-based prevalence of gallstones (based on all-listed diagnoses) was 0.7% (Table 173). Prevalence increased with age and was higher among women. It was highest among Hispanics, similar among Whites and Blacks, and lowest among Asians. It differed little by region.

**Table 173:**
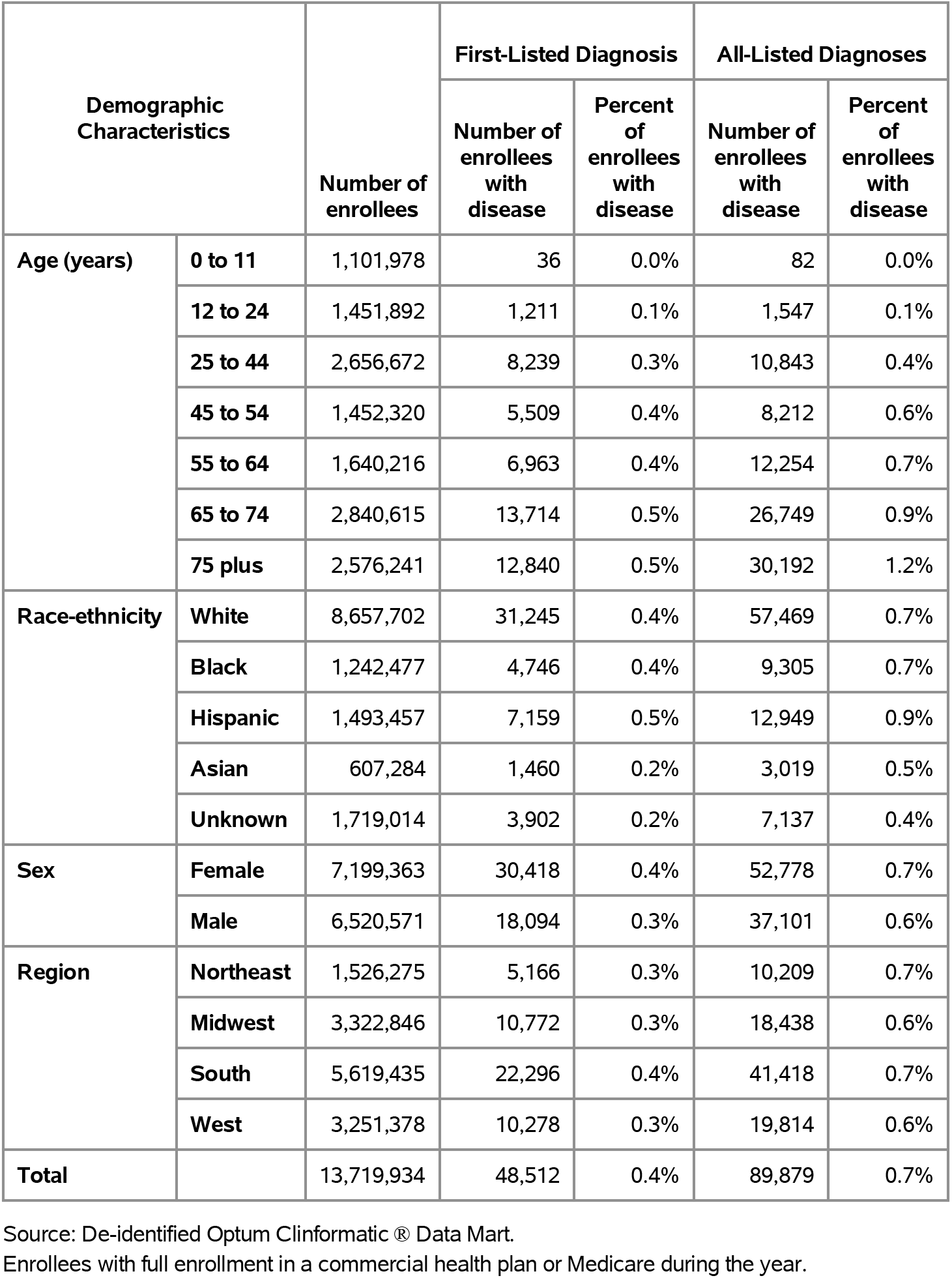
Gallstones: Claims-based prevalence with first-listed and all-listed diagnoses by age, race-ethnicity, sex and region among privately insured enrollees, 2020.

Among commercial insurance enrollees, ambulatory care visit rates with gallstones (all-listed diagnoses) increased with age and were higher among women compared with men (Table 174). Among persons with known race-ethnicity, rates were highest among Hispanics, followed by Blacks, then Whites, and lowest among Asians. Rates were highest in the South, followed by the Northeast, then the West, and lowest in the Midwest.

**Table 174:** Gallstones: Ambulatory care visits with first-listed and all-listed diagnoses by age, race-ethnicity, sex and region among privately insured enrollees, 2020.

| Demographic Characteristics |  | First-Listed Diagnosis |  | All-Listed Diagnoses |  |
| --- | --- | --- | --- | --- | --- |
|  |  | Number of events | Event rate per 100,000 | Number of events | Event rate per 100,000 |
| Age (years) | 0 to 11 | 51 | 5 | 115 | 10 |
|  | 12 to 24 | 2,211 | 152 | 2,883 | 199 |
|  | 25 to 44 | 14,812 | 558 | 20,158 | 759 |
|  | 45 to 54 | 9,409 | 648 | 14,121 | 972 |
|  | 55 to 64 | 10,982 | 670 | 19,041 | 1,161 |
|  | 65 to 74 | 19,796 | 697 | 37,835 | 1,332 |
|  | 75 plus | 16,815 | 653 | 38,397 | 1,490 |
| Race-ethnicity | White | 47,672 | 551 | 84,416 | 975 |
|  | Black | 7,180 | 578 | 13,137 | 1,057 |
|  | Hispanic | 11,029 | 738 | 19,525 | 1,307 |
|  | Asian | 2,152 | 354 | 4,656 | 767 |
|  | Unknown | 6,043 | 352 | 10,816 | 629 |
| Sex | Female | 48,371 | 672 | 82,723 | 1,149 |
|  | Male | 25,705 | 394 | 49,827 | 764 |
| Region | Northeast | 7,416 | 486 | 14,458 | 947 |
|  | Midwest | 16,514 | 497 | 27,831 | 838 |
|  | South | 34,550 | 615 | 61,224 | 1,090 |
|  | West | 15,596 | 480 | 29,037 | 893 |
| Total |  | 74,076 | 540 | 132,550 | 966 |

Source: De-identified Optum Clinformatic® Data Mart.
Enrollees with full enrollment in a commercial health plan or Medicare during the year.

Among commercial insurance enrollees, emergency department visit rates with gallstones (all-listed diagnoses) increased with age and were higher among women compared with men (Table 175). Among persons with known race-ethnicity, rates were highest among Blacks and Hispanics, followed by Whites, and lowest among Asians. Rates were lower in the Midwest compared with the South, West, and Northeast.

**Table 175:** Gallstones: Emergency department visits with first-listed and all-listed diagnoses by age, race-ethnicity, sex and region among privately insured enrollees, 2020.

| Demographic Characteristics |  | First-Listed Diagnosis |  | All-Listed Diagnoses |  |
| --- | --- | --- | --- | --- | --- |
|  |  | Number of events | Event rate per 100,000 | Number of events | Event rate per 100,000 |
| Age (years) | 0 to 11 | . | . | . | . |
|  | 12 to 24 | 33 | 2 | 52 | 4 |
|  | 25 to 44 | 328 | 12 | 542 | 20 |
|  | 45 to 54 | 301 | 21 | 503 | 35 |
|  | 55 to 64 | 693 | 42 | 1,215 | 74 |
|  | 65 to 74 | 2,783 | 98 | 5,048 | 178 |
|  | 75 plus | 2,979 | 116 | 6,000 | 233 |
| Race-ethnicity | White | 4,534 | 52 | 8,558 | 99 |
|  | Black | 789 | 64 | 1,515 | 122 |
|  | Hispanic | 1,016 | 68 | 1,814 | 121 |
|  | Asian | 172 | 28 | 340 | 56 |
|  | Unknown | 606 | 35 | 1,133 | 66 |
| Sex | Female | 4,215 | 59 | 7,754 | 108 |
|  | Male | 2,902 | 45 | 5,606 | 86 |
| Region | Northeast | 921 | 60 | 1,545 | 101 |
|  | Midwest | 1,499 | 45 | 2,706 | 81 |
|  | South | 2,981 | 53 | 5,800 | 103 |
|  | West | 1,716 | 53 | 3,309 | 102 |
| Total |  | 7,117 | 52 | 13,360 | 97 |

Source: De-identified Optum Clinformatic® Data Mart.
Enrollees with full enrollment in a commercial health plan or Medicare during the year.

Among commercial insurance enrollees, hospital discharge rates with gallstones (all-listed diagnoses) increased with age and differed little by sex (Table 176). Among persons with known race-ethnicity, rates were highest among Hispanics, followed by Blacks, then Whites, and lowest among Asians. Rates were highest in the Northeast, followed by the South, then the West, and lowest in the Midwest.

**Table 176:** Gallstones: Hospital discharges with first-listed and all-listed diagnoses by age, race-ethnicity, sex and region among privately insured enrollees, 2020.

| Demographic Characteristics |  | First-Listed Diagnosis |  | All-Listed Diagnoses |  |
| --- | --- | --- | --- | --- | --- |
|  |  | Number of events | Event rate per 100,000 | Number of events | Event rate per 100,000 |
| Age (years) | 0 to 11 | 3 | 0 | 29 | 3 |
|  | 12 to 24 | 132 | 9 | 259 | 18 |
|  | 25 to 44 | 830 | 31 | 1,766 | 66 |
|  | 45 to 54 | 670 | 46 | 1,716 | 118 |
|  | 55 to 64 | 1,109 | 68 | 3,526 | 215 |
|  | 65 to 74 | 2,847 | 100 | 9,549 | 336 |
|  | 75 plus | 4,128 | 160 | 15,044 | 584 |
| Race-ethnicity | White | 6,342 | 73 | 20,616 | 238 |
|  | Black | 926 | 75 | 3,565 | 287 |
|  | Hispanic | 1,441 | 96 | 4,412 | 295 |
|  | Asian | 264 | 43 | 837 | 138 |
|  | Unknown | 746 | 43 | 2,459 | 143 |
| Sex | Female | 5,441 | 76 | 16,727 | 232 |
|  | Male | 4,278 | 66 | 15,162 | 233 |
| Region | Northeast | 1,320 | 86 | 4,145 | 272 |
|  | Midwest | 2,059 | 62 | 6,564 | 198 |
|  | South | 4,204 | 75 | 14,278 | 254 |
|  | West | 2,136 | 66 | 6,902 | 212 |
| Total |  | 9,719 | 71 | 31,889 | 232 |

Source: De-identified Optum Clinformatic® Data Mart.
Enrollees with full enrollment in a commercial health plan or Medicare during the year.

Among Medicare beneficiaries, the claims-based prevalence of gallstones (based on all-listed diagnoses) was 1.7% (Table 177). Prevalence increased with age, was higher among men, and did not differ by race. It was lower in the Midwest compared with other regions.

**Table 177:** Gallstones: Claims-based prevalence with first-listed and all-listed diagnoses by age, race, sex and region among fee-for-service, age-eligible Medicare beneficiaries, 2019.

| Demographic Characteristics |  | Number of enrollees | First-Listed Diagnosis |  | All-Listed Diagnoses |  |
| --- | --- | --- | --- | --- | --- | --- |
|  |  |  | Number of enrollees with disease | Percent of enrollees with disease | Number of enrollees with disease | Percent of enrollees with disease |
| Age (years) | 65 to 69 | 8,066,320 | 56,600 | 0.7% | 105,120 | 1.3% |
|  | 70 to 74 | 6,936,940 | 53,220 | 0.8% | 105,220 | 1.5% |
|  | 75 to 79 | 4,894,060 | 41,060 | 0.8% | 84,680 | 1.7% |
|  | 80 to 84 | 3,335,340 | 32,620 | 1.0% | 70,320 | 2.1% |
|  | 85+ | 3,578,120 | 34,720 | 1.0% | 77,220 | 2.2% |
| Race | White | 22,023,800 | 178,220 | 0.8% | 359,020 | 1.6% |
|  | Black | 1,861,660 | 13,680 | 0.7% | 29,380 | 1.6% |
|  | Other | 2,925,320 | 26,320 | 0.9% | 54,160 | 1.9% |
| Sex | Female | 14,975,560 | 121,980 | 0.8% | 237,800 | 1.6% |
|  | Male | 11,835,220 | 96,240 | 0.8% | 204,760 | 1.7% |
| Region | Northeast | 4,748,700 | 37,900 | 0.8% | 80,840 | 1.7% |
|  | Midwest | 6,069,800 | 46,320 | 0.8% | 91,140 | 1.5% |
|  | South | 10,595,900 | 90,360 | 0.9% | 181,020 | 1.7% |
|  | West | 5,396,380 | 43,640 | 0.8% | 89,560 | 1.7% |
| Total |  | 26,810,780 | 218,220 | 0.8% | 442,560 | 1.7% |

Source: Centers for Medicare & Medicaid Services, 5% Denominator and Claims Files. Beneficiaries aged 65+ years with continuous and full Parts A and B enrollment and no Health Maintenance Organization enrollment during the year. Unweighted counts were multiplied by 20 to produce counts for this table.

Among Medicare beneficiaries, ambulatory care visit rates with gallstones (all-listed diagnoses) increased with age until 85 years and were higher among women compared with men and Whites compared with Blacks (Table 178). Rates were highest in the Northeast, intermediate in the South and West, and lowest in the Midwest.

**Table 178:** Gallstones: Ambulatory care visits with first-listed and all-listed diagnoses by age, race, sex and region among fee-for-service, age-eligible Medicare beneficiaries, 2019.

| Demographic Characteristics |  | First-Listed Diagnosis |  | All-Listed Diagnoses |  |
| --- | --- | --- | --- | --- | --- |
|  |  | Number of events | Event rate per 100,000 | Number of events | Event rate per 100,000 |
| Age (years) | 65 to 69 | 67,100 | 832 | 132,220 | 1,639 |
|  | 70 to 74 | 60,220 | 868 | 127,760 | 1,842 |
|  | 75 to 79 | 44,580 | 911 | 98,400 | 2,011 |
|  | 80 to 84 | 32,480 | 974 | 75,720 | 2,270 |
|  | 85+ | 29,720 | 831 | 73,600 | 2,057 |
| Race | White | 192,640 | 875 | 410,820 | 1,865 |
|  | Black | 12,440 | 668 | 29,220 | 1,570 |
|  | Other | 29,020 | 992 | 67,660 | 2,313 |
| Sex | Female | 135,260 | 903 | 286,460 | 1,913 |
|  | Male | 98,840 | 835 | 221,240 | 1,869 |
| Region | Northeast | 41,460 | 873 | 99,840 | 2,102 |
|  | Midwest | 49,200 | 811 | 101,620 | 1,674 |
|  | South | 98,260 | 927 | 203,340 | 1,919 |
|  | West | 45,180 | 837 | 102,900 | 1,907 |
| Total |  | 234,100 | 873 | 507,700 | 1,894 |

Source: Centers for Medicare & Medicaid Services, 5% Denominator and Claims Files. Beneficiaries aged 65+ years with continuous and full Parts A and B enrollment and no Health Maintenance Organization enrollment during the year. Unweighted counts were multiplied by 20 to produce counts for this table.

Among Medicare beneficiaries, emergency department visit rates with gallstones (all-listed diagnoses) increased with age and were higher among men compared with women and Blacks compared with Whites (Table 179). Rates were highest in the South, followed by the West, then the Northeast, and lowest in the Midwest.

**Table 179:** Gallstones: Emergency department visits with first-listed and all-listed diagnoses by age, race, sex and region among fee-for-service, age-eligible Medicare beneficiaries, 2019.

| Demographic Characteristics |  | First-Listed Diagnosis |  | All-Listed Diagnoses |  |
| --- | --- | --- | --- | --- | --- |
|  |  | Number of events | Event rate per 100,000 | Number of events | Event rate per 100,000 |
| Age (years) | 65 to 69 | 25,800 | 320 | 53,680 | 665 |
|  | 70 to 74 | 25,340 | 365 | 55,220 | 796 |
|  | 75 to 79 | 20,860 | 426 | 47,900 | 979 |
|  | 80 to 84 | 17,760 | 532 | 43,640 | 1,308 |
|  | 85+ | 23,180 | 648 | 58,780 | 1,643 |
| Race | White | 90,480 | 411 | 206,440 | 937 |
|  | Black | 8,400 | 451 | 21,460 | 1,153 |
|  | Other | 14,060 | 481 | 31,320 | 1,071 |
| Sex | Female | 60,720 | 405 | 134,780 | 900 |
|  | Male | 52,220 | 441 | 124,440 | 1,051 |
| Region | Northeast | 19,600 | 413 | 44,740 | 942 |
|  | Midwest | 23,120 | 381 | 54,360 | 896 |
|  | South | 46,140 | 435 | 107,020 | 1,010 |
|  | West | 24,080 | 446 | 53,100 | 984 |
| Total |  | 112,940 | 421 | 259,220 | 967 |

Source: Centers for Medicare & Medicaid Services, 5% Denominator and Claims Files. Beneficiaries aged 65+ years with continuous and full Parts A and B enrollment and no Health Maintenance Organization enrollment during the year. Unweighted counts were multiplied by 20 to produce counts for this table.

Among Medicare beneficiaries, hospital discharge rates with gallstones (all-listed diagnoses) increased with age and were higher among men compared with women and Blacks compared with Whites (Table 180). Rates were higher in the South compared with the Northeast, West, and Midwest.

**Table 180:**
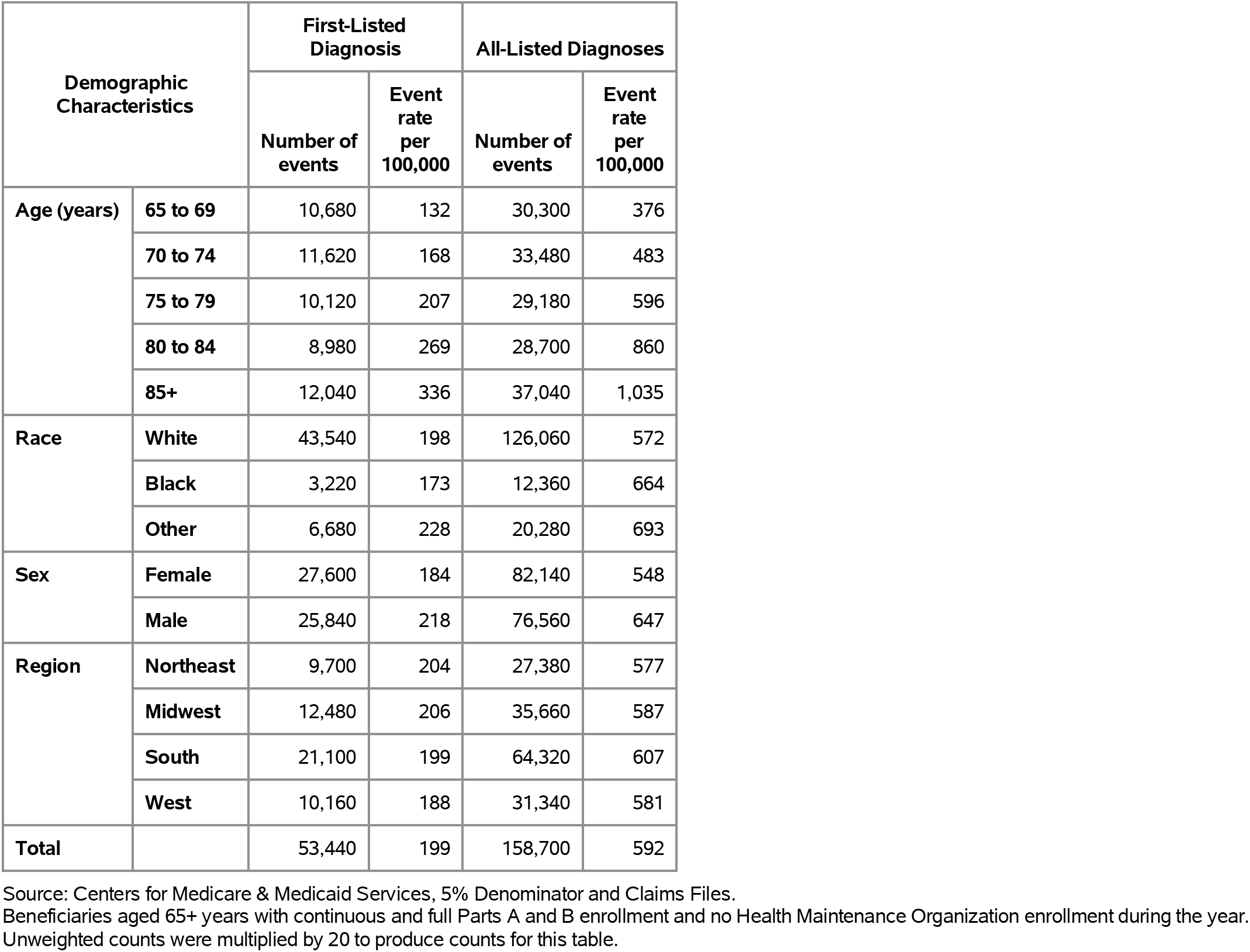
Gallstones: Hospital discharges with first-listed and all-listed diagnoses by age, race, sex and region among fee-for-service, age-eligible Medicare beneficiaries, 2019.

Acute pancreatitis contributed to 897,000 ambulatory visits (2015) (Table 181). Ambulatory care visit rates (all-listed diagnoses) peaked among persons 55-64 years. Age-adjusted ambulatory care visit rates were higher among women compared with men, Blacks compared with Whites and among non-Hispanics compared with Hispanics.

**Table 181:** Acute Pancreatitis: Ambulatory care visits with first-listed and all-listed diagnoses by age, race, ethnicity, and sex in the United States, 2015.

| Demographic Characteristics |  | First-Listed Diagnosis |  | All-Listed Diagnoses |  |
| --- | --- | --- | --- | --- | --- |
|  |  | Number in thousands | Rate per 100,000 | Number in thousands | Rate per 100,000 |
| Age (years) | 0 to 11 | 2 | 5 | 2 | 5 |
|  | 12 to 24 | 25 | 45 | 32 | 57 |
|  | 25 to 44 | 113 | 133 | 202 | 239 |
|  | 45 to 54 | 165 | 383 | 234 | 543 |
|  | 55 to 64 | 86 | 212 | 236 | 578 |
|  | 65 to 74 | 32 | 117 | 125 | 453 |
|  | 75 plus | 21 | 103 | 66 | 325 |
| Race | White | 391 | 158 | 720 | 275 |
|  | Black | 47 | 111 | 161 | 333 |
|  | Other | 7 | 31 | 16 | 78 |
| Ethnicity | Hispanic | 73 | 140 | 125 | 265 |
|  | Not Hispanic | 371 | 144 | 772 | 269 |
| Sex | Female | 239 | 155 | 484 | 282 |
|  | Male | 206 | 125 | 413 | 250 |
| Total |  | 445 | 140 | 897 | 264 |

Source: National Ambulatory Medical Care Survey (3-year average, 2014-2016). Rates for age groups are unadjusted and all other rates are age-adjusted.

Acute pancreatitis contributed to 585,000 emergency department visits in 2018 (Table 182). Emergency department visit rates (all-listed diagnoses) were similar among middle-aged and older adults. Age-adjusted emergency department visit rates were higher among men.

**Table 182:** Acute Pancreatitis: Emergency department visits with first-listed and all-listed diagnoses by age and sex in the United States, 2018.

| Demographic Characteristics |  | First-Listed Diagnosis |  | All-Listed Diagnoses |  |
| --- | --- | --- | --- | --- | --- |
|  |  | Number in thousands | Rate per 100,000 | Number in thousands | Rate per 100,000 |
| Age (years) | 0 to 11 | 2 | 4 | 3 | 6 |
|  | 12 to 24 | 20 | 36 | 31 | 55 |
|  | 25 to 44 | 123 | 141 | 177 | 203 |
|  | 45 to 54 | 82 | 197 | 122 | 294 |
|  | 55 to 64 | 73 | 173 | 115 | 273 |
|  | 65 to 74 | 44 | 144 | 74 | 243 |
|  | 75 plus | 33 | 152 | 62 | 284 |
| Sex | Female | 172 | 98 | 274 | 154 |
|  | Male | 206 | 125 | 310 | 187 |
| Total |  | 377 | 111 | 585 | 170 |

Source: Healthcare Cost and Utilization Project Nationwide Emergency Department Sample. Rates for age groups are unadjusted and all other rates are age-adjusted.

Acute pancreatitis contributed to 448,000 hospital discharges in 2018 (Table 183). Hospital discharge rates (all-listed diagnoses) were highest among persons 75 years and over. Age-adjusted rates were higher among women compared with men, Blacks compared with Whites and among non-Hispanics compared with Hispanics.

**Table 183:** Acute Pancreatitis: Hospital discharges with first-listed and all-listed diagnoses by age, race, ethnicity, and sex in the United States, 2018.

| Demographic Characteristics |  | First-Listed Diagnosis |  | All-Listed Diagnoses |  |
| --- | --- | --- | --- | --- | --- |
|  |  | Number in thousands | Rate per 100,000 | Number in thousands | Rate per 100,000 |
| Age (years) | 0 to 11 | 2 | 3 | 3 | 6 |
|  | 12 to 24 | 14 | 25 | 22 | 40 |
|  | 25 to 44 | 88 | 101 | 126 | 145 |
|  | 45 to 54 | 60 | 144 | 88 | 212 |
|  | 55 to 64 | 56 | 133 | 89 | 211 |
|  | 65 to 74 | 35 | 116 | 63 | 207 |
|  | 75 plus | 29 | 131 | 56 | 256 |
| Race | White | 221 | 82 | 351 | 127 |
|  | Black | 48 | 108 | 71 | 159 |
|  | Other | 17 | 65 | 29 | 112 |
| Ethnicity | Hispanic | 40 | 76 | 66 | 130 |
|  | Not Hispanic | 247 | 86 | 386 | 131 |
| Sex | Female | 131 | 74 | 213 | 118 |
|  | Male | 153 | 92 | 235 | 140 |
| Total |  | 283 | 83 | 448 | 129 |

Source: Healthcare Cost and Utilization Project National Inpatient Sample. Rates for age groups are unadjusted and all other rates are age-adjusted.

Acute pancreatitis contributed to 6,000 deaths in 2019 (Table 184). Mortality was uncommon among the youngest age groups after which rates (underlying or other cause) increased with age. Age-adjusted mortality rates were higher among men, Blacks, and non-Hispanics.

**Table 184:** Acute Pancreatitis: Deaths with underlying or underlying/other cause and lifetime years of life lost by age, race, ethnicity, and sex in the United States, 2019.

| Demographic Characteristics |  | Underlying Cause |  |  | Underlying/Other Cause |  |  |
| --- | --- | --- | --- | --- | --- | --- | --- |
|  |  | Number of Deaths | Rate per 100,000 | Years of potential life lost in thousands | Number of Deaths | Rate per 100,000 | Years of potential life lost in thousands |
| Age (years) | 0 to 11 | 2 | 0.0 | 0.1 | 4 | 0.0 | 0.3 |
|  | 12 to 24 | 39 | 0.1 | 2.3 | 67 | 0.1 | 3.9 |
|  | 25 to 44 | 398 | 0.5 | 17.2 | 721 | 0.8 | 31.1 |
|  | 45 to 54 | 335 | 0.8 | 10.3 | 698 | 1.7 | 21.5 |
|  | 55 to 64 | 586 | 1.4 | 13.4 | 1,186 | 2.8 | 27.1 |
|  | 65 to 74 | 599 | 1.9 | 9.4 | 1,216 | 3.9 | 19.2 |
|  | 75 plus | 861 | 3.8 | 6.4 | 1,802 | 8.0 | 13.2 |
| Race | White | 2,368 | 0.8 | 48.2 | 4,724 | 1.5 | 94.0 |
|  | Black | 342 | 0.8 | 8.0 | 720 | 1.6 | 16.4 |
|  | Other | 110 | 0.4 | 2.8 | 250 | 1.0 | 5.8 |
| Ethnicity | Hispanic | 178 | 0.4 | 4.7 | 409 | 0.9 | 10.4 |
|  | Not Hispanic | 2,642 | 0.8 | 54.3 | 5,285 | 1.6 | 105.9 |
| Sex | Female | 1,120 | 0.5 | 21.3 | 2,314 | 1.1 | 44.0 |
|  | Male | 1,700 | 1.0 | 37.7 | 3,380 | 1.9 | 72.3 |
| Total |  | 2,820 | 0.7 | 59.0 | 5,694 | 1.5 | 116.3 |

Source: Vital Statistics of the United States.
Rates for age groups are unadjusted and all other rates are age-adjusted.
Years of potential life lost is based on U.S. life table 2006-2018 estimates of life expectancy at age of death.

Among privately insured enrollees, the claims-based prevalence of acute pancreatitis (based on all-listed diagnoses) was 0.2% (Table 185). Prevalence was uncommon among children and adolescents and the youngest adults and was highest among persons 55 years and over. It did not differ by sex and was highest among Blacks, similar among Whites and Hispanics, and lowest among Asians. It did not differ by region.

**Table 185:**
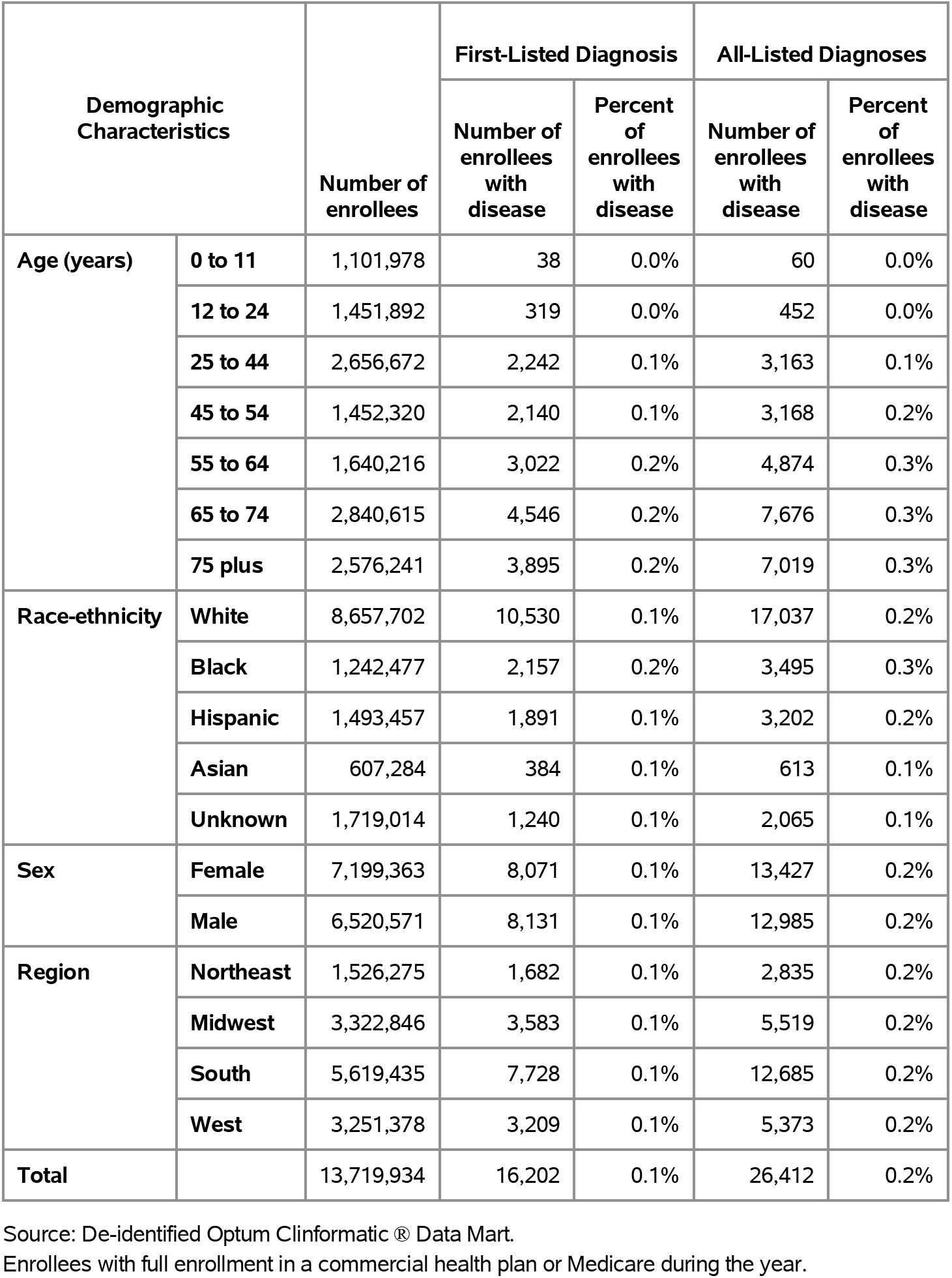
Acute Pancreatitis: Claims-based prevalence with first-listed and all-listed diagnoses by age, race-ethnicity, sex and region among privately insured enrollees, 2020.

Among commercial insurance enrollees, ambulatory care visit rates with acute pancreatitis (all-listed diagnoses) peaked among persons 55 to 64 years and were higher among men compared with women (Table 186). Among persons with known race-ethnicity, rates were highest among Blacks, followed by Whites and Hispanics, and lowest among Asians. Rates were higher in the South compared with the West, Northeast, and Midwest.

**Table 186:**
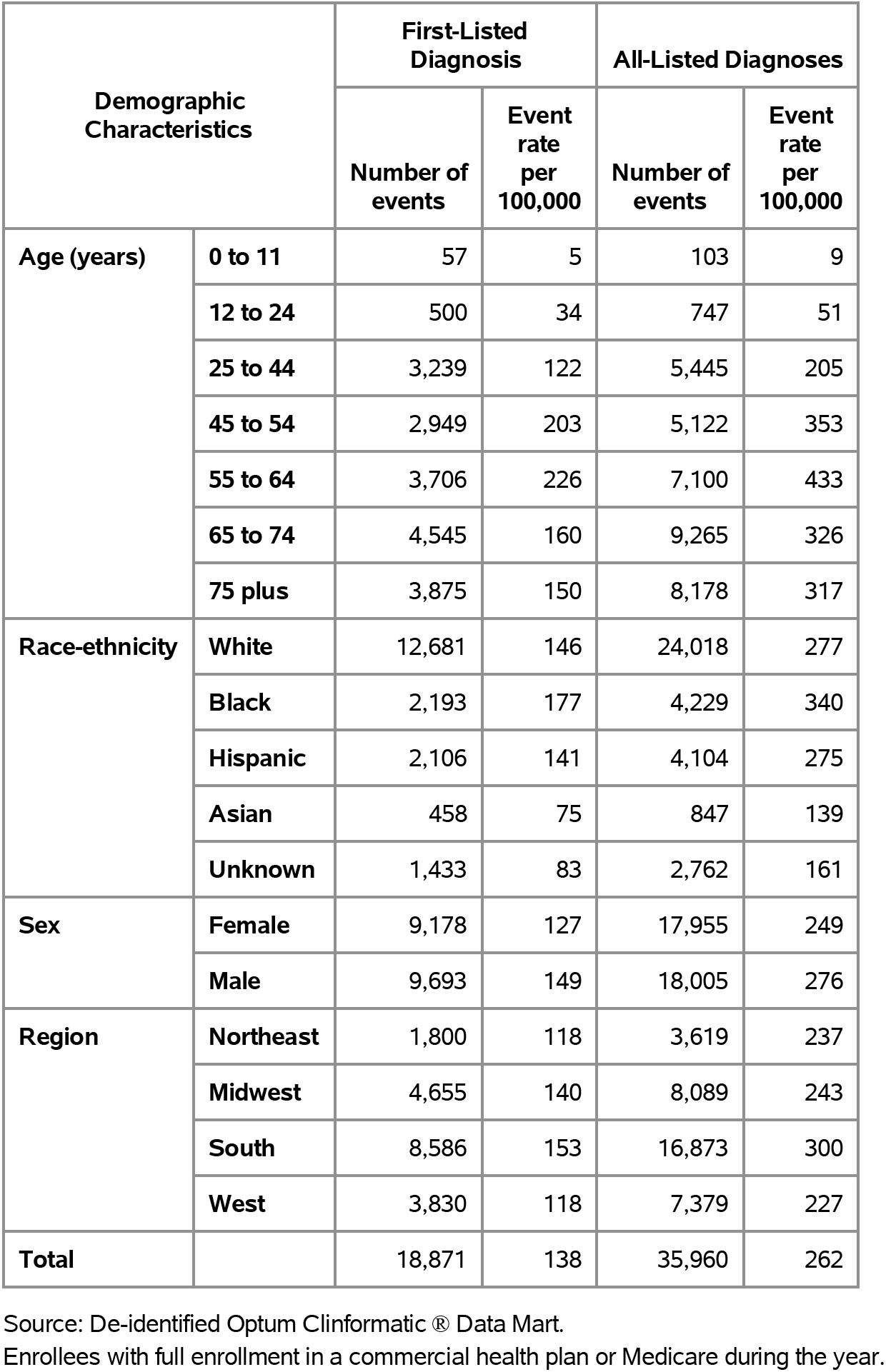
Acute Pancreatitis: Ambulatory care visits with first-listed and all-listed diagnoses by age, race-ethnicity, sex and region among privately insured enrollees, 2020.

| Demographic Characteristics |  | First-Listed Diagnosis |  | All-Listed Diagnoses |  |
| --- | --- | --- | --- | --- | --- |
|  |  | Number of events | Event rate per 100,000 | Number of events | Event rate per 100,000 |
| Age (years) | 0 to 11 | 57 | 5 | 103 | 9 |
|  | 12 to 24 | 500 | 34 | 747 | 51 |
|  | 25 to 44 | 3,239 | 122 | 5,445 | 205 |
|  | 45 to 54 | 2,949 | 203 | 5,122 | 353 |
|  | 55 to 64 | 3,706 | 226 | 7,100 | 433 |
|  | 65 to 74 | 4,545 | 160 | 9,265 | 326 |
|  | 75 plus | 3,875 | 150 | 8,178 | 317 |
| Race-ethnicity | White | 12,681 | 146 | 24,018 | 277 |
|  | Black | 2,193 | 177 | 4,229 | 340 |
|  | Hispanic | 2,106 | 141 | 4,104 | 275 |
|  | Asian | 458 | 75 | 847 | 139 |
|  | Unknown | 1,433 | 83 | 2,762 | 161 |
| Sex | Female | 9,178 | 127 | 17,955 | 249 |
|  | Male | 9,693 | 149 | 18,005 | 276 |
| Region | Northeast | 1,800 | 118 | 3,619 | 237 |
|  | Midwest | 4,655 | 140 | 8,089 | 243 |
|  | South | 8,586 | 153 | 16,873 | 300 |
|  | West | 3,830 | 118 | 7,379 | 227 |
| Total |  | 18,871 | 138 | 35,960 | 262 |

Source: De-identified Optum Clinformatics® Data Mart.
Enrollees with full enrollment in a commercial health plan or Medicare during the year.

Among commercial insurance enrollees, emergency department visit rates with acute pancreatitis (all-listed diagnoses) increased with age until 75 years and differed little by sex (Table 187). Among persons with known race-ethnicity, rates were highest among Blacks, followed by Hispanics and Whites, and lowest among Asians. Rates were highest in the South, followed by the Northeast, then the West, and lowest in the Midwest.

**Table 187:**
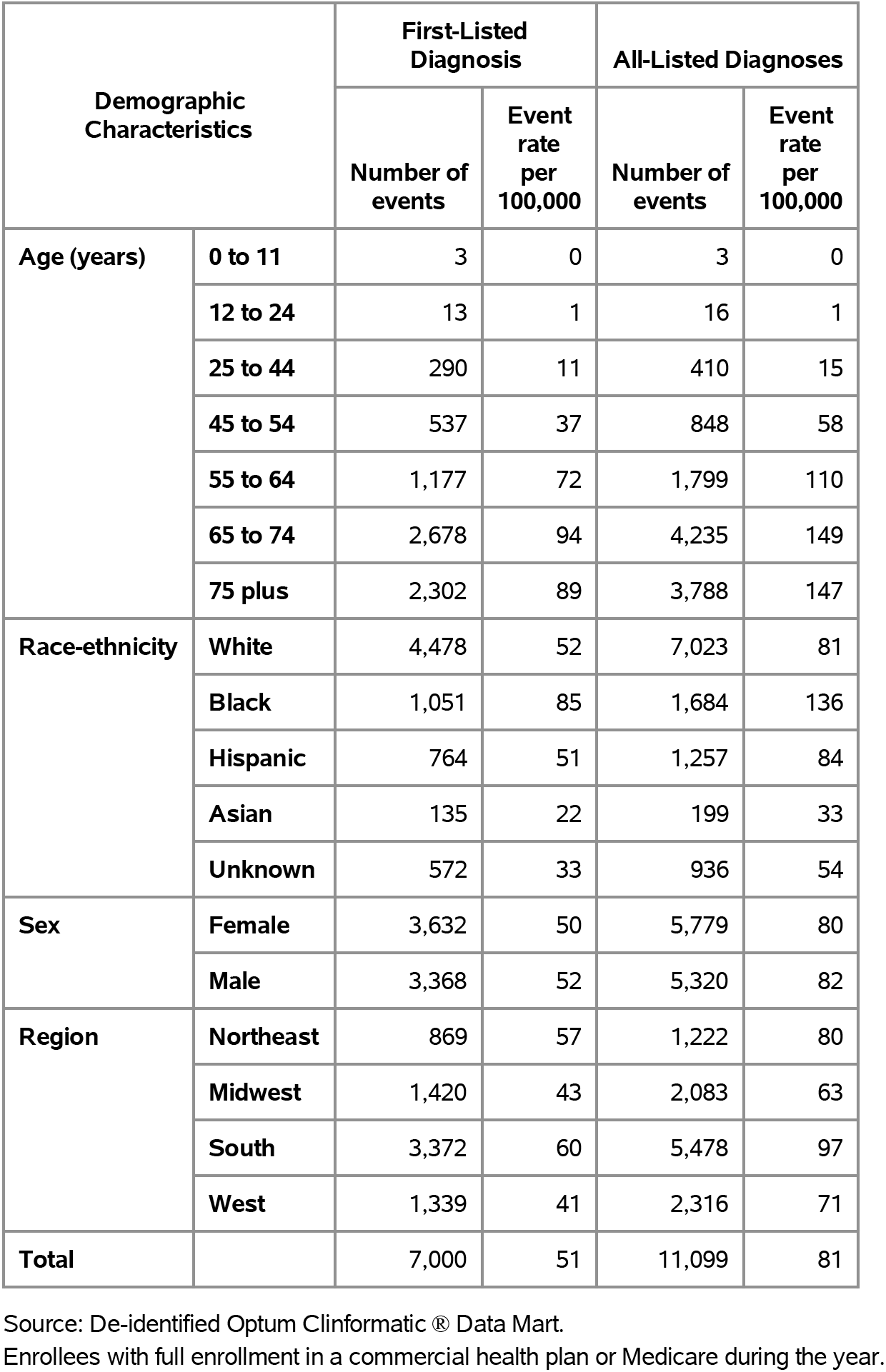
Acute Pancreatitis: Emergency department visits with first-listed and all-listed diagnoses by age, race-ethnicity, sex and region among privately insured enrollees, 2020.

| Demographic Characteristics |  | First-Listed Diagnosis |  | All-Listed Diagnoses |  |
| --- | --- | --- | --- | --- | --- |
|  |  | Number of events | Event rate per 100,000 | Number of events | Event rate per 100,000 |
| Age (years) | 0 to 11 | 3 | 0 | 3 | 0 |
|  | 12 to 24 | 13 | 1 | 16 | 1 |
|  | 25 to 44 | 290 | 11 | 410 | 15 |
|  | 45 to 54 | 537 | 37 | 848 | 58 |
|  | 55 to 64 | 1,177 | 72 | 1,799 | 110 |
|  | 65 to 74 | 2,678 | 94 | 4,235 | 149 |
|  | 75 plus | 2,302 | 89 | 3,788 | 147 |
| Race-ethnicity | White | 4,478 | 52 | 7,023 | 81 |
|  | Black | 1,051 | 85 | 1,684 | 136 |
|  | Hispanic | 764 | 51 | 1,257 | 84 |
|  | Asian | 135 | 22 | 199 | 33 |
|  | Unknown | 572 | 33 | 936 | 54 |
| Sex | Female | 3,632 | 50 | 5,779 | 80 |
|  | Male | 3,368 | 52 | 5,320 | 82 |
| Region | Northeast | 869 | 57 | 1,222 | 80 |
|  | Midwest | 1,420 | 43 | 2,083 | 63 |
|  | South | 3,372 | 60 | 5,478 | 97 |
|  | West | 1,339 | 41 | 2,316 | 71 |
| Total |  | 7,000 | 51 | 11,099 | 81 |

Source: De-identified Optum Clinformatic® Data Mart.
Enrollees with full enrollment in a commercial health plan or Medicare during the year.

Among commercial insurance enrollees, hospital discharge rates with acute pancreatitis (all-listed diagnoses) increased with age until 65 years and were higher among men compared with women (Table 188). Among persons with known race-ethnicity, rates were highest among Blacks, followed by Hispanics, then Whites, and lowest among Asians. Rates were highest in the South, followed by the Northeast, then the Midwest, and lowest in the West.

**Table 188:** Acute Pancreatitis: Hospital discharges with first-listed and all-listed diagnoses by age, race-ethnicity, sex and region among privately insured enrollees, 2020.

| Demographic Characteristics |  | First-Listed Diagnosis |  | All-Listed Diagnoses |  |
| --- | --- | --- | --- | --- | --- |
|  |  | Number of events | Event rate per 100,000 | Number of events | Event rate per 100,000 |
| Age (years) | 0 to 11 | 24 | 2 | 47 | 4 |
|  | 12 to 24 | 160 | 11 | 275 | 19 |
|  | 25 to 44 | 1,408 | 53 | 2,214 | 83 |
|  | 45 to 54 | 1,351 | 93 | 2,183 | 150 |
|  | 55 to 64 | 1,774 | 108 | 3,260 | 199 |
|  | 65 to 74 | 2,562 | 90 | 5,308 | 187 |
|  | 75 plus | 2,139 | 83 | 5,008 | 194 |
| Race-ethnicity | White | 6,064 | 70 | 11,760 | 136 |
|  | Black | 1,338 | 108 | 2,444 | 197 |
|  | Hispanic | 1,061 | 71 | 2,222 | 149 |
|  | Asian | 207 | 34 | 407 | 67 |
|  | Unknown | 748 | 44 | 1,462 | 85 |
| Sex | Female | 4,548 | 63 | 9,065 | 126 |
|  | Male | 4,870 | 75 | 9,230 | 142 |
| Region | Northeast | 948 | 62 | 2,008 | 132 |
|  | Midwest | 2,218 | 67 | 4,034 | 121 |
|  | South | 4,494 | 80 | 8,618 | 153 |
|  | West | 1,758 | 54 | 3,635 | 112 |
| Total |  | 9,418 | 69 | 18,295 | 133 |

Source: De-identified Optum Clinformatic® Data Mart.
Enrollees with full enrollment in a commercial health plan or Medicare during the year.

Among Medicare beneficiaries, the claims-based prevalence of acute pancreatitis (based on all-listed diagnoses) was 0.4% (Table 189). Prevalence differed little with age and was higher among men and Blacks. It differed little by region.

**Table 189:** Acute Pancreatitis: Claims-based prevalence with first-listed and all-listed diagnoses by age, race, sex and region among fee-for-service, age-eligible Medicare beneficiaries, 2019.

| Demographic Characteristics |  | Number of enrollees | First-Listed Diagnosis |  | All-Listed Diagnoses |  |
| --- | --- | --- | --- | --- | --- | --- |
|  |  |  | Number of enrollees with disease | Percent of enrollees with disease | Number of enrollees with disease | Percent of enrollees with disease |
| Age (years) | 65 to 69 | 8,066,320 | 16,500 | 0.2% | 26,020 | 0.3% |
|  | 70 to 74 | 6,936,940 | 15,740 | 0.2% | 25,420 | 0.4% |
|  | 75 to 79 | 4,894,060 | 9,940 | 0.2% | 16,660 | 0.3% |
|  | 80 to 84 | 3,335,340 | 7,680 | 0.2% | 13,300 | 0.4% |
|  | 85+ | 3,578,120 | 8,300 | 0.2% | 13,740 | 0.4% |
| Race | White | 22,023,800 | 47,040 | 0.2% | 75,520 | 0.3% |
|  | Black | 1,861,660 | 4,440 | 0.2% | 7,920 | 0.4% |
|  | Other | 2,925,320 | 6,680 | 0.2% | 11,700 | 0.4% |
| Sex | Female | 14,975,560 | 30,500 | 0.2% | 50,440 | 0.3% |
|  | Male | 11,835,220 | 27,660 | 0.2% | 44,700 | 0.4% |
| Region | Northeast | 4,748,700 | 8,820 | 0.2% | 14,740 | 0.3% |
|  | Midwest | 6,069,800 | 13,340 | 0.2% | 21,160 | 0.3% |
|  | South | 10,595,900 | 24,840 | 0.2% | 40,080 | 0.4% |
|  | West | 5,396,380 | 11,160 | 0.2% | 19,160 | 0.4% |
| Total |  | 26,810,780 | 58,160 | 0.2% | 95,140 | 0.4% |

Source: Centers for Medicare & Medicaid Services, 5% Denominator and Claims Files.
Beneficiaries aged 65+ years with continuous and full Parts A and B enrollment and no Health Maintenance Organization enrollment during the year. Unweighted counts were multiplied by 20 to produce counts for this table.

Among Medicare beneficiaries, ambulatory care visit rates with acute pancreatitis (all-listed diagnoses) were relatively stable with age and were higher among men compared with women and Whites compared with Blacks (Table 190). Rates were higher in the Midwest and South compared with the Northeast and West.

**Table 190:** Acute Pancreatitis: Ambulatory care visits with first-listed and all-listed diagnoses by age, race, sex and region among fee-for-service, age-eligible Medicare beneficiaries, 2019.

| Demographic Characteristics |  | First-Listed Diagnosis |  | All-Listed Diagnoses |  |
| --- | --- | --- | --- | --- | --- |
|  |  | Number of events | Event rate per 100,000 | Number of events | Event rate per 100,000 |
| Age (years) | 65 to 69 | 14,000 | 174 | 30,100 | 373 |
|  | 70 to 74 | 14,300 | 206 | 29,240 | 422 |
|  | 75 to 79 | 9,320 | 190 | 18,500 | 378 |
|  | 80 to 84 | 6,620 | 198 | 14,180 | 425 |
|  | 85+ | 7,920 | 221 | 14,780 | 413 |
| Race | White | 44,560 | 202 | 88,520 | 402 |
|  | Black | 2,500 | 134 | 6,360 | 342 |
|  | Other | 5,100 | 174 | 11,920 | 407 |
| Sex | Female | 25,920 | 173 | 53,540 | 358 |
|  | Male | 26,240 | 222 | 53,260 | 450 |
| Region | Northeast | 9,060 | 191 | 18,140 | 382 |
|  | Midwest | 12,320 | 203 | 25,220 | 415 |
|  | South | 21,280 | 201 | 43,080 | 407 |
|  | West | 9,500 | 176 | 20,360 | 377 |
| Total |  | 52,160 | 195 | 106,800 | 398 |

Source: Centers for Medicare & Medicaid Services, 5% Denominator and Claims Files. Beneficiaries aged 65+ years with continuous and full Parts A and B enrollment and no Health Maintenance Organization enrollment during the year. Unweighted counts were multiplied by 20 to produce counts for this table.

Among Medicare beneficiaries, emergency department visit rates with acute pancreatitis (all-listed diagnoses) generally increased with age and were higher among men compared with women and Blacks compared with Whites (Table 191). Rates were highest in the South, followed by the West, then the Midwest, and lowest in the Northeast.

**Table 191:** Acute Pancreatitis: Emergency department visits with first-listed and all-listed diagnoses by age, race, sex and region among fee-for-service, age-eligible Medicare beneficiaries, 2019.

| Demographic Characteristics |  | First-Listed Diagnosis |  | All-Listed Diagnoses |  |
| --- | --- | --- | --- | --- | --- |
|  |  | Number of events | Event rate per 100,000 | Number of events | Event rate per 100,000 |
| Age (years) | 65 to 69 | 15,480 | 192 | 23,280 | 289 |
|  | 70 to 74 | 13,920 | 201 | 21,100 | 304 |
|  | 75 to 79 | 8,920 | 182 | 14,560 | 298 |
|  | 80 to 84 | 6,900 | 207 | 11,640 | 349 |
|  | 85+ | 7,700 | 215 | 12,660 | 354 |
| Race | White | 42,480 | 193 | 65,660 | 298 |
|  | Black | 4,480 | 241 | 7,440 | 400 |
|  | Other | 5,960 | 204 | 10,140 | 347 |
| Sex | Female | 27,700 | 185 | 43,820 | 293 |
|  | Male | 25,220 | 213 | 39,420 | 333 |
| Region | Northeast | 8,060 | 170 | 12,620 | 266 |
|  | Midwest | 11,940 | 197 | 17,860 | 294 |
|  | South | 22,640 | 214 | 36,080 | 341 |
|  | West | 10,280 | 190 | 16,680 | 309 |
| Total |  | 52,920 | 197 | 83,240 | 310 |

Source: Centers for Medicare & Medicaid Services, 5% Denominator and Claims Files. Beneficiaries aged 65+ years with continuous and full Parts A and B enrollment and no Health Maintenance Organization enrollment during the year. Unweighted counts were multiplied by 20 to produce counts for this table.

Among Medicare beneficiaries, hospital discharge rates with acute pancreatitis (all-listed diagnoses) generally increased with age and were higher among men compared with women and Blacks compared with Whites (Table 192). Rates were highest in the South and Midwest, followed by the West, and lowest in the Northeast.

**Table 192:** Acute Pancreatitis: Hospital discharges with first-listed and all-listed diagnoses by age, race, sex and region among fee-for-service, age-eligible Medicare beneficiaries, 2019.

| Demographic Characteristics |  | First-Listed Diagnosis |  | All-Listed Diagnoses |  |
| --- | --- | --- | --- | --- | --- |
|  |  | Number of events | Event rate per 100,000 | Number of events | Event rate per 100,000 |
| Age (years) | 65 to 69 | 9,840 | 122 | 17,340 | 215 |
|  | 70 to 74 | 9,160 | 132 | 16,200 | 234 |
|  | 75 to 79 | 5,600 | 114 | 10,920 | 223 |
|  | 80 to 84 | 4,480 | 134 | 8,860 | 266 |
|  | 85+ | 5,360 | 150 | 10,420 | 291 |
| Race | White | 28,480 | 129 | 51,340 | 233 |
|  | Black | 2,460 | 132 | 4,900 | 263 |
|  | Other | 3,500 | 120 | 7,500 | 256 |
| Sex | Female | 17,760 | 119 | 33,120 | 221 |
|  | Male | 16,680 | 141 | 30,620 | 259 |
| Region | Northeast | 5,100 | 107 | 9,820 | 207 |
|  | Midwest | 8,660 | 143 | 15,020 | 247 |
|  | South | 14,700 | 139 | 26,540 | 250 |
|  | West | 5,980 | 111 | 12,360 | 229 |
| Total |  | 34,440 | 128 | 63,740 | 238 |

Source: Centers for Medicare & Medicaid Services, 5% Denominator and Claims Files. Beneficiaries aged 65+ years with continuous and full Parts A and B enrollment and no Health Maintenance Organization enrollment during the year. Unweighted counts were multiplied by 20 to produce counts for this table.

Chronic pancreatitis contributed to 245,000 ambulatory visits (2015) (Table 193). Ambulatory care visit rates (all-listed diagnoses) peaked in middle age. Age-adjusted rates were higher among men compared with women, Whites compared with Blacks, and Hispanics compared with non-Hispanics.

**Table 193:** Chronic Pancreatitis: Ambulatory care visits with first-listed and all-listed diagnoses by age, race, ethnicity, and sex in the United States, 2015.

| Demographic Characteristics |  | First-Listed Diagnosis |  | All-Listed Diagnoses |  |
| --- | --- | --- | --- | --- | --- |
|  |  | Number in thousands | Rate per 100,000 | Number in thousands | Rate per 100,000 |
| Age (years) | 0 to 11 | . | . | . | . |
|  | 12 to 24 | . | . | 3 | 5 |
|  | 25 to 44 | 16 | 19 | 54 | 64 |
|  | 45 to 54 | 40 | 92 | 86 | 199 |
|  | 55 to 64 | 9 | 22 | 49 | 119 |
|  | 65 to 74 | 31 | 114 | 36 | 129 |
|  | 75 plus | 1 | 7 | 19 | 93 |
| Race | White | 86 | 30 | 228 | 81 |
|  | Black | 11 | 27 | 16 | 37 |
|  | Other | . | . | 2 | 7 |
| Ethnicity | Hispanic | 74 | 168 | 91 | 206 |
|  | Not Hispanic | 23 | 9 | 154 | 52 |
| Sex | Female | 40 | 28 | 107 | 67 |
|  | Male | 57 | 30 | 138 | 80 |
| Total |  | 97 | 28 | 245 | 71 |

Source: National Ambulatory Medical Care Survey (3-year average, 2014-2016). Rates for age groups are unadjusted and all other rates are age-adjusted.

Chronic pancreatitis contributed to 263,000 emergency department visits in 2018 (Table 194). Emergency department visit rates (all-listed diagnoses) peaked in middle age. Age-adjusted rates were higher among men compared with women.

**Table 194:**
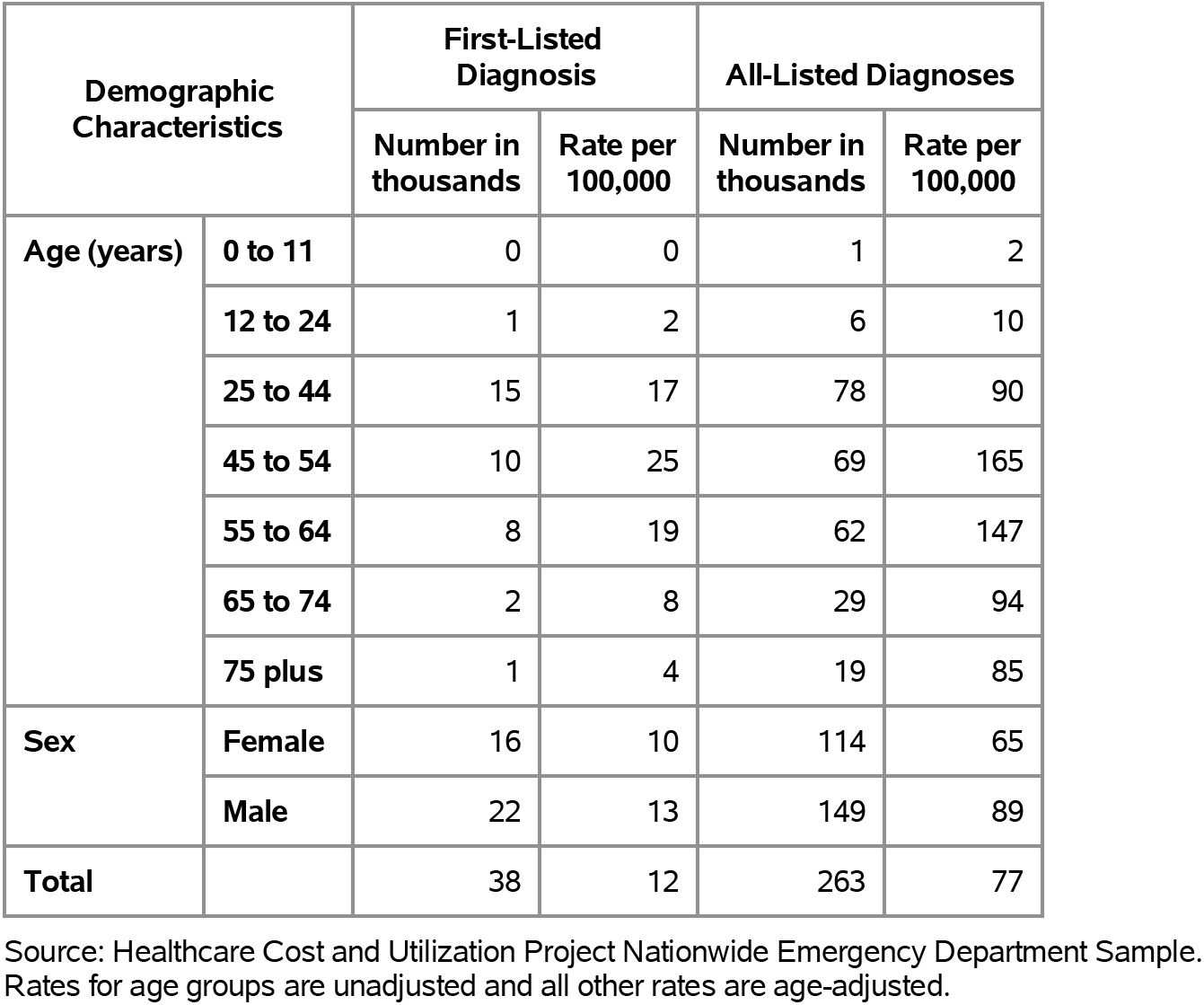
Chronic Pancreatitis: Emergency department visits with first-listed and all-listed diagnoses by age and sex in the United States, 2018.

| Demographic Characteristics |  | First-Listed Diagnosis |  | All-Listed Diagnoses |  |
| --- | --- | --- | --- | --- | --- |
|  |  | Number in thousands | Rate per 100,000 | Number in thousands | Rate per 100,000 |
| Age (years) | 0 to 11 | 0 | 0 | 1 | 2 |
|  | 12 to 24 | 1 | 2 | 6 | 10 |
|  | 25 to 44 | 15 | 17 | 78 | 90 |
|  | 45 to 54 | 10 | 25 | 69 | 165 |
|  | 55 to 64 | 8 | 19 | 62 | 147 |
|  | 65 to 74 | 2 | 8 | 29 | 94 |
|  | 75 plus | 1 | 4 | 19 | 85 |
| Sex | Female | 16 | 10 | 114 | 65 |
|  | Male | 22 | 13 | 149 | 89 |
| Total |  | 38 | 12 | 263 | 77 |

Source: Healthcare Cost and Utilization Project Nationwide Emergency Department Sample. Rates for age groups are unadjusted and all other rates are age-adjusted.

Chronic pancreatitis contributed to 192,000 hospital discharges in 2018 (Table 195). Hospital discharge rates (all-listed diagnoses) peaked in middle age. Age-adjusted rates were higher among men compared with women, Blacks compared with Whites, and non-Hispanics compared with Hispanics.

**Table 195:** Chronic Pancreatitis: Hospital discharges with first-listed and all-listed diagnoses by age, race, ethnicity, and sex in the United States, 2018.

| Demographic Characteristics |  | First-Listed Diagnosis |  | All-Listed Diagnoses |  |
| --- | --- | --- | --- | --- | --- |
|  |  | Number in thousands | Rate per 100,000 | Number in thousands | Rate per 100,000 |
| Age (years) | 0 to 11 | 0 | 0 | 1 | 2 |
|  | 12 to 24 | 1 | 1 | 5 | 8 |
|  | 25 to 44 | 3 | 4 | 51 | 58 |
|  | 45 to 54 | 3 | 7 | 47 | 112 |
|  | 55 to 64 | 2 | 5 | 47 | 112 |
|  | 65 to 74 | 1 | 4 | 25 | 83 |
|  | 75 plus | 1 | 3 | 17 | 76 |
| Race | White | 8 | 3 | 143 | 52 |
|  | Black | 2 | 5 | 42 | 93 |
|  | Other | 1 | 2 | 9 | 34 |
| Ethnicity | Hispanic | 1 | 2 | 18 | 35 |
|  | Not Hispanic | 10 | 4 | 176 | 60 |
| Sex | Female | 5 | 3 | 84 | 47 |
|  | Male | 6 | 3 | 109 | 64 |
| Total |  | 11 | 3 | 192 | 55 |

Source: Healthcare Cost and Utilization Project National Inpatient Sample. Rates for age groups are unadjusted and all other rates are age-adjusted.

Chronic pancreatitis contributed to 2,000 deaths in 2019 (Table 196). Mortality was uncommon among the youngest age groups after which rates (underlying or other cause) increased with age. Age-adjusted mortality rates were higher among men, Blacks, and non-Hispanics.

**Table 196:**
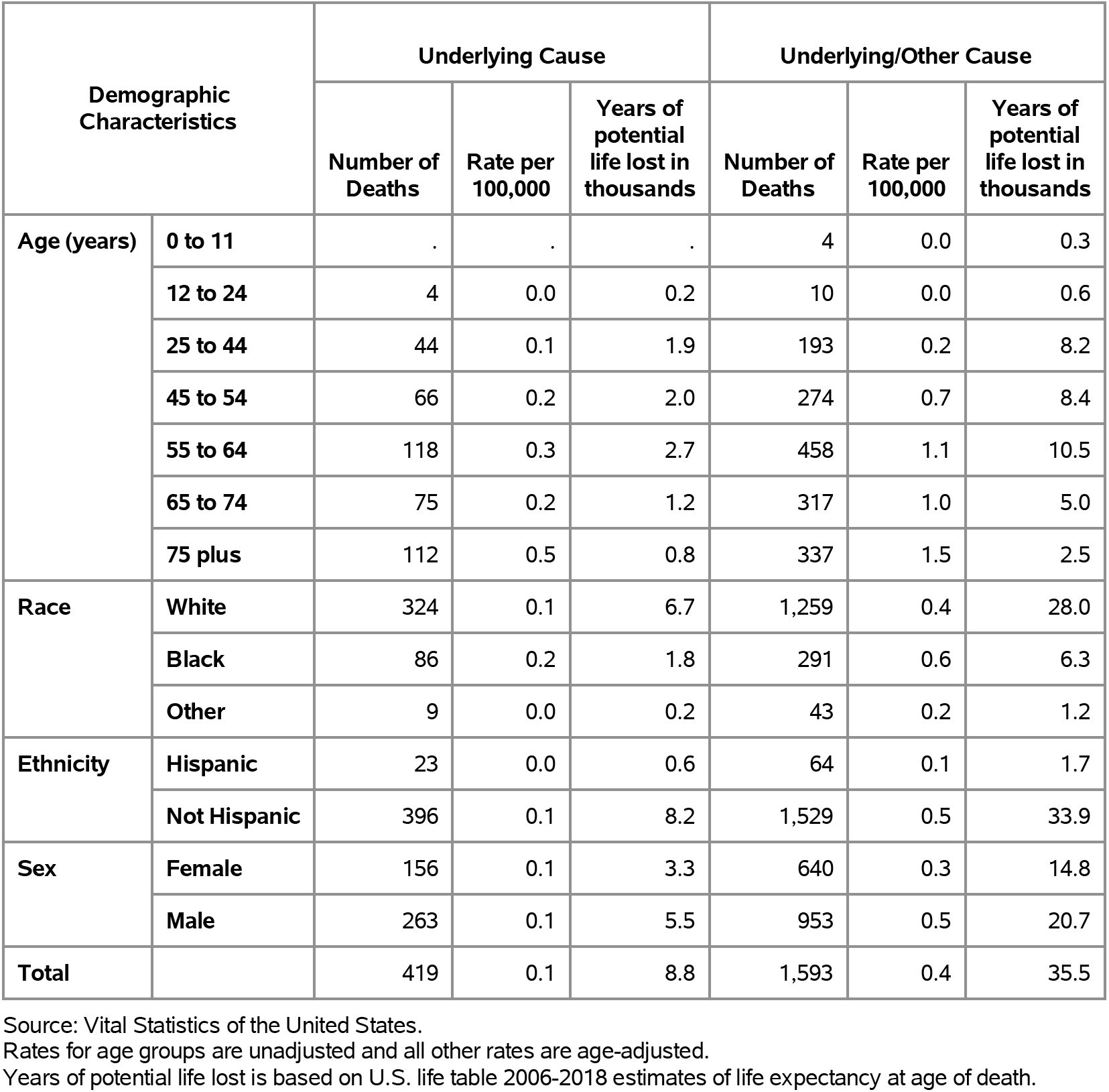
Chronic Pancreatitis: Deaths with underlying or underlying/other cause and lifetime years of life lost by age, race, ethnicity, and sex in the United States, 2019.

Among privately insured enrollees, the claims-based prevalence of chronic pancreatitis (based on all-listed diagnoses) was 0.1% (Table 197). Chronic pancreatitis was uncommon among children and adolescents and the youngest adults and prevalence was highest among persons 55 years and over. Prevalence did not differ by sex and was highest among Blacks and similar among Whites, Hispanics, and Asians. It did not differ by region.

**Table 197:** Chronic Pancreatitis: Claims-based prevalence with first-listed and all-listed diagnoses by age, race-ethnicity, sex and region among privately insured enrollees, 2020.

| Demographic Characteristics |  | Number of enrollees | First-Listed Diagnosis |  | All-Listed Diagnoses |  |
| --- | --- | --- | --- | --- | --- | --- |
|  |  |  | Number of enrollees with disease | Percent of enrollees with disease | Number of enrollees with disease | Percent of enrollees with disease |
| Age (years) | 0 to 11 | 1,101,978 | 9 | 0.0% | 15 | 0.0% |
|  | 12 to 24 | 1,451,892 | 54 | 0.0% | 110 | 0.0% |
|  | 25 to 44 | 2,656,672 | 484 | 0.0% | 1,168 | 0.0% |
|  | 45 to 54 | 1,452,320 | 669 | 0.0% | 1,704 | 0.1% |
|  | 55 to 64 | 1,640,216 | 1,157 | 0.1% | 3,288 | 0.2% |
|  | 65 to 74 | 2,840,615 | 1,530 | 0.1% | 4,823 | 0.2% |
|  | 75 plus | 2,576,241 | 963 | 0.0% | 4,092 | 0.2% |
| Race-ethnicity | White | 8,657,702 | 3,269 | 0.0% | 9,973 | 0.1% |
|  | Black | 1,242,477 | 705 | 0.1% | 2,201 | 0.2% |
|  | Hispanic | 1,493,457 | 404 | 0.0% | 1,520 | 0.1% |
|  | Asian | 607,284 | 131 | 0.0% | 368 | 0.1% |
|  | Unknown | 1,719,014 | 357 | 0.0% | 1,138 | 0.1% |
| Sex | Female | 7,199,363 | 2,364 | 0.0% | 7,456 | 0.1% |
|  | Male | 6,520,571 | 2,502 | 0.0% | 7,744 | 0.1% |
| Region | Northeast | 1,526,275 | 532 | 0.0% | 1,579 | 0.1% |
|  | Midwest | 3,322,846 | 1,101 | 0.0% | 3,151 | 0.1% |
|  | South | 5,619,435 | 2,312 | 0.0% | 7,271 | 0.1% |
|  | West | 3,251,378 | 921 | 0.0% | 3,199 | 0.1% |
| Total |  | 13,719,934 | 4,866 | 0.0% | 15,200 | 0.1% |

Source: De-identified Optum Clinformatic® Data Mart.
Enrollees with full enrollment in a commercial health plan or Medicare during the year.

Among commercial insurance enrollees, ambulatory care visit rates with chronic pancreatitis (all-listed diagnoses) peaked among persons 55 to 64 years and were higher among men compared with women (Table 198). Among persons with known race-ethnicity, rates were highest among Blacks, followed by Whites, then Hispanics, and lowest among Asians. Rates were higher in the South compared with the Northeast, West, and Midwest.

**Table 198:** Chronic Pancreatitis: Ambulatory care visits with first-listed and all-listed diagnoses by age, race-ethnicity, sex and region among privately insured enrollees, 2020.

| Demographic Characteristics |  | First-Listed Diagnosis |  | All-Listed Diagnoses |  |
| --- | --- | --- | --- | --- | --- |
|  |  | Number of events | Event rate per 100,000 | Number of events | Event rate per 100,000 |
| Age (years) | 0 to 11 | 9 | 1 | 25 | 2 |
|  | 12 to 24 | 84 | 6 | 211 | 15 |
|  | 25 to 44 | 1,006 | 38 | 2,884 | 109 |
|  | 45 to 54 | 1,429 | 98 | 4,514 | 311 |
|  | 55 to 64 | 2,269 | 138 | 7,853 | 479 |
|  | 65 to 74 | 2,453 | 86 | 9,100 | 320 |
|  | 75 plus | 1,468 | 57 | 6,576 | 255 |
| Race-ethnicity | White | 5,832 | 67 | 20,509 | 237 |
|  | Black | 1,253 | 101 | 4,667 | 376 |
|  | Hispanic | 711 | 48 | 2,906 | 195 |
|  | Asian | 242 | 40 | 703 | 116 |
|  | Unknown | 680 | 40 | 2,378 | 138 |
| Sex | Female | 4,342 | 60 | 15,938 | 221 |
|  | Male | 4,376 | 67 | 15,225 | 233 |
| Region | Northeast | 866 | 57 | 2,994 | 196 |
|  | Midwest | 2,044 | 62 | 6,668 | 201 |
|  | South | 4,187 | 75 | 14,983 | 267 |
|  | West | 1,621 | 50 | 6,518 | 200 |
| Total |  | 8,718 | 64 | 31,163 | 227 |

Source: De-identified Optum Clinformatic® Data Mart.
Enrollees with full enrollment in a commercial health plan or Medicare during the year.

Among commercial insurance enrollees, emergency department visit rates with chronic pancreatitis (all-listed diagnoses) peaked among persons 55 to 64 years and were higher among men compared with women (Table 199). Among persons with known race-ethnicity, rates were highest among Blacks, followed by Whites, then Hispanics, and lowest among Asians. Rates were highest in the South, followed by the Northeast, and lowest in the Midwest and West.

**Table 199:** Chronic Pancreatitis: Emergency department visits with first-listed and all-listed diagnoses by age, race-ethnicity, sex and region among privately insured enrollees, 2020.

| Demographic Characteristics |  | First-Listed Diagnosis |  | All-Listed Diagnoses |  |
| --- | --- | --- | --- | --- | --- |
|  |  | Number of events | Event rate per 100,000 | Number of events | Event rate per 100,000 |
| Age (years) | 0 to 11 | 1 | 0 | 1 | 0 |
|  | 12 to 24 | . | . | 1 | 0 |
|  | 25 to 44 | 58 | 2 | 119 | 4 |
|  | 45 to 54 | 114 | 8 | 264 | 18 |
|  | 55 to 64 | 202 | 12 | 455 | 28 |
|  | 65 to 74 | 155 | 5 | 395 | 14 |
|  | 75 plus | 50 | 2 | 177 | 7 |
| Race-ethnicity | White | 330 | 4 | 862 | 10 |
|  | Black | 146 | 12 | 294 | 24 |
|  | Hispanic | 41 | 3 | 116 | 8 |
|  | Asian | 5 | 1 | 14 | 2 |
|  | Unknown | 58 | 3 | 126 | 7 |
| Sex | Female | 249 | 3 | 631 | 9 |
|  | Male | 331 | 5 | 781 | 12 |
| Region | Northeast | 52 | 3 | 157 | 10 |
|  | Midwest | 121 | 4 | 270 | 8 |
|  | South | 335 | 6 | 760 | 14 |
|  | West | 72 | 2 | 225 | 7 |
| Total |  | 580 | 4 | 1,412 | 10 |

Source: De-identified Optum Clinformatic® Data Mart.
Enrollees with full enrollment in a commercial health plan or Medicare during the year.

Among commercial insurance enrollees, hospital discharge rates with chronic pancreatitis (all-listed diagnoses) peaked among persons 55 to 64 years and were higher among men compared with women (Table 200). Among persons with known race-ethnicity, rates were highest among Blacks, followed by Whites, then Hispanics, and lowest among Asians. Rates were highest in the South, followed by the Northeast and Midwest, and lowest in the West.

**Table 200:** Chronic Pancreatitis: Hospital discharges with first-listed and all-listed diagnoses by age, race-ethnicity, sex and region among privately insured enrollees, 2020.

| Demographic Characteristics |  | First-Listed Diagnosis |  | All-Listed Diagnoses |  |
| --- | --- | --- | --- | --- | --- |
|  |  | Number of events | Event rate per 100,000 | Number of events | Event rate per 100,000 |
| Age (years) | 0 to 11 | 1 | 0 | 9 | 1 |
|  | 12 to 24 | 4 | 0 | 58 | 4 |
|  | 25 to 44 | 34 | 1 | 746 | 28 |
|  | 45 to 54 | 51 | 4 | 1,046 | 72 |
|  | 55 to 64 | 78 | 5 | 1,893 | 115 |
|  | 65 to 74 | 75 | 3 | 2,211 | 78 |
|  | 75 plus | 22 | 1 | 1,611 | 63 |
| Race-ethnicity | White | 169 | 2 | 4,784 | 55 |
|  | Black | 54 | 4 | 1,369 | 110 |
|  | Hispanic | 23 | 2 | 700 | 47 |
|  | Asian | 3 | 0 | 117 | 19 |
|  | Unknown | 16 | 1 | 604 | 35 |
| Sex | Female | 140 | 2 | 3,521 | 49 |
|  | Male | 125 | 2 | 4,053 | 62 |
| Region | Northeast | 28 | 2 | 819 | 54 |
|  | Midwest | 62 | 2 | 1,726 | 52 |
|  | South | 125 | 2 | 3,708 | 66 |
|  | West | 50 | 2 | 1,321 | 41 |
| Total |  | 265 | 2 | 7,574 | 55 |

Source: De-identified Optum Clinformatic® Data Mart.
Enrollees with full enrollment in a commercial health plan or Medicare during the year.

Among Medicare beneficiaries, the claims-based prevalence of chronic pancreatitis (based on all-listed diagnoses) was 0.2% (Table 201). Prevalence did not differ by age or sex and was higher among Blacks. It did not differ by region.

**Table 201:** Chronic Pancreatitis: Claims-based prevalence with first-listed and all-listed diagnoses by age, race, sex and region among fee-for-service, age-eligible Medicare beneficiaries, 2019.

| Demographic Characteristics |  | Number of enrollees | First-Listed Diagnosis |  | All-Listed Diagnoses |  |
| --- | --- | --- | --- | --- | --- | --- |
|  |  |  | Number of enrollees with disease | Percent of enrollees with disease | Number of enrollees with disease | Percent of enrollees with disease |
| Age (years) | 65 to 69 | 8,066,320 | 5,540 | 0.1% | 16,180 | 0.2% |
|  | 70 to 74 | 6,936,940 | 4,700 | 0.1% | 13,000 | 0.2% |
|  | 75 to 79 | 4,894,060 | 2,940 | 0.1% | 9,740 | 0.2% |
|  | 80 to 84 | 3,335,340 | 1,780 | 0.1% | 6,740 | 0.2% |
|  | 85+ | 3,578,120 | 1,640 | 0.0% | 6,260 | 0.2% |
| Race | White | 22,023,800 | 13,880 | 0.1% | 41,980 | 0.2% |
|  | Black | 1,861,660 | 1,380 | 0.1% | 5,160 | 0.3% |
|  | Other | 2,925,320 | 1,340 | 0.0% | 4,780 | 0.2% |
| Sex | Female | 14,975,560 | 7,920 | 0.1% | 26,220 | 0.2% |
|  | Male | 11,835,220 | 8,680 | 0.1% | 25,700 | 0.2% |
| Region | Northeast | 4,748,700 | 2,600 | 0.1% | 9,020 | 0.2% |
|  | Midwest | 6,069,800 | 3,240 | 0.1% | 11,400 | 0.2% |
|  | South | 10,595,900 | 7,660 | 0.1% | 22,200 | 0.2% |
|  | West | 5,396,380 | 3,100 | 0.1% | 9,300 | 0.2% |
| Total |  | 26,810,780 | 16,600 | 0.1% | 51,920 | 0.2% |

Source: Centers for Medicare & Medicaid Services, 5% Denominator and Claims Files.
Beneficiaries aged 65+ years with continuous and full Parts A and B enrollment and no Health Maintenance Organization enrollment during the year. Unweighted counts were multiplied by 20 to produce counts for this table.

Among Medicare beneficiaries, ambulatory care visit rates with chronic pancreatitis (all-listed diagnoses) were generally stable before declining after 85 years and were higher among men compared with women and Blacks compared with Whites (Table 202). Rates were highest in the South, followed by the Midwest, and lowest in the West and Northeast.

**Table 202:**
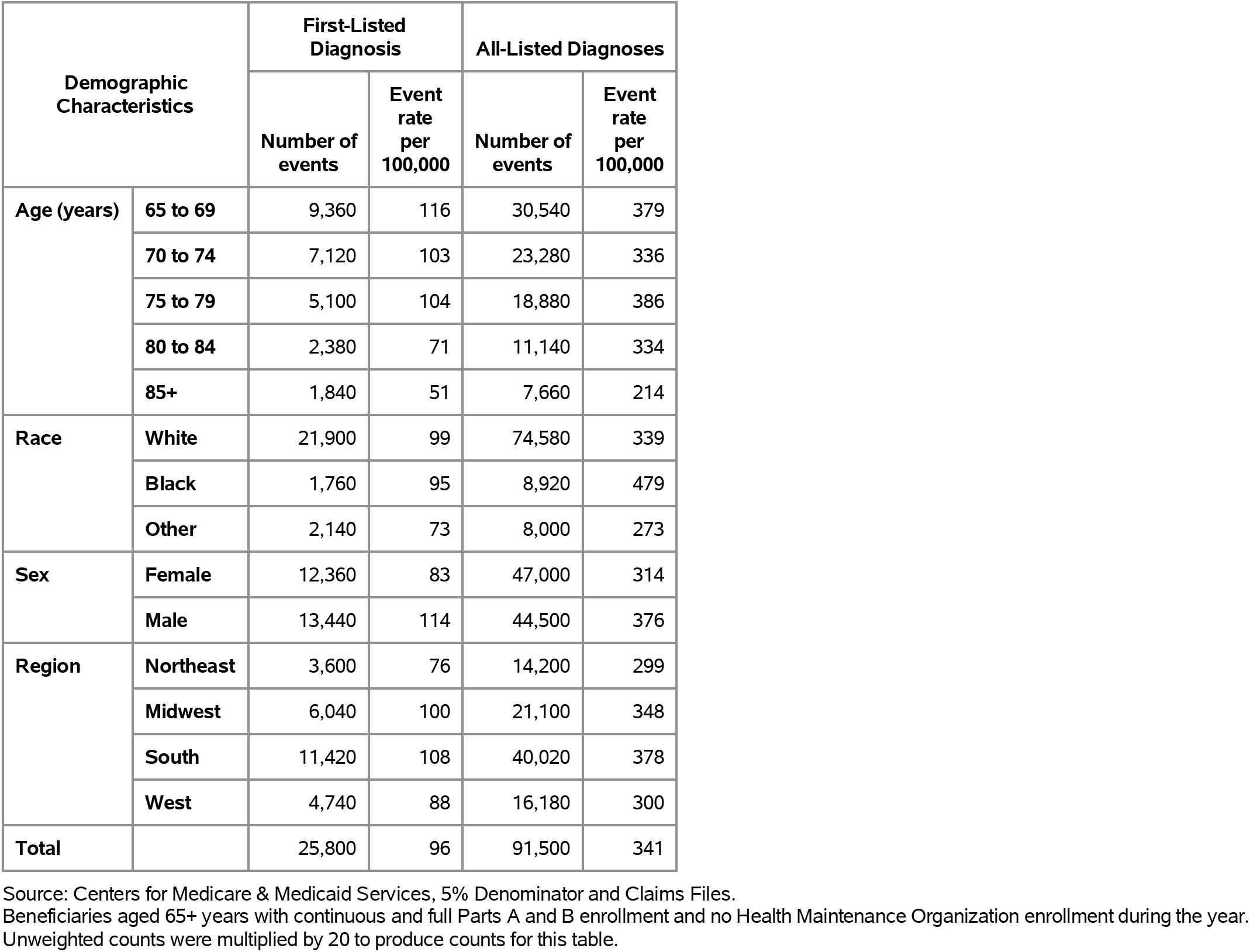
Chronic Pancreatitis: Ambulatory care visits with first-listed and all-listed diagnoses by age, race, sex and region among fee-for-service, age-eligible Medicare beneficiaries, 2019.

| Demographic Characteristics |  | First-Listed Diagnosis |  | All-Listed Diagnoses |  |
| --- | --- | --- | --- | --- | --- |
|  |  | Number of events | Event rate per 100,000 | Number of events | Event rate per 100,000 |
| Age (years) | 65 to 69 | 9,360 | 116 | 30,540 | 379 |
|  | 70 to 74 | 7,120 | 103 | 23,280 | 336 |
|  | 75 to 79 | 5,100 | 104 | 18,880 | 386 |
|  | 80 to 84 | 2,380 | 71 | 11,140 | 334 |
|  | 85+ | 1,840 | 51 | 7,660 | 214 |
| Race | White | 21,900 | 99 | 74,580 | 339 |
|  | Black | 1,760 | 95 | 8,920 | 479 |
|  | Other | 2,140 | 73 | 8,000 | 273 |
| Sex | Female | 12,360 | 83 | 47,000 | 314 |
|  | Male | 13,440 | 114 | 44,500 | 376 |
| Region | Northeast | 3,600 | 76 | 14,200 | 299 |
|  | Midwest | 6,040 | 100 | 21,100 | 348 |
|  | South | 11,420 | 108 | 40,020 | 378 |
|  | West | 4,740 | 88 | 16,180 | 300 |
| Total |  | 25,800 | 96 | 91,500 | 341 |

Source: Centers for Medicare & Medicaid Services, 5% Denominator and Claims Files. Beneficiaries aged 65+ years with continuous and full Parts A and B enrollment and no Health Maintenance Organization enrollment during the year. Unweighted counts were multiplied by 20 to produce counts for this table.

Among Medicare beneficiaries, emergency department visit rates with chronic pancreatitis (all-listed diagnoses) were highest among persons 65 to 69 years and were higher among men compared with women and over twice as high among Blacks compared with Whites (Table 203). Rates were highest in the Midwest, followed by the South, then the West, and lowest in the Northeast.

**Table 203:**
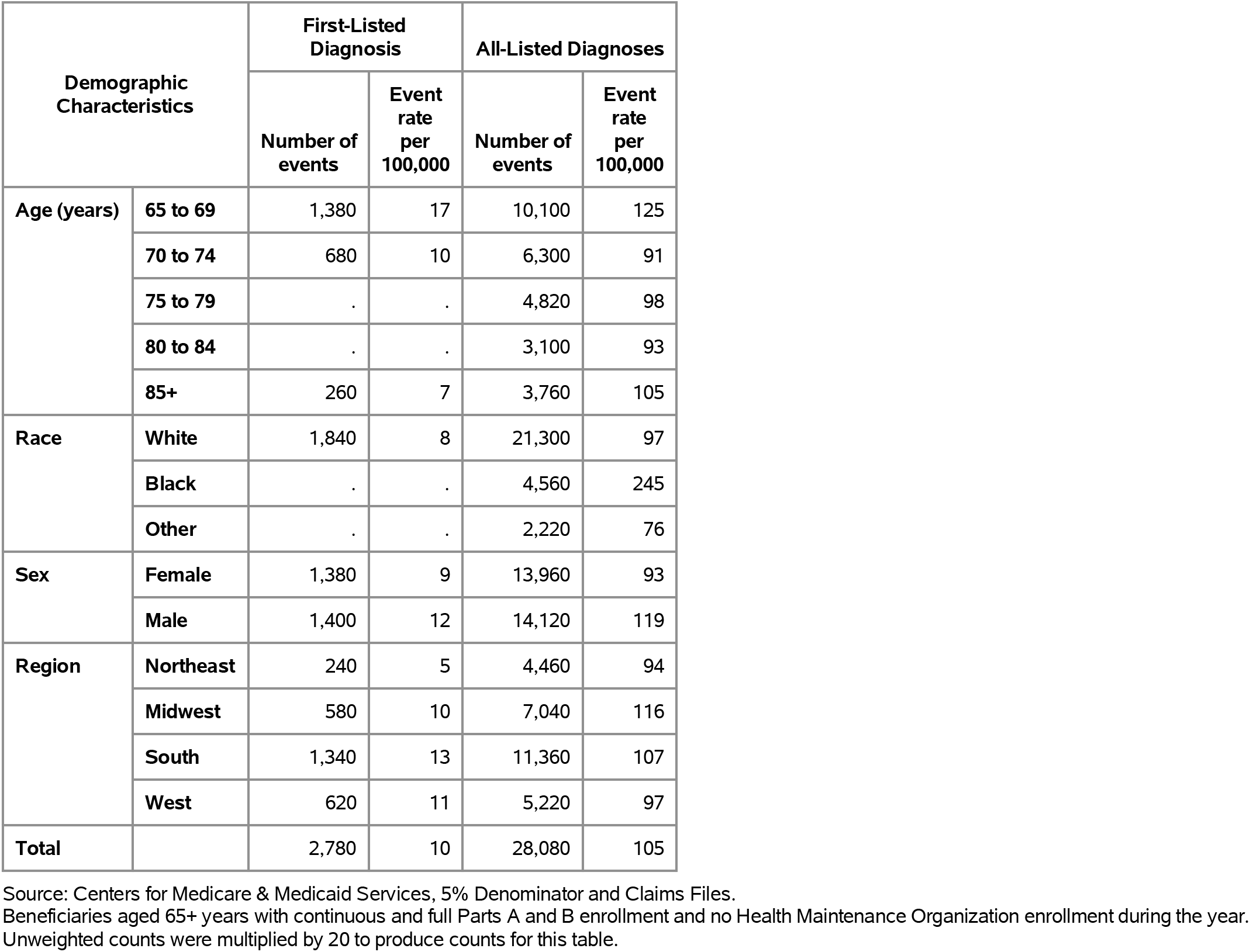
Chronic Pancreatitis: Emergency department visits with first-listed and all-listed diagnoses by age, race, sex and region among fee-for-service, age-eligible Medicare beneficiaries, 2019.

| Demographic Characteristics |  | First-Listed Diagnosis |  | All-Listed Diagnoses |  |
| --- | --- | --- | --- | --- | --- |
|  |  | Number of events | Event rate per 100,000 | Number of events | Event rate per 100,000 |
| Age (years) | 65 to 69 | 1,380 | 17 | 10,100 | 125 |
|  | 70 to 74 | 680 | 10 | 6,300 | 91 |
|  | 75 to 79 | . | . | 4,820 | 98 |
|  | 80 to 84 | . | . | 3,100 | 93 |
|  | 85+ | 260 | 7 | 3,760 | 105 |
| Race | White | 1,840 | 8 | 21,300 | 97 |
|  | Black | . | . | 4,560 | 245 |
|  | Other | . | . | 2,220 | 76 |
| Sex | Female | 1,380 | 9 | 13,960 | 93 |
|  | Male | 1,400 | 12 | 14,120 | 119 |
| Region | Northeast | 240 | 5 | 4,460 | 94 |
|  | Midwest | 580 | 10 | 7,040 | 116 |
|  | South | 1,340 | 13 | 11,360 | 107 |
|  | West | 620 | 11 | 5,220 | 97 |
| Total |  | 2,780 | 10 | 28,080 | 105 |

Source: Centers for Medicare & Medicaid Services, 5% Denominator and Claims Files.
Beneficiaries aged 65+ years with continuous and full Parts A and B enrollment and no Health Maintenance Organization enrollment during the year. Unweighted counts were multiplied by 20 to produce counts for this table.

Among Medicare beneficiaries, hospital discharge rates with chronic pancreatitis (all-listed diagnoses) were highest among persons 65 to 69 years and were higher among men compared with women and over twice as high among Blacks compared with Whites (Table 204). Rates were highest in the Midwest, followed by the South, then the Northeast, and lowest in the West.

**Table 204:** Chronic Pancreatitis: Hospital discharges with first-listed and all-listed diagnoses by age, race, sex and region among fee-for-service, age-eligible Medicare beneficiaries, 2019.

| Demographic Characteristics |  | First-Listed Diagnosis |  | All-Listed Diagnoses |  |
| --- | --- | --- | --- | --- | --- |
|  |  | Number of events | Event rate per 100,000 | Number of events | Event rate per 100,000 |
| Age (years) | 65 to 69 | 360 | 4 | 8,280 | 103 |
|  | 70 to 74 | 280 | 4 | 5,940 | 86 |
|  | 75 to 79 | . | . | 4,800 | 98 |
|  | 80 to 84 | . | . | 2,480 | 74 |
|  | 85+ | . | . | 3,000 | 84 |
| Race | White | 640 | 3 | 19,100 | 87 |
|  | Black | . | . | 3,520 | 189 |
|  | Other | . | . | 1,880 | 64 |
| Sex | Female | 540 | 4 | 12,340 | 82 |
|  | Male | 340 | 3 | 12,160 | 103 |
| Region | Northeast | . | . | 3,880 | 82 |
|  | Midwest | . | . | 6,480 | 107 |
|  | South | 440 | 4 | 10,040 | 95 |
|  | West | . | . | 4,100 | 76 |
| Total |  | 880 | 3 | 24,500 | 91 |

Source: Centers for Medicare & Medicaid Services, 5% Denominator and Claims Files. Beneficiaries aged 65+ years with continuous and full Parts A and B enrollment and no Health Maintenance Organization enrollment during the year. Unweighted counts were multiplied by 20 to produce counts for this table.

Celiac disease contributed to 611,000 ambulatory visits (2015) (Table 205). Ambulatory care visit rates (all-listed diagnoses) were highest among persons 55-64 years and second highest among children. Age-adjusted rates were higher among women compared with men, Whites compared with Blacks, and Hispanics compared with non-Hispanics.

**Table 205:** Celiac Disease: Ambulatory care visits with first-listed and all-listed diagnoses by age, race, ethnicity, and sex in the United States, 2015.

| Demographic Characteristics |  | First-Listed Diagnosis |  | All-Listed Diagnoses |  |
| --- | --- | --- | --- | --- | --- |
|  |  | Number in thousands | Rate per 100,000 | Number in thousands | Rate per 100,000 |
| Age (years) | 0 to 11 | 5 | 9 | 122 | 251 |
|  | 12 to 24 | 11 | 19 | 37 | 65 |
|  | 25 to 44 | 2 | 2 | 138 | 164 |
|  | 45 to 54 | 18 | 42 | 80 | 186 |
|  | 55 to 64 | 13 | 31 | 192 | 470 |
|  | 65 to 74 | 8 | 30 | 19 | 70 |
|  | 75 plus | 7 | 35 | 22 | 109 |
| Race | White | 63 | 23 | 507 | 199 |
|  | Black | . | . | 44 | 80 |
|  | Other | . | . | 59 | 257 |
| Ethnicity | Hispanic | 3 | 6 | 118 | 195 |
|  | Not Hispanic | 60 | 21 | 493 | 180 |
| Sex | Female | 47 | 27 | 390 | 247 |
|  | Male | 16 | 10 | 220 | 126 |
| Total |  | 63 | 18 | 611 | 187 |

Source: National Ambulatory Medical Care Survey (3-year average, 2014-2016). Rates for age groups are unadjusted and all other rates are age-adjusted.

Celiac disease contributed to 68,000 emergency department visits in 2018 (Table 206). Emergency department visit rates (all-listed diagnoses) increased with age. Age-adjusted rates were higher among women compared with men.

**Table 206:** Celiac Disease: Emergency department visits with first-listed and all-listed diagnoses by age and sex in the United States, 2018.

| Demographic Characteristics |  | First-Listed Diagnosis |  | All-Listed Diagnoses |  |
| --- | --- | --- | --- | --- | --- |
|  |  | Number in thousands | Rate per 100,000 | Number in thousands | Rate per 100,000 |
| Age (years) | 0 to 11 | 0 | 0 | 3 | 5 |
|  | 12 to 24 | 0 | 1 | 10 | 18 |
|  | 25 to 44 | 0 | 0 | 18 | 21 |
|  | 45 to 54 | 0 | 0 | 9 | 21 |
|  | 55 to 64 | 0 | 0 | 9 | 22 |
|  | 65 to 74 | 0 | 0 | 8 | 26 |
|  | 75 plus | 0 | 1 | 11 | 51 |
| Sex | Female | 1 | 1 | 50 | 29 |
|  | Male | 0 | 0 | 18 | 11 |
| Total |  | 1 | 0 | 68 | 20 |

Source: Healthcare Cost and Utilization Project Nationwide Emergency Department Sample. Rates for age groups are unadjusted and all other rates are age-adjusted.

Celiac disease contributed to 42,000 hospital discharges in 2018 (Table 207). Hospital discharge rates (all-listed diagnoses) increased with age. Age-adjusted rates were higher among women compared with men, Whites compared with Blacks, and non-Hispanics compared with Hispanics.

**Table 207:** Celiac Disease: Hospital discharges with first-listed and all-listed diagnoses by age, race, ethnicity, and sex in the United States, 2018.

| Demographic Characteristics |  | First-Listed Diagnosis |  | All-Listed Diagnoses |  |
| --- | --- | --- | --- | --- | --- |
|  |  | Number in thousands | Rate per 100,000 | Number in thousands | Rate per 100,000 |
| Age (years) | 0 to 11 | 0 | 0 | 1 | 2 |
|  | 12 to 24 | 0 | 0 | 4 | 8 |
|  | 25 to 44 | 0 | 0 | 10 | 11 |
|  | 45 to 54 | 0 | 0 | 5 | 12 |
|  | 55 to 64 | 0 | 0 | 6 | 15 |
|  | 65 to 74 | 0 | 0 | 7 | 23 |
|  | 75 plus | 0 | 0 | 9 | 41 |
| Race | White | 1 | 0 | 40 | 14 |
|  | Black | 0 | 0 | 2 | 3 |
|  | Other | 0 | 0 | 1 | 4 |
| Ethnicity | Hispanic | 0 | 0 | 2 | 4 |
|  | Not Hispanic | 1 | 0 | 40 | 13 |
| Sex | Female | 0 | 0 | 30 | 16 |
|  | Male | 0 | 0 | 13 | 7 |
| Total |  | 1 | 0 | 42 | 12 |

Source: Healthcare Cost and Utilization Project National Inpatient Sample. Rates for age groups are unadjusted and all other rates are age-adjusted.

Celiac disease contributed to <1,000 deaths in 2019 (Table 208). Mortality was uncommon with the highest rate (underlying or other cause) among persons 75 years and over. Age-adjusted mortality rates were higher among women, Whites, and non-Hispanics.

**Table 208:** Celiac Disease: Deaths with underlying or underlying/other cause and lifetime years of life lost by age, race, ethnicity, and sex in the United States, 2019.

| Demographic Characteristics |  | Underlying Cause |  |  | Underlying/Other Cause |  |  |
| --- | --- | --- | --- | --- | --- | --- | --- |
|  |  | Number of Deaths | Rate per 100,000 | Years of potential life lost in thousands | Number of Deaths | Rate per 100,000 | Years of potential life lost in thousands |
| Age (years) | 0 to 11 | . | . | . | 1 | 0.0 | 0.1 |
|  | 12 to 24 | . | . | . | . | . | . |
|  | 25 to 44 | 2 | 0.0 | 0.1 | 4 | 0.0 | 0.2 |
|  | 45 to 54 | 1 | 0.0 | 0.0 | 5 | 0.0 | 0.2 |
|  | 55 to 64 | 3 | 0.0 | 0.1 | 26 | 0.1 | 0.6 |
|  | 65 to 74 | 10 | 0.0 | 0.2 | 44 | 0.1 | 0.7 |
|  | 75 plus | 28 | 0.1 | 0.2 | 128 | 0.6 | 0.8 |
| Race | White | 42 | 0.0 | 0.5 | 202 | 0.1 | 2.4 |
|  | Black | 1 | 0.0 | 0.0 | 2 | 0.0 | 0.0 |
|  | Other | 1 | 0.0 | 0.0 | 4 | 0.0 | 0.1 |
| Ethnicity | Hispanic | 2 | 0.0 | 0.1 | 6 | 0.0 | 0.1 |
|  | Not Hispanic | 42 | 0.0 | 0.5 | 202 | 0.1 | 2.4 |
| Sex | Female | 25 | 0.0 | 0.3 | 130 | 0.1 | 1.5 |
|  | Male | 19 | 0.0 | 0.3 | 78 | 0.0 | 1.1 |
| Total |  | 44 | 0.0 | 0.6 | 208 | 0.1 | 2.6 |

Source: Vital Statistics of the United States.
Rates for age groups are unadjusted and all other rates are age-adjusted.
Years of potential life lost is based on U.S. life table 2006-2018 estimates of life expectancy at age of death.

Among privately insured enrollees, the claims-based prevalence of celiac disease (based on all-listed diagnoses) was 0.1% (Table 209). Prevalence was highest among adolescents and the youngest adults and among women. It was highest among Whites, similar among Blacks and Hispanics, and lowest among Asians. It differed little by region.

**Table 209:** Celiac Disease: Claims-based prevalence with first-listed and all-listed diagnoses by age, race-ethnicity, sex and region among privately insured enrollees, 2020.

| Demographic Characteristics |  | Number of enrollees | First-Listed Diagnosis |  | All-Listed Diagnoses |  |
| --- | --- | --- | --- | --- | --- | --- |
|  |  |  | Number of enrollees with disease | Percent of enrollees with disease | Number of enrollees with disease | Percent of enrollees with disease |
| Age (years) | 0 to 11 | 1,101,978 | 492 | 0.0% | 904 | 0.1% |
|  | 12 to 24 | 1,451,892 | 1,012 | 0.1% | 2,476 | 0.2% |
|  | 25 to 44 | 2,656,672 | 1,072 | 0.0% | 3,555 | 0.1% |
|  | 45 to 54 | 1,452,320 | 554 | 0.0% | 2,163 | 0.1% |
|  | 55 to 64 | 1,640,216 | 564 | 0.0% | 2,299 | 0.1% |
|  | 65 to 74 | 2,840,615 | 954 | 0.0% | 3,555 | 0.1% |
|  | 75 plus | 2,576,241 | 587 | 0.0% | 2,909 | 0.1% |
| Race-ethnicity | White | 8,657,702 | 4,083 | 0.0% | 14,136 | 0.2% |
|  | Black | 1,242,477 | 203 | 0.0% | 710 | 0.1% |
|  | Hispanic | 1,493,457 | 351 | 0.0% | 1,178 | 0.1% |
|  | Asian | 607,284 | 89 | 0.0% | 298 | 0.0% |
|  | Unknown | 1,719,014 | 509 | 0.0% | 1,539 | 0.1% |
| Sex | Female | 7,199,363 | 3,588 | 0.0% | 12,546 | 0.2% |
|  | Male | 6,520,571 | 1,647 | 0.0% | 5,315 | 0.1% |
| Region | Northeast | 1,526,275 | 976 | 0.1% | 2,948 | 0.2% |
|  | Midwest | 3,322,846 | 1,353 | 0.0% | 4,605 | 0.1% |
|  | South | 5,619,435 | 1,710 | 0.0% | 5,957 | 0.1% |
|  | West | 3,251,378 | 1,196 | 0.0% | 4,351 | 0.1% |
| Total |  | 13,719,934 | 5,235 | 0.0% | 17,861 | 0.1% |

Source: De-identified Optum Clinformatic® Data Mart.
Enrollees with full enrollment in a commercial health plan or Medicare during the year.

Among commercial insurance enrollees, ambulatory care visit rates with celiac disease (all-listed diagnoses) were higher among adolescents and the youngest adults compared with other age groups and were over twice as high among women compared with men (Table 210). Among persons with known race-ethnicity, rates were highest among Whites, followed by Hispanics, then Blacks, and lowest among Asians. Rates were highest in the Northeast, followed by the West and Midwest, and lowest in the South.

**Table 210:** Celiac Disease: Ambulatory care visits with first-listed and all-listed diagnoses by age, race-ethnicity, sex and region among privately insured enrollees, 2020.

| Demographic Characteristics |  | First-Listed Diagnosis |  | All-Listed Diagnoses |  |
| --- | --- | --- | --- | --- | --- |
|  |  | Number of events | Event rate per 100,000 | Number of events | Event rate per 100,000 |
| Age (years) | 0 to 11 | 710 | 64 | 1,666 | 151 |
|  | 12 to 24 | 1,405 | 97 | 4,473 | 308 |
|  | 25 to 44 | 1,448 | 55 | 5,917 | 223 |
|  | 45 to 54 | 713 | 49 | 3,727 | 257 |
|  | 55 to 64 | 743 | 45 | 3,947 | 241 |
|  | 65 to 74 | 1,296 | 46 | 6,340 | 223 |
|  | 75 plus | 786 | 31 | 5,350 | 208 |
| Race-ethnicity | White | 5,516 | 64 | 24,758 | 286 |
|  | Black | 280 | 23 | 1,268 | 102 |
|  | Hispanic | 488 | 33 | 2,251 | 151 |
|  | Asian | 128 | 21 | 460 | 76 |
|  | Unknown | 689 | 40 | 2,683 | 156 |
| Sex | Female | 4,857 | 67 | 22,158 | 308 |
|  | Male | 2,244 | 34 | 9,262 | 142 |
| Region | Northeast | 1,328 | 87 | 5,253 | 344 |
|  | Midwest | 1,798 | 54 | 7,704 | 232 |
|  | South | 2,338 | 42 | 10,788 | 192 |
|  | West | 1,637 | 50 | 7,675 | 236 |
| Total |  | 7,101 | 52 | 31,420 | 229 |

Source: De-identified Optum Clinformatic® Data Mart.
Enrollees with full enrollment in a commercial health plan or Medicare during the year.

Among commercial insurance enrollees, emergency department visit rates with celiac disease (all-listed diagnoses) were highest among persons 25 to 44 years and differed little by sex (Table 211). Among persons with known race-ethnicity, rates were higher among Whites compared with other race-ethnicities. Rates were highest in the Northeast, followed by the Midwest and West, and lowest in the South.

**Table 211:**
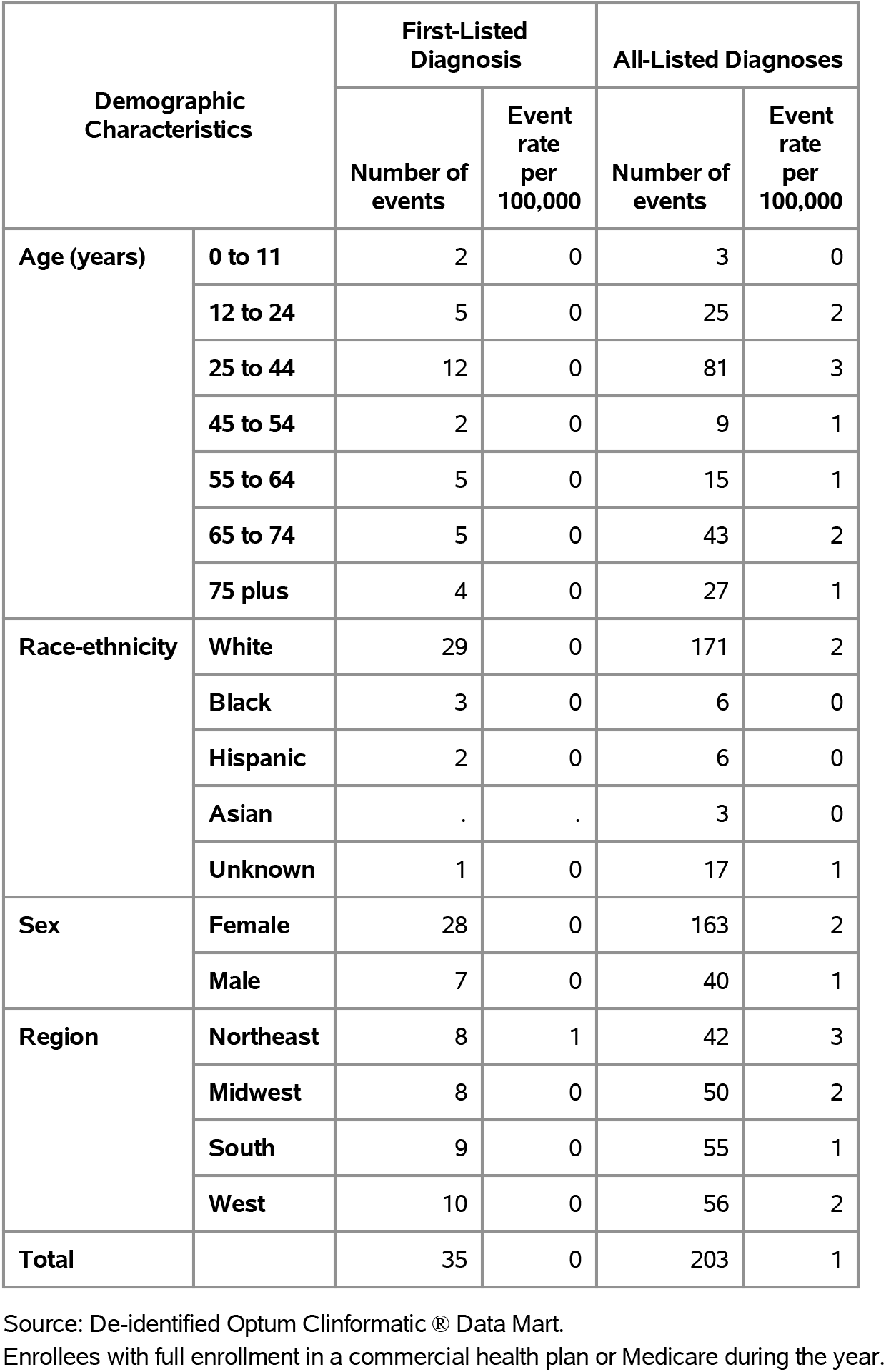
Celiac Disease: Emergency department visits with first-listed and all-listed diagnoses by age, race-ethnicity, sex and region among privately insured enrollees, 2020.

Among commercial insurance enrollees, hospital discharge rates with celiac disease (all-listed diagnoses) were highest among persons 75 years and over and were twice as high among women compared with men (Table 212). Among persons with known race-ethnicity, rates were highest among Whites, followed by Blacks, then Hispanics, and lowest among Asians. Rates were highest in the Northeast, followed by the Midwest, then the West, and lowest in the South.

**Table 212:** Celiac Disease: Hospital discharges with first-listed and all-listed diagnoses by age, race-ethnicity, sex and region among privately insured enrollees, 2020.

| Demographic Characteristics |  | First-Listed Diagnosis |  | All-Listed Diagnoses |  |
| --- | --- | --- | --- | --- | --- |
|  |  | Number of events | Event rate per 100,000 | Number of events | Event rate per 100,000 |
| Age (years) | 0 to 11 | 2 | 0 | 12 | 1 |
|  | 12 to 24 | 3 | 0 | 157 | 11 |
|  | 25 to 44 | . | . | 302 | 11 |
|  | 45 to 54 | 2 | 0 | 139 | 10 |
|  | 55 to 64 | 4 | 0 | 276 | 17 |
|  | 65 to 74 | 7 | 0 | 459 | 16 |
|  | 75 plus | 3 | 0 | 769 | 30 |
| Race-ethnicity | White | 14 | 0 | 1,691 | 20 |
|  | Black | . | . | 122 | 10 |
|  | Hispanic | 2 | 0 | 113 | 8 |
|  | Asian | . | . | 29 | 5 |
|  | Unknown | 5 | 0 | 159 | 9 |
| Sex | Female | 11 | 0 | 1,480 | 21 |
|  | Male | 10 | 0 | 634 | 10 |
| Region | Northeast | 3 | 0 | 310 | 20 |
|  | Midwest | 7 | 0 | 609 | 18 |
|  | South | 6 | 0 | 674 | 12 |
|  | West | 5 | 0 | 521 | 16 |
| Total |  | 21 | 0 | 2,114 | 15 |

Source: De-identified Optum Clinformatic® Data Mart.
Enrollees with full enrollment in a commercial health plan or Medicare during the year.

Among Medicare beneficiaries, the claims-based prevalence of celiac disease (based on all-listed diagnoses) was 0.2% (Table 213). Prevalence was lowest in the oldest age group and higher among women and Whites. It was highest in the Northeast, and lowest in the South.

**Table 213:** Celiac Disease: Claims-based prevalence with first-listed and all-listed diagnoses by age, race, sex and region among fee-for-service, age-eligible Medicare beneficiaries, 2019.

| Demographic Characteristics |  | Number of enrollees | First-Listed Diagnosis |  | All-Listed Diagnoses |  |
| --- | --- | --- | --- | --- | --- | --- |
|  |  |  | Number of enrollees with disease | Percent of enrollees with disease | Number of enrollees with disease | Percent of enrollees with disease |
| Age (years) | 65 to 69 | 8,066,320 | 3,400 | 0.0% | 13,660 | 0.2% |
|  | 70 to 74 | 6,936,940 | 3,200 | 0.0% | 12,720 | 0.2% |
|  | 75 to 79 | 4,894,060 | 2,720 | 0.1% | 9,500 | 0.2% |
|  | 80 to 84 | 3,335,340 | 1,840 | 0.1% | 7,220 | 0.2% |
|  | 85+ | 3,578,120 | 940 | 0.0% | 4,880 | 0.1% |
| Race | White | 22,023,800 | 11,320 | 0.1% | 44,340 | 0.2% |
|  | Black | 1,861,660 | . | . | 680 | 0.0% |
|  | Other | 2,925,320 | . | . | 2,960 | 0.1% |
| Sex | Female | 14,975,560 | 8,560 | 0.1% | 32,980 | 0.2% |
|  | Male | 11,835,220 | 3,540 | 0.0% | 15,000 | 0.1% |
| Region | Northeast | 4,748,700 | 3,380 | 0.1% | 12,320 | 0.3% |
|  | Midwest | 6,069,800 | 2,540 | 0.0% | 11,100 | 0.2% |
|  | South | 10,595,900 | 4,100 | 0.0% | 15,760 | 0.1% |
|  | West | 5,396,380 | 2,080 | 0.0% | 8,800 | 0.2% |
| Total |  | 26,810,780 | 12,100 | 0.0% | 47,980 | 0.2% |

Source: Centers for Medicare & Medicaid Services, 5% Denominator and Claims Files.
Beneficiaries aged 65+ years with continuous and full Parts A and B enrollment and no Health Maintenance Organization enrollment during the year. Unweighted counts were multiplied by 20 to produce counts for this table.

Among Medicare beneficiaries, ambulatory care visit rates with celiac disease (all-listed diagnoses) increased with age until 85 years and were higher among women compared with men and almost five times higher among Whites compared with Blacks (Table 214). Rates were highest in the Northeast, followed by the Midwest, and lowest in the South and West.

**Table 214:**
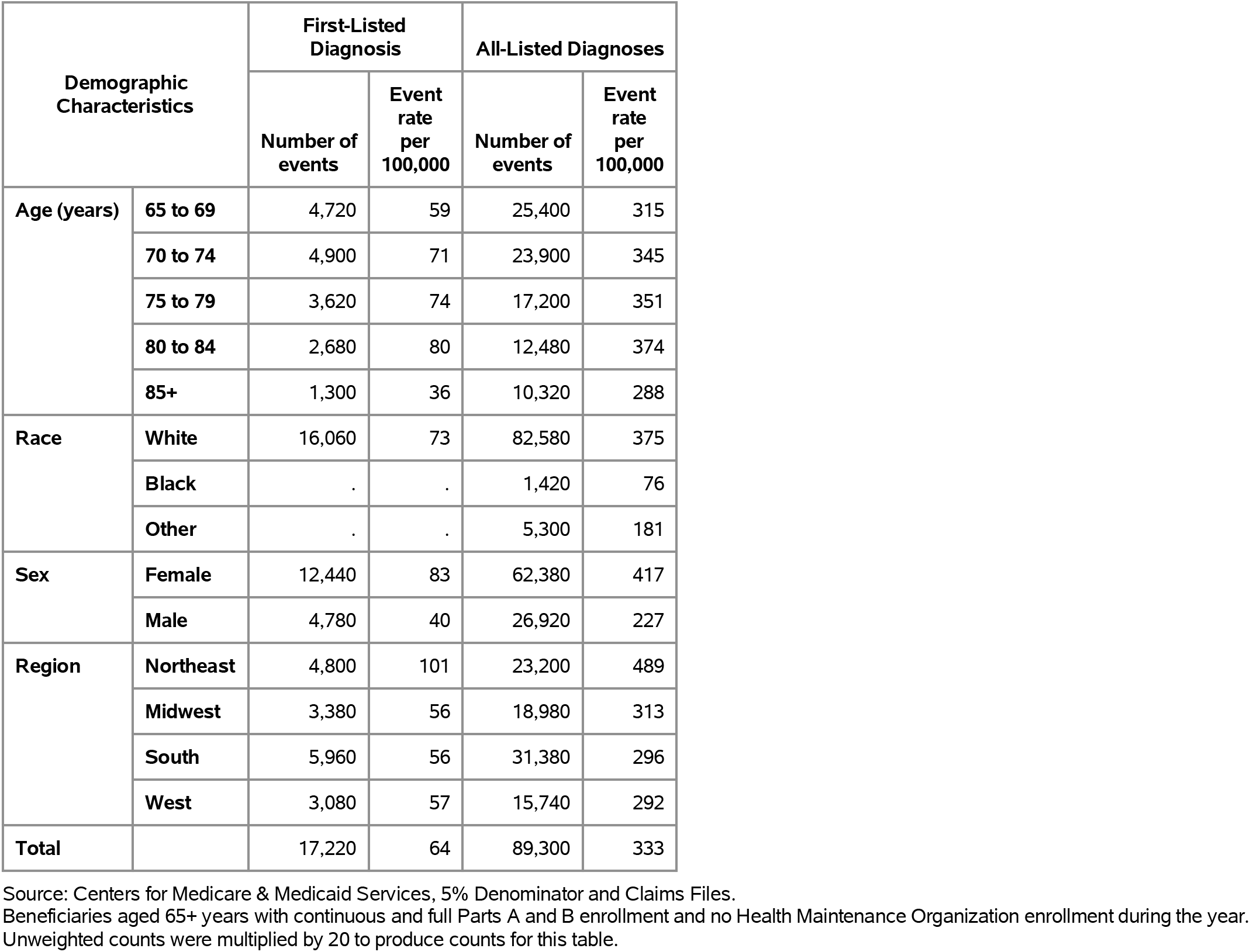
Celiac Disease: Ambulatory care visits with first-listed and all-listed diagnoses by age, race, sex and region among fee-for-service, age-eligible Medicare beneficiaries, 2019.

Among Medicare beneficiaries, emergency department visits with celiac disease were uncommon among Blacks (Table 215). Rates (all-listed diagnoses) increased with age until 85 years and were twice as high among women compared with men. Rates were higher in the Northeast compared with the Midwest, South, and West.

**Table 215:** Celiac Disease: Emergency department visits with first-listed and all-listed diagnoses by age, race, sex and region among fee-for-service, age-eligible Medicare beneficiaries, 2019.

| Demographic Characteristics |  | First-Listed Diagnosis |  | All-Listed Diagnoses |  |
| --- | --- | --- | --- | --- | --- |
|  |  | Number of events | Event rate per 100,000 | Number of events | Event rate per 100,000 |
| Age (years) | 65 to 69 | . | . | 2,580 | 32 |
|  | 70 to 74 | . | . | 3,140 | 45 |
|  | 75 to 79 | . | . | 3,300 | 67 |
|  | 80 to 84 | . | . | 2,780 | 83 |
|  | 85+ | . | . | 2,400 | 67 |
| Race | White | . | . | 13,260 | 60 |
|  | Black | . | . | . | . |
|  | Other | . | . | . | . |
| Sex | Female | . | . | 10,240 | 68 |
|  | Male | . | . | 3,960 | 33 |
| Region | Northeast | . | . | 3,540 | 75 |
|  | Midwest | . | . | 2,940 | 48 |
|  | South | . | . | 5,100 | 48 |
|  | West | . | . | 2,620 | 49 |
| Total |  | . | . | 14,200 | 53 |

Source: Centers for Medicare & Medicaid Services, 5% Denominator and Claims Files. Beneficiaries aged 65+ years with continuous and full Parts A and B enrollment and no Health Maintenance Organization enrollment during the year. Unweighted counts were multiplied by 20 to produce counts for this table.

Among Medicare beneficiaries, hospital discharges with celiac disease were uncommon among Blacks (Table 216). Rates (all-listed diagnoses) increased with age and were higher among women compared with men. Rates were higher in the Northeast compared with the South, Midwest, and West.

**Table 216:** Celiac Disease: Hospital discharges with first-listed and all-listed diagnoses by age, race, sex and region among fee-for-service, age-eligible Medicare beneficiaries, 2019.

| Demographic Characteristics |  | First-Listed Diagnosis |  | All-Listed Diagnoses |  |
| --- | --- | --- | --- | --- | --- |
|  |  | Number of events | Event rate per 100,000 | Number of events | Event rate per 100,000 |
| Age (years) | 65 to 69 | . | . | 2,160 | 27 |
|  | 70 to 74 | . | . | 2,560 | 37 |
|  | 75 to 79 | . | . | 2,560 | 52 |
|  | 80 to 84 | . | . | 1,940 | 58 |
|  | 85+ | . | . | 2,140 | 60 |
| Race | White | . | . | 10,580 | 48 |
|  | Black | . | . | . | . |
|  | Other | . | . | . | . |
| Sex | Female | . | . | 7,880 | 53 |
|  | Male | . | . | 3,480 | 29 |
| Region | Northeast | . | . | 2,960 | 62 |
|  | Midwest | . | . | 2,420 | 40 |
|  | South | . | . | 3,820 | 36 |
|  | West | . | . | 2,160 | 40 |
| Total |  | . | . | 11,360 | 42 |

Source: Centers for Medicare & Medicaid Services, 5% Denominator and Claims Files. Beneficiaries aged 65+ years with continuous and full Parts A and B enrollment and no Health Maintenance Organization enrollment during the year. Unweighted counts were multiplied by 20 to produce counts for this table.

Gastrointestinal infections (excluding C. difficile) contributed to 1.6 million ambulatory visits (2015) (Table 217). Ambulatory care visit rates (all-listed diagnoses) were highest among children and adolescents. Age-adjusted rates were higher among women compared with men, Blacks compared with Whites, and Hispanics compared with non-Hispanics.

**Table 217:** Gastrointestinal Infections (exc. C. difficile): Ambulatory care visits with first-listed and all-listed diagnoses by age, race, ethnicity, and sex in the United States, 2015.

| Demographic Characteristics |  | First-Listed Diagnosis |  | All-Listed Diagnoses |  |
| --- | --- | --- | --- | --- | --- |
|  |  | Number in thousands | Rate per 100,000 | Number in thousands | Rate per 100,000 |
| Age (years) | 0 to 11 | 339 | 696 | 468 | 962 |
|  | 12 to 24 | 323 | 576 | 377 | 671 |
|  | 25 to 44 | 270 | 320 | 318 | 377 |
|  | 45 to 54 | 84 | 196 | 123 | 286 |
|  | 55 to 64 | 119 | 291 | 167 | 410 |
|  | 65 to 74 | 37 | 135 | 108 | 393 |
|  | 75 plus | 77 | 382 | 81 | 400 |
| Race | White | 940 | 394 | 1,257 | 519 |
|  | Black | 286 | 627 | 354 | 786 |
|  | Other | 24 | 104 | 31 | 136 |
| Ethnicity | Hispanic | 217 | 394 | 316 | 551 |
|  | Not Hispanic | 1,033 | 422 | 1,327 | 528 |
| Sex | Female | 692 | 455 | 952 | 608 |
|  | Male | 557 | 368 | 690 | 454 |
| Total |  | 1,250 | 408 | 1,642 | 527 |

Source: National Ambulatory Medical Care Survey (3-year average, 2014-2016). Rates for age groups are unadjusted and all other rates are age-adjusted.

Gastrointestinal infections (excluding C. difficile) contributed to 698,000 emergency department visits in 2018 (Table 218). Emergency department visit rates (all-listed diagnoses) were highest among children and second highest among persons 75 years and over. Age-adjusted rates were higher among women compared with men.

**Table 218:** Gastrointestinal Infections (exc. C. difficile): Emergency department visits with first-listed and all-listed diagnoses by age and sex in the United States, 2018.

| Demographic Characteristics |  | First-Listed Diagnosis |  | All-Listed Diagnoses |  |
| --- | --- | --- | --- | --- | --- |
|  |  | Number in thousands | Rate per 100,000 | Number in thousands | Rate per 100,000 |
| Age (years) | 0 to 11 | 174 | 360 | 208 | 429 |
|  | 12 to 24 | 83 | 149 | 101 | 182 |
|  | 25 to 44 | 108 | 124 | 143 | 164 |
|  | 45 to 54 | 40 | 97 | 60 | 144 |
|  | 55 to 64 | 38 | 90 | 63 | 149 |
|  | 65 to 74 | 32 | 106 | 57 | 185 |
|  | 75 plus | 35 | 161 | 67 | 304 |
| Sex | Female | 287 | 179 | 394 | 239 |
|  | Male | 224 | 144 | 304 | 193 |
| Total |  | 510 | 162 | 698 | 217 |

Source: Healthcare Cost and Utilization Project Nationwide Emergency Department Sample. Rates for age groups are unadjusted and all other rates are age-adjusted.

Gastrointestinal infections (excluding C. difficile) contributed to 240,000 hospital discharges in 2018 (Table 219). Hospital discharge rates (all-listed diagnoses) were higher among children compared with adolescents and younger adults and then increased with age. Age-adjusted rates were higher among women compared with men, Blacks compared with Whites, and non-Hispanics compared with Hispanics.

**Table 219:** Gastrointestinal Infections (exc. C. difficile): Hospital discharges with first-listed and all-listed diagnoses by age, race, ethnicity, and sex in the United States, 2018.

| Demographic Characteristics |  | First-Listed Diagnosis |  | All-Listed Diagnoses |  |
| --- | --- | --- | --- | --- | --- |
|  |  | Number in thousands | Rate per 100,000 | Number in thousands | Rate per 100,000 |
| Age (years) | 0 to 11 | 16 | 32 | 31 | 64 |
|  | 12 to 24 | 7 | 12 | 14 | 25 |
|  | 25 to 44 | 16 | 18 | 36 | 42 |
|  | 45 to 54 | 13 | 30 | 28 | 68 |
|  | 55 to 64 | 16 | 39 | 39 | 92 |
|  | 65 to 74 | 17 | 55 | 40 | 133 |
|  | 75 plus | 21 | 97 | 51 | 231 |
| Race | White | 85 | 30 | 190 | 67 |
|  | Black | 14 | 32 | 35 | 78 |
|  | Other | 7 | 29 | 17 | 66 |
| Ethnicity | Hispanic | 16 | 30 | 34 | 65 |
|  | Not Hispanic | 91 | 31 | 208 | 69 |
| Sex | Female | 63 | 34 | 137 | 74 |
|  | Male | 43 | 26 | 102 | 61 |
| Total |  | 106 | 30 | 240 | 68 |

Source: Healthcare Cost and Utilization Project National Inpatient Sample. Rates for age groups are unadjusted and all other rates are age-adjusted.

Gastrointestinal infections (excluding C. difficile) contributed to 5,000 deaths in 2019 (Table 220). Mortality rates (underlying or other cause) were higher among children compared with adolescents and younger adults and then increased with age. Age-adjusted mortality rates were higher among women and non-Hispanics and did not differ by race.

**Table 220:** Gastrointestinal Infections (exc. C. difficile): Deaths with underlying or underlying/other cause and lifetime years of life lost by age, race, ethnicity, and sex in the United States, 2019.

| Demographic Characteristics |  | Underlying Cause |  |  | Underlying/Other Cause |  |  |
| --- | --- | --- | --- | --- | --- | --- | --- |
|  |  | Number of Deaths | Rate per 100,000 | Years of potential life lost in thousands | Number of Deaths | Rate per 100,000 | Years of potential life lost in thousands |
| Age (years) | 0 to 11 | 215 | 0.4 | 16.5 | 279 | 0.6 | 21.4 |
|  | 12 to 24 | 9 | 0.0 | 0.6 | 19 | 0.0 | 1.2 |
|  | 25 to 44 | 70 | 0.1 | 3.1 | 146 | 0.2 | 6.4 |
|  | 45 to 54 | 112 | 0.3 | 3.5 | 235 | 0.6 | 7.3 |
|  | 55 to 64 | 313 | 0.7 | 7.2 | 632 | 1.5 | 14.6 |
|  | 65 to 74 | 537 | 1.7 | 8.5 | 1,060 | 3.4 | 16.8 |
|  | 75 plus | 1,580 | 7.0 | 10.3 | 2,905 | 12.9 | 19.4 |
| Race | White | 2,398 | 0.7 | 36.9 | 4,519 | 1.3 | 66.9 |
|  | Black | 332 | 0.8 | 10.5 | 566 | 1.3 | 16.1 |
|  | Other | 106 | 0.4 | 2.2 | 191 | 0.8 | 4.0 |
| Ethnicity | Hispanic | 237 | 0.6 | 6.3 | 402 | 1.0 | 9.7 |
|  | Not Hispanic | 2,599 | 0.7 | 43.3 | 4,874 | 1.4 | 77.2 |
| Sex | Female | 1,701 | 0.8 | 27.5 | 3,103 | 1.4 | 48.1 |
|  | Male | 1,135 | 0.7 | 22.1 | 2,173 | 1.3 | 38.9 |
| Total |  | 2,836 | 0.7 | 49.6 | 5,276 | 1.3 | 87.0 |

Source: Vital Statistics of the United States.
Rates for age groups are unadjusted and all other rates are age-adjusted.
Years of potential life lost is based on U.S. life table 2006-2018 estimates of life expectancy at age of death.

Among privately insured enrollees, the claims-based prevalence of gastrointestinal infections (excluding C. difficile) (based on all-listed diagnoses) was 0.5% (Table 221). Prevalence was highest among children and higher among women. It was higher among Blacks and Hispanics compared with Whites and Asians. It was highest in the South, and lowest in the Midwest and West.

**Table 221:** Gastrointestinal Infections (exc. C. difficile): Claims-based prevalence with first-listed and all-listed diagnoses by age, race-ethnicity, sex and region among privately insured enrollees, 2020.

| Demographic Characteristics |  | Number of enrollees | First-Listed Diagnosis |  | All-Listed Diagnoses |  |
| --- | --- | --- | --- | --- | --- | --- |
|  |  |  | Number of enrollees with disease | Percent of enrollees with disease | Number of enrollees with disease | Percent of enrollees with disease |
| Age (years) | 0 to 11 | 1,101,978 | 6,863 | 0.6% | 9,069 | 0.8% |
|  | 12 to 24 | 1,451,892 | 4,595 | 0.3% | 6,335 | 0.4% |
|  | 25 to 44 | 2,656,672 | 7,506 | 0.3% | 11,473 | 0.4% |
|  | 45 to 54 | 1,452,320 | 3,713 | 0.3% | 6,703 | 0.5% |
|  | 55 to 64 | 1,640,216 | 4,038 | 0.2% | 7,891 | 0.5% |
|  | 65 to 74 | 2,840,615 | 6,251 | 0.2% | 13,250 | 0.5% |
|  | 75 plus | 2,576,241 | 5,661 | 0.2% | 12,428 | 0.5% |
| Race-ethnicity | White | 8,657,702 | 21,074 | 0.2% | 37,052 | 0.4% |
|  | Black | 1,242,477 | 3,801 | 0.3% | 7,231 | 0.6% |
|  | Hispanic | 1,493,457 | 5,039 | 0.3% | 9,396 | 0.6% |
|  | Asian | 607,284 | 1,370 | 0.2% | 2,421 | 0.4% |
|  | Unknown | 1,719,014 | 7,343 | 0.4% | 11,049 | 0.6% |
| Sex | Female | 7,199,363 | 21,975 | 0.3% | 38,875 | 0.5% |
|  | Male | 6,520,571 | 16,652 | 0.3% | 28,274 | 0.4% |
| Region | Northeast | 1,526,275 | 4,349 | 0.3% | 7,660 | 0.5% |
|  | Midwest | 3,322,846 | 8,389 | 0.3% | 13,719 | 0.4% |
|  | South | 5,619,435 | 19,018 | 0.3% | 33,725 | 0.6% |
|  | West | 3,251,378 | 6,871 | 0.2% | 12,045 | 0.4% |
| Total |  | 13,719,934 | 38,627 | 0.3% | 67,149 | 0.5% |

Source: De-identified Optum Clinformatic® Data Mart.
Enrollees with full enrollment in a commercial health plan or Medicare during the year.

Among commercial insurance enrollees, ambulatory care visit rates with gastrointestinal infections (excluding C. difficile) (all-listed diagnoses) were highest among children and were higher among women compared with men (Table 222). Among persons with known race-ethnicity, rates were highest among Hispanics, followed by Blacks, then Asians, and lowest among Whites. Rates were highest in the South, followed by the Northeast, and lowest in the Midwest and West.

**Table 222:** Gastrointestinal Infections (exc. C. difficile): Ambulatory care visits with first-listed and all-listed diagnoses by age, race-ethnicity, sex and region among privately insured enrollees, 2020.

| Demographic Characteristics |  | First-Listed Diagnosis |  | All-Listed Diagnoses |  |
| --- | --- | --- | --- | --- | --- |
|  |  | Number of events | Event rate per 100,000 | Number of events | Event rate per 100,000 |
| Age (years) | 0 to 11 | 7,126 | 647 | 9,592 | 870 |
|  | 12 to 24 | 4,791 | 330 | 6,796 | 468 |
|  | 25 to 44 | 8,008 | 301 | 12,919 | 486 |
|  | 45 to 54 | 3,852 | 265 | 7,267 | 500 |
|  | 55 to 64 | 4,017 | 245 | 8,065 | 492 |
|  | 65 to 74 | 6,298 | 222 | 13,223 | 465 |
|  | 75 plus | 5,716 | 222 | 11,860 | 460 |
| Race-ethnicity | White | 21,410 | 247 | 37,151 | 429 |
|  | Black | 3,880 | 312 | 7,281 | 586 |
|  | Hispanic | 5,365 | 359 | 10,559 | 707 |
|  | Asian | 1,466 | 241 | 2,765 | 455 |
|  | Unknown | 7,687 | 447 | 11,966 | 696 |
| Sex | Female | 22,588 | 314 | 40,674 | 565 |
|  | Male | 17,220 | 264 | 29,048 | 445 |
| Region | Northeast | 4,512 | 296 | 8,008 | 525 |
|  | Midwest | 8,538 | 257 | 13,571 | 408 |
|  | South | 19,559 | 348 | 34,967 | 622 |
|  | West | 7,199 | 221 | 13,176 | 405 |
| Total |  | 39,808 | 290 | 69,722 | 508 |

Source: De-identified Optum Clinformatic® Data Mart.
Enrollees with full enrollment in a commercial health plan or Medicare during the year.

Among commercial insurance enrollees, emergency department visit rates with gastrointestinal infections (excluding C. difficile) (all-listed diagnoses) increased with age and were higher among women compared with men (Table 223). Among persons with known race-ethnicity, rates were highest among Blacks, followed by Hispanics, then Whites, and lowest among Asians. Rates were highest in the South, followed by the Midwest, then the Northeast, and lowest in the West.

**Table 223:**
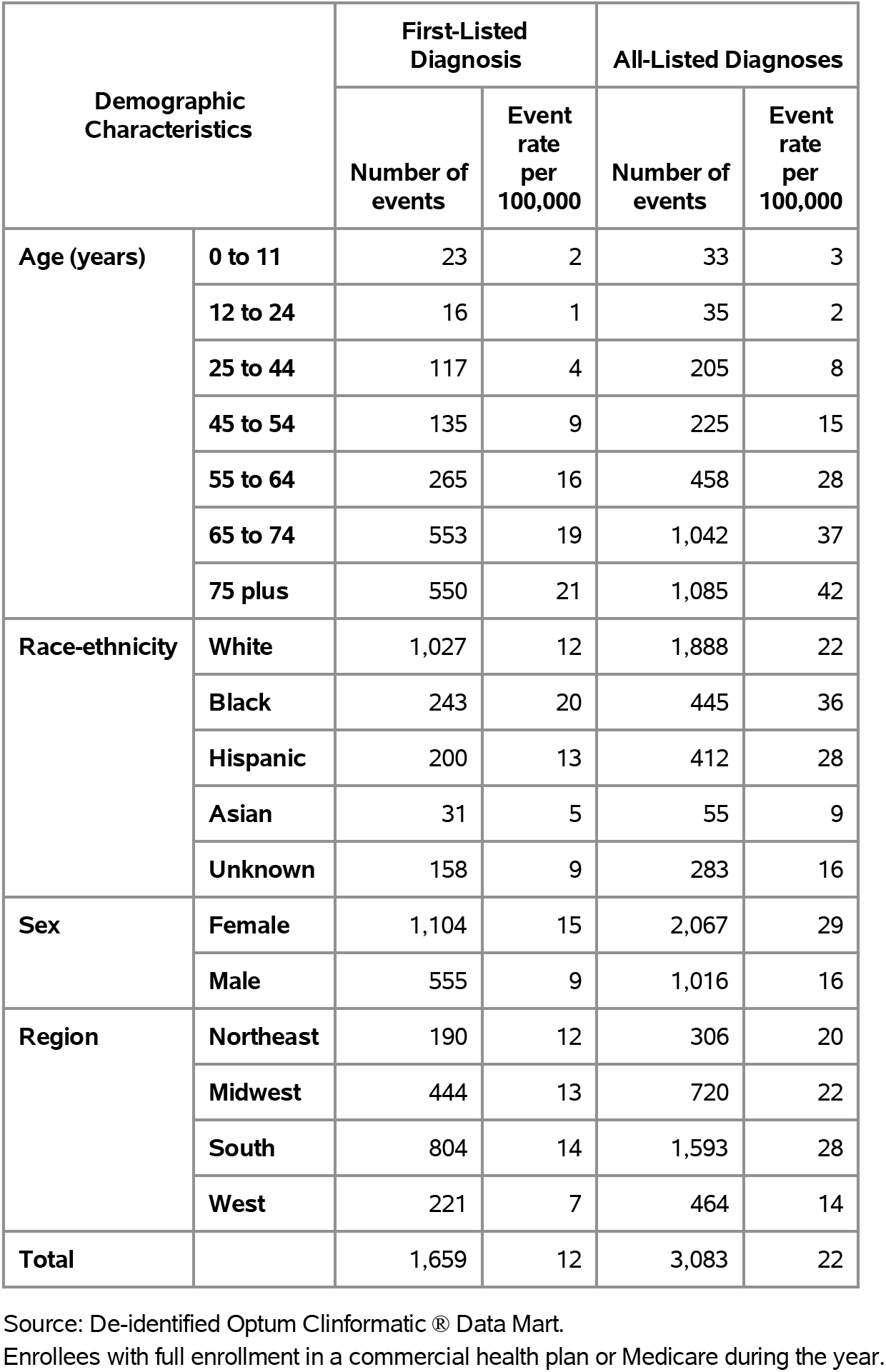
Gastrointestinal Infections (exc. C. difficile): Emergency department visits with first-listed and all-listed diagnoses by age, race-ethnicity, sex and region among privately insured enrollees, 2020.

Among commercial insurance enrollees, hospital discharge rates with gastrointestinal infections (excluding C. difficile) (all-listed diagnoses) were higher among children compared with adolescents and the youngest adults and then increased with age and were higher among women compared with men (Table 224). Among persons with known race-ethnicity, rates were highest among Blacks, followed by Whites and Hispanics, and lowest among Asians. Rates were highest in the Northeast, followed by the South, then the Midwest, and lowest in the West.

**Table 224:** Gastrointestinal Infections (exc. C. difficile): Hospital discharges with first-listed and all-listed diagnoses by age, race-ethnicity, sex and region among privately insured enrollees, 2020.

| Demographic Characteristics |  | First-Listed Diagnosis |  | All-Listed Diagnoses |  |
| --- | --- | --- | --- | --- | --- |
|  |  | Number of events | Event rate per 100,000 | Number of events | Event rate per 100,000 |
| Age (years) | 0 to 11 | 105 | 10 | 222 | 20 |
|  | 12 to 24 | 69 | 5 | 194 | 13 |
|  | 25 to 44 | 151 | 6 | 550 | 21 |
|  | 45 to 54 | 213 | 15 | 741 | 51 |
|  | 55 to 64 | 366 | 22 | 1,511 | 92 |
|  | 65 to 74 | 744 | 26 | 3,331 | 117 |
|  | 75 plus | 1,132 | 44 | 4,530 | 176 |
| Race-ethnicity | White | 1,839 | 21 | 7,104 | 82 |
|  | Black | 347 | 28 | 1,535 | 124 |
|  | Hispanic | 263 | 18 | 1,216 | 81 |
|  | Asian | 51 | 8 | 231 | 38 |
|  | Unknown | 280 | 16 | 993 | 58 |
| Sex | Female | 1,818 | 25 | 6,754 | 94 |
|  | Male | 962 | 15 | 4,325 | 66 |
| Region | Northeast | 366 | 24 | 1,571 | 103 |
|  | Midwest | 603 | 18 | 2,423 | 73 |
|  | South | 1,352 | 24 | 5,246 | 93 |
|  | West | 459 | 14 | 1,839 | 57 |
| Total |  | 2,780 | 20 | 11,079 | 81 |

Source: De-identified Optum Clinformatic® Data Mart.
Enrollees with full enrollment in a commercial health plan or Medicare during the year.

Among Medicare beneficiaries, the claims-based prevalence of gastrointestinal infections (excluding C. difficile) (based on all-listed diagnoses) was 0.8% (Table 225). Prevalence was highest among persons 75 years and over, higher among women, and did not differ by race. It was highest in the Northeast and South, and lowest in the Midwest.

**Table 225:** Gastrointestinal Infections (exc. C. difficile): Claims-based prevalence with first-listed and all-listed diagnoses by age, race, sex and region among fee-for-service, age-eligible Medicare beneficiaries, 2019.

| Demographic Characteristics |  | Number of enrollees | First-Listed Diagnosis |  | All-Listed Diagnoses |  |
| --- | --- | --- | --- | --- | --- | --- |
|  |  |  | Number of enrollees with disease | Percent of enrollees with disease | Number of enrollees with disease | Percent of enrollees with disease |
| Age (years) | 65 to 69 | 8,066,320 | 30,860 | 0.4% | 56,600 | 0.7% |
|  | 70 to 74 | 6,936,940 | 30,000 | 0.4% | 54,300 | 0.8% |
|  | 75 to 79 | 4,894,060 | 22,540 | 0.5% | 41,700 | 0.9% |
|  | 80 to 84 | 3,335,340 | 17,660 | 0.5% | 30,960 | 0.9% |
|  | 85+ | 3,578,120 | 18,360 | 0.5% | 33,000 | 0.9% |
| Race | White | 22,023,800 | 96,880 | 0.4% | 174,260 | 0.8% |
|  | Black | 1,861,660 | 7,740 | 0.4% | 14,300 | 0.8% |
|  | Other | 2,925,320 | 14,800 | 0.5% | 28,000 | 1.0% |
| Sex | Female | 14,975,560 | 77,640 | 0.5% | 139,240 | 0.9% |
|  | Male | 11,835,220 | 41,780 | 0.4% | 77,320 | 0.7% |
| Region | Northeast | 4,748,700 | 22,820 | 0.5% | 41,000 | 0.9% |
|  | Midwest | 6,069,800 | 24,620 | 0.4% | 43,700 | 0.7% |
|  | South | 10,595,900 | 50,380 | 0.5% | 90,780 | 0.9% |
|  | West | 5,396,380 | 21,600 | 0.4% | 41,080 | 0.8% |
| Total |  | 26,810,780 | 119,420 | 0.4% | 216,560 | 0.8% |

Source: Centers for Medicare & Medicaid Services, 5% Denominator and Claims Files. Beneficiaries aged 65+ years with continuous and full Parts A and B enrollment and no Health Maintenance Organization enrollment during the year. Unweighted counts were multiplied by 20 to produce counts for this table.

Among Medicare beneficiaries, ambulatory care visit rates with gastrointestinal infections (excluding C. difficile) (all-listed diagnoses) increased with age and were higher among women compared with men and Whites compared with Blacks (Table 226). Rates were highest in the Northeast and South, followed by the West, and lowest in the Midwest.

**Table 226:** Gastrointestinal Infections (exc. C. difficile): Ambulatory care visits with first-listed and all-listed diagnoses by age, race, sex and region among fee-for-service, age-eligible Medicare beneficiaries, 2019.

| Demographic Characteristics |  | First-Listed Diagnosis |  | All-Listed Diagnoses |  |
| --- | --- | --- | --- | --- | --- |
|  |  | Number of events | Event rate per 100,000 | Number of events | Event rate per 100,000 |
| Age (years) | 65 to 69 | 24,660 | 306 | 52,760 | 654 |
|  | 70 to 74 | 23,740 | 342 | 49,320 | 711 |
|  | 75 to 79 | 18,760 | 383 | 38,600 | 789 |
|  | 80 to 84 | 13,260 | 398 | 26,480 | 794 |
|  | 85+ | 14,620 | 409 | 28,760 | 804 |
| Race | White | 76,520 | 347 | 154,820 | 703 |
|  | Black | 5,620 | 302 | 12,840 | 690 |
|  | Other | 12,900 | 441 | 28,260 | 966 |
| Sex | Female | 61,700 | 412 | 126,620 | 846 |
|  | Male | 33,340 | 282 | 69,300 | 586 |
| Region | Northeast | 18,980 | 400 | 37,820 | 796 |
|  | Midwest | 17,460 | 288 | 34,960 | 576 |
|  | South | 40,720 | 384 | 84,140 | 794 |
|  | West | 17,880 | 331 | 39,000 | 723 |
| Total |  | 95,040 | 354 | 195,920 | 731 |

Source: Centers for Medicare & Medicaid Services, 5% Denominator and Claims Files. Beneficiaries aged 65+ years with continuous and full Parts A and B enrollment and no Health Maintenance Organization enrollment during the year. Unweighted counts were multiplied by 20 to produce counts for this table.

Among Medicare beneficiaries, emergency department visit rates with gastrointestinal infections (excluding C. difficile) (all-listed diagnoses) increased with age and were higher among women compared with men and Blacks compared with Whites (Table 227). Rates were highest in the Northeast, followed by the Midwest, then the South, and lowest in the West.

**Table 227:**
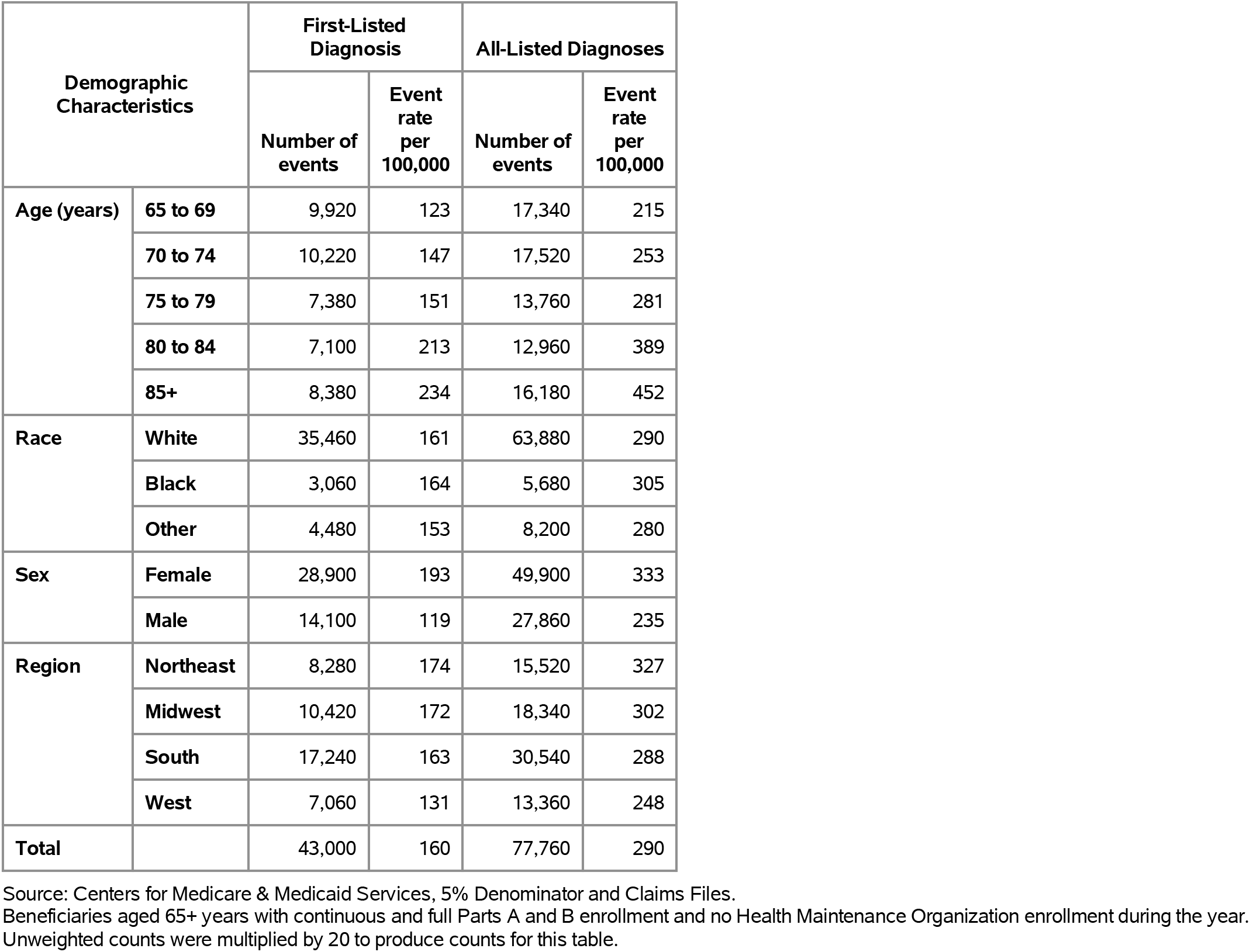
Gastrointestinal Infections (exc. C. difficile): Emergency department visits with first-listed and all-listed diagnoses by age, race, sex and region among fee-for-service, age-eligible Medicare beneficiaries, 2019.

| Demographic Characteristics |  | First-Listed Diagnosis |  | All-Listed Diagnoses |  |
| --- | --- | --- | --- | --- | --- |
|  |  | Number of events | Event rate per 100,000 | Number of events | Event rate per 100,000 |
| Age (years) | 65 to 69 | 9,920 | 123 | 17,340 | 215 |
|  | 70 to 74 | 10,220 | 147 | 17,520 | 253 |
|  | 75 to 79 | 7,380 | 151 | 13,760 | 281 |
|  | 80 to 84 | 7,100 | 213 | 12,960 | 389 |
|  | 85+ | 8,380 | 234 | 16,180 | 452 |
| Race | White | 35,460 | 161 | 63,880 | 290 |
|  | Black | 3,060 | 164 | 5,680 | 305 |
|  | Other | 4,480 | 153 | 8,200 | 280 |
| Sex | Female | 28,900 | 193 | 49,900 | 333 |
|  | Male | 14,100 | 119 | 27,860 | 235 |
| Region | Northeast | 8,280 | 174 | 15,520 | 327 |
|  | Midwest | 10,420 | 172 | 18,340 | 302 |
|  | South | 17,240 | 163 | 30,540 | 288 |
|  | West | 7,060 | 131 | 13,360 | 248 |
| Total |  | 43,000 | 160 | 77,760 | 290 |

Source: Centers for Medicare & Medicaid Services, 5% Denominator and Claims Files. Beneficiaries aged 65+ years with continuous and full Parts A and B enrollment and no Health Maintenance Organization enrollment during the year. Unweighted counts were multiplied by 20 to produce counts for this table.

Among Medicare beneficiaries, hospital discharge rates with gastrointestinal infections (excluding C. difficile) (all-listed diagnoses) increased with age and were higher among women compared with men but differed little by race (Table 228). Rates were highest in the Northeast, followed by the Midwest, then the South, and lowest in the West.

**Table 228:** Gastrointestinal Infections (exc. C. difficile): Hospital discharges with first-listed and all-listed diagnoses by age, race, sex and region among fee-for-service, age-eligible Medicare beneficiaries, 2019.

| Demographic Characteristics |  | First-Listed Diagnosis |  | All-Listed Diagnoses |  |
| --- | --- | --- | --- | --- | --- |
|  |  | Number of events | Event rate per 100,000 | Number of events | Event rate per 100,000 |
| Age (years) | 65 to 69 | 4,660 | 58 | 11,600 | 144 |
|  | 70 to 74 | 5,380 | 78 | 12,060 | 174 |
|  | 75 to 79 | 3,920 | 80 | 10,040 | 205 |
|  | 80 to 84 | 4,080 | 122 | 9,520 | 285 |
|  | 85+ | 4,880 | 136 | 12,040 | 336 |
| Race | White | 18,920 | 86 | 45,520 | 207 |
|  | Black | 1,680 | 90 | 3,940 | 212 |
|  | Other | 2,320 | 79 | 5,800 | 198 |
| Sex | Female | 15,200 | 101 | 34,760 | 232 |
|  | Male | 7,720 | 65 | 20,500 | 173 |
| Region | Northeast | 5,020 | 106 | 12,000 | 253 |
|  | Midwest | 5,500 | 91 | 13,160 | 217 |
|  | South | 8,720 | 82 | 20,580 | 194 |
|  | West | 3,680 | 68 | 9,520 | 176 |
| Total |  | 22,920 | 85 | 55,260 | 206 |

Source: Centers for Medicare & Medicaid Services, 5% Denominator and Claims Files. Beneficiaries aged 65+ years with continuous and full Parts A and B enrollment and no Health Maintenance Organization enrollment during the year. Unweighted counts were multiplied by 20 to produce counts for this table.

Clostridium difficile contributed to 249,000 ambulatory visits (2015) (Table 229). Ambulatory care visit rates (all-listed diagnoses) were higher among children compared with adolescents and younger adults and then increased with age. Age-adjusted rates were higher among women compared with men, Whites compared with Blacks, and non-Hispanics compared with Hispanics.

**Table 229:**
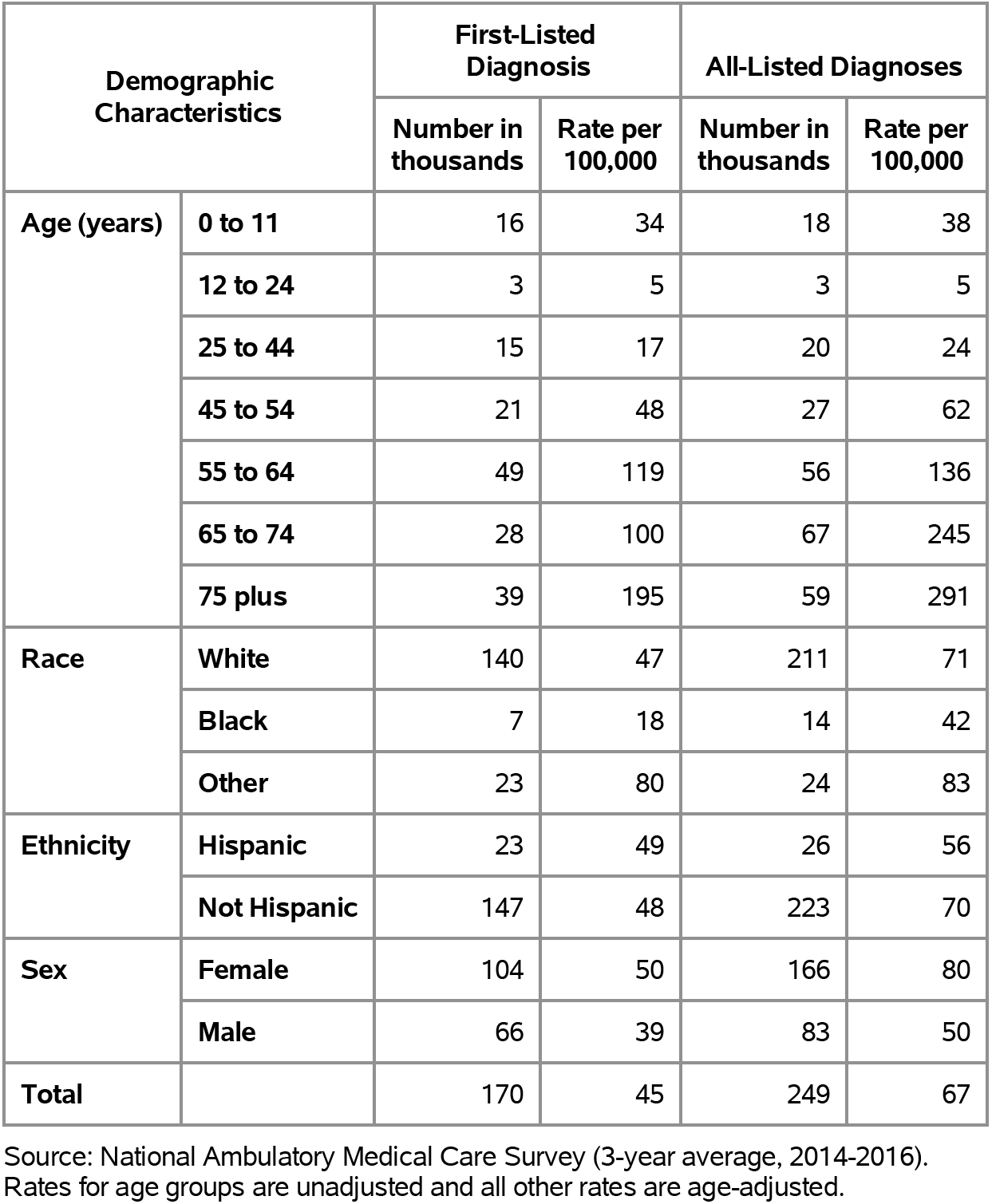
Clostridium difficile: Ambulatory care visits with first-listed and all-listed diagnoses by age, race, ethnicity, and sex in the United States, 2015.

| Demographic Characteristics |  | First-Listed Diagnosis |  | All-Listed Diagnoses |  |
| --- | --- | --- | --- | --- | --- |
|  |  | Number in thousands | Rate per 100,000 | Number in thousands | Rate per 100,000 |
| Age (years) | 0 to 11 | 16 | 34 | 18 | 38 |
|  | 12 to 24 | 3 | 5 | 3 | 5 |
|  | 25 to 44 | 15 | 17 | 20 | 24 |
|  | 45 to 54 | 21 | 48 | 27 | 62 |
|  | 55 to 64 | 49 | 119 | 56 | 136 |
|  | 65 to 74 | 28 | 100 | 67 | 245 |
|  | 75 plus | 39 | 195 | 59 | 291 |
| Race | White | 140 | 47 | 211 | 71 |
|  | Black | 7 | 18 | 14 | 42 |
|  | Other | 23 | 80 | 24 | 83 |
| Ethnicity | Hispanic | 23 | 49 | 26 | 56 |
|  | Not Hispanic | 147 | 48 | 223 | 70 |
| Sex | Female | 104 | 50 | 166 | 80 |
|  | Male | 66 | 39 | 83 | 50 |
| Total |  | 170 | 45 | 249 | 67 |

Source: National Ambulatory Medical Care Survey (3-year average, 2014-2016). Rates for age groups are unadjusted and all other rates are age-adjusted.

Clostridium difficile contributed to 293,000 emergency department visits in 2018 (Table 230). Emergency department visit rates (all-listed diagnoses) increased with age. Age-adjusted rates were higher among women compared with men.

**Table 230:** Clostridium difficile: Emergency department visits with first-listed and all-listed diagnoses by age and sex in the United States, 2018.

| Demographic Characteristics |  | First-Listed Diagnosis |  | All-Listed Diagnoses |  |
| --- | --- | --- | --- | --- | --- |
|  |  | Number in thousands | Rate per 100,000 | Number in thousands | Rate per 100,000 |
| Age (years) | 0 to 11 | 3 | 6 | 5 | 11 |
|  | 12 to 24 | 4 | 8 | 9 | 16 |
|  | 25 to 44 | 14 | 16 | 32 | 37 |
|  | 45 to 54 | 12 | 29 | 31 | 75 |
|  | 55 to 64 | 18 | 43 | 52 | 123 |
|  | 65 to 74 | 21 | 68 | 63 | 208 |
|  | 75 plus | 36 | 162 | 100 | 454 |
| Sex | Female | 70 | 35 | 173 | 85 |
|  | Male | 39 | 23 | 120 | 69 |
| Total |  | 108 | 29 | 293 | 78 |

Source: Healthcare Cost and Utilization Project Nationwide Emergency Department Sample. Rates for age groups are unadjusted and all other rates are age-adjusted.

Clostridium difficile contributed to 307,000 hospital discharges in 2018 (Table 231). Hospital discharge rates (all-listed diagnoses) increased with age. Age-adjusted rates were higher among women compared with men, Blacks compared with Whites, and non-Hispanics compared with Hispanics.

**Table 231:** Clostridium difficile: Hospital discharges with first-listed and all-listed diagnoses by age, race, ethnicity, and sex in the United States, 2018.

| Demographic Characteristics |  | First-Listed Diagnosis |  | All-Listed Diagnoses |  |
| --- | --- | --- | --- | --- | --- |
|  |  | Number in thousands | Rate per 100,000 | Number in thousands | Rate per 100,000 |
| Age (years) | 0 to 11 | 2 | 4 | 5 | 10 |
|  | 12 to 24 | 3 | 5 | 8 | 15 |
|  | 25 to 44 | 9 | 10 | 29 | 33 |
|  | 45 to 54 | 9 | 21 | 30 | 73 |
|  | 55 to 64 | 15 | 35 | 57 | 135 |
|  | 65 to 74 | 19 | 63 | 71 | 233 |
|  | 75 plus | 33 | 149 | 106 | 485 |
| Race | White | 77 | 25 | 254 | 81 |
|  | Black | 10 | 23 | 40 | 93 |
|  | Other | 3 | 15 | 15 | 63 |
| Ethnicity | Hispanic | 7 | 18 | 27 | 63 |
|  | Not Hispanic | 82 | 25 | 282 | 84 |
| Sex | Female | 57 | 28 | 175 | 85 |
|  | Male | 32 | 19 | 132 | 75 |
| Total |  | 89 | 23 | 307 | 80 |

Source: Healthcare Cost and Utilization Project National Inpatient Sample. Rates for age groups are unadjusted and all other rates are age-adjusted.

Clostridium difficile contributed to 8,000 deaths in 2019 (Table 232). Mortality was uncommon among the youngest age groups after which rates (underlying or other cause) increased with age. Age-adjusted mortality rates were higher among men, Whites, and non-Hispanics.

**Table 232:**
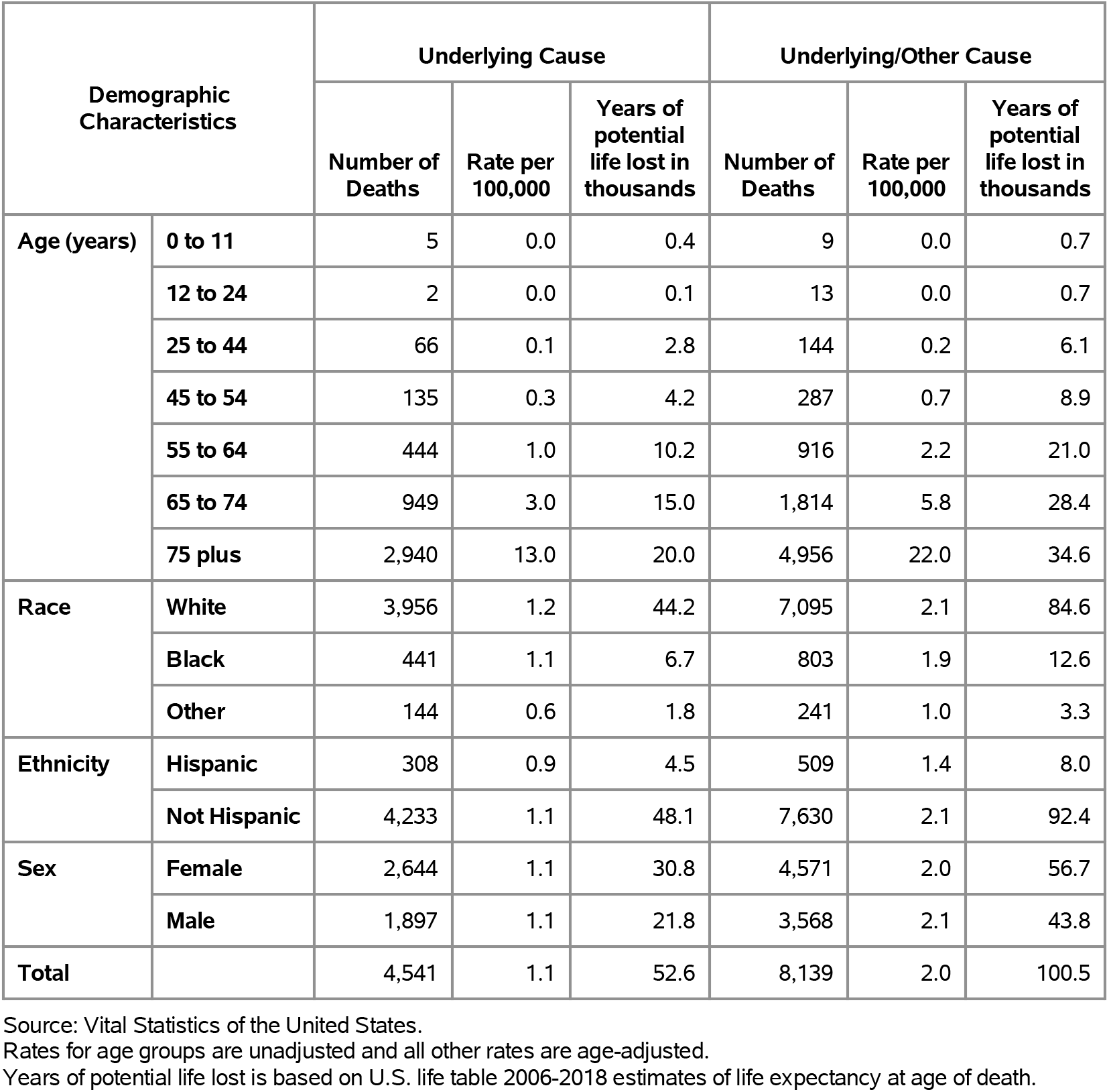
Clostridium difficile: Deaths with underlying or underlying/other cause and lifetime years of life lost by age, race, ethnicity, and sex in the United States, 2019.

Among privately insured enrollees, the claims-based prevalence of Clostridium difficile (based on all-listed diagnoses) was 0.1% (Table 233). C. difficile was uncommon among children and adolescents and the youngest adults and then prevalence increased with age. Prevalence was higher among women and higher among Whites and Blacks compared with Hispanics and Asians. It differed little by region.

**Table 233:**
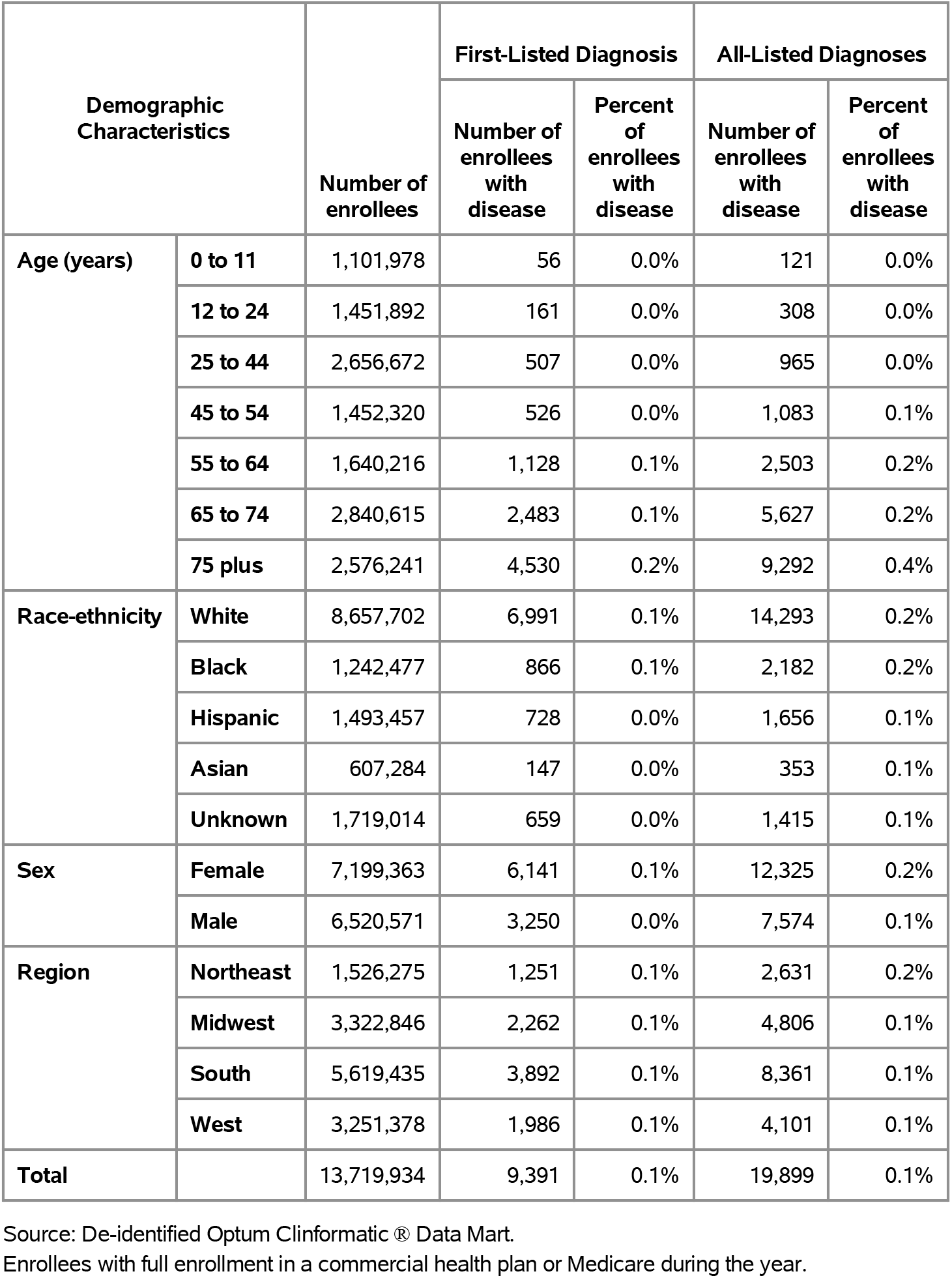
Clostridium difficile: Claims-based prevalence with first-listed and all-listed diagnoses by age, race-ethnicity, sex and region among privately insured enrollees, 2020.

Among commercial insurance enrollees, ambulatory care visit rates with Clostridium difficile (all-listed diagnoses) increased with age and were higher among women compared with men (Table 234). Among persons with known race-ethnicity, rates were highest among Whites, followed by Blacks, then Hispanics, and lowest among Asians. Rates were highest in the Northeast, followed by the Midwest and West, and lowest in the South.

**Table 234:** Clostridium difficile: Ambulatory care visits with first-listed and all-listed diagnoses by age, race-ethnicity, sex and region among privately insured enrollees, 2020.

| Demographic Characteristics |  | First-Listed Diagnosis |  | All-Listed Diagnoses |  |
| --- | --- | --- | --- | --- | --- |
|  |  | Number of events | Event rate per 100,000 | Number of events | Event rate per 100,000 |
| Age (years) | 0 to 11 | 63 | 6 | 147 | 13 |
|  | 12 to 24 | 222 | 15 | 454 | 31 |
|  | 25 to 44 | 744 | 28 | 1,533 | 58 |
|  | 45 to 54 | 765 | 53 | 1,670 | 115 |
|  | 55 to 64 | 1,771 | 108 | 4,123 | 251 |
|  | 65 to 74 | 4,218 | 148 | 10,143 | 357 |
|  | 75 plus | 9,474 | 368 | 21,614 | 839 |
| Race-ethnicity | White | 13,316 | 154 | 29,790 | 344 |
|  | Black | 1,376 | 111 | 3,631 | 292 |
|  | Hispanic | 1,206 | 81 | 3,107 | 208 |
|  | Asian | 221 | 36 | 573 | 94 |
|  | Unknown | 1,138 | 66 | 2,583 | 150 |
| Sex | Female | 11,239 | 156 | 25,525 | 355 |
|  | Male | 6,018 | 92 | 14,159 | 217 |
| Region | Northeast | 2,588 | 170 | 5,918 | 388 |
|  | Midwest | 3,967 | 119 | 9,593 | 289 |
|  | South | 6,684 | 119 | 14,887 | 265 |
|  | West | 4,018 | 124 | 9,286 | 286 |
| Total |  | 17,257 | 126 | 39,684 | 289 |

Source: De-identified Optum Clinformatic® Data Mart.
Enrollees with full enrollment in a commercial health plan or Medicare during the year.

Among commercial insurance enrollees, emergency department visit rates with Clostridium difficile (all-listed diagnoses) increased with age and were higher among women compared with men (Table 235). Among persons with known race-ethnicity, rates were highest among Whites, followed by Blacks, then Hispanics, and lowest among Asians. Rates were higher in the Northeast and Midwest compared with the South and West.

**Table 235:**
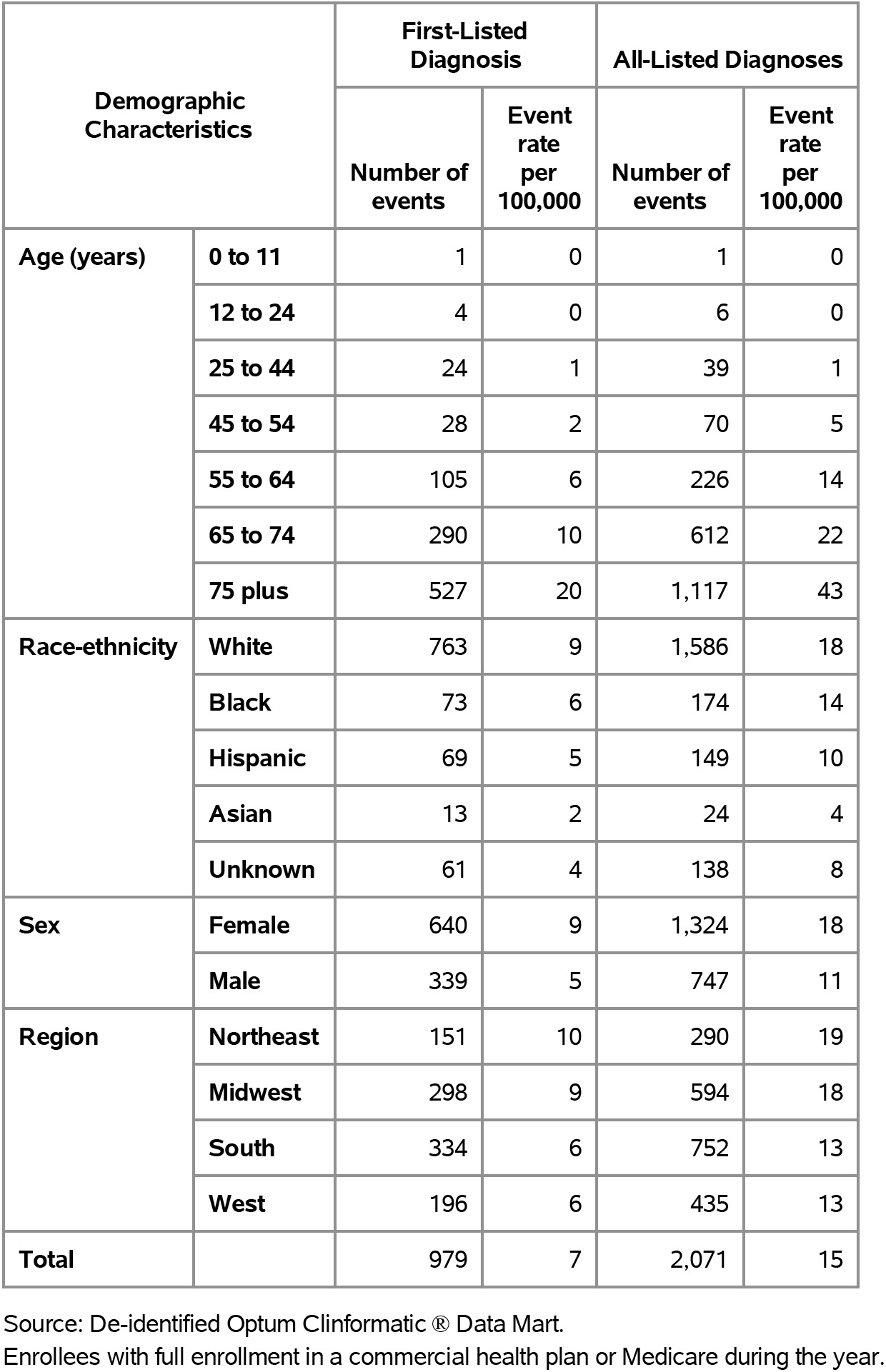
Clostridium difficile: Emergency department visits with first-listed and all-listed diagnoses by age, race-ethnicity, sex and region among privately insured enrollees, 2020.

| Demographic Characteristics |  | First-Listed Diagnosis |  | All-Listed Diagnoses |  |
| --- | --- | --- | --- | --- | --- |
|  |  | Number of events | Event rate per 100,000 | Number of events | Event rate per 100,000 |
| Age (years) | 0 to 11 | 1 | 0 | 1 | 0 |
|  | 12 to 24 | 4 | 0 | 6 | 0 |
|  | 25 to 44 | 24 | 1 | 39 | 1 |
|  | 45 to 54 | 28 | 2 | 70 | 5 |
|  | 55 to 64 | 105 | 6 | 226 | 14 |
|  | 65 to 74 | 290 | 10 | 612 | 22 |
|  | 75 plus | 527 | 20 | 1,117 | 43 |
| Race-ethnicity | White | 763 | 9 | 1,586 | 18 |
|  | Black | 73 | 6 | 174 | 14 |
|  | Hispanic | 69 | 5 | 149 | 10 |
|  | Asian | 13 | 2 | 24 | 4 |
|  | Unknown | 61 | 4 | 138 | 8 |
| Sex | Female | 640 | 9 | 1,324 | 18 |
|  | Male | 339 | 5 | 747 | 11 |
| Region | Northeast | 151 | 10 | 290 | 19 |
|  | Midwest | 298 | 9 | 594 | 18 |
|  | South | 334 | 6 | 752 | 13 |
|  | West | 196 | 6 | 435 | 13 |
| Total |  | 979 | 7 | 2,071 | 15 |

Source: De-identified Optum Clinformatics® Data Mart.
Enrollees with full enrollment in a commercial health plan or Medicare during the year.

Among commercial insurance enrollees, hospital discharge rates with Clostridium difficile (all-listed diagnoses) increased with age and were higher among women compared with men (Table 236). Among persons with known race-ethnicity, rates were highest among Blacks, followed by Whites, then Hispanics, and lowest among Asians. Rates were highest in the Northeast, followed by the South, then the Midwest, and lowest in the West.

**Table 236:** Clostridium difficile: Hospital discharges with first-listed and all-listed diagnoses by age, race-ethnicity, sex and region among privately insured enrollees, 2020.

| Demographic Characteristics |  | First-Listed Diagnosis |  | All-Listed Diagnoses |  |
| --- | --- | --- | --- | --- | --- |
|  |  | Number of events | Event rate per 100,000 | Number of events | Event rate per 100,000 |
| Age (years) | 0 to 11 | 14 | 1 | 65 | 6 |
|  | 12 to 24 | 33 | 2 | 133 | 9 |
|  | 25 to 44 | 90 | 3 | 393 | 15 |
|  | 45 to 54 | 156 | 11 | 662 | 46 |
|  | 55 to 64 | 371 | 23 | 1,904 | 116 |
|  | 65 to 74 | 841 | 30 | 4,305 | 152 |
|  | 75 plus | 1,595 | 62 | 6,788 | 263 |
| Race-ethnicity | White | 2,240 | 26 | 9,845 | 114 |
|  | Black | 336 | 27 | 1,869 | 150 |
|  | Hispanic | 269 | 18 | 1,297 | 87 |
|  | Asian | 37 | 6 | 227 | 37 |
|  | Unknown | 218 | 13 | 1,012 | 59 |
| Sex | Female | 2,053 | 29 | 8,320 | 116 |
|  | Male | 1,047 | 16 | 5,930 | 91 |
| Region | Northeast | 403 | 26 | 1,895 | 124 |
|  | Midwest | 683 | 21 | 3,449 | 104 |
|  | South | 1,446 | 26 | 6,209 | 110 |
|  | West | 568 | 17 | 2,697 | 83 |
| Total |  | 3,100 | 23 | 14,250 | 104 |

Source: De-identified Optum Clinformatic® Data Mart.
Enrollees with full enrollment in a commercial health plan or Medicare during the year.

Among Medicare beneficiaries, the claims-based prevalence of Clostridium difficile (based on all-listed diagnoses) was 0.5% (Table 237). Prevalence increased with age, was higher among women, and did not differ by race. It differed little by region.

**Table 237:** Clostridium difficile: Claims-based prevalence with first-listed and all-listed diagnoses by age, race, sex and region among fee-for-service, age-eligible Medicare beneficiaries, 2019.

| Demographic Characteristics |  | Number of enrollees | First-Listed Diagnosis |  | All-Listed Diagnoses |  |
| --- | --- | --- | --- | --- | --- | --- |
|  |  |  | Number of enrollees with disease | Percent of enrollees with disease | Number of enrollees with disease | Percent of enrollees with disease |
| Age (years) | 65 to 69 | 8,066,320 | 10,820 | 0.1% | 23,040 | 0.3% |
|  | 70 to 74 | 6,936,940 | 13,600 | 0.2% | 26,500 | 0.4% |
|  | 75 to 79 | 4,894,060 | 14,440 | 0.3% | 25,760 | 0.5% |
|  | 80 to 84 | 3,335,340 | 10,100 | 0.3% | 20,060 | 0.6% |
|  | 85+ | 3,578,120 | 16,520 | 0.5% | 29,880 | 0.8% |
| Race | White | 22,023,800 | 57,120 | 0.3% | 106,640 | 0.5% |
|  | Black | 1,861,660 | 4,020 | 0.2% | 8,940 | 0.5% |
|  | Other | 2,925,320 | 4,340 | 0.1% | 9,660 | 0.3% |
| Sex | Female | 14,975,560 | 42,500 | 0.3% | 78,120 | 0.5% |
|  | Male | 11,835,220 | 22,980 | 0.2% | 47,120 | 0.4% |
| Region | Northeast | 4,748,700 | 12,040 | 0.3% | 23,480 | 0.5% |
|  | Midwest | 6,069,800 | 16,480 | 0.3% | 30,080 | 0.5% |
|  | South | 10,595,900 | 26,420 | 0.2% | 50,320 | 0.5% |
|  | West | 5,396,380 | 10,540 | 0.2% | 21,360 | 0.4% |
| Total |  | 26,810,780 | 65,480 | 0.2% | 125,240 | 0.5% |

Source: Centers for Medicare & Medicaid Services, 5% Denominator and Claims Files. Beneficiaries aged 65+ years with continuous and full Parts A and B enrollment and no Health Maintenance Organization enrollment during the year. Unweighted counts were multiplied by 20 to produce counts for this table.

Among Medicare beneficiaries, ambulatory care visit rates with Clostridium difficile (all-listed diagnoses) increased with age and were higher among women compared with men and Whites compared with Blacks (Table 238). Rates were highest in the Northeast, followed by the Midwest, then the South, and lowest in the West.

**Table 238:** Clostridium difficile: Ambulatory care visits with first-listed and all-listed diagnoses by age, race, sex and region among fee-for-service, age-eligible Medicare beneficiaries, 2019.

| Demographic Characteristics |  | First-Listed Diagnosis |  | All-Listed Diagnoses |  |
| --- | --- | --- | --- | --- | --- |
|  |  | Number of events | Event rate per 100,000 | Number of events | Event rate per 100,000 |
| Age (years) | 65 to 69 | 16,700 | 207 | 41,420 | 513 |
|  | 70 to 74 | 19,480 | 281 | 49,680 | 716 |
|  | 75 to 79 | 23,980 | 490 | 52,540 | 1,074 |
|  | 80 to 84 | 18,820 | 564 | 42,920 | 1,287 |
|  | 85+ | 31,020 | 867 | 70,900 | 1,981 |
| Race | White | 96,780 | 439 | 225,020 | 1,022 |
|  | Black | 6,340 | 341 | 15,880 | 853 |
|  | Other | 6,880 | 235 | 16,560 | 566 |
| Sex | Female | 74,480 | 497 | 169,100 | 1,129 |
|  | Male | 35,520 | 300 | 88,360 | 747 |
| Region | Northeast | 22,440 | 473 | 55,240 | 1,163 |
|  | Midwest | 25,820 | 425 | 59,400 | 979 |
|  | South | 43,440 | 410 | 98,420 | 929 |
|  | West | 18,300 | 339 | 44,400 | 823 |
| Total |  | 110,000 | 410 | 257,460 | 960 |

Source: Centers for Medicare & Medicaid Services, 5% Denominator and Claims Files. Beneficiaries aged 65+ years with continuous and full Parts A and B enrollment and no Health Maintenance Organization enrollment during the year. Unweighted counts were multiplied by 20 to produce counts for this table.

Among Medicare beneficiaries, emergency department visit rates with Clostridium difficile (all-listed diagnoses) increased with age and were higher among women compared with men and Blacks compared with Whites (Table 239). Rates were highest in the Northeast and Midwest, followed by the South, and lowest in the West.

**Table 239:** Clostridium difficile: Emergency department visits with first-listed and all-listed diagnoses by age, race, sex and region among fee-for-service, age-eligible Medicare beneficiaries, 2019.

| Demographic Characteristics |  | First-Listed Diagnosis |  | All-Listed Diagnoses |  |
| --- | --- | --- | --- | --- | --- |
|  |  | Number of events | Event rate per 100,000 | Number of events | Event rate per 100,000 |
| Age (years) | 65 to 69 | 5,200 | 64 | 15,920 | 197 |
|  | 70 to 74 | 6,680 | 96 | 18,140 | 261 |
|  | 75 to 79 | 7,000 | 143 | 18,040 | 369 |
|  | 80 to 84 | 5,140 | 154 | 14,560 | 437 |
|  | 85+ | 8,800 | 246 | 22,140 | 619 |
| Race | White | 28,300 | 128 | 73,460 | 334 |
|  | Black | 2,180 | 117 | 7,980 | 429 |
|  | Other | 2,340 | 80 | 7,360 | 252 |
| Sex | Female | 21,560 | 144 | 54,480 | 364 |
|  | Male | 11,260 | 95 | 34,320 | 290 |
| Region | Northeast | 6,260 | 132 | 17,020 | 358 |
|  | Midwest | 8,080 | 133 | 21,580 | 356 |
|  | South | 13,400 | 126 | 34,560 | 326 |
|  | West | 5,080 | 94 | 15,640 | 290 |
| Total |  | 32,820 | 122 | 88,800 | 331 |

Source: Centers for Medicare & Medicaid Services, 5% Denominator and Claims Files. Beneficiaries aged 65+ years with continuous and full Parts A and B enrollment and no Health Maintenance Organization enrollment during the year. Unweighted counts were multiplied by 20 to produce counts for this table.

Among Medicare beneficiaries, hospital discharge rates with Clostridium difficile (all-listed diagnoses) increased with age and were higher among women compared with men and Blacks compared with Whites (Table 240). Rates were highest in the Midwest, followed by the Northeast and South, and lowest in the West.

**Table 240:** Clostridium difficile: Hospital discharges with first-listed and all-listed diagnoses by age, race, sex and region among fee-for-service, age-eligible Medicare beneficiaries, 2019.

| Demographic Characteristics |  | First-Listed Diagnosis |  | All-Listed Diagnoses |  |
| --- | --- | --- | --- | --- | --- |
|  |  | Number of events | Event rate per 100,000 | Number of events | Event rate per 100,000 |
| Age (years) | 65 to 69 | 4,420 | 55 | 18,420 | 228 |
|  | 70 to 74 | 5,400 | 78 | 19,940 | 287 |
|  | 75 to 79 | 5,900 | 121 | 19,660 | 402 |
|  | 80 to 84 | 4,240 | 127 | 15,420 | 462 |
|  | 85+ | 7,740 | 216 | 22,320 | 624 |
| Race | White | 24,140 | 110 | 78,640 | 357 |
|  | Black | 1,860 | 100 | 8,900 | 478 |
|  | Other | 1,700 | 58 | 8,220 | 281 |
| Sex | Female | 18,220 | 122 | 57,100 | 381 |
|  | Male | 9,480 | 80 | 38,660 | 327 |
| Region | Northeast | 4,880 | 103 | 17,520 | 369 |
|  | Midwest | 7,220 | 119 | 23,820 | 392 |
|  | South | 11,840 | 112 | 38,560 | 364 |
|  | West | 3,760 | 70 | 15,860 | 294 |
| Total |  | 27,700 | 103 | 95,760 | 357 |

Source: Centers for Medicare & Medicaid Services, 5% Denominator and Claims Files. Beneficiaries aged 65+ years with continuous and full Parts A and B enrollment and no Health Maintenance Organization enrollment during the year. Unweighted counts were multiplied by 20 to produce counts for this table.

Esophageal cancer accounted for 16,000 incident cases in 2016 (Table 241). Esophageal cancer was rare among children and adolescents and incidence rates increased with age throughout adulthood. Age-adjusted incidence rates were much higher among men compared with women, and higher among Whites compared with Blacks, and non-Hispanics compared with Hispanics.

**Table 241:**
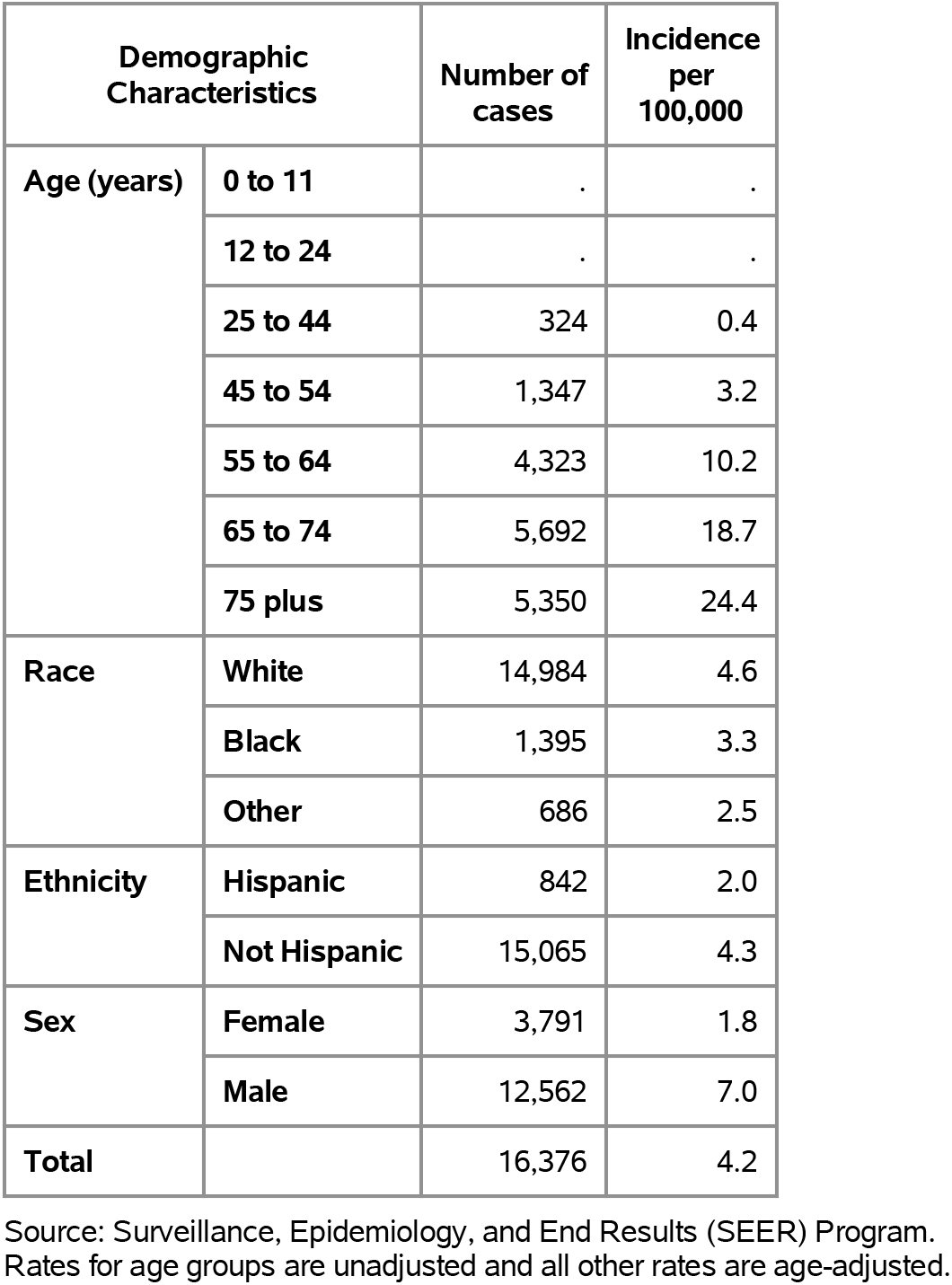
Esophageal Cancer: Incidence rates by age, race, ethnicity, and sex, 2016.

Esophageal cancer accounted contributed to 40,000 hospital discharges in 2018 (Table 242). Esophageal cancer hospitalization was rare among children and adolescents and hospital discharge rates increased with age throughout adulthood. Age-adjusted hospital discharge rates (all-listed diagnoses) were much higher among men compared with women, and higher among Whites compared with Blacks, and non-Hispanics compared with Hispanics.

**Table 242:**
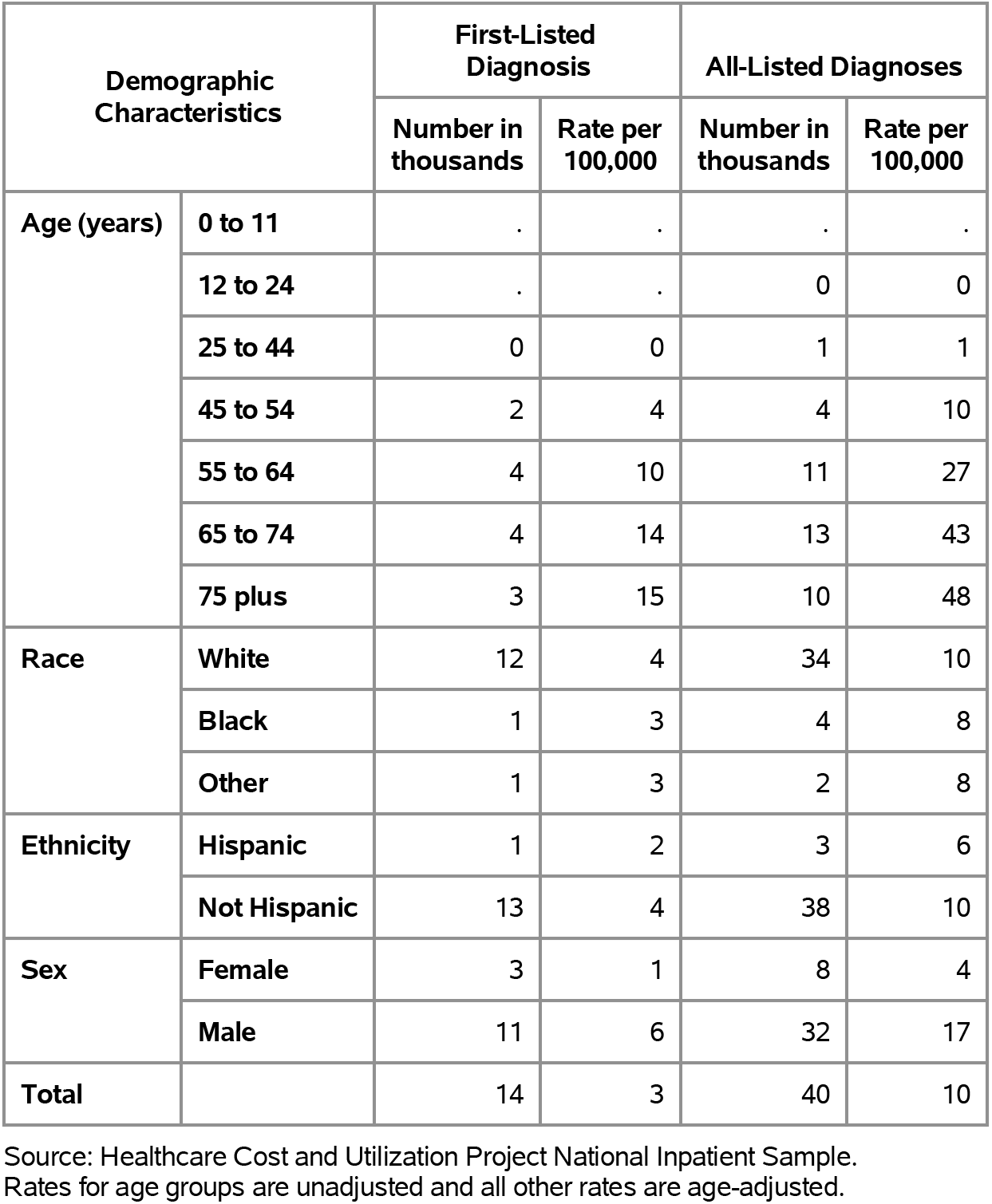
Esophageal Cancer: Hospital discharges with first-listed and all-listed diagnoses by age, race, ethnicity, and sex in the United States, 2018.

| Demographic Characteristics |  | First-Listed Diagnosis |  | All-Listed Diagnoses |  |
| --- | --- | --- | --- | --- | --- |
|  |  | Number in thousands | Rate per 100,000 | Number in thousands | Rate per 100,000 |
| Age (years) | 0 to 11 | . | . | . | . |
|  | 12 to 24 | . | . | 0 | 0 |
|  | 25 to 44 | 0 | 0 | 1 | 1 |
|  | 45 to 54 | 2 | 4 | 4 | 10 |
|  | 55 to 64 | 4 | 10 | 11 | 27 |
|  | 65 to 74 | 4 | 14 | 13 | 43 |
|  | 75 plus | 3 | 15 | 10 | 48 |
| Race | White | 12 | 4 | 34 | 10 |
|  | Black | 1 | 3 | 4 | 8 |
|  | Other | 1 | 3 | 2 | 8 |
| Ethnicity | Hispanic | 1 | 2 | 3 | 6 |
|  | Not Hispanic | 13 | 4 | 38 | 10 |
| Sex | Female | 3 | 1 | 8 | 4 |
|  | Male | 11 | 6 | 32 | 17 |
| Total |  | 14 | 3 | 40 | 10 |

Source: Healthcare Cost and Utilization Project National Inpatient Sample. Rates for age groups are unadjusted and all other rates are age-adjusted.

Esophageal cancer contributed to 18,000 deaths in 2019 (Table 243). Esophageal cancer mortality was rare among children and adolescents and mortality rates increased with age throughout adulthood. Age-adjusted mortality rates (underlying or other cause) were much higher among men compared with women, and higher among Whites compared with Blacks, and non-Hispanics compared with Hispanics.

**Table 243:**
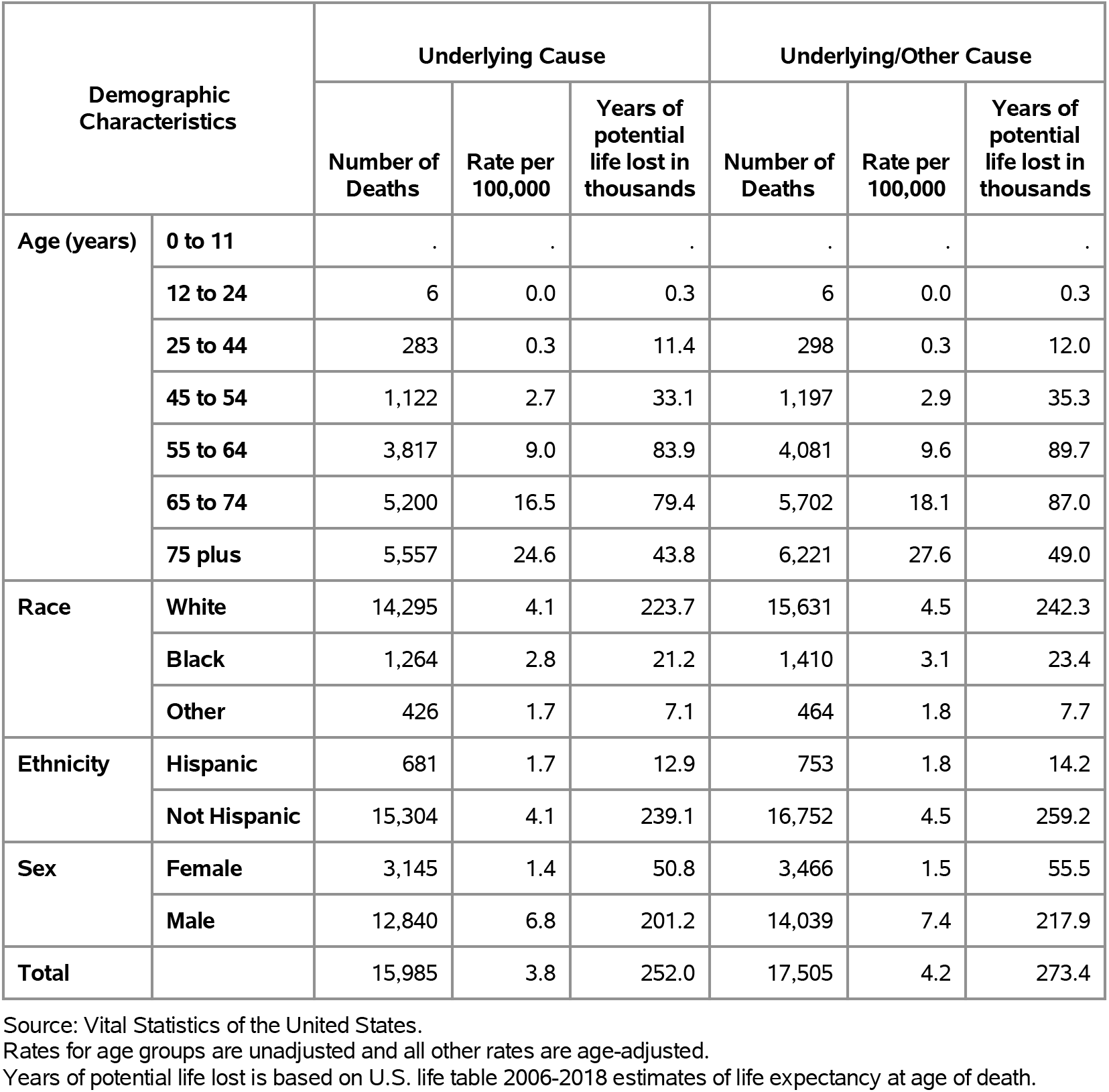
Esophageal Cancer: Deaths with underlying or underlying/other cause and lifetime years of life lost by age, race, ethnicity, and sex in the United States, 2019.

| Demographic Characteristics |  | Underlying Cause |  |  | Underlying/Other Cause |  |  |
| --- | --- | --- | --- | --- | --- | --- | --- |
|  |  | Number of Deaths | Rate per 100,000 | Years of potential life lost in thousands | Number of Deaths | Rate per 100,000 | Years of potential life lost in thousands |
| Age (years) | 0 to 11 | . | . | . | . | . | . |
|  | 12 to 24 | 6 | 0.0 | 0.3 | 6 | 0.0 | 0.3 |
|  | 25 to 44 | 283 | 0.3 | 11.4 | 298 | 0.3 | 12.0 |
|  | 45 to 54 | 1,122 | 2.7 | 33.1 | 1,197 | 2.9 | 35.3 |
|  | 55 to 64 | 3,817 | 9.0 | 83.9 | 4,081 | 9.6 | 89.7 |
|  | 65 to 74 | 5,200 | 16.5 | 79.4 | 5,702 | 18.1 | 87.0 |
|  | 75 plus | 5,557 | 24.6 | 43.8 | 6,221 | 27.6 | 49.0 |
| Race | White | 14,295 | 4.1 | 223.7 | 15,631 | 4.5 | 242.3 |
|  | Black | 1,264 | 2.8 | 21.2 | 1,410 | 3.1 | 23.4 |
|  | Other | 426 | 1.7 | 7.1 | 464 | 1.8 | 7.7 |
| Ethnicity | Hispanic | 681 | 1.7 | 12.9 | 753 | 1.8 | 14.2 |
|  | Not Hispanic | 15,304 | 4.1 | 239.1 | 16,752 | 4.5 | 259.2 |
| Sex | Female | 3,145 | 1.4 | 50.8 | 3,466 | 1.5 | 55.5 |
|  | Male | 12,840 | 6.8 | 201.2 | 14,039 | 7.4 | 217.9 |
| Total |  | 15,985 | 3.8 | 252.0 | 17,505 | 4.2 | 273.4 |

Source: Vital Statistics of the United States.
Rates for age groups are unadjusted and all other rates are age-adjusted.
Years of potential life lost is based on U.S. life table 2006-2018 estimates of life expectancy at age of death.

Gastric cancer accounted for 24,000 incident cases in 2016 (Table 244). Gastric cancer was rare among children and adolescents and incidence rates increased with age throughout adulthood. Age-adjusted incidence rates were higher among men compared with women, Blacks compared with Whites, and Hispanics compared with non-Hispanics.

**Table 244:** Gastric Cancer: Incidence rates by age, race, ethnicity, and sex, 2016.

| Demographic Characteristics |  | Number of cases | Incidence per 100,000 |
| --- | --- | --- | --- |
| Age (years) | 0 to 11 | . | . |
|  | 12 to 24 | . | 0.2 |
|  | 25 to 44 | 1,346 | 1.5 |
|  | 45 to 54 | 2,262 | 5.4 |
|  | 55 to 64 | 5,412 | 12.8 |
|  | 65 to 74 | 7,184 | 23.6 |
|  | 75 plus | 8,093 | 36.9 |
| Race | White | 17,077 | 5.4 |
|  | Black | 3,483 | 8.5 |
|  | Other | 2,422 | 9.1 |
| Ethnicity | Hispanic | 3,486 | 8.6 |
|  | Not Hispanic | 19,874 | 5.9 |
| Sex | Female | 9,182 | 4.5 |
|  | Male | 14,343 | 8.2 |
| Total |  | 23,539 | 6.2 |

Source: Surveillance, Epidemiology, and End Results (SEER) Program. Rates for age groups are unadjusted and all other rates are age-adjusted.

Gastric cancer contributed to 41,000 hospital discharges in 2018 (Table 245). Gastric cancer hospitalization was rare among children and adolescents and hospital discharge rates increased with age throughout adulthood. Age-adjusted hospital discharges rates (all-listed diagnoses) were higher among men compared with women, Blacks compared with Whites, and Hispanics compared with non-Hispanics.

**Table 245:** Gastric Cancer: Hospital discharges with first-listed and all-listed diagnoses by age, race, ethnicity, and sex in the United States, 2018.

| Demographic Characteristics |  | First-Listed Diagnosis |  | All-Listed Diagnoses |  |
| --- | --- | --- | --- | --- | --- |
|  |  | Number in thousands | Rate per 100,000 | Number in thousands | Rate per 100,000 |
| Age (years) | 0 to 11 | . | . | 0 | 0 |
|  | 12 to 24 | 0 | 0 | 0 | 0 |
|  | 25 to 44 | 1 | 2 | 3 | 4 |
|  | 45 to 54 | 2 | 5 | 5 | 12 |
|  | 55 to 64 | 5 | 11 | 10 | 24 |
|  | 65 to 74 | 5 | 16 | 11 | 37 |
|  | 75 plus | 5 | 24 | 12 | 53 |
| Race | White | 13 | 4 | 29 | 9 |
|  | Black | 3 | 7 | 7 | 16 |
|  | Other | 3 | 11 | 5 | 22 |
| Ethnicity | Hispanic | 4 | 8 | 7 | 17 |
|  | Not Hispanic | 15 | 4 | 34 | 10 |
| Sex | Female | 7 | 3 | 15 | 7 |
|  | Male | 12 | 7 | 26 | 14 |
| Total |  | 19 | 5 | 41 | 11 |

Source: Healthcare Cost and Utilization Project National Inpatient Sample. Rates for age groups are unadjusted and all other rates are age-adjusted.

Gastric cancer contributed to 12,000 deaths in 2019 (Table 246). Gastric cancer mortality was rare among children and adolescents and mortality rates increased with age throughout adulthood. Age-adjusted mortality rates (underlying or other cause) were higher among men compared with women, Blacks compared with Whites, and Hispanics compared with non-Hispanics.

**Table 246:** Gastric Cancer: Deaths with underlying or underlying/other cause and lifetime years of life lost by age, race, ethnicity, and sex in the United States, 2019.

| Demographic Characteristics |  | Underlying Cause |  |  | Underlying/Other Cause |  |  |
| --- | --- | --- | --- | --- | --- | --- | --- |
|  |  | Number of Deaths | Rate per 100,000 | Years of potential life lost in thousands | Number of Deaths | Rate per 100,000 | Years of potential life lost in thousands |
| Age (years) | 0 to 11 | . | . | . | . | . | . |
|  | 12 to 24 | 21 | 0.0 | 1.2 | 23 | 0.0 | 1.3 |
|  | 25 to 44 | 559 | 0.6 | 24.0 | 583 | 0.7 | 25.1 |
|  | 45 to 54 | 1,021 | 2.5 | 32.1 | 1,055 | 2.6 | 33.2 |
|  | 55 to 64 | 2,074 | 4.9 | 47.2 | 2,208 | 5.2 | 50.2 |
|  | 65 to 74 | 2,925 | 9.3 | 45.7 | 3,149 | 10.0 | 49.1 |
|  | 75 plus | 4,513 | 20.0 | 34.3 | 5,039 | 22.3 | 38.2 |
| Race | White | 8,137 | 2.5 | 136.3 | 8,829 | 2.7 | 145.4 |
|  | Black | 1,941 | 4.6 | 30.5 | 2,114 | 5.0 | 32.9 |
|  | Other | 1,035 | 4.1 | 17.9 | 1,114 | 4.5 | 18.8 |
| Ethnicity | Hispanic | 1,709 | 4.1 | 38.4 | 1,828 | 4.4 | 40.5 |
|  | Not Hispanic | 9,404 | 2.6 | 146.3 | 10,229 | 2.8 | 156.7 |
| Sex | Female | 4,478 | 2.1 | 79.1 | 4,827 | 2.2 | 84.1 |
|  | Male | 6,635 | 3.7 | 105.6 | 7,230 | 4.0 | 113.1 |
| Total |  | 11,113 | 2.8 | 184.7 | 12,057 | 3.0 | 197.2 |

Source: Vital Statistics of the United States.
Rates for age groups are unadjusted and all other rates are age-adjusted.
Years of potential life lost is based on U.S. life table 2006-2018 estimates of life expectancy at age of death.

Cancer of the small intestine accounted for 9,000 incident cases in 2016 (Table 247). Small intestinal cancer was rare among children and adolescents and incidence rates increased with age throughout adulthood. Age-adjusted incidence rates were higher among men compared with women, Blacks compared with Whites, and among non-Hispanics compared with Hispanics. Between 2004 and 2016, age-adjusted incidence rates (per 100,000) increased by a third from 1.8 to 2.4.(4)

**Table 247:** Cancer of Small Intestine: Incidence rates by age, race, ethnicity, and sex, 2016.

| Demographic Characteristics |  | Number of cases | Incidence per 100,000 |
| --- | --- | --- | --- |
| Age (years) | 0 to 11 | . | . |
|  | 12 to 24 | . | 0.0 |
|  | 25 to 44 | 577 | 0.7 |
|  | 45 to 54 | 1,168 | 2.8 |
|  | 55 to 64 | 2,081 | 4.9 |
|  | 65 to 74 | 2,873 | 9.4 |
|  | 75 plus | 2,630 | 12.0 |
| Race | White | 7,256 | 2.3 |
|  | Black | 1,607 | 3.8 |
|  | Other | 455 | 1.7 |
| Ethnicity | Hispanic | 674 | 1.6 |
|  | Not Hispanic | 8,157 | 2.5 |
| Sex | Female | 4,247 | 2.1 |
|  | Male | 4,795 | 2.7 |
| Total |  | 9,044 | 2.4 |

Source: Surveillance, Epidemiology, and End Results (SEER) Program. Rates for age groups are unadjusted and all other rates are age-adjusted.

Cancer of the small intestine contributed to 9,000 hospital discharges in 2018 (Table 248). Small intestinal cancer hospitalization was rare among children and adolescents and hospital discharge rates increased with age throughout adulthood. Age-adjusted hospital discharge rates (all-listed diagnoses) were higher among men compared with women and Blacks compared with Whites but did not differ by ethnicity. Between 2004 and 2018, age-adjusted hospital discharge rates (per 100,000) with an all-listed diagnosis decreased by a third from 3 to 2.(4)

**Table 248:** Cancer of Small Intestine: Hospital discharges with first-listed and all-listed diagnoses by age, race, ethnicity, and sex in the United States, 2018.

| Demographic Characteristics |  | First-Listed Diagnosis |  | All-Listed Diagnoses |  |
| --- | --- | --- | --- | --- | --- |
|  |  | Number in thousands | Rate per 100,000 | Number in thousands | Rate per 100,000 |
| Age (years) | 0 to 11 | . | . | . | . |
|  | 12 to 24 | 0 | 0 | 0 | 0 |
|  | 25 to 44 | 0 | 0 | 0 | 1 |
|  | 45 to 54 | 0 | 1 | 1 | 3 |
|  | 55 to 64 | 1 | 2 | 2 | 5 |
|  | 65 to 74 | 1 | 4 | 3 | 9 |
|  | 75 plus | 1 | 6 | 3 | 13 |
| Race | White | 3 | 1 | 7 | 2 |
|  | Black | 1 | 2 | 2 | 4 |
|  | Other | 0 | 1 | 1 | 3 |
| Ethnicity | Hispanic | 0 | 1 | 1 | 2 |
|  | Not Hispanic | 4 | 1 | 9 | 2 |
| Sex | Female | 2 | 1 | 4 | 2 |
|  | Male | 2 | 1 | 5 | 3 |
| Total |  | 4 | 1 | 9 | 2 |

Source: Healthcare Cost and Utilization Project National Inpatient Sample. Rates for age groups are unadjusted and all other rates are age-adjusted.

Cancer of the small intestine contributed to 2,000 deaths in 2019 (Table 249). Small intestinal cancer mortality was rare among children and adolescents and mortality rates increased with age throughout adulthood. Age-adjusted mortality rates (underlying or other cause) were higher among men compared with women, Blacks compared with Whites, and non-Hispanics compared with Hispanics. Between 2004 and 2018, age-adjusted hospital discharge rates (per 100,000) with an all-listed diagnosis increased by a fourth from 0.4 to 0.5.(4)

**Table 249:** Cancer of Small Intestine: Deaths with underlying or underlying/other cause and lifetime years of life lost by age, race, ethnicity, and sex in the United States, 2019.

| Demographic Characteristics |  | Underlying Cause |  |  | Underlying/Other Cause |  |  |
| --- | --- | --- | --- | --- | --- | --- | --- |
|  |  | Number of Deaths | Rate per 100,000 | Years of potential life lost in thousands | Number of Deaths | Rate per 100,000 | Years of potential life lost in thousands |
| Age (years) | 0 to 11 | . | . | . | . | . | . |
|  | 12 to 24 | 3 | 0.0 | 0.2 | 3 | 0.0 | 0.2 |
|  | 25 to 44 | 51 | 0.1 | 2.1 | 54 | 0.1 | 2.3 |
|  | 45 to 54 | 107 | 0.3 | 3.3 | 119 | 0.3 | 3.6 |
|  | 55 to 64 | 286 | 0.7 | 6.5 | 315 | 0.7 | 7.1 |
|  | 65 to 74 | 490 | 1.6 | 7.7 | 542 | 1.7 | 8.5 |
|  | 75 plus | 771 | 3.4 | 5.9 | 883 | 3.9 | 6.8 |
| Race | White | 1,362 | 0.4 | 20.1 | 1,528 | 0.4 | 22.3 |
|  | Black | 277 | 0.6 | 4.6 | 308 | 0.7 | 5.2 |
|  | Other | 69 | 0.3 | 1.0 | 80 | 0.3 | 1.1 |
| Ethnicity | Hispanic | 100 | 0.3 | 1.8 | 120 | 0.3 | 2.2 |
|  | Not Hispanic | 1,608 | 0.4 | 23.9 | 1,796 | 0.5 | 26.3 |
| Sex | Female | 782 | 0.3 | 12.4 | 867 | 0.4 | 13.6 |
|  | Male | 926 | 0.5 | 13.3 | 1,049 | 0.6 | 14.9 |
| Total |  | 1,708 | 0.4 | 25.7 | 1,916 | 0.5 | 28.5 |

Source: Vital Statistics of the United States.
Rates for age groups are unadjusted and all other rates are age-adjusted.
Years of potential life lost is based on U.S. life table 2006-2018 estimates of life expectancy at age of death.

Colorectal cancer accounted for 132,000 incident cases in 2016 (Table 250). Colorectal cancer was rare among children and incidence rates increased with age throughout adulthood. Age-adjusted incidence rates were higher among men compared with women, Blacks compared with Whites, and non-Hispanics compared with Hispanics. Between 2004 and 2016, age-adjusted incidence rates (per 100,000) decreased by 26% from 47.5 to 35.0 (4)

**Table 250:** Colorectal Cancer: Incidence rates by age, race, ethnicity, and sex, 2016.

| Demographic Characteristics |  | Number of cases | Incidence per 100,000 |
| --- | --- | --- | --- |
| Age (years) | 0 to 11 | . | 0.0 |
|  | 12 to 24 | 877 | 1.6 |
|  | 25 to 44 | 8,814 | 10.1 |
|  | 45 to 54 | 19,593 | 47.1 |
|  | 55 to 64 | 29,710 | 70.3 |
|  | 65 to 74 | 33,730 | 110.6 |
|  | 75 plus | 43,342 | 197.6 |
| Race | White | 106,708 | 34.3 |
|  | Black | 16,878 | 40.6 |
|  | Other | 9,409 | 34.7 |
| Ethnicity | Hispanic | 13,543 | 33.0 |
|  | Not Hispanic | 115,855 | 35.3 |
| Sex | Female | 63,289 | 31.2 |
|  | Male | 68,385 | 39.4 |
| Total |  | 131,692 | 35.0 |

Source: Surveillance, Epidemiology, and End Results (SEER) Program. Rates for age groups are unadjusted and all other rates are age-adjusted.

Colorectal cancer contributed to 250,000 hospital discharges in 2018 (Table 251). Colorectal cancer hospitalization was rare among children and hospital discharge rates increased with age throughout adulthood. Age-adjusted hospital discharge rates (all-listed diagnoses) were higher among men compared with women, Blacks compared with Whites, and non-Hispanics compared with Hispanics. Between 2004 and 2018, age-adjusted hospital discharge rates (per 100,000) with an all-listed diagnosis decreased by 26% from 87 to 64.(4,6)

**Table 251:** Colorectal Cancer: Hospital discharges with first-listed and all-listed diagnoses by age, race, ethnicity, and sex in the United States, 2018.

| Demographic Characteristics |  | First-Listed Diagnosis |  | All-Listed Diagnoses |  |
| --- | --- | --- | --- | --- | --- |
|  |  | Number in thousands | Rate per 100,000 | Number in thousands | Rate per 100,000 |
| Age (years) | 0 to 11 | 0 | 0 | 0 | 0 |
|  | 12 to 24 | 0 | 0 | 0 | 1 |
|  | 25 to 44 | 7 | 8 | 16 | 18 |
|  | 45 to 54 | 18 | 44 | 36 | 87 |
|  | 55 to 64 | 29 | 68 | 59 | 139 |
|  | 65 to 74 | 33 | 108 | 66 | 215 |
|  | 75 plus | 38 | 175 | 73 | 333 |
| Race | White | 103 | 32 | 202 | 63 |
|  | Black | 15 | 34 | 32 | 74 |
|  | Other | 9 | 36 | 18 | 71 |
| Ethnicity | Hispanic | 12 | 29 | 25 | 59 |
|  | Not Hispanic | 114 | 33 | 227 | 66 |
| Sex | Female | 59 | 28 | 117 | 56 |
|  | Male | 66 | 37 | 133 | 74 |
| Total |  | 126 | 32 | 250 | 64 |

Source: Healthcare Cost and Utilization Project National Inpatient Sample. Rates for age groups are unadjusted and all other rates are age-adjusted.

Colorectal cancer contributed to 60,000 deaths in 2019 (Table 252). Colorectal cancer mortality was rare among children and mortality rates increased with age throughout adulthood. Age-adjusted mortality rates (underlying or other cause) were higher among men compared with women, Blacks compared with Whites, and non-Hispanics compared with Hispanics. Between 2004 and 2019, age-adjusted mortality rates (per 100,000) with colorectal cancer as underlying or other cause decreased by 31% from 21.4 to 14.7.(4)

**Table 252:** Colorectal Cancer: Deaths with underlying or underlying/other cause and lifetime years of life lost by age, race, ethnicity, and sex in the United States, 2019.

| Demographic Characteristics |  | Underlying Cause |  |  | Underlying/Other Cause |  |  |
| --- | --- | --- | --- | --- | --- | --- | --- |
|  |  | Number of Deaths | Rate per 100,000 | Years of potential life lost in thousands | Number of Deaths | Rate per 100,000 | Years of potential life lost in thousands |
| Age (years) | 0 to 11 | . | . | . | . | . | . |
|  | 12 to 24 | 38 | 0.1 | 2.2 | 42 | 0.1 | 2.4 |
|  | 25 to 44 | 1,708 | 1.9 | 71.0 | 1,772 | 2.0 | 73.7 |
|  | 45 to 54 | 4,734 | 11.6 | 146.4 | 4,988 | 12.2 | 154.1 |
|  | 55 to 64 | 10,048 | 23.7 | 229.2 | 10,845 | 25.5 | 247.1 |
|  | 65 to 74 | 12,684 | 40.3 | 199.8 | 14,354 | 45.6 | 225.4 |
|  | 75 plus | 22,494 | 99.6 | 164.0 | 27,798 | 123.1 | 199.0 |
| Race | White | 42,411 | 12.6 | 654.6 | 49,354 | 14.6 | 728.6 |
|  | Black | 7,024 | 16.3 | 117.8 | 7,907 | 18.5 | 129.6 |
|  | Other | 2,271 | 9.0 | 40.1 | 2,538 | 10.1 | 43.4 |
| Ethnicity | Hispanic | 3,654 | 8.8 | 74.4 | 4,063 | 9.9 | 80.6 |
|  | Not Hispanic | 48,052 | 13.3 | 738.1 | 55,736 | 15.3 | 821.1 |
| Sex | Female | 24,124 | 10.7 | 377.1 | 27,678 | 12.2 | 415.7 |
|  | Male | 27,582 | 15.2 | 435.4 | 32,121 | 17.8 | 486.0 |
| Total |  | 51,706 | 12.8 | 812.5 | 59,799 | 14.7 | 901.6 |

Source: Vital Statistics of the United States.
Rates for age groups are unadjusted and all other rates are age-adjusted.
Years of potential life lost is based on U.S. life table 2006-2018 estimates of life expectancy at age of death.

Primary liver cancer accounted for 26,000 incident cases in 2016 (Table 253). Primary liver cancer was uncommon in childhood through young adulthood and incidence rates peaked among persons 65-74 years. Age-adjusted incidence rates were higher among men compared with women, Blacks compared with Whites, and Hispanics compared with non-Hispanics.

**Table 253:**
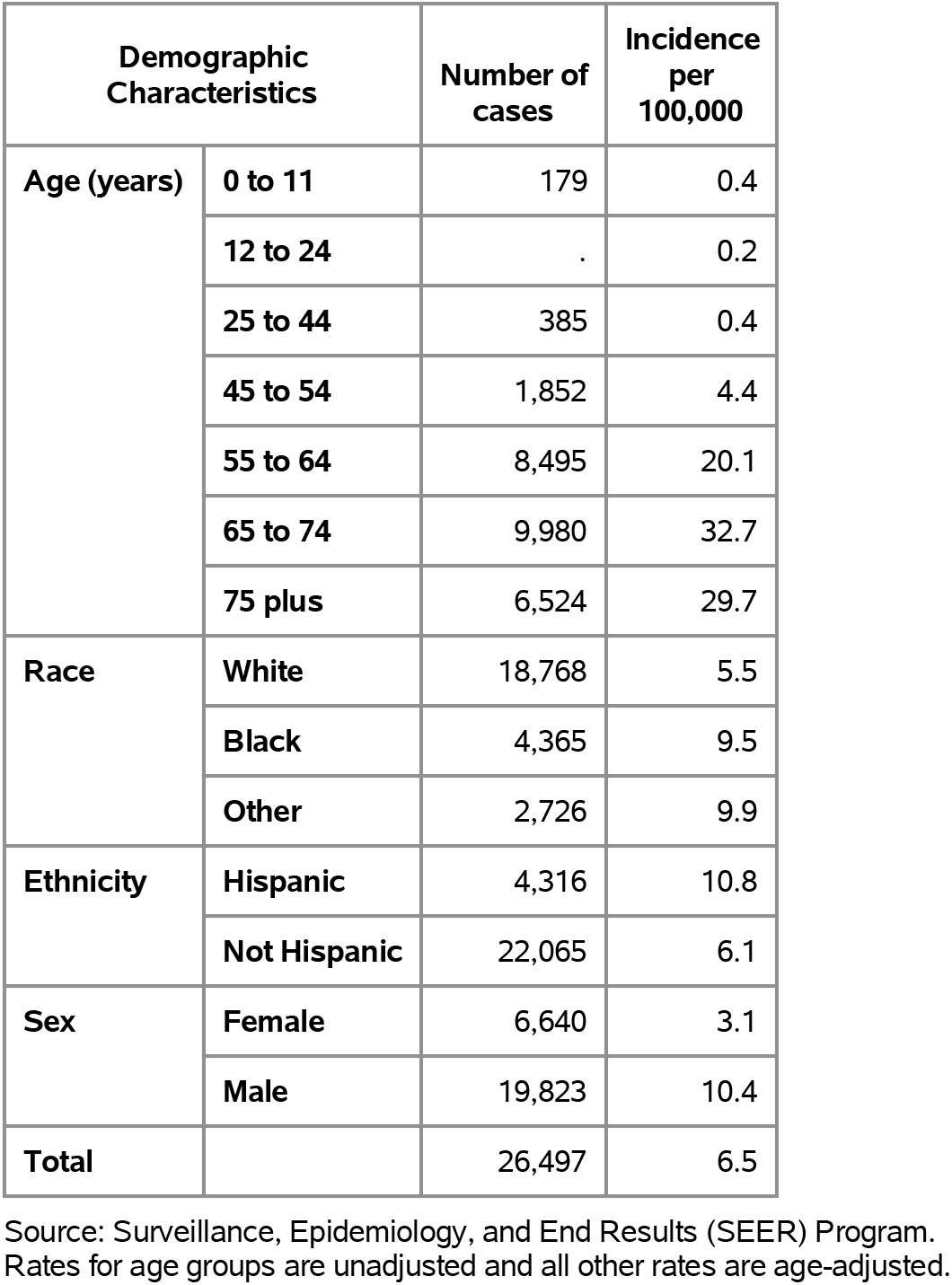
Primary Liver Cancer: Incidence rates by age, race, ethnicity, and sex, 2016.

Primary liver cancer contributed to 65,000 hospital discharges in 2018 (Table 254). Primary liver cancer hospitalization was uncommon in childhood through young adulthood and rates peaked among persons 65-74 years. Age-adjusted hospital discharge rates (all-listed diagnoses) were higher among men compared with women, Blacks compared with Whites, and Hispanics compared with non-Hispanics.

**Table 254:** Primary Liver Cancer: Hospital discharges with first-listed and all-listed diagnoses by age, race, ethnicity, and sex in the United States, 2018.

| Demographic Characteristics |  | First-Listed Diagnosis |  | All-Listed Diagnoses |  |
| --- | --- | --- | --- | --- | --- |
|  |  | Number in thousands | Rate per 100,000 | Number in thousands | Rate per 100,000 |
| Age (years) | 0 to 11 | 0 | 1 | 2 | 4 |
|  | 12 to 24 | 0 | 0 | 0 | 1 |
|  | 25 to 44 | 0 | 0 | 2 | 2 |
|  | 45 to 54 | 1 | 3 | 6 | 14 |
|  | 55 to 64 | 6 | 13 | 23 | 54 |
|  | 65 to 74 | 5 | 16 | 21 | 67 |
|  | 75 plus | 3 | 14 | 13 | 58 |
| Race | White | 11 | 3 | 49 | 14 |
|  | Black | 2 | 5 | 10 | 20 |
|  | Other | 2 | 9 | 7 | 28 |
| Ethnicity | Hispanic | 3 | 6 | 11 | 26 |
|  | Not Hispanic | 13 | 4 | 55 | 15 |
| Sex | Female | 4 | 2 | 18 | 8 |
|  | Male | 11 | 6 | 47 | 24 |
| Total |  | 16 | 4 | 65 | 16 |

Source: Healthcare Cost and Utilization Project National Inpatient Sample. Rates for age groups are unadjusted and all other rates are age-adjusted.

Primary liver cancer contributed to 23,000 deaths in 2019 (Table 255). Primary liver cancer mortality rates increased with age. Age-adjusted mortality rates (underlying or other cause) were higher among men compared with women, Blacks compared with Whites, and Hispanics compared with non-Hispanics.

**Table 255:** Primary Liver Cancer: Deaths with underlying or underlying/other cause and lifetime years of life lost by age, race, ethnicity, and sex in the United States, 2019.

| Demographic Characteristics |  | Underlying Cause |  |  | Underlying/Other Cause |  |  |
| --- | --- | --- | --- | --- | --- | --- | --- |
|  |  | Number of Deaths | Rate per 100,000 | Years of potential life lost in thousands | Number of Deaths | Rate per 100,000 | Years of potential life lost in thousands |
| Age (years) | 0 to 11 | 28 | 0.1 | 2.1 | 32 | 0.1 | 2.4 |
|  | 12 to 24 | 29 | 0.1 | 1.7 | 30 | 0.1 | 1.8 |
|  | 25 to 44 | 233 | 0.3 | 9.6 | 260 | 0.3 | 10.7 |
|  | 45 to 54 | 963 | 2.4 | 28.4 | 1,110 | 2.7 | 32.8 |
|  | 55 to 64 | 5,547 | 13.1 | 121.0 | 6,235 | 14.7 | 136.2 |
|  | 65 to 74 | 7,041 | 22.4 | 110.0 | 7,957 | 25.3 | 124.2 |
|  | 75 plus | 6,353 | 28.1 | 51.1 | 7,051 | 31.2 | 56.7 |
| Race | White | 15,745 | 4.4 | 250.6 | 17,711 | 4.9 | 283.0 |
|  | Black | 2,971 | 6.2 | 49.2 | 3,314 | 6.9 | 54.9 |
|  | Other | 1,478 | 5.8 | 24.2 | 1,650 | 6.5 | 26.9 |
| Ethnicity | Hispanic | 2,410 | 5.8 | 44.9 | 2,722 | 6.5 | 51.3 |
|  | Not Hispanic | 17,784 | 4.6 | 279.1 | 19,953 | 5.1 | 313.6 |
| Sex | Female | 5,513 | 2.4 | 85.9 | 6,118 | 2.6 | 95.9 |
|  | Male | 14,681 | 7.4 | 238.1 | 16,557 | 8.3 | 269.0 |
| Total |  | 20,194 | 4.7 | 324.0 | 22,675 | 5.3 | 364.9 |

Source: Vital Statistics of the United States.
Rates for age groups are unadjusted and all other rates are age-adjusted.
Years of potential life lost is based on U.S. life table 2006-2018 estimates of life expectancy at age of death.

Bile duct cancer accounted for 13,000 incident cases in 2016 (Table 256). Bile duct cancer was rare among children and adolescents and incidence rates increased with age throughout adulthood. Age-adjusted mortality rates (underlying or other cause) were higher among men compared with women, Whites compared with Blacks, and Hispanics compared with non-Hispanics. Between 2004 and 2016, age-adjusted incidence rates (per 100,000) increased by 43% from 2.3 to 3.3.(4)

**Table 256:** Bile Duct Cancer: Incidence rates by age, race, ethnicity, and sex, 2016.

| Demographic Characteristics |  | Number of cases | Incidence per 100,000 |
| --- | --- | --- | --- |
| Age (years) | 0 to 11 | . | 0.0 |
|  | 12 to 24 | . | . |
|  | 25 to 44 | 425 | 0.5 |
|  | 45 to 54 | 936 | 2.2 |
|  | 55 to 64 | 2,889 | 6.8 |
|  | 65 to 74 | 4,056 | 13.3 |
|  | 75 plus | 4,932 | 22.5 |
| Race | White | 10,361 | 3.2 |
|  | Black | 1,217 | 3.0 |
|  | Other | 1,090 | 4.0 |
| Ethnicity | Hispanic | 1,542 | 4.1 |
|  | Not Hispanic | 11,007 | 3.2 |
| Sex | Female | 5,878 | 2.8 |
|  | Male | 6,840 | 3.9 |
| Total |  | 12,720 | 3.3 |

Source: Surveillance, Epidemiology, and End Results (SEER) Program. Rates for age groups are unadjusted and all other rates are age-adjusted.

Bile duct cancer contributed to 34,000 hospital discharges in 2018 (Table 257). Bile duct cancer hospitalization was rare among children and adolescents and hospital discharge rates increased with age throughout adulthood. Age-adjusted hospital discharge rates (all-listed diagnoses) were higher among men compared with women and Hispanics compared with non-Hispanics but did not differ by race. Between 2004 and 2018, age-adjusted hospital discharge rates (per 100,000) with an all-listed diagnosis increased by a third from 6 to 8.(4)

**Table 257:** Bile Duct Cancer: Hospital discharges with first-listed and all-listed diagnoses by age, race, ethnicity, and sex in the United States, 2018.

| Demographic Characteristics |  | First-Listed Diagnosis |  | All-Listed Diagnoses |  |
| --- | --- | --- | --- | --- | --- |
|  |  | Number in thousands | Rate per 100,000 | Number in thousands | Rate per 100,000 |
| Age (years) | 0 to 11 | . | . | 0 | 0 |
|  | 12 to 24 | 0 | 0 | 0 | 0 |
|  | 25 to 44 | 0 | 0 | 2 | 2 |
|  | 45 to 54 | 1 | 3 | 3 | 8 |
|  | 55 to 64 | 3 | 7 | 8 | 19 |
|  | 65 to 74 | 4 | 12 | 11 | 35 |
|  | 75 plus | 3 | 16 | 10 | 44 |
| Race | White | 9 | 3 | 27 | 8 |
|  | Black | 1 | 3 | 3 | 8 |
|  | Other | 1 | 5 | 4 | 14 |
| Ethnicity | Hispanic | 1 | 4 | 4 | 10 |
|  | Not Hispanic | 10 | 3 | 30 | 8 |
| Sex | Female | 5 | 2 | 16 | 7 |
|  | Male | 6 | 3 | 18 | 10 |
| Total |  | 11 | 3 | 34 | 8 |

Source: Healthcare Cost and Utilization Project National Inpatient Sample. Rates for age groups are unadjusted and all other rates are age-adjusted.

Bile duct cancer contributed to 11,000 deaths in 2019 (Table 258). Bile duct cancer mortality was rare among children and adolescents and mortality rates increased with age throughout adulthood. Age-adjusted mortality rates (underlying or other cause) were higher among men compared with women, Whites compared with Blacks, and non-Hispanics compared with Hispanics. Between 2004 and 2019, age-adjusted mortality rates (per 100,000) with bile duct cancer as underlying or other cause increased by 44% from 1.8 to 2.6.(4)

**Table 258:** Bile Duct Cancer: Deaths with underlying or underlying/other cause and lifetime years of life lost by age, race, ethnicity, and sex in the United States, 2019.

| Demographic Characteristics |  | Underlying Cause |  |  | Underlying/Other Cause |  |  |
| --- | --- | --- | --- | --- | --- | --- | --- |
|  |  | Number of Deaths | Rate per 100,000 | Years of potential life lost in thousands | Number of Deaths | Rate per 100,000 | Years of potential life lost in thousands |
| Age (years) | 0 to 11 | . | . | . | . | . | . |
|  | 12 to 24 | 10 | 0.0 | 0.6 | 11 | 0.0 | 0.6 |
|  | 25 to 44 | 247 | 0.3 | 10.5 | 267 | 0.3 | 11.4 |
|  | 45 to 54 | 617 | 1.5 | 19.3 | 664 | 1.6 | 20.8 |
|  | 55 to 64 | 1,963 | 4.6 | 45.1 | 2,142 | 5.0 | 49.1 |
|  | 65 to 74 | 3,130 | 9.9 | 49.8 | 3,422 | 10.9 | 54.4 |
|  | 75 plus | 3,871 | 17.1 | 31.4 | 4,250 | 18.8 | 34.5 |
| Race | White | 8,175 | 2.4 | 129.5 | 8,942 | 2.6 | 141.2 |
|  | Black | 946 | 2.2 | 15.8 | 1,045 | 2.4 | 17.5 |
|  | Other | 717 | 2.9 | 11.4 | 769 | 3.1 | 12.0 |
| Ethnicity | Hispanic | 909 | 2.2 | 18.2 | 1,004 | 2.4 | 20.0 |
|  | Not Hispanic | 8,929 | 2.4 | 138.5 | 9,752 | 2.6 | 150.9 |
| Sex | Female | 4,756 | 2.1 | 78.4 | 5,147 | 2.3 | 84.7 |
|  | Male | 5,082 | 2.7 | 78.3 | 5,609 | 3.0 | 86.2 |
| Total |  | 9,838 | 2.4 | 156.7 | 10,756 | 2.6 | 170.8 |

Source: Vital Statistics of the United States.
Rates for age groups are unadjusted and all other rates are age-adjusted.
Years of potential life lost is based on U.S. life table 2006-2018 estimates of life expectancy at age of death.

Gallbladder cancer accounted for an estimated 4,000 incident cases in 2016 (Table 259). Gallbladder cancer was rare among children, adolescents, and younger adults and incidence rates increased with age throughout middle and older age. In contrast to all other common digestive cancers, age-adjusted incidence rates were higher among women compared with men. Incidence rates were higher among Blacks compared with Whites and among Hispanics compared with non-Hispanics. Since 2004, age-adjusted rates (per 100,000) for incidence have remained stable at 1.1.(4)

**Table 259:** Gallbladder Cancer: Incidence rates by age, race, ethnicity, and sex, 2016.

| Demographic Characteristics |  | Number of cases | Incidence per 100,000 |
| --- | --- | --- | --- |
| Age (years) | 0 to 11 | . | . |
|  | 12 to 24 | . | . |
|  | 25 to 44 | . | 0.1 |
|  | 45 to 54 | 326 | 0.8 |
|  | 55 to 64 | 604 | 1.4 |
|  | 65 to 74 | 1,315 | 4.3 |
|  | 75 plus | 2,009 | 9.2 |
| Race | White | 2,886 | 0.9 |
|  | Black | 748 | 1.9 |
|  | Other | 433 | 1.6 |
| Ethnicity | Hispanic | 583 | 1.6 |
|  | Not Hispanic | 3,551 | 1.1 |
| Sex | Female | 2,923 | 1.4 |
|  | Male | 1,254 | 0.8 |
| Total |  | 4,173 | 1.1 |

Source: Surveillance, Epidemiology, and End Results (SEER) Program. Rates for age groups are unadjusted and all other rates are age-adjusted.

Gallbladder cancer contributed to 8,000 hospital discharges in 2018 (Table 260). Gallbladder cancer hospitalization was rare among children, adolescents, and younger adults and hospital discharge rates increased with age throughout middle and older age. In contrast to all other common digestive cancers, age-adjusted hospital discharge rates (all-listed diagnoses) were higher among women compared with men. Hospital discharge rates were higher among Blacks compared with Whites and among Hispanics compared with non-Hispanics. Since 2004, age-adjusted hospital discharge rates have remained stable at 2.(4)

**Table 260:** Gallbladder Cancer: Hospital discharges with first-listed and all-listed diagnoses by age, race, ethnicity, and sex in the United States, 2018.

| Demographic Characteristics |  | First-Listed Diagnosis |  | All-Listed Diagnoses |  |
| --- | --- | --- | --- | --- | --- |
|  |  | Number in thousands | Rate per 100,000 | Number in thousands | Rate per 100,000 |
| Age (years) | 0 to 11 | . | . | 0 | 0 |
|  | 12 to 24 | . | . | . | . |
|  | 25 to 44 | 0 | 0 | 0 | 0 |
|  | 45 to 54 | 0 | 1 | 1 | 2 |
|  | 55 to 64 | 1 | 2 | 2 | 4 |
|  | 65 to 74 | 1 | 3 | 2 | 8 |
|  | 75 plus | 1 | 4 | 3 | 11 |
| Race | White | 2 | 1 | 5 | 2 |
|  | Black | 1 | 1 | 1 | 3 |
|  | Other | 0 | 1 | 1 | 4 |
| Ethnicity | Hispanic | 0 | 1 | 1 | 3 |
|  | Not Hispanic | 2 | 1 | 7 | 2 |
| Sex | Female | 2 | 1 | 5 | 2 |
|  | Male | 1 | 0 | 2 | 1 |
| Total |  | 3 | 1 | 8 | 2 |

Source: Healthcare Cost and Utilization Project National Inpatient Sample. Rates for age groups are unadjusted and all other rates are age-adjusted.

Gallbladder cancer contributed to 2,000 deaths in 2019 (Table 261). Gallbladder cancer mortality was rare among children, adolescents, and younger adults and mortality rates increased with age throughout middle and older age. In contrast to all other common digestive cancers, age-adjusted mortality rates (underlying or other cause) were higher among women compared with men. Mortality rates were higher among Blacks compared with Whites and among Hispanics compared with non-Hispanics. Between 2004 and 2019, age-adjusted mortality rates (per 100,000) with gallbladder cancer as underlying or other cause decreased by 14% from 0.7 to 0.6.(4)

**Table 261:** Gallbladder Cancer: Deaths with underlying or underlying/other cause and lifetime years of life lost by age, race, ethnicity, and sex in the United States, 2019.

| Demographic Characteristics |  | Underlying Cause |  |  | Underlying/Other Cause |  |  |
| --- | --- | --- | --- | --- | --- | --- | --- |
|  |  | Number of Deaths | Rate per 100,000 | Years of potential life lost in thousands | Number of Deaths | Rate per 100,000 | Years of potential life lost in thousands |
| Age (years) | 0 to 11 | . | . | . | . | . | . |
|  | 12 to 24 | . | . | . | . | . | . |
|  | 25 to 44 | 18 | 0.0 | 0.8 | 21 | 0.0 | 0.9 |
|  | 45 to 54 | 138 | 0.3 | 4.4 | 147 | 0.4 | 4.7 |
|  | 55 to 64 | 404 | 1.0 | 9.5 | 429 | 1.0 | 10.1 |
|  | 65 to 74 | 628 | 2.0 | 10.2 | 671 | 2.1 | 10.9 |
|  | 75 plus | 952 | 4.2 | 7.7 | 1,036 | 4.6 | 8.3 |
| Race | White | 1,614 | 0.5 | 24.4 | 1,732 | 0.5 | 26.1 |
|  | Black | 374 | 0.9 | 5.9 | 404 | 0.9 | 6.3 |
|  | Other | 152 | 0.6 | 2.3 | 168 | 0.7 | 2.6 |
| Ethnicity | Hispanic | 289 | 0.7 | 5.8 | 312 | 0.8 | 6.4 |
|  | Not Hispanic | 1,851 | 0.5 | 26.8 | 1,992 | 0.5 | 28.6 |
| Sex | Female | 1,436 | 0.6 | 22.9 | 1,537 | 0.7 | 24.4 |
|  | Male | 704 | 0.4 | 9.8 | 767 | 0.4 | 10.6 |
| Total |  | 2,140 | 0.5 | 32.6 | 2,304 | 0.6 | 34.9 |

Source: Vital Statistics of the United States.
Rates for age groups are unadjusted and all other rates are age-adjusted.
Years of potential life lost is based on U.S. life table 2006-2018 estimates of life expectancy at age of death.

Pancreatic cancer accounted for 52,000 incident cases in 2016 (Table 262). Pancreatic cancer was rare among children and adolescents and incidence rates increased with age throughout adulthood. Age-adjusted incidence rates were higher among men compared with women, Blacks compared with Whites, and non-Hispanics compared with Hispanics. Between 2004 and 2016, age-adjusted incidence rates (per 100,000) increased by 18% from 11.3 to 13.3 (4)

**Table 262:** Pancreatic Cancer: Incidence rates by age, race, ethnicity, and sex, 2016.

| Demographic Characteristics |  | Number of cases | Incidence per 100,000 |
| --- | --- | --- | --- |
| Age (years) | 0 to 11 | . | 0.0 |
|  | 12 to 24 | . | 0.1 |
|  | 25 to 44 | 1,194 | 1.4 |
|  | 45 to 54 | 3,851 | 9.3 |
|  | 55 to 64 | 11,352 | 26.9 |
|  | 65 to 74 | 17,152 | 56.3 |
|  | 75 plus | 20,125 | 91.7 |
| Race | White | 43,366 | 13.3 |
|  | Black | 6,430 | 15.6 |
|  | Other | 3,001 | 11.1 |
| Ethnicity | Hispanic | 4,925 | 12.8 |
|  | Not Hispanic | 45,662 | 13.3 |
| Sex | Female | 24,496 | 11.5 |
|  | Male | 27,063 | 15.4 |
| Total |  | 51,567 | 13.3 |

Source: Surveillance, Epidemiology, and End Results (SEER) Program. Rates for age groups are unadjusted and all other rates are age-adjusted.

Pancreatic cancer contributed to 110,000 hospital discharges in 2018 (Table 263). Pancreatic cancer hospitalization was rare among children and adolescents and hospital discharge rates increased with age throughout adulthood. Age-adjusted hospital discharge rates (all-listed diagnoses) were higher among men compared with women, Blacks compared with Whites, and non-Hispanics compared with Hispanics. Between 2004 and 2018, age-adjusted hospital discharge rates (per 100,000) with an all-listed diagnosis increased by 17% from 23 to 27.(4)

**Table 263:** Pancreatic Cancer: Hospital discharges with first-listed and all-listed diagnoses by age, race, ethnicity, and sex in the United States, 2018.

| Demographic Characteristics |  | First-Listed Diagnosis |  | All-Listed Diagnoses |  |
| --- | --- | --- | --- | --- | --- |
|  |  | Number in thousands | Rate per 100,000 | Number in thousands | Rate per 100,000 |
| Age (years) | 0 to 11 | 0 | 0 | 0 | 0 |
|  | 12 to 24 | 0 | 0 | 0 | 0 |
|  | 25 to 44 | 1 | 1 | 3 | 3 |
|  | 45 to 54 | 3 | 8 | 10 | 23 |
|  | 55 to 64 | 9 | 22 | 27 | 65 |
|  | 65 to 74 | 12 | 39 | 37 | 122 |
|  | 75 plus | 12 | 53 | 33 | 152 |
| Race | White | 30 | 9 | 89 | 26 |
|  | Black | 5 | 12 | 16 | 35 |
|  | Other | 3 | 12 | 7 | 30 |
| Ethnicity | Hispanic | 3 | 8 | 10 | 24 |
|  | Not Hispanic | 34 | 9 | 102 | 28 |
| Sex | Female | 19 | 9 | 54 | 24 |
|  | Male | 18 | 10 | 57 | 30 |
| Total |  | 37 | 9 | 110 | 27 |

Source: Healthcare Cost and Utilization Project National Inpatient Sample. Rates for age groups are unadjusted and all other rates are age-adjusted.

Pancreatic cancer contributed to 48,000 deaths in 2019 (Table 264). Pancreatic cancer mortality was rare among children and adolescents and mortality rates increased with age throughout adulthood. Age-adjusted mortality rates (underlying or other cause) were higher among men compared with women, Blacks compared with Whites, and non-Hispanics compared with Hispanics. Between 2004 and 2019, age-adjusted mortality rates (per 100,000) with pancreatic cancer as underlying or other cause increased by 3% from 11.3 to 11.6.(4)

**Table 264:** Pancreatic Cancer: Deaths with underlying or underlying/other cause and lifetime years of life lost by age, race, ethnicity, and sex in the United States, 2019.

| Demographic Characteristics |  | Underlying Cause |  |  | Underlying/Other Cause |  |  |
| --- | --- | --- | --- | --- | --- | --- | --- |
|  |  | Number of Deaths | Rate per 100,000 | Years of potential life lost in thousands | Number of Deaths | Rate per 100,000 | Years of potential life lost in thousands |
| Age (years) | 0 to 11 | 1 | 0.0 | 0.1 | 1 | 0.0 | 0.1 |
|  | 12 to 24 | 7 | 0.0 | 0.4 | 8 | 0.0 | 0.5 |
|  | 25 to 44 | 534 | 0.6 | 21.5 | 565 | 0.6 | 22.7 |
|  | 45 to 54 | 2,454 | 6.0 | 74.7 | 2,551 | 6.2 | 77.7 |
|  | 55 to 64 | 9,068 | 21.4 | 205.6 | 9,456 | 22.3 | 214.4 |
|  | 65 to 74 | 14,448 | 45.9 | 227.7 | 15,173 | 48.2 | 238.9 |
|  | 75 plus | 19,444 | 86.1 | 155.9 | 20,570 | 91.1 | 164.7 |
| Race | White | 38,345 | 11.0 | 566.0 | 40,315 | 11.6 | 593.2 |
|  | Black | 5,810 | 13.3 | 92.0 | 6,123 | 14.1 | 96.7 |
|  | Other | 1,801 | 7.3 | 27.8 | 1,886 | 7.6 | 29.0 |
| Ethnicity | Hispanic | 2,941 | 7.4 | 53.4 | 3,087 | 7.8 | 56.0 |
|  | Not Hispanic | 43,015 | 11.4 | 632.4 | 45,237 | 12.0 | 662.8 |
| Sex | Female | 22,176 | 9.6 | 333.4 | 23,224 | 10.1 | 348.3 |
|  | Male | 23,780 | 12.7 | 352.4 | 25,100 | 13.4 | 370.5 |
| Total |  | 45,956 | 11.0 | 685.8 | 48,324 | 11.6 | 718.8 |

Source: Vital Statistics of the United States.
Rates for age groups are unadjusted and all other rates are age-adjusted.
Years of potential life lost is based on U.S. life table 2006-2018 estimates of life expectancy at age of death.

## Data Availability

This work used existing data and no new data were collected. Data sources are listed in the appendices.

## ACKNOWLEDGMENTS

The authors thank Bryan Sayer, Helen Corns, Laura Fang, Ying Li, and Joe Evans for statistical programming.

Financial Support: The work was supported by contracts from the National Institute of Diabetes and Digestive and Kidney Diseases (HHSN275201700074U and 75N94022F00050).

