## Appendix 1 for "The Burden of Digestive Diseases in the United States Population"

Table 1: International Classification of Diseases (ICD) Code Digestive Disease Definitions

| Disease Code | Digestive Disease | ICD-9-CM Codes for Morbidity (1979-9/30/15) | ICD-10-CM Codes for Morbidity (since 10/1/15) | ICD-10 Codes for Mortality (since 1999) |
| --- | --- | --- | --- | --- |
| 1 | Gastrointestinal infections (excluding C. difficile) | 001-008.44, 008.46-009 | A00-A04.6, A04.8-A09 | A00-A04.6, A04.8-A09 |
| 2 | Clostridium difficile | 008.45 | A04.7 | A04.7 |
| 3 | Hepatitis A | 070.0, 070.1 | B15 | B15 |
| 4 | Hepatitis B | 1991 to present:  070.42, 070.52  All Years:  070.2, 070.3 | B16, B17.0, B18.0, B18.1, B19.1 | B16, B17.0, B18.0, B18.1 |
| 5 | Hepatitis C | 1991 to present:  070.41, 070.44, 070.51, 070.54, 070.7  Before 1991:  070.4, 070.5 | B17.1, B18.2, B19.2 | B17.1, B18.2 |
| 6 | Other viral hepatitis | 1991 to present:  070.43, 070.49, 070.53, 070.59  All Years  070.6, 070.9 | B17.2, B17.8, B17.9, B18.8, B18.9, B19.0, B19.9 | B17.2, B17.8, B17.9, B18.8, B18.9, B19 |
| 7 | Esophageal cancer | 150 | C15 | C15 |
| 8 | Gastric cancer | 151 | C16 | C16 |
| 9 | Cancer of small intestine | 152 | C17 | C17 |
| 10 | Colorectal cancer | 153, 154.0, 154.1 | C18-C20 | C18-C20 |
| 11 | Primary liver cancer | 155.0, 155.2 | C22.0, C22.2-C22.9 | C22.0, C22.2-C22.9 |
| 12 | Bile duct cancer | 155.1, 156.1-156.9 | C22.1, C24 | C22.1, C24 |
| 13 | Gallbladder cancer | 156.0 | C23 | C23 |
| 14 | Pancreatic cancer | 157 | C25 | C25 |
| 15 | Other digestive cancers | 154.2, 154.3, 154.8, 158, 159.0, 159.8, 159.9, 196.2, 197.4-197.8 | C21, C26.0, C26.8, C26.9, C45.1, C46.4, C48,  C7A.01, C7A.02,  C7A.092, C7A.094-C7A.096,  C7B.02, C7B.04,  C77.2, C78.4-C78.8 | C21, C26.0, C26.8, C26.9, C45.1, C48, C77.2, C78.4-C78.8 |
| 16 | Hemorrhoids | 455 | I84, K64 | I84, K64 |
| 17 | Gastroesophageal reflux disease | 530.1-530.3, 530.81 | K20, K21, K22.1, K22.2 | K20, K21, K22.1, K22.2 |
| 18 | Peptic ulcer disease | 531-534 | K25-K28 | K25-K28 |
| 19 | Functional disorders | 536, 564 | K30, K31.0, K31.83, K31.84, K58, K59, K91.0, K91.1, K91.8, K94.2 | K30, K31.0, K31.8, K58, K59, K91.0, K91.1, K91.8 |
| 20 | Appendicitis | 540-543 | K35-K38 | K35-K38 |
| 21 | Abdominal wall hernia | 550, 551.0-551.2, 551.8, 551.9, 552.0-552.2, 552.8, 552.9, 553.0-553.2, 553.8, 553.9 | K40-K43, K45, K46 | K40-K43, K45, K46 |
| 22 | Crohn’s disease | 555 | K50 | K50 |
| 23 | Ulcerative colitis | 556 | K51 | K51 |
| 24 | Diverticular disease | 562 | K57 | K57 |
| 25 | Liver disease | 570-573 | K70-K76 | K70-K76 |
| 26 | Gallstones | 574 | K80 | K80 |
| 27 | Acute pancreatitis | 577.0 | K85 | K85 |
| 28 | Chronic pancreatitis | 577.1 | K86.0, K86.1 | K86.0, K86.1 |
| 29 | Celiac disease | 579.0 | K90.0 | K90.0 |
| 30 | Other digestive diseases | 014, 017.8, 021.1, 022.2, 032.83, 040.2, 060, 072.3, 072.71, 075, 086.1, 091.1, 091.62, 095.2, 095.3, 098.7, 098.86, 099.52, 099.56,  112.84, 112.85, 120-129, 130.5, 176.3,  211, 230.1-230.9, 235.2-235.5, 239.0, 251.4-251.9, 271.3, 273.4, 275.0, 275.1, 277.01, 277.03, 277.1, 277.4, 279.01, 280.8, 281.0, 286.0-286.5, 286.7, 289.2,  306.4, 307.54, 307.7,  452, 453.0, 456.0-456.2,  530.0, 530.4-530.7, 530.82-530.89, 530.9, 535, 537, 538,  539, 551.3, 552.3, 553.3, 557, 558, 560, 565-569, 575, 576, 577.2, 577.8, 577.9, 578, 579.1-579.9,  643, 646.7, 671.8,  750.3-750.9, 751, 772.4, 773.4, 774.2-774.7, 776.0, 777, 779.3, 782.4, 787, 789.0, 789.1, 789.3-789.9, 792.1, 793.3, 793.4, 793.6, 794.8,  862.22, 862.32, 863, 864, 868.02-868.04, 868.12-868.14,  935.1, 935.2, 936-938, 947.2, 947.3, 973, 988.1, 996.82, 996.86, 996.87, 997.4,  V01.0, V02.0-V02.3, V02.6, V03.0, V03.1, V04.4, V05.3, V06.0, V10.00, V10.03-V10.09, V12.7, V16.0, V18.5, V42.7, V42.83, V42.84, V44.1-V44.4, V45.3, V45.72, V45.75, V45.86, V47.3, V53.5, V55.1-V55.4, V58.75, V59.6, V73.4, V74.0, V75.5-V75.7, V76.41, V76.5,  E858.4, E870.7, E879.5, E943 | A18.3, A18.83, A21.3, A22.2, A36.89, A42.1, A51.1, A51.45, A52.74, A54.6, A54.85, A56.3, A60.1, A74.81, A95,  B00.81, B05.4, B25.1, B25.2, B26.3, B26.81, B27, B37.81, B37.82, B46.2, B57.3, B58.1,  B65.1, B65.2, B65.8, B65.9, B66.0-B66.3, B66.5-B66.9, B67.0, B67.32, B67.39, B67.4-B67.9, B68, B69.89, B69.9, B70-B77, B78.0, B78.7, B78.9, B79-B82, B83.0, B83.1, B83.3, B83.8, B83.9, B87.82, B94.2,  D00.1, D00.2, D01, D12, D13, D17.5, D19.1, D20, D3A.01, D3A.02, D3A.092, D37.1-D37.9, D48.3, D48.4, D49.0, D50.1, D51.0, D66, D67, D68.0-D68.2, D68.31, D68.4, D80.2,  E16.3-E16.9, E73, E74.3, E80, E83.0, E83.1, E84.1, E88.01,  F55.0, F55.2, F98.1,  I81, I82.0, I85, I86.4, I88.0,  K22.0, K22.3-K22.9, K23, K29, K31.1-K31.7, K31.81, K31.82, K31.89, K31.9, K44, K52, K55, K56, K60-K63, K65-K68, K77, K81-K83, K86.2-K86.9, K87, K90.1-K90.9, K91.2-K91.7, K92, K94.0, K94.1, K94.3, K95,  O21, O22.4, O26.6, O87.2, O98.4, O99.6, O99.84,  P15.0, P35.3, P53, P54.0-P54.3, P57, P59, P76-P78, P92.0, P92.1,  Q39-Q45,  R10.0, R10.1, R10.3, R10.8,  R10.9, R11-R15, R16.0, P16.2, R17, R18, R19.0-R19.5, R19.7, R19.8, R74.0, R85, R93.2, R93.3, R93.5, R94.5,  S27.81, S30.3, S30.817, S30.827, S30.857, S30.867, S30.877, S30.98, S31.83, S36.1-S36.9,  T18.1-T18.9, T28.1, T28.2, T28.6, T28.7, T47, T62.0, T85.5, T86.4, T86.85, T86.89, Y84.5,  Z11.0, Z12.0, Z12.1, Z13.81, Z20.0, Z20.5, Z22.0, Z22.1, Z43.1-Z43.4, Z46.5, Z48.23, Z48.815, Z52.6, Z80.0, Z83.7,  Z85.0, Z86.010, Z87.1, Z87.738, Z90.3, Z90.4,  Z93.1-Z93.4, Z94.4, Z94.82, Z94.83, Z98.0, Z98.84 | A18.3, A21.3, A22.2, A51.1, A54.6, A56.3, A60.1, A74.8, A95,  B25.1, B25.2, B26.3, B27, B46.2, B57.3, B58.1, B65-B83, B94.2,  D00.1, D00.2, D01, D12, D13, D19.1, D20, D37.1-D37.9, D48.3, D48.4, D50.1, D51.0, D66, D67, D68.0-D68.4, D80.2,  E16.3-E16.9, E73, E74.3, E80, E83.0, E83.1, E84.1, E88.0,  F50.5, F98.1,  I81, I82.0, I85, I86.4, I88.0,  K22.0, K22.3-K22.9, K29, K31.1-K31.9, K44, K52, K55, K56, K60-K63, K65, K66, K81-K83, K86.2-K86.9, K90.1-K90.9, K91.2-K91.9, K92,  O21, O22.4, O26.6, O87.2,  P53, P54.0-P54.3, P57, P59,  P76-P78, P92.0, P92.1,  Q39-Q45,  R10.0, R10.1, R10.3, R10.4, R11-R15, R16.0, R16.2, R17-R19, R93.2, R93.3, R93.5, R94.5,  S36.1-S36.9,  T18.1-T18.9, T28.1, T28.2, T28.6, T28.7, T47, T62.0, T85.5, T86.4,  Y53, Y60.7, Y84.5 |
|  | All digestive diseases  (1-30) |  |  |  |

### Sources for ICD Codes

ICD-9-CM: https://ftp.cdc.gov/pub/Health_Statistics/NCHS/Publications/ICD9-CM/2011

ICD-10-CM: https://ftp.cdc.gov/pub/Health_Statistics/NCHS/Publications/ICD10CM/2020

ICD-10: https://ftp.cdc.gov/pub/Health_Statistics/NCHS/Publications/ICD10
