## Appendix 2 for "The Burden of Digestive Diseases in the United States Population"

NATIONAL AMBULATORY MEDICAL CARE SURVEY (NAMCS)

| SPONSOR: | Ambulatory Care Statistics Branch  Division of Health Care Statistics  National Center for Health Statistics  Centers for Disease Control and Prevention  U.S. Department of Health and Human Services  3311 Toledo Road  Hyattsville, MD 20782  301-458-4600  https://www.cdc.gov/nchs/ahcd/index.htm |
| --- | --- |
| DESIGN: | The National Ambulatory Medical Care Survey (NAMCS) is a continuing series of nationally representative sample surveys of office-based physicians in the United States. The survey includes all non-federal office-based physicians who are primarily engaged in direct patient care. Anesthesiologists, pathologists, and radiologists are excluded. The design is a multistage stratified probability sample of geographically defined areas, physician practices within these areas, and patient visits within physician practices. Physicians are asked to complete a patient encounter form for a systematic sample of office visits occurring during a randomly assigned one-week reporting period.  The study design is described in: National Center for Health Statistics, Bryant E, Shimuzu I. Sample design, sampling variance, and estimation procedures for the National Ambulatory Medical Care Survey. Hyattsville, Maryland: Public Health Service, 1988; DHHS Publication No. (PHS) 88-1382. (Vital and health statistics, Series 2, No. 108). |
| TIMEFRAME: | Data were collected annually from 1974 through 1981, and in 1985; since then, data have been collected annually beginning in 1989. |
| SAMPLE SIZE: | Through 1981, the sample included 3,000 total physicians, about 1,925 responding physicians, and about 51,000 patient visits. The 1985 sample included about 5,000 total physicians, 2,900 responding physicians, and 70,000 patient visits. Beginning in 1989, the sample included 2,500 total physicians, about 1,600 responding physicians, and about 42,000 patient visits (unweighted). Sample size for patient visits decreased in 2015 and 2016 to 28,332 in 2015 and 13,165 in 2016. |
| CONTENT RELEVANT TO  DIGESTIVE DISEASES: | Demographic data, reason for visit, physician's diagnostic and therapeutic services ordered or provided, diagnosis and disposition decision, and drugs prescribed are included. International Classification of Diseases (ICD) codes are given for the first four physician diagnoses. The reason for office visit is the principal reason given by the patient, which in the physician's judgment is the most appropriate one. Up to two or four additional symptoms or other reasons for visit can be coded. Beginning with the 2016 data, ICD-10 codes replace the ICD-9 codes. |
| STRENGTHS: | Survey form is completed from provider records. Trend data are available for about 30 years. Visits can be compared with those of the National Health Interview Survey, in which the conditions are similarly defined. Since 1980, data have been collected on the number and names of specific drugs prescribed in office-based practice. The sample allows estimates for specific physician subspecialties. ICD codes are used for diagnoses. |
| LIMITATIONS: | The sample is limited to office-based physicians, a group that has become a less inclusive source for ambulatory care. There may be more than one report per person, because the report reflects a visit rather than an individual. The sample size is small, so estimates of fewer than 200,000 are statistically unreliable. Because ambulatory care in federal facilities is not included, ambulatory care rates based on the U.S. population are underestimates. |
| AVAILABILITY OF DATA: | The National Center for Health Statistics Vital and health statistics, Series 13 <http://www.cdc.gov/nchs/products/pubs/pubd/series/ser.htm#sr13> and advance data <http://www.cdc.gov/nchs/products/pubs/pubd/ad/ad.htm> can be referred. Data are available for public use on the National Center for Health Statistics Web site in an easy-to-use form with input statements. However, to use the full seven-digit ICD-10 codes needed for the definition of digestive diseases requires using the Research Data Center (RDC) at the National Center for Health Statistics (NCHS). |

HEALTHCARE COST AND UTILIZATION PROJECT-NATIONWIDE EMERGENCY DEPARTMENT SAMPLE (HCUP NEDS)

| SPONSORS: | Agency for Healthcare Research and Quality  Department of Health and Human Services  5600 Fishers Lane  Rockville, MD 20857  301-427-1364  1-866-290-HCUP  <https://www.hcup-us.ahrq.gov/nedsoverview.jsp> |
| --- | --- |
| DESIGN: | The Healthcare Cost and Utilization Project Nationwide Emergency Department Sample (HCUP NEDS) is the largest all-payer emergency department database in the United States, yielding national estimates of hospital-based emergency department visits. Unweighted, it contains data from approximately 31 million emergency department visits each year. Weighted, it estimates roughly 143 million emergency department visits. Developed through a federal-state-industry partnership sponsored by the Agency for Healthcare Research and Quality, HCUP data inform decision making at the national, state, and community levels. |
| TIMEFRAME: | Data were collected annually since 2006. |
| SAMPLE SIZE: | Approximately 31 million emergency department visits (unweighted) each year. |
| CONTENT RELEVANT TO  DIGESTIVE DISEASES: | Data for each emergency department visit include patient demographics (gender, age, race), admission status, total charges, expected payment source, diagnosis and surgical proce­dures coded using ICD and CPT codes, and hospital characteristics (ownership, size, teaching status). |
| STRENGTHS: | The NEDS is the largest all-payer emergency department database in the U.S. Data are weighted to be nationally representative of non-federal hospitals in the U.S. NEDS is the only emergency department database containing charge information on all patients, regardless of payer. |
| LIMITATIONS: | Not all states are included. Only hospital owned emergency departments are included. The universe of hospital-owned emergency departments is therefore defined as the AHA community, non-rehabilitation hospitals that reported total emergency departments visits. The AHA defines community hospitals as "all non-federal, short-term, general, and other specialty hospitals." Included among community hospitals are pediatric institutions, public hospitals, and academic medical centers. |
| AVAILABILITY OF DATA: | Summary statistics are published by the U.S. Agency for Healthcare Research and Quality <http://www.hcup-us.ahrq.gov/reports.jsp> with an online database, HCUP-Net, <http://hcupnet.ahrq.gov/> that allows users to easily generate certain statistics. Selected data sets can be purchased for analysis. |

HEALTHCARE COST AND UTILIZATION PROJECT-NATIONAL (NATIONWIDE) INPATIENT SAMPLE (HCUP NIS)

| SPONSORS: | Agency for Healthcare Research and Quality  U.S. Department of Health and Human Services  5600 Fishers Lane  Rockville, MD 20857  301-427-1364  1-866-290-HCUP  <https://hcup-us.ahrq.gov/db/nation/nis/nisdbdocumentation.jsp> |
| --- | --- |
| DESIGN: | The Healthcare Cost and Utilization Project National (Nationwide) Inpatient Sample (HCUP NIS) is a database of hospital inpatient stays. It utilizes a stratified sample of hospitals drawn from the subset of hospitals in the states that make their data available to HCUP. Hospitals are stratified by region, location/teaching status, bed-size category, and ownership. All discharges from sampled hospitals are included. The 2004 NIS includes all discharges from more than 1,000 hospitals, an approximate 20 percent stratified sample of U.S. community hospitals. NIS data are weighted to represent the annual discharges from non-federal hospitals in the United States.  Several revisions have been made to the NIS sampling design since its inception. First, the sampling frame changed over time as more states made their data available to HCUP. The 1988 NIS was drawn from a sampling frame of eight states, representing 31 percent of all hospital discharges in the United States. In contrast, the sampling frame in recent years included 46 states plus the District of Columbia, representing 85 to 90 percent of all hospital discharges in the United States. Second, in 1998, the sampling method was changed to better reflect the cross-sectional population of hospitals. The hospital stratification variables were redefined, short-term rehabilitation facilities were dropped from the target universe, and sampling preference was no longer given to prior-year NIS hospitals.  Beginning with the 2012 data year, the NIS approximates a 20-percent stratified sample of all discharges from U.S. community hospitals, excluding rehabilitation and long-term acute care hospitals. The NIS contains information on all patients, regardless of payer, including individuals covered by Medicare, Medicaid, or private insurance, and uninsured. The NIS is sampled from the [State Inpatient Databases (SID](https://www.hcup-us.ahrq.gov/sidoverview.jsp)), which include all inpatient data that are currently contributed to HCUP.  Beginning with 2012 data, the NIS was redesigned to improve national estimates. To highlight the design change, beginning with 2012 data, AHRQ renamed the NIS from the "Nationwide Inpatient Sample" to the "National Inpatient Sample." The redesign incorporates three major types of changes:   - Revisions to the sample design - the NIS is now a sample of discharge records from all HCUP-participating hospitals, rather than a sample of hospitals from which all discharges were retained. - Revisions to how hospitals are defined - the NIS now uses the definitions of hospitals and discharges supplied by the statewide data organizations that contribute to HCUP, rather than the definitions used by the AHA Annual Survey. - Revisions to enhance confidentiality - the NIS now eliminates state and hospital identifiers and other data elements that are not uniformly available across states.   Key features of the NIS (2016) include:   - The NIS is drawn from all states participating in HCUP, representing more than 97 percent of the U.S. population. - The NIS approximates a 20-percent stratified sample of discharges from U.S. community hospitals, excluding rehabilitation and long-term acute care hospitals. - The self-weighting design of the new NIS reduces the margin of error for estimates and delivers more stable and precise estimates than previous versions of the NIS. - The NIS protects patient confidentiality because state and hospital identifiers are no longer provided. - The new NIS retains a large sample size, which enables analyses of rare conditions, uncommon treatments, and special patient populations. |
| TIMEFRAME: | Data have been collected annually since 1988. |
| SAMPLE SIZE: | The sample size is approximately 20% of hospital stays. |
| CONTENT RELEVANT TO  DIGESTIVE DISEASES: | Data for each hospital stay include patient demographics (gender, age, race, median income for ZIP Code), admission and discharge status, length of stay, total charges, expected payment source, up to 15 diagnoses and 7 surgical proce­dures coded using International Classification of Diseases (ICD) codes, and hospital characteristics (ownership, size, teaching status). |
| STRENGTHS: | The NIS is the largest all-payer inpatient care database in the United States. Data are weighted to be nationally representative of non-federal hospitals in the United States. The NIS is the only national hospital database containing charge information on all patients, regardless of payer. |
| LIMITATIONS: | Not all states participate. Not all participating states collect data on race-ethnicity. The charge information is for the facility only; no information on physician fees is available. Data on medications are not supplied, although medication costs are included in the charge total. |
| AVAILABILITY OF DATA: | Summary statistics are published by the U.S. Agency for Healthcare Research and Quality <http://www.hcup-us.ahrq.gov/reports.jsp> with an online database, HCUP-Net, <http://hcupnet.ahrq.gov/> that allows users to easily generate certain statistics. Selected data sets can be purchased for analysis. |

VITAL STATISTICS OF THE UNITED STATES: MULTIPLE CAUSE-OF-DEATH

| SPONSORS: | Mortality Statistics Branch  Division of Vital Statistics  National Center for Health Statistics  Centers for Disease Control and Prevention  U.S. Department of Health and Human Services  3311 Toledo Road, 7^th^ floor  Hyattsville, MD 20782  301-458-4666  <http://www.cdc.gov/nchs/deaths.htm> |
| --- | --- |
| DESIGN: | Multiple cause-of-death mortality data from the National Vital Statistics System provide mortality data by multiple cause of death for all deaths occurring within the United States. Each record in the microdata is based on information abstracted from death certificates filed in vital statistics offices of each state and the District of Columbia. Causes of death were coded according to the International Classification of Diseases (ICD)-9 for 1979 through 1998, and according to ICD-10, beginning in 1999.  The study design is described in: National Center for Health Statistics, Data systems of the National Center for Health Statistics. Hyattsville, Maryland: Public Health Service, 1981; DHHS Publication No. (PHS) 82-1318. (Vital and health statistics: Series 1, No. 16). |
| TIMEFRAME: | Data have been collected annually since 1968. |
| SAMPLE SIZE: | The sample is a 100 percent count of deaths occurring in the United States. |
| CONTENT RELEVANT TO  DIGESTIVE DISEASES: | Demographic data (age, sex, race, residence) and underlying and contributing causes of death are included. |
| STRENGTHS: | A complete count of deaths in the United States is included, along with a number of diagnosis fields. Trend data are available for more than 35 years. For digestive diseases with high mortality rates, such as cirrhosis, death records are the most comprehensive data source. Mortality statistics may be the only reliable data source for uncommon fatal conditions. Annual age-adjusted mortality rates are useful for examining trends over time, assuming case-fatality rates do not change significantly. Mortality rates for diseases that are usually fatal are often used as estimates of incidence rates when the latter are not available. |
| LIMITATIONS: | Quality is dependent on the accuracy of death certificates, which may vary, according to condition. Chronic diseases that contribute to mortality are frequently underreported. |
| AVAILABILITY OF DATA: | National Center for Health Statistics. Vital statistics of the United States, Vol. II <http://www.cdc.gov/nchs/products/pubs/pubd/vsus/vsus.htm> Mortality Parts A and B; <http://www.cdc.gov/nchs/products/nvsr.htm> National vital statistics reports; and <http://www.cdc.gov/nchs/products/pubs/pubd/series/ser.htm#sr20> Vital and health statistics, Series 20. Data available through National Bureau of Economic Research Web site <http://www.nber.org/data/multicause.html> in an easy-to-use form with input statements in addition to the CDC website <https://www.cdc.gov/nchs/data_access/vitalstatsonline.htm>. |

UNITED STATES POPULATION ESTIMATES

| SPONSOR: | Division of Population Projections  U.S. Census Bureau  From CDC Wonder  Centers for Disease Control and Prevention (CDC)  U.S. Department of Health and Human Services  1600 Clifton Road  Atlanta, GA 30333  404-639-3311  404-639-3534 and 800-311-3435 (public inquiries)  <https://www.cdc.gov/nchs/nvss/bridged_race.htm> |
| --- | --- |
| DESIGN: | The population estimates are mid-year (July 1) population counts by age, sex, and race. The counts are used with all national samples as the denominator for all estimates of rates. The year 2000 estimates are also used for age adjusting. These estimates are not used for cancer statistics from the Surveillance, Epidemiology, and End Results (SEER) program, which has its own population counts. |
| TIMEFRAME: | Estimates for all years were used in this report. |
| SAMPLE SIZE: | The U.S. population is the sample. |
| CONTENT RELEVANT TO  DlGESTIVE DISEASES: | Denominators are provided for calculating rate per 100,000 persons by age, race, ethnicity, and sex. |

SURVEILLANCE, EPIDEMIOLOGY, AND END RESULTS (SEER) PROGRAM

| SPONSORS: | Cancer Statistics Branch  Surveillance Research Program  Division of Cancer Control and Population Sciences  National Cancer Institute  National Institutes of Health  U.S. Department of Health and Human Services  6116 Executive Boulevard  Suite 504, MSC 8316  Bethesda, MD 20892-8316  301-496-8510  http://seer.cancer.gov/ |
| --- | --- |
| DESIGN: | A total of 17 population-based registries in the United States pro­vide data on all residents diagnosed with cancer and follow-up information on all previously diagnosed patients. Data are compiled twice a year. Cancer mortality data are obtained from vital statistics for the entire United States. |
| TIMEFRAME: | Data have been collected annually since 1975. |
| SAMPLE SIZE: | Surveillance, Epidemiology, and End Results (SEER) program data for trends are 100 percent counts from Atlanta, Georgia; Con­necticut; Detroit, Michigan; Hawaii; Iowa; New Mexico; San Francisco/Oakland, California; Seattle/Puget Sound, Washington; and Utah. SEER data for 2004 are 100 percent counts from the 9 registries above, plus Los Angeles, California; San Jose-Monterey, California; Rural Georgia; the Alaska Native Tumor Registry; Greater California; Kentucky; Louisiana; and New Jersey. |
| CONTENT RELEVANT TO  DIGESTIVE DISEASES: | Data regarding cancer incidence and mortality, including current and pro­jected trends, are collected for selected sites such as esophagus, stomach, colon, rectum, liver, and pancreas. Demographic data include age, sex, and race. |
| STRENGTHS: | SEER data are verified for quality and completeness. Data are estimated to be 99 percent complete from the registry sites. |
| LIMITATIONS: | SEER data represent only 17 areas of the country. Although the data are weighted to provide national estimates, these data are not statistically repre­sentative of the United States. |
| AVAILABILITY OF DATA: | The National Cancer Institute <http://seer.cancer.gov/publications/> and certain statistics can easily be generated online <http://seer.cancer.gov/statistics/> with selected data sets are available for analysis. |

OPTUM CLINFORMATICS®  DATA MART (CDM)

| SPONSOR: | UnitedHealth Group  P.O. Box 1459  Minneapolis, MN 55440-1459  (800) 328-5979  <https://www.optum.com/> |
| --- | --- |
| DESIGN: | The Clinformatics® Data Mart (CDM) is a database comprised of administrative health claims for members of a large national managed care company affiliated with Optum. These administrative claims are submitted for payment by providers and pharmacies are verified, adjudicated, adjusted, and de-identified prior to inclusion in CDM.  Data are included for only those covered lives with both medical and prescription drug coverage, to enable users to evaluate the claims related to the complete health care experience. Additionally, the CDM includes results for outpatient lab tests processed by large national lab vendors under contract with the managed care organization. The CDM is comprised of commercial health plan data and Medicare Advantage members. The population is geographically diverse, spanning all 50 states.  In addition to medical claims, pharmacy claims, and lab results, CDM includes data tables related to member inpatient confinements and member eligibility data. The internal database is updated monthly. Customers’ datasets can be refreshed annually, semi-annually, or quarterly, as designated by the contractual agreement.  CDM includes the following members from the affiliate company:   - Fully Insured Lives - ASO (Administrative Services Only) Lives - Legacy Medicare Choice Lives (prior to January 2006) - Medicare Advantage (MAPD starting in January 2006)   In all categories above, only members with both medical and pharmacy benefits are included.  ASO lives represent companies who contract with the affiliate company to administer health benefits for their employees. The affiliate company is not financially accountable for their healthcare service utilization. Inclusion of ASO members in the database both increases sample size and provides a wider breadth in the data available.  Medicare Advantage with Part D (MAPD) members beginning in 2006 is included. |
| TIMEFRAME: | Data have been collected monthly since 2001. |
| SAMPLE SIZE: | The CDM includes data with service dates beginning 1/2001 through the present date. The database includes approximately 15 – 18 million annual covered lives for a total of roughly 98 million unique lives over a seventeen-year period (1/2001 – 12/2017). |
| CONTENT RELEVANT TO  DIGESTIVE DISEASES: | Demographics including age, gender, race, and region; date of death; and data for each medical or confinement claim include patient admission status, total costs and charges, diagnoses and surgical proce­dures coded using ICD and CPT codes |
| STRENGTHS: | Data are on a member-level, allowing members to be tracked across different plans through different enrollment periods. Month and year of death are based on social security number plus one additional piece of PII. |
| LIMITATIONS: | CDM contains lab results from certain laboratory chains with contractual relationships with the major insurance company sourcing CDM. There are no incentives on the part of the members or the providers to use these lab vendors. Therefore, the lab result data should be considered incomplete. Not all members have lab results. Even if a member has lab results in the CDM, this does not guarantee that all labs are represented. Diagnoses are not recorded with pharmacy claims. In 2011, some states stopped contributing dates of death to the Social Security office for privacy concerns, and so state reported information is no longer used. This caused a 30% drop in records. It is important to avoid comparing data from before this time to after this time because of this. |
| AVAILABILITY OF DATA: | Selected data sets can be purchased for analysis.  Either date-of-death or socioeconomic status versions are available. |

CENTERS FOR MEDICARE AND MEDICAID SERVICES (CMS) MEDICARE 5% SAMPLE

| SPONSOR: | Centers for Medicare and Medicaid Services  U.S Department of Health & Human Services  Hubert H. Humphrey Building  200 Independence Avenue, S.W.  Washington, D.C. 20201 |
| --- | --- |
| DESIGN: | The Centers for Medicare and Medicaid Services (CMS) Chronic Conditions Data Warehouse (CCW) is a research database engine that contains an amalgamation of linked datasets containing Medicare, Medicaid, Assessments and Part D Prescription Drug Event administrative data.  CMS creates the 5% Medicare sample files to be representative of the entire datasets of Medicare beneficiaries. Data are linked by a unique, unidentifiable beneficiary key that can be used to follow beneficiaries across care settings and time.  The Medicare datasets contain fee-for-service institutional and non-institutional claims and enrollment/eligibility information (Part A, Part B, Part D and Medicare Advantage). Assessment data and Part D Prescription Drug Event data which includes plan characteristics, pharmacy characteristics and prescriber characteristics are also available (2006 onward for Part D).  These administrative data are obtained for billing purposes and contain information on claims, diagnoses, measures of costs, healthcare utilization e.g., number of visits, type of admissions, lengths of stays etc., mortality and dates of death, and demographic information including sex, race, insurance type, age, geographic location etc. at an individual Medicare beneficiary level. |
| TIMEFRAME: | Medicare files are available in the CCW from 1999 onward and Part D Prescription Drug Event data files are available from 2006 onward. NDI Cause of Death Information is available from 1999 onward. |
| SAMPLE SIZE: | A 5% sample created by CMS representative of the dataset of Medicare beneficiaries. |
| CONTENT RELEVANT TO  DlGESTIVE DISEASES: | Primary and other diagnoses (up to 25 in some datasets) are coded using ICD-9 prior to 2015 quarter 4 and ICD-10 for 2015 quarter 4 onward. Procedures are coded using CPT codes. Utilization includes number of claims, number of visits, types of procedures, patient admission type/date/length, and associated charges and costs. Demographic information includes sex, race, insurance type, age, and geographic location. Death information is available from 2007 onward, including date of death and cause of death diagnosis. |
| STRENGTHS: | These datasets allow beneficiaries to be followed across multiple care settings and years. |
| LIMITATIONS: | Linking all entries for one specific event can be complicated, as separate claims are generated for institutional (Part A) and physician/supplier (Part B) services. Information is not available on stage/severity or histology of diseases. The Part D event file contains no diagnosis code. Different care settings use different procedure codes (ICD codes for inpatient care and CPT/HCPCS codes for outpatient care.) Exact timing of events can be difficult to calculate. |
| AVAILABILITY OF DATA: | Data can be accessed through the CCW Virtual Research Data Center (VRDC). Costs vary depending on beneficiary cohort size, years of data and files requested.  <https://www.resdac.org/>  <https://www2.ccwdata.org/web/guest/home/> |
