## Appendix 3 for "The Burden of Digestive Diseases in the United States Population"

This appendix provides information on the sources and computations for the tables of digestive diseases.

### National Event-Level Data Sources

#### Disease Definitions

Digestive diseases were coded based on either the International Classification of Diseases ICD-9 CM (Clinical Modification) https://www.cdc.gov/nchs/icd/icd9cm.htm for events prior to the 4^th^ quarter of 2015 or ICD-10 for events beginning in 4^th^ quarter of 2015.

Mortality is coded with either ICD-9 <https://www.cdc.gov/nchs/icd/icd9.htm> for deaths prior to 1999, or ICD-10 <https://www.cdc.gov/nchs/icd/icd10.htm> for deaths in 1999–present. See Appendix 1 for the complete list of codes. The first-listed diagnosis was considered the primary diagnosis. All remaining diagnoses were considered secondary and were included (along with the primary diagnosis) under the category “All-Listed Diagnoses.” In the tables for ambulatory care visits, emergency department visits, hospital discharges, and mortality, diagnoses were counted only once under the all-listed category, irrespective of the number of actual diagnoses. For example, in the chapter on all digestive diseases, only one digestive disease diagnosis was counted even though more than one could have been listed on a medical record or death certificate.

While the coding for digestive disease mortality is generally consistent between ICD-9 and ICD-10, the World Health Organization (WHO), which produces the ICD code definitions, advises that series are not necessarily comparable across versions of the ICD code book. The “Other Digestive Disease” category (not a separate disease but included in All Digestive Diseases) shows the greatest difference between ICD-9 and ICD-10.

#### Demographic Categories for National Event Level Data

For the purpose of calculating rates for the U.S. population, annual population data were derived from the national population estimates program of the U.S. Census Bureau and the Centers for Disease Control and Prevention (CDC) <http://wonder.cdc.gov/population.html> for national estimates from survey data. Population estimates from claims data (Medicare and Optum) are specific to the claims and are based on the enrollment in the respective programs.

For the survey data, race was coded as “White” or “Black;” or “Other,” if another category was specified. Missing race data were not considered “Other.” The HCUP NIS data combine Hispanic origin with race, so it was impossible to know whether Hispanics were white or black. In order not to undercount the totals, we assumed all Hispanics were white. As a result, discharges for whites were slightly overstated and for blacks slightly understated.

HCUP NIS data came from the individual states, and a varying number of states did not report race, depending on the year. To adjust for this limitation, we created a separate weight for race, based on the existing weight times the inverse of the proportion of each race in the states that did report race to the total for all states in that year of the NIS. Note that these are counts of persons, based on the mid-year population estimates, and not the proportion of discharges. We did not report separate counts for “Other Race,” because the definition of “Other Race” in the HCUP NIS and the population counts may not be the same. A similar weight was created for Hispanic origin.

#### Age-Adjustment

Age-adjustment through direct standardization allowed for comparisons across race, sex, and time that were not influenced by differences in age distribution for the groups being compared. Year-specific population data in 19 age groups, plus the National Center for Health Statistics (NCHS) standard year 2000 population, were used for age-adjusting http://www.cdc.gov/nchs/data/nvsr/nvsr47/nvs47_03.pdf. Age-specific rates were calculated for each of the 18 age groups (age 0–4, 5-year age groups through age 84, and age 85 and older), and the results were multiplied by the year 2000 standard population proportion in each of the age groups. These results were then summed to arrive at the age-adjusted population rate estimate. Further details can be found in Anderson and Rosenberg.^1^

**Morbidity Estimates**

Ambulatory Care Visits

Estimates in the tables for ambulatory care visits are based on 3-year averages. Multiple years were combined to have sufficient observations to meet the minimum threshold for reporting and for more stable estimates. The 3 years of data were averaged by dividing the sampling weight by 3, in accordance with the general instructions from NCHS.

First-Listed Diagnosis

The primary diagnosis for an outpatient visit was the first diagnosis listed in the record. A visit was considered to have been for a digestive disease if the first diagnosis listed on the record fell into the subject category. Estimates for first-listed diagnosis for digestive diseases included the number of visits and the rate of visits per 100,000 of the population. The rate per 100,000 was the number of visits, not the number of individuals with a visit, divided by the number of persons (in 100,000s) in the population in the specific subgroup.

The weighted count of visits with a first-listed diagnosis of each of the digestive diseases was the count (in thousands) listed in the table under “Ambulatory Care Visits,” “First-Listed Diagnosis,” “Number in Thousands.” The “Rate per 100,000” was calculated by dividing the count of visits by the number of people (in 100,000s) in the population in the specific subgroup.

All-Listed Diagnoses

Each outpatient record could have multiple diagnoses listed. A visit was considered to have been for a specific digestive disease if any of the diagnoses listed on the record fell into the subject category. Therefore, any individual record could be counted for more than one digestive disease. However, a given record was not counted more than once for a specific disease. For example, a record having the ICD-9-CM diagnosis codes of “001” and “002” was only counted once in the category of Gastrointestinal Infections. The weighted count of visits with all-listed diagnoses of each of the digestive diseases was the count (in thousands) listed in the table under “Ambulatory Care Visits,” “All-Listed Diagnoses,” “Number in Thousands.” The “Rate per 100,000” was calculated by dividing the count of visits by the number of persons in the population (in 100,000s) in the demographic subgroup.

Hospital Discharges

Hospital discharges were based on inpatient stays of at least 1 night. Emergency room visits that did not result in an admission to the hospital with an overnight stay were not counted. Data in the tables came from the HCUP NIS file of hospital discharges from participating states. Sampling weights inflated the discharges to the U.S. total, based on information from the American Hospital Association.

First-Listed Diagnosis

The primary diagnosis for a hospital discharge was the first diagnosis listed in the record. Inpatient estimates for first-listed diagnosis for digestive diseases included the number of discharges and the rate of discharges per 100,000 of the population. The weighted count of hospital discharges with a primary diagnosis of each of the digestive diseases was the count (in thousands) listed in the table under “Hospital Discharges,” “First-Listed Diagnosis,” “Number in Thousands.” The “Rate per 100,000” was the number of discharges, not the number of individuals with an inpatient stay, divided by the number of persons (in 100,000s) in the population in the specific subgroup.

All-Listed Diagnoses

Each hospital discharge record could have multiple diagnoses listed. A hospitalization was considered to have been for a specific digestive disease, if any of the diagnoses listed on the record fell into the subject category. Therefore, any individual record could be counted for more than one digestive disease. As with ambulatory care visits, a given record was not counted more than once for a specific disease. For example, ICD-9-CM diagnostic codes of “001” and “002” were only counted once in the category of Gastrointestinal Infections. The weighted count of hospital discharges with all-listed diagnoses of each of the digestive diseases was the count (in thousands) listed in the table under “Hospital Discharges,” “All-Listed Diagnosis,” “Number in Thousands.” The “Rate per 100,000” was calculated by dividing the count of hospital discharges by the number of persons in the population (in 100,000s) in the demographic subgroup.

**Mortality Estimates**

Counts of deaths from digestive diseases were derived from the Multiple Cause-of-Death data files from the Division of Vital Statistics, CDC. These data are a complete accounting of all deaths in the United States (although not necessarily for all U.S. citizens). Cause of death is organized on a record axis, with a specific underlying cause of death and contributing causes for each decedent.

Underlying Cause of Death

The underlying cause of death was determined from the list of all causes on the death certificate by professional coders. Underlying cause is analogous to a first-listed diagnosis for morbidity. The “Number of Deaths” column for “Underlying Cause” was a count of the number of records in the file with a digestive disease as the underlying cause of death.

The “Rate per 100,000” column was determined by dividing the number of deaths with the underlying cause by the population (in 100,000s) in the demographic subgroup. The race- and sex-specific estimates were age-adjusted, while the age-specific rates were not age-adjusted.

“Years of Potential Life Lost” assumed life expectancy of 75 years had individuals not died before that age. Because age at death is reported in full years, we added 0.5 years to each age at death. Thus, for the purpose of calculating years of life lost, a person whose age at death was listed as 65 was counted as having been 65.5 years old. The age 65.5 represented the average age of all people who died at age 65, and each contributed 9.5 years of potential life lost (75-65.5 = 9.5). The tables show the total number of years of life lost to age 75 in thousands.

Underlying or Other Cause of Death

The record axis of the death certificate can list a number of contributing causes in addition to the underlying cause. A record of any of the unique digestive diseases was noted for each of the possible causes, and any duplicate digestive diseases were eliminated. A death was attributed to one of the digestive diseases if any of the unduplicated digestive diseases were recorded. Therefore, a death could appear under more than one of the digestive diseases in the “Underlying or Other Cause” column of the tables.

“Number of Deaths” (in 100,000s) was the count of all deaths that had the specified digestive disease listed in any position on the record axis. A death could appear under more than one disease if any of the diagnoses were listed; however, no death appeared more than once for a given disease.

The “Rate per 100,000” column was determined by dividing the number of deaths for underlying or other cause by the population (in 100,000s) in the demographic subgroup. The race- and sex-specific estimates were age-adjusted, while the age-specific rates and the total were not age-adjusted.

Cancer Incidence

Cancer incidence was derived from SEER registry data. The registries do not cover the entire United States and do not necessarily represent the entire population. Instead, each registry covers a specific set of counties, across diverse sections of the country. (For more information on registries, see SEER in Appendix 2.) Population counts used for rates and age-adjustment were also restricted to the counties covered by the registry. Only estimates based on unweighted counts of 17 or more cases were shown, following the reporting standard set by NCI.

Cancer incidence was estimated for the entire country from the rates for the registries multiplied by the annual U.S. population. This yielded an estimated annual number of new cases for the United States. The unadjusted and age-adjusted incidence rates were based only on the registry areas. Unadjusted rates were calculated from the number of new cases divided by the population in the demographic subgroup. Age-adjusted incidence rates were calculated from the age-specific rates within the demographic subgroup multiplied by the U.S. standard 2000 population as described above under “Age-Adjustment”.

### Person-Level Data Sources

#### Optum Clinformatics® Data Mart

Data source

The Optum© Clinformatics® Data Mart (CDM) database is comprised of de-identified adjudicated administrative health claims for 15-18 million individuals covered annually by commercial insurance in all 50 states. All members were covered for medical services and prescription drugs. Member eligibility files containing birth year, gender, race/ethnicity, geographic region, and eligibility period were linked with inpatient confinement, medical, medical diagnosis, and pharmacy claims. Inpatient confinement files summarized services in acute care hospitals or skilled nursing facilities. Medical files included reimbursement claims for healthcare professional services provided in all service locations (e.g., inpatient hospital, outpatient facilities, physician office, and laboratory). Medical diagnosis files included diagnosis information related to each medical claim. Diagnoses were identified using ICD-10-CM codes, from participants’ medical claims.

Study sample

The study population covered by these analyses included all privately insured enrollees with a single consistent birth year recorded in CDM who resided in the United States and were continuously enrolled for at least one full calendar year.

#### Measures

Enrollees were considered digestive disease patients if they met the study population inclusion criteria and had one or more claims with an ICD-10-CM diagnostic code indicative of a digestive disease in any diagnosis field in the medical files. Inpatient hospitalizations were defined as having an inpatient hospital place of service. Ambulatory visits were evaluation & management visits with an office, outpatient hospital, or ambulatory surgical center place of service. Evaluation  & management visits were additionally characterized by a non-facility location with a type of service including office visits, surgery, ER, consultations, or home health/hospice visits, and excluding non-encounter services; capitated primary, specialist, or professional encounters, fee-for-services charged to Cap Organization; and fee-for-service for OXFORD/Parallax/NICE only. Other covariates included age (0-11, 12-24, 25-44, 45-54, 55-64, 65-74, or 75+), gender (male or female), race/ethnicity (non-Hispanic White, non-Hispanic Black, Hispanic, Asian, or unknown), and geographical region (Northeast, Midwest, South, or West).

#### Digestive disease prevalence

The annual number and percentage of privately insured enrollees in CDM who qualified as digestive disease patients were calculated. Analyses were reported overall and stratified by age, gender, race/ethnicity, and region.

### Centers for Medicare and Medicaid Services (CMS) Medicare 5% sample

Data source

The Centers for Medicare and Medicaid Services (CMS) Chronic Conditions Warehouse (CCW) is a research database engine that contains an amalgamation of linked datasets containing Medicare, Assessments and Part D Prescription Drug Event administrative data. CMS creates the Medicare 5% datasets to be a representative sample of the entire set of Medicare beneficiaries. It is sufficiently representative as to allow population-level estimation. Data for a beneficiary can be linked across care settings and years.

Study sample (Denominator)

The study population/denominator consisted of beneficiaries who were 65 years or older as of January 1^st^ of the year and resided in the 50 U.S states or Washington D.C, and were continuously and fully enrolled in Part A and Part B Medicare benefits throughout the year or until death and were not enrolled in the Medicare Advantage program (Part C/Health Maintenance Organization benefits). Enrollment in Part C has doubled in terms of percentage of the total Medicare population over time. The denominator file was at an individual level and contained demographic and enrollment data and was linkable to other Medicare data by the beneficiary unique identifier. Figure 1 shows the count and percent of individuals aged 65 plus who are eligible for inclusion. As can be seen in the figure, the absolute number of people defined for the denominator stays relatively constant at about 25 to 26 million per year for 2006-2016. However, the overall population of people aged 65 plus increases from about 35 million to 47 million. Correspondingly, the percent of the 65 plus population in the denominator falls from 75% to less than 60% over the period.

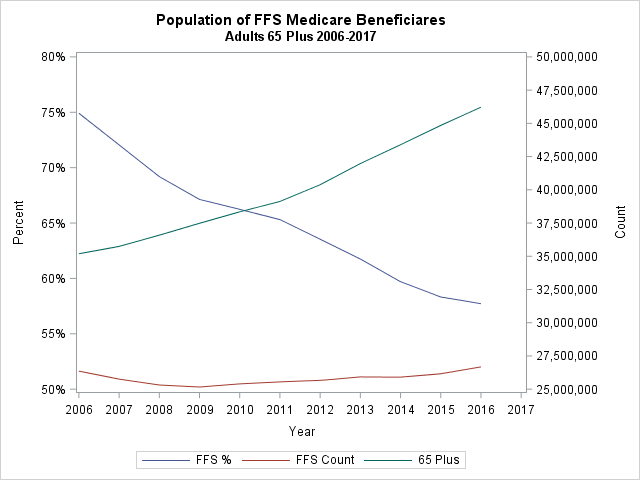

*Figure 1: Eligible Medicare Beneficiaries*

Measures

Enrollees were considered digestive disease patients (the claims-based prevalence measure) if they met the study population inclusion criteria and had one or more claims with an ICD-10-CM diagnostic code indicative of a digestive disease in the primary or any diagnosis field in the medical claim files. The medical claims files used for analyses were institutional and non-institutional claims files. Institutional claims files for fee-for-service contain information on claims submitted by health care institutions for reimbursement of facility costs. There are 5 types of institutional files for the different types of institutional claims: hospital inpatient stays, hospital outpatient services, skilled nursing facilities, home health agencies, and hospice care organizations. Non-institutional claims (also called Part B claims) contain claims from health-care professional (e.g., doctors) for reimbursement and for supplies and services such as laboratory tests, etc. Both institutional and non-institutional claims files have a base file that contains information at a claim level including the corresponding diagnoses. A revenue detail file exists for each type of institutional claim file and a line-level file for the non-institutional claim file. These line/revenue level files contain line-item details of charges and contain information such as medical procedure Healthcare Common Procedure Coding System codes (HCPCS) or place of service and type of service. Both types of line level files can be linked back to the appropriate base file using beneficiary and claim ids, but institutional claims for a given event (such as inpatient stay) are independent of supplier claims and may have different diagnosis codes as a result.

For each of the measures, the number of qualified persons and events having each digestive disease was reported. CMS restricts reporting to measures having at least 5 unweighted cases. All measures from 5% samples are multiplied by 20 to represent national estimates. Thus, any measure in a table under 100 is suppressed.

Inpatient hospitalizations

All claims in the hospital inpatients stays institutional dataset were used to identify a person as having an inpatient hospitalization.

Emergency department visits

People with emergency department visits were identified from the institutional hospital inpatients stays, institutional hospital outpatient services and the non-institutional Part B claims file. From the institutional files a claim was from an emergency department visit if the “revenue center” code was 0450-0459 or 0981 as these codes relate to the emergency room use or professional fees charged for emergency room. In the non-institutional file, a claim was from an emergency department visit if the service place indicated “emergency room” and service type indicated “medical care,” “surgery” or “consultation.” People with at least one claim were included in the analysis.

Ambulatory Care Visits

People with ambulatory care visits were identified when an evaluation & management (E & M) visit occurred at an office, outpatient hospital, or other place of service. These were identified from institutional hospital outpatient services and the non-institutional Part B claims file. A claim was classified as from an ambulatory E-M visit from the institutional hospital outpatient services file using either of two criteria: if the HCPCS code^[1]^ indicated an office or other outpatient visit for evaluation & management; or if the revenue code indicated a clinic visit^[2]^. From the non-institutional Part B claims file a claim was considered to result from an ambulatory E-M visit if the Service Place did not indicate “pharmacy,” “ambulance,” “mass immunization center,” “independent laboratory,” or “inpatient” and Service Type indicated “medical care,” “surgery” or “consultation.”

Both emergency department visit and ambulatory care visit events were matched on service date across data files (e.g., outpatient/Part B). This was because one event could have different claims in different files for the same visit (a facility charge and a service provider charge).

People with annual utilization for digestive diseases (Prevalence)

People qualified as digestive disease patients from any of the medical claims. All people identified from any of the following claim types: inpatient stays; hospital outpatient services; Part B services; skilled nursing facilities; home health agencies; hospice care organizations; and durable medical equipment were used to identify a person having a digestive disease event. People were included as a prevalent case who had at least one event in any of the datasets with a digestive disease in any diagnosis. Although it varies by digestive disease, anywhere from 7% to 69% of prevalent cases consist of a single ambulatory claim. This is lowest for cancers, but rather high for hemorrhoids, functional disorders, and diverticular disease, suggesting that many cases may not be chronic, or may be rule outs.

Mortality

Mortality was identified using the National Death Index (NDI) segment file. Enrollees were considered digestive disease patients if they met the study population inclusion criteria and had an ICD-10 cause of death code indicative of a digestive disease or in any of the entity axis cause of death fields depending on the table (primary/any cause of death).

Other covariates included in all tables and analyses were age (65-69, 70-74, 75-79, 80-84 or 85+), gender (male or female), race/ethnicity (White, Black, Hispanic, or unknown), and geographical region (Northeast, Midwest, South, or West). Results were reported overall and stratified by age, gender, race/ethnicity, and region.

1. 92002 – 92004,92012 – 92014,99201 – 99205, 99211 – 99215,99241 – 99245,99304 – 99310, 99315 – 99316, 99318,99324 – 99328, 99334 – 99337, 99341 – 99345, 99347 – 99350, 99381 – 99387, 99391 – 99397, 99401 – 99404, 99411 – 99412, 99420,99429, 99461, G0463, T1015
2. 0510-0529, 0982-0983
